# Pharmacotherapy for Children and adolescents with overweight and obesity: a systematic review and network meta-analysis of randomized controlled trials

**DOI:** 10.1101/2023.07.31.23293470

**Authors:** Li Luo, Tingting Huang, Hui Wang, Jianglin Zhao, Yunyun Qi, Zijing Yan, Chunmei Zhu, Chufeng Wang, Na Su, Ting Xu, Shengzhao Zhang

## Abstract

**Background:** Overweight and obesity are widespread among children and adolescents. We aimed to summarize the evidence for the pharmacotherapy as an adjunct to lifestyle interventions in overweight or obese children and adolescents by comparing the benefits and harms.

**Methods:** RCTs (randomized controlled trials) were sourced from PubMed, Embase (using the OVID platform), the Cochrane Library (CENTRAL), as well as the trial registers ICTRP (WHO) and ClinicalTrials.gov. Searches were undertaken from inception to April 25, 2023. A network meta-analysis was performed using the frequentists framework based on random-effects model. We used GRADE (Grading of Recommendations Assessment, Development, and Evaluation) approach to evaluate the overall certainty of evidence and categorized the interventions.

**Results:** In total, 42 RCTs (n=3883) comparing 8 different pharmacotherapy strategies were included in this study. Evidence strongly suggested that phentermine-topiramate reduced BMI the most (the mean difference (MD) -4.83 [95% CI, -7.46 to -2.20] kg/m^2^) and weight (MD, -14.59 [95% CI, -19.37, -9.81] kg) in children and adolescents with overweight or obesity. Compared to lifestyle intervention alone, phentermine-topiramate was associated with an additional 557 events per 1000 person-years in terms of the proportion of participants achieving a BMI reduction of ≥5%, but there was no increased harm in total gastrointestinal adverse effects and discontinuation due to adverse events.

**Conclusions:** Phentermine-topiramate was closely related to weight loss and showed a good tolerability, proving to be the optimal treatment strategy for overweight or obese children and adolescents.

**Registration:** PROSPERO registry number: CRD42022329226

## Background

Overweight and obesity have profound implications for numerous children and adolescents, with the data indicating a persistent upward trend in affected populations. Recent estimates by the World Health Organization (WHO) indicate that in 2020, there were 39 million children under the age of 5 years who were affected by overweight or obesity[1]. Disturbingly, the global prevalence of obesity among individuals aged 5-19 has experienced a notable surge between 1975 and 2020. Among girls, the rates have escalated from less than 1% to 8%, while among boys, the rates have reached 10%[1–3]. Beyond the heightened risk of physical health conditions[4–6], overweight and obesity also significantly impact the societal and emotional well-being of the youth[7, 8]. Clearly, such circumstances hinder the optimal growth and development of children and adolescents, emphasizing the urgency of addressing this matter.

Lifestyle interventions serve as the cornerstone for managing excess weight in children and adolescents grappling with overweight or obesity. These interventions encompass a range of measures, such as making dietary adjustments, promoting physical activity, and fostering behavioral changes.[9, 10]. However, it is worth acknowledging the inherent challenges associated with modifying deeply ingrained behavioral habits, especially among younger individuals[11]. The necessity for long-term commitment further compounds the issue, often resulting in suboptimal patient compliance. Consequently, treatment failures or rapid weight regain following initial progress become all too common[12]. To address these obstacles and enhance outcomes, the incorporation of effective pharmacotherapy strategies is crucial, acting as adjunctive therapy in the maintenance of weight loss[13].

Despite numerous studies have explored pharmacological interventions for weight reduction in adults, the availability of approved drugs for children and adolescents remains limited[14]. Evidence is lacking on the optimal combination of drug therapy with lifestyle intervention for weight loss. Consequently, there is a pressing need for further investigation to evaluate the efficacy and safety of pharmacological approaches in the pediatric and adolescent population. In this network meta-analysis, we aim to summarize the evidence for the use of drug as adjunctive therapy to lifestyle interventions in overweight or obese children and adolescents by comparing the benefits and harms.

## Methods

### Study design

This network meta-analysis was conducted according to the Preferred Reporting Items for Systematic Reviews and Meta-Analyses (PRISMA) 2020 guidelines and the statement standards for network meta-analysis (PRISMA-NMA)[15, 16]. The review was registered in the International Prospective Register of Systematic Reviews, PROSPERO (CRD42022329226)

### Eligibility criteria

We included randomized controlled trials (RCTs) of pharmacological monotherapy or combined therapy for weight reduction in children and adolescents (aged ≤ 18 years) with obesity or overweight, regardless of the presence of the weight-related complications. Trials were excluded if they (1) enrolled patients who were medication-induced obesity, pregnancy, or normal bodyweight; (2) enrolled patients with malignant tumors or blood diseases; (3) were not published in English; (4) had no available results related to weight loss.

### Search strategy

PubMed, Embase (using the OVID platform), and the Cochrane Library (CENTRAL) were conducted from inception to September 24, 2022 for the initial search. Due to the long interval, in order to avoid overlooking any newly published relevant studies during this period and ensure a comprehensive screening, these databases were re-searched, with the deadline set at April 25, 2023. Furthermore, to supplement the identified citations, we also searched International clinical trials registry platform (ICTRP) and ClinicalTrials.gov. Appendix 2 shows the detailed search strategy.

### Study selection

The PRISMA flow diagram was applied to guide the process of study selection. The retrieved studies were checked for duplication using Endnote X9, and duplicate records were removed. Two independent researchers conducted initial screening based on article titles and abstracts and then performed full-texts, excluding irrelevant studies. Discrepancies were resolved through mutual discussion or consultation with a senior member.

### Outcomes

Primary outcomes as determined by our assessment were change in BMI and weight from baseline, as well as the percentage of participants achieving BMI reduction of at least 5%. Secondary outcomes included changes in BMI z-score and BMI standard deviation score (SDS) from baseline, percentage of participants achieving BMI reduction of at least 10%, and adverse outcome, such as total gastrointestinal adverse events, discontinuation due to adverse events, serious adverse events, nausea events, vomiting events and diarrhea events.

### Data extraction and risk of bias assessment

Two reviewers independently extracted relevant data from the included studies and organized them in an Excel spreadsheet, with any discrepancies resolved through discussion with a third reviewer. The extracted information included study characteristics (first author, year of publication, intervention measures), participant information (such as age, gender, sample size) and outcome indicators.

Risk of bias (ROB) for all included randomized controlled trials was assessed by two independent assessors with the Cochrane Risk of Bias 2 (ROB-2) tool[17] and categorized as “high,” “low,” or “some concern,” based on evaluations across five domains: (1) the randomization process, (2) deviations from intended interventions, (3) missing outcome data, (4) measurement of the outcome, (5) selection of the reported result. Disagreements were resolved through discussion with a third assessor.

### Publication bias

We employed funnel plots to assess the potential publication bias of included studies. The symmetry of the funnel plot was visually examined as an initial step. If the number of trials included in the analysis exceeded 10, Egger’s test, Begg-Mazumdar test, and Thompson-Sharp test were conducted to further evaluate publication bias. A p-value greater than 0.05 in these tests indicates no significant publication bias.

### Statistical analysis

The network meta-analyses in this study were performed using the frequentist framework, employing a graph-theoretical method[18]. For dichotomous outcomes, odds ratios (ORs) were calculated, while mean differences (MDs) were utilized for continuous outcomes. Both measures were accompanied by 95% confidence intervals (CIs) to provide a range of estimated effect sizes. A random-effects model was used to account for variability among the included studies. To assess heterogeneity among the studies, Cochran’s Q test was utilized, where a p-value greater than 0.05 indicated no significant heterogeneity. The transitivity assumption was assessed by comparing the distribution of potential effect modifiers across different intervention comparisons. Factors such as baseline age, gender proportion, and mean BMI at baseline were considered as effect modifiers. A detailed description of this analysis can be found in Appendix 6.6. The node-splitting model was applied to analyze the inconsistency between direct and indirect comparison results[19, 20].

In order to rank the performance of interventions in terms of weight loss and safety, we utilized a metric called the P-score. This score is assigned to each intervention and falls within the range of 0 to 1. A higher P-score indicates a greater likelihood of the intervention being ranked as the most effective option.[21]. The absolute risk difference (RD) of the intervention versus lifestyle modification alone was estimated from the relative effect and the estimate of baseline risk, see Appendix 3.3 for detailed calculation formulas.

To ensure the robustness of the main analyses, six sensitivity analyses were conducted. These sensitivity analyses aimed to explore the impact of different factors on the results obtained. The following aspects were considered:

- Sequential exclusion of studies with fewer than 40 participants.
- Exclusion of studies without a placebo control.
- Exclusion of studies that did not report baseline BMI changes.
- Exclusion of studies with a drug treatment duration of less than three months.
- Exclusion of studies judged to have a high risk of bias.
- Network meta-analysis using a random-effects model within the Bayesian framework, as an alternative to the previously employed frequentist method.

To explore the potential influence of mean age, gender, mean BMI and weight at baseline, as well as the duration of trial follow-up on the outcomes, a meta-regression analysis with the restricted maximum likelihood estimator method (REML) was conducted.

All statistical analyses, including the sensitivity analyses and meta-regression, were performed using the R software version 4.2.2.

### Quality of evidence (GRADE) and categorization of interventions

We assessed certainty of the evidence by applying the GRADE (Grading of Recommendations Assessment, Development, and Evaluation) approach, thus reflecting the methodological reliability of the network meta-analysis[22, 23]. We rated the certainty for each comparison and the network estimate results as high, moderate, low, or very low, based on risk of bias, heterogeneity, publication bias, transitivity process, inconsistency, imprecision.

The minimal important differences (MID) of the primary outcomes were identified by searching through previous research studies. We classified the interventions into four categories: among the best, intermediate-possibly better, intermediate-possibly worse, and among the worst, based on whether the effect estimate values were greater than or less than MID[24]. For the outcome of discontinuation due to adverse events and total gastrointestinal events, we set three categories of effect class: among the worst, intermediate, among the best, based on comparisons with other drugs and lifestyle interventions. A detailed description is provided in Appendix 3.5.

## Result

### Characteristics of the included studies

We screened 2072 records from the databases (PubMed, Embase, the Cochrane Library), ICTRP and ClinicalTrials.gov and 42 eligible trials were identified in this network meta-analysis, including 40 RCTs (published between 2001 and 2022) and 2 ongoing RCTs (without any publications, only results available on clinical trial websites). The retrieval flow diagram is shown in Figure 1. A total of 3883 participants were included, with a mean age ranging from 8.1 to 16.1 years. The baseline mean BMI ranged from 26.2 to 41.7 kg/m^2^. Males accounted for 38.14% of the participants. The duration of drugs intervention ranged from 1.25 to 28.2 months. Appendix 4 details the study characteristics.

**Fig. 1.**
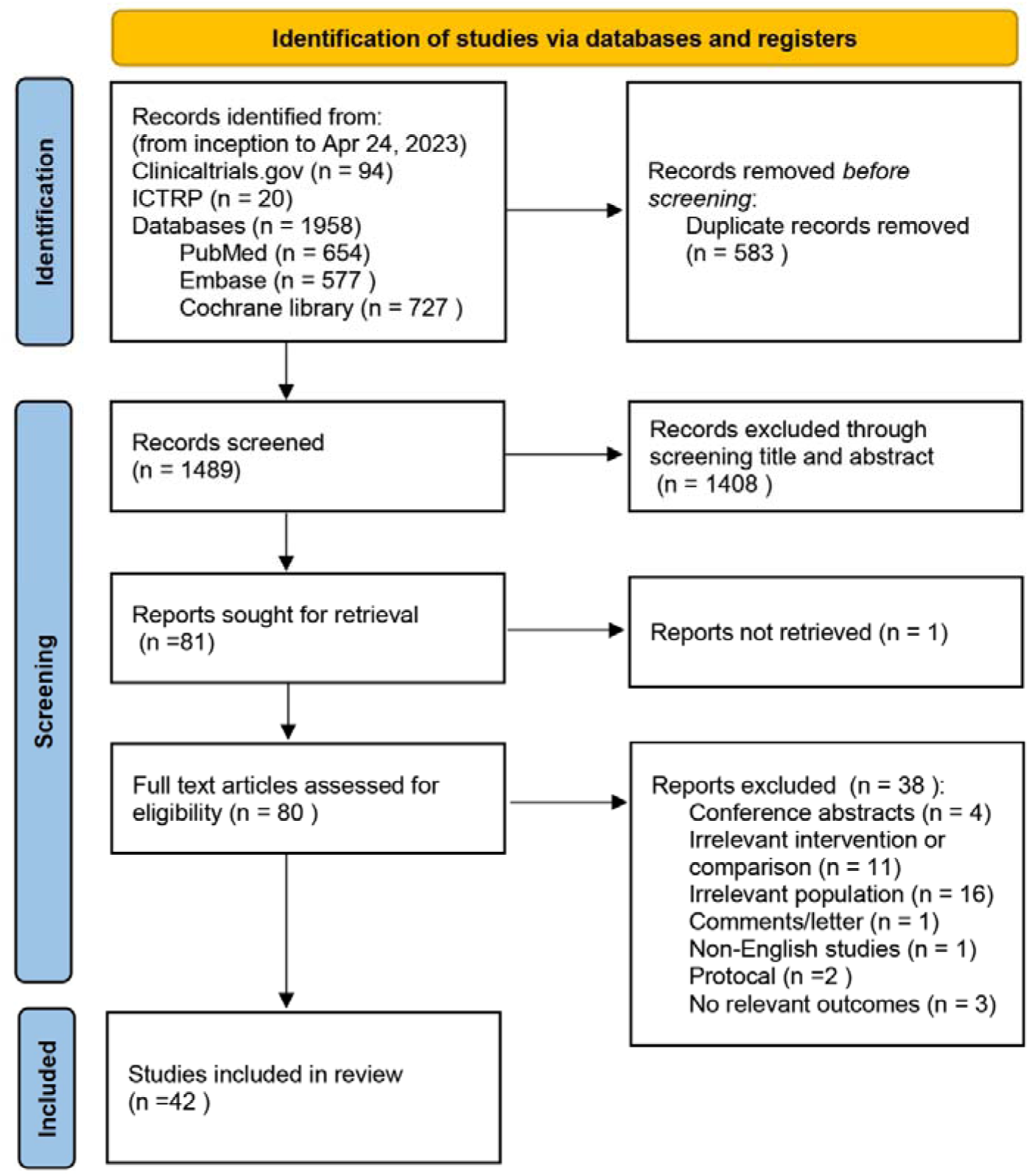
PRISMA flow diagram of the study selection process.

### Risk of bias assessment

A summary of the results on the risk of bias is presented in Appendix 5. One trial was rated as having a high risk of bias using the Rob 2 tool, which was generated by the domain of randomization process. 24 trials included in this network meta-analysis raised some concerns, primarily related to deviations from intended interventions, the randomization process, and missing outcome data.

### Network plots

We established networks involving 12 outcomes. The network plots of the primary outcome are shown in Figure 2. Each circle represents an intervention, and the size of the circle is proportional to the number of participants in that intervention. The line indicates that there is a direct comparison between two interventions. The thickness of the line is proportional to the number of trials. Change in BMI was included in 9 intervention measures, change in weight was included in 7 intervention measures, and the percentage of participants achieving BMI reduction of at least 5% was included in 5 intervention measures. The network diagram for the secondary outcomes can be found in Appendix 6.1.

**Fig. 2.**
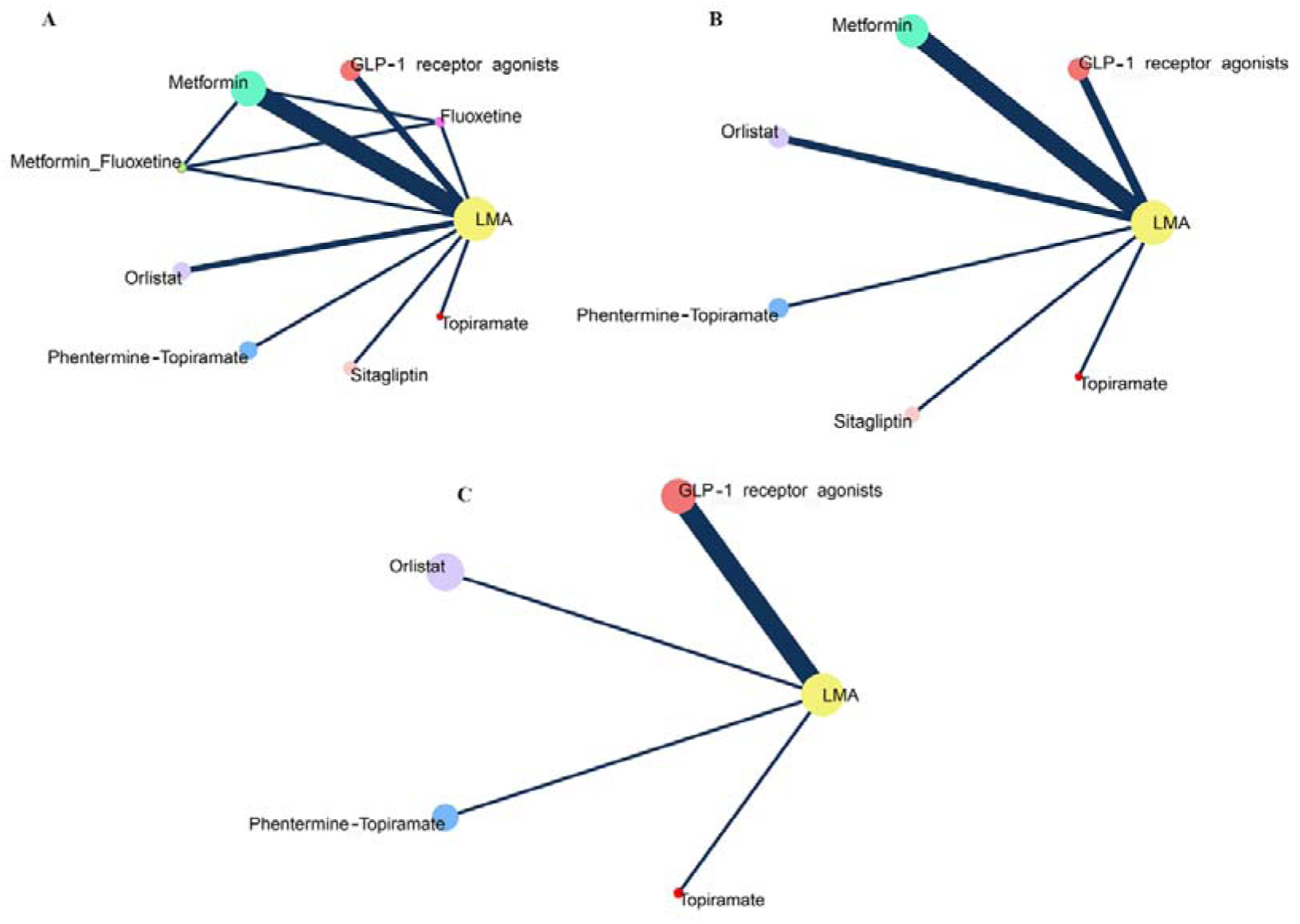
Network meta-analysis plots. **(A)** Change in BMI from baseline, **(B)** Change in weight from baseline, **(C)** Percentage of participants achieving BMI reduction of at least 5%. Each circle represents an intervention, and the size of the circle is proportional to the number of participants in that intervention. The line indicates that there is a direct comparison between two interventions. The thickness of the line is proportional to the number of trials.

### Primary outcomes

Thirty RCTs, involving 2,596 participants, reported the effects of the drugs on changes in BMI from baseline. As shown in Figure 3, Glucagon-Like Peptide-1 (GLP-1) receptor agonists, metformin, orlistat, and phentermine-topiramate showed better effects in reducing BMI compared to lifestyle modification alone, with evidence graded from low to high. In comparison to other interventions, except for fluoxetine and metformin combined with fluoxetine, the BMI reduction values for phentermine-topiramate were as follows: -3.02 (MD, [95%CI, -5.99 to -0.05] kg/m^2^, high-quality evidence) compared to GLP-1 receptor agonists, -4.83 (MD, [95%CI, -7.46 to -2.20] kg/m^2^, high-quality evidence) compared to lifestyle modification alone, -3.73 (MD, [95%CI, -6.45, to -1.01] kg/m^2^, moderate-quality evidence) compared to metformin, -3.17 (MD, [95%CI, -6.23 to -0.10] kg/m^2^, low-quality evidence) compared to orlistat, -5.53 (MD, [95%CI, -9.29 to -1.77] kg/m^2^, high-quality evidence) compared to sitagliptin, -3.97 (MD, [95%CI, -7.91 to -0.03] kg/m^2^, high-quality evidence) compared to topiramate.

**Fig. 3.**
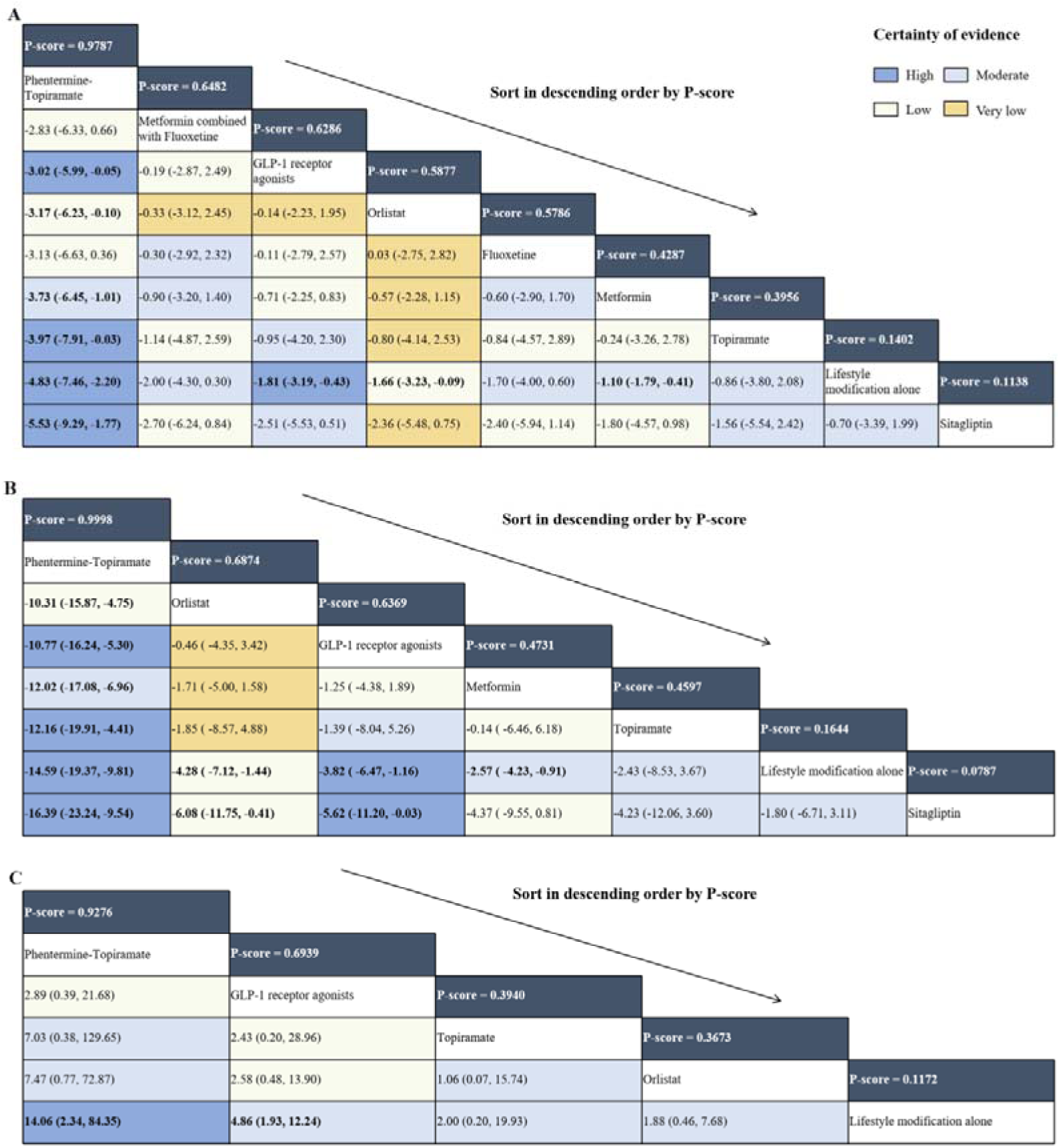
Relative effect sizes of pharmacotherapy and GRADE Rating. The league table for results of network meta-analysis**(A)** Change in BMI from baseline (mean differences, 95% CI), **(B)** Change in weight from baseline (mean differences, 95% CI), **(C)** Percentage of participants achieving BMI reduction of at least 5% (odd ratios, 95% Cl). The league tables showed the relative effects for each primary outcome, with colors representing different levels of evidence (high, moderate, low and very low). Bold text indicates statistical significance. White numbers in the dark blue boxes are P-score. The league table is arranged from left to right in descending order of P-score.

Twenty-four RCTs, involving 2,098 participants, reported the effects of the drugs on change in weight from baseline. GLP-1 receptor agonists and phentermine-topiramate both demonstrated significant weight loss effects, with bodyweight reductions of -3.82 (MD, [95% CI, -6.47 to -1.16] kg) and -14.59 (MD, [95% CI, -19.37, -9.81] kg), respectively, compared to lifestyle modification alone. The evidence quality for the two comparisons was rated as high. The MD of orlistat compared to lifestyle modification alone was -4.28 [95%, -7.12 to -1.44] kg, which was evaluated as a low-quality evidence. Most worthy of mention was that phentermine-topiramate exhibited a stronger weight loss effect, with a reduction of -10.77 (MD, [95% CI, -16.24, -5.30] kg, high-quality evidence) compared to GLP-1 receptor agonists. Furthermore, compared to orlistat, it resulted in a weight reduction of -10.31 (MD, [95% CI, -15.87, -4.75] kg, low-quality evidence).

For the outcome of percentage of participants achieving a BMI reduction of at least 5%, low- to moderate-quality evidence indicated no significant differences among the different drug therapies. However, compared to lifestyle modification alone, the proportion of participants who achieved a 5% or more BMI reduction might increase when using phentermine-topiramate and GLP-1 receptor agonists, with odds ratios (OR) of 14.06 ([95% CI, 2.34 to 84.35], high-quality evidence) and 4.86 ([95% CI, 1.93 to 12.24], moderate-quality evidence), respectively.

### Secondary outcomes

The relative effects of secondary outcome measures are shown in the Appendix 6.2. For the outcome of percentage of participants achieving a BMI reduction of at least 10%, high- and moderate-quality evidence indicated phentermine-topiramate and GLP-1 receptor agonists were better than lifestyle modification alone, respectively. Additionally, GLP-1 receptor agonists also showed a strong effect on the change of BMI SDS from baseline. However, in terms of gastrointestinal adverse effects (such as nausea events, vomiting events, diarrhea events), high-quality evidence demonstrated that GLP-1 receptor agonists occurred significantly more frequently than lifestyle interventions. Concerning the change in BMI z-score from baseline, metformin and orlistat resulted in greater reductions than lifestyle modification alone (high-quality evidence). Furthermore, high-quality evidence indicated that metformin had a higher incidence of nausea events compared to lifestyle intervention and orlistat, while orlistat was associated with a higher incidence of diarrhea events (high-quality evidence) and a higher likelihood of discontinuation due to any adverse events (moderate-quality evidence) compared to lifestyle intervention. No significant differences were observed between all drugs and lifestyle intervention concerning severe adverse events.

### Categorization of interventions

The estimated values of the lifestyle intervention in a single arm trial were used as MID (minimal important differences) for weight loss, which was -4 kg, and the MID for BMI change, which was -1.4 kg/m^2^ [25]. The MID for the percentage of participants achieving BMI reduction of at least 5% was set as approximately double the percentage of the lifestyle modification alone group (see Appendix 3.4). The primary benefit outcomes were assessed according to the MID to determine clinical importance. Additionally, we classified the harm outcomes of interventions into three categories: among the worst, intermediate and among the best. These categorizations were based on the comparisons with lifestyle modification alone and other drugs. Figure 4 illustrates the categorization of interventions.

**Fig. 4.**
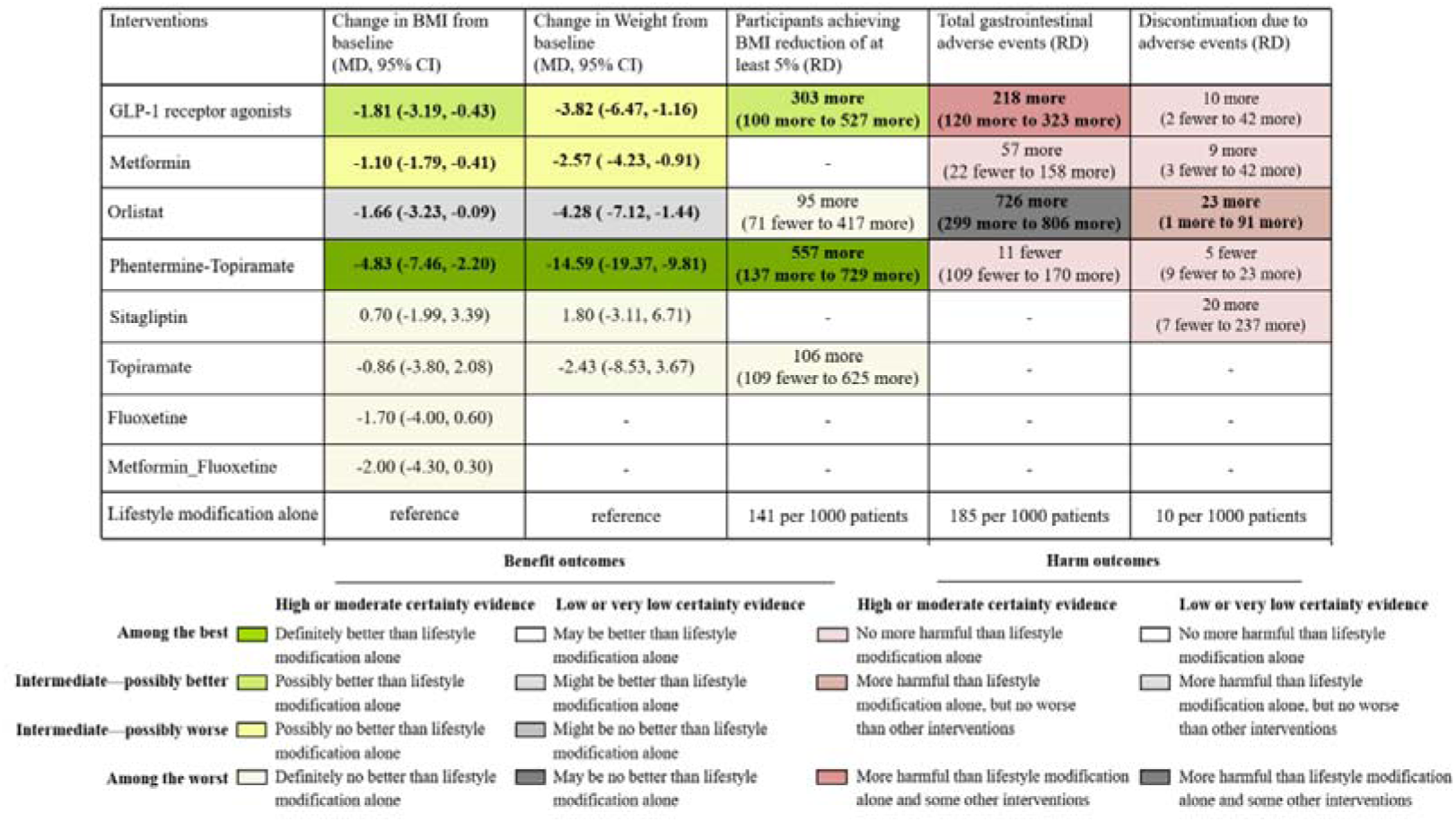
Relative effects and absolute effects of interventions on benefit and harm outcomes. According to MID and effect estimate values, we classified the effectiveness of interventions into four categories: among the best, intermediate-possibly better, intermediate-possibly worse, and among the worst. For the outcome of discontinuation due to adverse events and total gastrointestinal events, we set three categories of effect class: among the worst, intermediate, among the best, based on comparisons with other drugs and lifestyle interventions.

For the outcomes of changes in BMI and weight from baseline, phentermine-topiramate was evaluated as the most effective drug (high-quality evidence), with the upper limit of the 95% CI for its estimated effect size was smaller than the MID. Considering the point estimate of orlistat is lower than the MID and its 95% CI intersects with the MID, it was categorized as might be better than lifestyle modification alone (low-quality evidence). GLP-1 receptor agonists proved to be intermediate-possibly better in terms of BMI change (high-quality evidence). But in terms of weight change, GLP-1 receptor agonists proved to be intermediate-possibly worse as the 95% CI intersected with the MID and the point estimate was greater than the MID (high-quality evidence).

We calculated the absolute effects for dichotomous outcomes (see Figure 4 and Appendix 6.3). Lifestyle modification alone resulted in 141 achieving a BMI reduction of at least 5% per 1000 person-years. Phentermine-topiramate had 557 more events per 1000 person-years with high-quality evidence, followed by GLP-1 receptor agonists with 303 more events (moderate-quality evidence). They were respectively evaluated as among the best and intermediate-possibly better.

Regarding discontinuation due to adverse events, except for orlistat (had 557 more events per 1000 person-years than lifestyle modification alone), there were no significant differences observed between the other drugs and lifestyle modification alone. For total gastrointestinal adverse events, both GLP-1 receptor agonists and orlistat were evaluated as more harmful than lifestyle modification alone and some other drugs.

### Assessment of heterogeneity, transitivity, inconsistency and publication bias

Heterogeneity, transitivity, and inconsistency of the network meta-analysis were evaluated. The heterogeneity assessment results are described in Appendix 6.5. For transitivity, there was no significant differences in the distribution of the possible effect modifiers across intervention comparisons were observed in all networks (Appendix 6.6). The node-splitting method indicated that there was no inconsistency between indirect and direct estimates in the outcome of BMI change from baseline (Appendix 6.7). The funnel plots appeared quite symmetrical, suggesting the absence of publication bias for all the outcomes (Appendix 6.8).

### Meta-regression and sensitivity analyses

Meta-regression analysis revealed a significant correlation between age and change in BMI from baseline (coefficient: 0.4455, 95%CI: 0.1201 to 0.7708, p-value: 0.0073). The weight loss was proportional to the duration of the intervention (coefficient: 0.3456, 95%CI: 0.0938 to 0.5975, p-value: 0.0071). In meta-regression models, participants’ age, BMI and weight at baseline, gender, and length of drug therapy were not associated with the percentage of participants achieving BMI reduction of at least 5%. Appendix 9 details other outcomes. Sensitivity analyses showed that the trend of MD and OR values were consistent with the primary results, confirming the robustness of our findings (Appendix 10).

## Discussion

Based on 42 trials and 3883 participants, a frequentist network meta-analysis was performed to evaluate the efficacy and safety for eight drugs targeting overweight or obese children and adolescents. Compared to lifestyle modification alone, GLP-1 receptor agonists, metformin, orlistat, and phentermine-topiramate all showed significant benefits in terms of BMI reduction and bodyweight loss (high-to low-quality evidence). Phentermine-topiramate was proved to be the optimal treatment strategy for weight management in overweight or obese children and adolescents to manage body weight, with the greatest impact on BMI reduction, weight loss, and the proportion of participants who achieved a reduction in BMI ≥ 5%. Moreover, it was not more harmful than lifestyle modification alone in terms of gastrointestinal adverse effects.

However, our study found some heterogeneity. For the terms of BMI change and weight change from baseline, heterogeneity was observed when comparing lifestyle modification alone with metformin and orlistat. Several factors may contribute to the heterogeneity. Ozkan B 2004[26], assessed as having a high risk of bias due to issues with the randomization process, was included in the orlistat trials to analysis the outcomes of change in BMI and weight, which may be a source of heterogeneity. The inclusion of non-blinded studies in the network meta-analysis may lead to an increase in heterogeneity of the results. Similarly, some trials were assessed with some concerns regarding the randomization process, deviations from the intended intervention and missing outcome data, which were included in the metformin trials. The small sample sizes may also lead to a lower power to detect differences in the outcomes. Additionally, meta-regression indicated an association between age and BMI changes, as well as between study duration and weight changes, suggesting that the heterogeneity is likely a result of differences in study duration and age of participant. In the percentage of participants who achieved a BMI reduction of at least 5%, there was also heterogeneity when comparing GLP-1 receptor agonists to lifestyle modification alone. The heterogeneity may be attributed to Diene G 2022[27], because of its relatively small sample size and some concerns regarding randomization process. Despite the presence of these heterogeneities, transitivity assessment yielded favorable results in this network meta-analysis. To ensure the reliability of our findings, we downgraded the quality of evidence for comparisons with heterogeneity and risk of bias during the GRADE rating. Additionally, sensitivity analyses conducted by excluding trials from different perspectives also confirmed the robustness of the results.

Phentermine-topiramate received its pediatric first approval on 20 July, 2022 by the FDA (Food and Drug Administration), as an adjunct to a dietary habit modification and increased physical activity, for chronic weight management in pediatric obese patients aged ≥ 12 years[28, 29]. This indication was supported by a 56-week, double-blind, placebo-controlled trial[30], which was included in our study. Of note, this medication is not approved for use by the EMA (The European Medicines Agency). Phentermine as a noradrenaline reuptake inhibition can reduce appetite and induce a feeling of satiety[31]. Topiramate reduces caloric intake by enhancing the activity of GABA (γ-aminobutyric acid) neurotransmitter and inhibiting carbonic anhydrase[32]. The complementary mechanisms of both medications allow for reducing the dosage of each drug while achieving weight loss, thereby enhancing safety and tolerability[33, 34]. We are the first to include phentermine-topiramate in a network meta-analysis of pediatric drug therapy. Compared to various intervention measures, phentermine-topiramate exhibited satisfactory results in terms of reducing BMI and promoting weight loss. It is widely acknowledged that lifestyle intervention is the primary weight management strategy for overweight or obese children and adolescents. Importantly, high-quality evidence suggested that compared to lifestyle intervention alone, phentermine-topiramate was associated with an additional 557 events per 1000 person-years in terms of the proportion of participants achieving a BMI reduction of ≥ 5% in our analysis. A previous study in adult populations had also suggested a significant beneficial effect on weight loss[35]. The difference was that phentermine-topiramate had a higher incidence of gastrointestinal adverse effects and discontinuation due to any adverse event than lifestyle modification alone in adult populations, which was not observed in children and adolescent patients.

GLP-1 receptor agonists, which stimulate postprandial insulin secretion and inhibit glucagon secretion in a glucose-dependent manner, were originally used to treat T2DM (type 2 diabetes mellitus) [36]. Additionally, this anti-diabetic drug was shown to benefit weight loss in subsequent studies[37–40]. Its mechanism involves targeting receptors in the hypothalamus to suppress appetite, controlling body weight through both a direct effect of reducing food intake and an indirect effect of slowing gastric emptying[41]. Currently, two GLP-1 receptor agonists, liraglutide and semaglutide, have been approved by the FDA and EMA for weight management in adolescents[42–45]. Review previous studies on weight loss in children and adolescents, Ryan PM 2021 identified an effect size of -1.24 (MD, [95% CI, -1.71 to -0.77] kg/m^2^, 5 trials) on BMI reduction and -1.50 (MD, [95% CI, -2.50 to 0.50] kg, 7 trials) on weight loss for GLP-1 receptor agonists compared with placebo[46], In Chadda KR 2021, GLP-1 agonist therapy resulted in a reduction in body weight of -2.74 (MD, [95% CI, -3.77 to -1.70] kg, 6 trials) and a decrease in BMI of -1.25 (MD, [95% CI, -1.70 to -0.80] kg/m^2^, 4 trials) [47]. The observed smaller effects in both of these reviews compared to our findings may be attributed to the differences in the number of included studies. In addition to the favorable changes in BMI and weight, we found that 303 more participants per 1000 person-years experienced a 5% decrease in BMI compared to lifestyle modification alone. This result is slightly inferior to phentermine-topiramate. Of note, it is associated with a higher incidence of gastrointestinal adverse effects.

Orlistat is currently the only weight-loss drug approved by the FDA for the long-term treatment of obesity in individuals 12 years or older[48]. By inhibiting gastrointestinal lipase, it reduces the absorption of approximately 30% of dietary fats. However, its frequent gastrointestinal side effects, such as urgency of bowel movements, increased frequency of bowel movements, and oily stools, limit treatment compliance[49, 50]. Our study also provided evidence of a higher incidence of gastrointestinal adverse events and poorer tolerance. For BMI reduction, a systematic review of treatment for pediatric obesity estimated an effect size of -0.7 (MD, [95% CI, -1.2 to -0.3] kg/m^2^) compared to placebo[51]. Similar findings were reported in three additional studies, with the observed reduction in BMI of -0.83 (MD, [95% CI, -1.19 to-0.47] kg/m^2^, 2 trials)[52], -0.79 (MD, [95% CI, -1.08 to -0.51] kg/m^2^, 2 trials and 1 ongoing study)[53], and -0.76 (MD, [95% CI, -1.07 to -0.44] kg/m^2^, 2 trials)[54], respectively. In our study, we included the additional orlistat trials and performed a comprehensive pooling of four studies (3 trials and 1 ongoing study), which yielded a point estimate of -1.66 (MD, [95%CI, -3.23 to -0.09] kg/m^2^). Consistent with our findings, a meta-analysis encompassing 3 orlistat trials also demonstrated the estimate of -1.67 (MD, [95% CI, -3.52 to -0.18] kg/m^2^)[55], thereby reinforcing the validity and reliability of our results.

Metformin, a biguanide medication, is also an FDA-approved drug for T2DM in children aged 10 years and older, but not approved for weight loss. Its potential appetite-suppressing and weight-reducing effects make it popular among obese children and adolescents[56, 57]. Mead E 2016 conducted a review that pooled 8 trials studying the effect of metformin on BMI reduction and reported the MD value of -1.35 [95% CI, -2 to -0.69] kg/m^2^[53]. Our network meta-analysis yielded comparable results, with the MD of -1.10 ([95%CI, -1.79 to -0.41] kg/m^2^, 18 trials). As a treatment for T2DM, the BMI-reducing effect of metformin is relatively mild; however, it is safer. A study conducted in a real-world clinical setting supports this result, showing that metformin as a weight loss aid in youth with obesity provides modest benefits and generally well tolerated[58].

As far as we know, this network meta-analysis represents the most comprehensive evidence base to date on pharmacological treatment in overweight or obese children and adolescents, and the findings are not limited by unpublished trials. We employed comprehensive statistical methods to analyze each outcome, including heterogeneity, inconsistency, and transitivity. The multi-faceted sensitivity analyses and GRADE assessments further validated the reliability of our results. In addition, we calculated the absolute effects, and classified interventions according to minimal important differences.

We acknowledge several limitations of this review. Firstly, the number of included studies was limited, and the available data were insufficient to evaluate the effects of each individual drug within the GLP-1 receptor agonist class. Due to the same reasons, a dose-response analysis was not conducted. Secondly, the focus of our analysis was exclusively on gastrointestinal-related adverse events, as there were insufficient studies to support including adverse reactions from other organ systems as outcome measures. Thirdly, due to fewer studies included in our analysis reported the extent of long-term weight maintenance after discontinuing drug treatment, it was not possible to analyze the possibility of weight gain after discontinuation.

## Conclusions

In conclusion, this network meta-analysis summarized and compared the pharmacological treatments for overweight or obese children and adolescents, providing information for clinical decision-making. The evidence suggests that phentermine-topiramate as an adjunct medication to lifestyle interventions is closely related to weight management and has fewer adverse effects, although this needs larger studies in a range of young clinical populations to verify the effectiveness and long-term maintenance of weight loss.

## Data Availability

The data can be obtained by contacting the corresponding author.

## Declaration

### Availability of data and materials

The data can be obtained by contacting the corresponding author.

### Authors’ contributors

LL and TH contributed equally. SZ, LL, TH and JZ conceived and designed the study. LL, TH and HW screened articles, LL, TH and CZ extracted the data. YQ, ZY, CW assessed ROB. LL, TH, HW, TX, NS contributed to the statistical analysis. SZ obtained funding for this study, and supervised the work. LL, TH and SZ rated the certainty of evidence. LL,TH, YQ and ZY drafted the manuscript. TH, LL, TX, NS and SZ contributed to the revision and discussed the fnal edition. All authors read and approved the final manuscript.

## References

1. The Lancet Public H: Childhood obesity beyond COVID-19. Lancet Public Health. 2021; 6(8):e534.

2. Di Cesare M, Sorić M, Bovet P, Miranda JJ, Bhutta Z, Stevens GA, Laxmaiah A, Kengne AP, Bentham J: The epidemiological burden of obesity in childhood: a worldwide epidemic requiring urgent action. BMC Med. 2019; 17(1):212.

3. World Obesity Federation. World Obesity Atlas 2023. [https://data.worldobesity.org/publications/?cat=19]

4. Peinado Fabregat MI, Saynina O, Sanders LM: Obesity and Overweight Among Children With Medical Complexity. Pediatrics. 2023; 151(1).

5. Kumar S, Kelly AS: Review of Childhood Obesity: From Epidemiology, Etiology, and Comorbidities to Clinical Assessment and Treatment. Mayo Clin Proc. 2017; 92(2):251–265.

6. Lanigan J, Sauven N: Treatment of childhood obesity: a multidisciplinary approach. Clinics in Integrated Care. 2020; 3:100026.

7. Rao W-W, Zong Q-Q, Zhang J-W, An F-R, Jackson T, Ungvari GS, Xiang Y, Su Y-Y, D’Arcy C, Xiang Y-T: Obesity increases the risk of depression in children and adolescents: Results from a systematic review and meta-analysis. J of Affective Disorders. 2020; 267:78–85.

8. Halfon N, Larson K, Slusser W: Associations Between Obesity and Comorbid Mental Health, Developmental, and Physical Health Conditions in a Nationally Representative Sample of US Children Aged 10 to 17. Acad Pediatr. 2013; 13(1):6–13.

9. Calcaterra V, Rossi V, Mari A, Casini F, Bergamaschi F, Zuccotti GV, Fabiano V: Medical treatment of weight loss in children and adolescents with obesity. Pharmacol Res. 2022; 185:106471.

10. Ebbeling CB, Pawlak DB, Ludwig DS: Childhood obesity: public-health crisis, common sense cure. The Lancet. 2002; 360(9331):473–482.

11. Sothern MS: EXERCISE AS A MODALITY IN THE TREATMENT OF CHILDHOOD OBESITY. Pediatr Clin N Am. 2001; 48(4):995–1015.

12. Varkevisser RDM, van Stralen MM, Kroeze W, Ket JCF, Steenhuis IHM: Determinants of weight loss maintenance: a systematic review. Obes Rev. 2019; 20(2):171–211.

13. Steinbeck KS, Lister NB, Gow ML, Baur LA: Treatment of adolescent obesity. Nat Rev Endocrinol. 2018; 14(6):331–344.

14. Güngör NK: Overweight and obesity in children and adolescents. J Clin Res Pediatr Endocrinol. 2014; 6(3):129–143.

15. Hutton B, Salanti G, Caldwell DM, Chaimani A, Schmid CH, Cameron C, Ioannidis JP, Straus S, Thorlund K, Jansen JP et al: The PRISMA extension statement for reporting of systematic reviews incorporating network meta-analyses of health care interventions: checklist and explanations. Ann Intern Med. 2015; 162(11):777–784.

16. Page MJ, McKenzie JE, Bossuyt PM, Boutron I, Hoffmann TC, Mulrow CD, Shamseer L, Tetzlaff JM, Akl EA, Brennan SE, et al: The PRISMA 2020 statement: an updated guideline for reporting systematic reviews. Bmj. 2021; 372:n71.

17. Sterne JAC, Savović J, Page MJ, Elbers RG, Blencowe NS, Boutron I, Cates CJ, Cheng HY, Corbett MS, Eldridge SM et al: RoB 2: a revised tool for assessing risk of bias in randomised trials. Bmj. 2019; 366:l4898.

18. Shim SR, Kim SJ, Lee J, Rücker G: Network meta-analysis: application and practice using R software. Epidemiol Health. 2019; 41:e2019013.

19. Jiang N, Rao F, Xiao J, Yang J, Wang W, Li Z, Huang R, Liu Z, Guo T: Evaluation of different surgical dressings in reducing postoperative surgical site infection of a closed wound: A network meta-analysis. Int J Surg. 2020; 82:24–29.

20. Higgins JP, Jackson D, Barrett JK, Lu G, Ades AE, White IR: Consistency and inconsistency in network meta-analysis: concepts and models for multi-arm studies. Res Synth Methods. 2012; 3(2):98–110.

21. Rücker G, Schwarzer G: Ranking treatments in frequentist network meta-analysis works without resampling methods. BMC Med Res Methodol. 2015; 15:58.

22. Puhan MA, Schünemann HJ, Murad MH, Li T, Brignardello-Petersen R, Singh JA, Kessels AG, Guyatt GH: A GRADE Working Group approach for rating the quality of treatment effect estimates from network meta-analysis. Bmj. 2014; 349:g5630.

23. Brignardello-Petersen R, Bonner A, Alexander PE, Siemieniuk RA, Furukawa TA, Rochwerg B, Hazlewood GS, Alhazzani W, Mustafa RA, Murad MH et al: Advances in the GRADE approach to rate the certainty in estimates from a network meta-analysis. J Clin Epidemiol. 2018; 93:36–44.

24. Zeng L, Brignardello-Petersen R, Hultcrantz M, Siemieniuk RAC, Santesso N, Traversy G, Izcovich A, Sadeghirad B, Alexander PE, Devji T et al: GRADE guidelines 32: GRADE offers guidance on choosing targets of GRADE certainty of evidence ratings. J Clin Epidemiol. 2021; 137:163–175.

25. Chen AK, Roberts CK, Barnard RJ: Effect of a short-term diet and exercise intervention on metabolic syndrome in overweight children. Metabolism. 2006; 55(7):871–878.

26. Ozkan B, Bereket A, Turan S, Keskin S: Addition of orlistat to conventional treatment in adolescents with severe obesity. Eur J Pediatr. 2004; 163(12):738–741.

27. Diene G, Angulo M, Hale PM, Jepsen CH, Hofman PL, Hokken-Koelega A, Ramesh C, Turan S, Tauber M: Liraglutide for Weight Management in Children and Adolescents With Prader-Willi Syndrome and Obesity. J Clin Endocrinol Metab. 2022; 108(1):4–12.

28. Dhillon S: Phentermine/Topiramate: Pediatric First Approval. Paediatr Drugs. 2022; 24(6):715–720.

29. FDA Approves Treatment for Chronic Weight Management in Pediatric Patients Aged 12 Years and Older [https://www.fda.gov/drugs/news-events-human-drugs/fda-approves-treatment-chronic-weight-management-pediatric-patients-aged-12-years-and-older]

30. Kelly AS, Bensignor MO, Hsia DS, Shoemaker AH, Shih W, Peterson C, Varghese ST: Phentermine/Topiramate for the Treatment of Adolescent Obesity. NEJM Evid. 2022; 1(6).

31. Heal DJ, Gosden J, Smith SL: What is the prognosis for new centrally-acting anti-obesity drugs? Neuropharmacology. 2012; 63(1):132–146.

32. Astrup A, Toubro S: Topiramate: a new potential pharmacological treatment for obesity. Obes Res. 2004; 12 Suppl:167s–173s.

33. Gadde KM, Allison DB, Ryan DH, Peterson CA, Troupin B, Schwiers ML, Day WW: Effects of low-dose, controlled-release, phentermine plus topiramate combination on weight and associated comorbidities in overweight and obese adults (CONQUER): a randomised, placebo-controlled, phase 3 trial. Lancet. 2011; 377(9774):1341–1352.

34. Garvey WT, Ryan DH, Look M, Gadde KM, Allison DB, Peterson CA, Schwiers M, Day WW, Bowden CH: Two-year sustained weight loss and metabolic benefits with controlled-release phentermine/topiramate in obese and overweight adults (SEQUEL): a randomized, placebo-controlled, phase 3 extension study. Am J Clin Nutr. 2012; 95(2):297–308.

35. Shi Q, Wang Y, Hao Q, Vandvik PO, Guyatt G, Li J, Chen Z, Xu S, Shen Y, Ge L et al: Pharmacotherapy for adults with overweight and obesity: a systematic review and network meta-analysis of randomised controlled trials. Lancet. 2022; 399(10321):259–269.

36. Fruci B, Giuliano S, Mazza A, Malaguarnera R, Belfiore A: Nonalcoholic Fatty liver: a possible new target for type 2 diabetes prevention and treatment. Int J Mol Sci. 2013; 14(11):22933–22966.

37. Drucker DJ: GLP-1 physiology informs the pharmacotherapy of obesity. Mol Metab. 2022; 57:101351.

38. Basolo A, Burkholder J, Osgood K, Graham A, Bundrick S, Frankl J, Piaggi P, Thearle MS, Krakoff J: Exenatide has a pronounced effect on energy intake but not energy expenditure in non-diabetic subjects with obesity: A randomized, double-blind, placebo-controlled trial. Metabolism. 2018; 85:116–125.

39. Wadden TA, Walsh OA, Berkowitz RI, Chao AM, Alamuddin N, Gruber K, Leonard S, Mugler K, Bakizada Z, Tronieri JS: Intensive Behavioral Therapy for Obesity Combined with Liraglutide 3.0 mg: A Randomized Controlled Trial. Obesity (Silver Spring). 2019; 27(1):75–86.

40. Wadden TA, Hollander P, Klein S, Niswender K, Woo V, Hale PM, Aronne L: Weight maintenance and additional weight loss with liraglutide after low-calorie-diet-induced weight loss: the SCALE Maintenance randomized study. Int J Obes (Lond). 2013; 37(11):1443–1451.

41. Vergès B, Bonnard C, Renard E: Beyond glucose lowering: glucagon-like peptide-1 receptor agonists, body weight and the cardiovascular system. Diabetes Metab. 2011; 37(6):477–488.

42. Food and Drug Administration. FDA Approves Weight Management Drug for Patients Aged 12 and Older [https://www.fda.gov/drugs/news-events-human-drugs/fda-approves-weight-management-drug-patients-aged-12-and-older]

43. MedwireNews. EMA Approves Liraglutide for Teenagers with Obesity [https://diabetes.medicinematters.com/en-GB/liraglutide--obesity-/adolescents/ema-approves-liraglutide-for-teenagers-with-obesity/19028106]

44. MedwireNews. FDA Approves High-dose Semaglutide for Adolescents with Obesity [https://diabetes.medicinematters.com/en-GB/semaglutide/obesity/us-fda-children-obesity/23902216]

45. European Medicines Agency. Union Register of Medicinal Products for Human Use [https://ec.europa.eu/health/documents/community-register/html/h1608.htm]

46. Ryan PM, Seltzer S, Hayward NE, Rodriguez DA, Sless RT, Hawkes CP: Safety and Efficacy of Glucagon-Like Peptide-1 Receptor Agonists in Children and Adolescents with Obesity: A Meta-Analysis. J Pediatr. 2021; 236:137–147.e113.

47. Chadda KR, Cheng TS, Ong KK: GLP-1 agonists for obesity and type 2 diabetes in children: Systematic review and meta-analysis. Obes Rev. 2021; 22(6):e13177.

48. Sherafat-Kazemzadeh R, Yanovski SZ, Yanovski JA: Pharmacotherapy for childhood obesity: present and future prospects. Int J Obesity. 2013; 37(1):1–15.

49. Chanoine JP, Hampl S, Jensen C, Boldrin M, Hauptman J: Effect of orlistat on weight and body composition in obese adolescents: a randomized controlled trial. Jama. 2005; 293(23):2873–2883.

50. Davidson MH, Hauptman J, DiGirolamo M, Foreyt JP, Halsted CH, Heber D, Heimburger DC, Lucas CP, Robbins DC, Chung J et al: Weight control and risk factor reduction in obese subjects treated for 2 years with orlistat: a randomized controlled trial. Jama. 1999; 281(3):235–242.

51. McGovern L, Johnson JN, Paulo R, Hettinger A, Singhal V, Kamath C, Erwin PJ, Montori VM: Clinical review: treatment of pediatric obesity: a systematic review and meta-analysis of randomized trials. J Clin Endocrinol Metab. 2008; 93(12):4600–4605.

52. Viner RM, Hsia Y, Tomsic T, Wong IC: Efficacy and safety of anti-obesity drugs in children and adolescents: systematic review and meta-analysis. Obes Rev. 2010; 11(8):593–602.

53. Mead E, Atkinson G, Richter B, Metzendorf MI, Baur L, Finer N, Corpeleijn E, O’Malley C, Ells LJ: Drug interventions for the treatment of obesity in children and adolescents. Cochrane Database Syst Rev. 2016; 11(11):Cd012436.

54. Oude Luttikhuis H, Baur L, Jansen H, Shrewsbury VA, O’Malley C, Stolk RP, Summerbell CD: Interventions for treating obesity in children. Cochrane Database Syst Rev. 2009; (1):Cd001872.

55. Czernichow S, Lee CM, Barzi F, Greenfield JR, Baur LA, Chalmers J, Woodward M, Huxley RR: Efficacy of weight loss drugs on obesity and cardiovascular risk factors in obese adolescents: a meta-analysis of randomized controlled trials. Obes Rev. 2010; 11(2):150–158.

56. Day EA, Ford RJ, Smith BK, Mohammadi-Shemirani P, Morrow MR, Gutgesell RM, Lu R, Raphenya AR, Kabiri M, McArthur AG et al: Metformin-induced increases in GDF15 are important for suppressing appetite and promoting weight loss. Nat Metab. 2019; 1(12):1202–1208.

57. Malin SK, Kashyap SR: Effects of metformin on weight loss: potential mechanisms. Curr Opin Endocrinol Diabetes Obes. 2014; 21(5):323–329.

58. Kyler KE, Kadakia RB, Palac HL, Kwon S, Ariza AJ, Binns HJ: Use of Metformin for Weight Management in Children and Adolescents With Obesity in the Clinical Setting. Clin Pediatr (Phila). 2018; 57(14):1677–1685.

