## Supplementary appendix for "Pharmacotherapy for Children and adolescents with overweight and obesity: a systematic review and network meta-analysis of randomized controlled trials"

### Appendix 1: PRISMA Checklist

| **Section and Topic** | **Item #** | **Checklist item** | **Location where item is reported** |
| --- | --- | --- | --- |
| **TITLE** | | |  |
| Title | 1 | Identify the report as a systematic review. | Line 1-2 |
| **ABSTRACT** | | |  |
| Abstract | 2 | See the PRISMA 2020 for Abstracts checklist. | Line 19-41 |
| **INTRODUCTION** | | |  |
| Rationale | 3 | Describe the rationale for the review in the context of existing knowledge. | Line 45-69 |
| Objectives | 4 | Provide an explicit statement of the objective(s) or question(s) the review addresses. | Line 70-71 |
| **METHODS** | | |  |
| Eligibility criteria | 5 | Specify the inclusion and exclusion criteria for the review and how studies were grouped for the syntheses. | Line 78-84 |
| Information sources | 6 | Specify all databases, registers, websites, organisations, reference lists and other sources searched or consulted to identify studies. Specify the date when each source was last searched or consulted. | Line 86-91 |
| Search strategy | 7 | Present the full search strategies for all databases, registers and websites, including any filters and limits used. | Appendix 2 |
| Selection process | 8 | Specify the methods used to decide whether a study met the inclusion criteria of the review, including how many reviewers screened each record and each report retrieved, whether they worked independently, and if applicable, details of automation tools used in the process. | Line 92-97 |
| Data collection process | 9 | Specify the methods used to collect data from reports, including how many reviewers collected data from each report, whether they worked independently, any processes for obtaining or confirming data from study investigators, and if applicable, details of automation tools used in the process. | Line 106-109 |
| Data items | 10a | List and define all outcomes for which data were sought. Specify whether all results that were compatible with each outcome domain in each study were sought (e.g. for all measures, time points, analyses), and if not, the methods used to decide which results to collect. | Line 98-104, 79-81 |
|  | 10b | List and define all other variables for which data were sought (e.g. participant and intervention characteristics, funding sources). Describe any assumptions made about any missing or unclear information. | Line 108-109 |
| Study risk of bias assessment | 11 | Specify the methods used to assess risk of bias in the included studies, including details of the tool(s) used, how many reviewers assessed each study and whether they worked independently, and if applicable, details of automation tools used in the process. | Line 110-114,  Appendix 3.2 |
| Effect measures | 12 | Specify for each outcome the effect measure(s) (e.g. risk ratio, mean difference) used in the synthesis or presentation of results. | Line 122-125 |
| Synthesis methods | 13a | Describe the processes used to decide which studies were eligible for each synthesis (e.g. tabulating the study intervention characteristics and comparing against the planned groups for each synthesis (item #5)). | Line 122-125 |
|  | 13b | Describe any methods required to prepare the data for presentation or synthesis, such as handling of missing summary statistics, or data conversions. | Line 122-138, |
|  | 13c | Describe any methods used to tabulate or visually display results of individual studies and syntheses. | Line 133-138, |
|  | 13d | Describe any methods used to synthesize results and provide a rationale for the choice(s). If meta-analysis was performed, describe the model(s), method(s) to identify the presence and extent of statistical heterogeneity, and software package(s) used. | Line 122-153,  Appendix 3 |
|  | 13e | Describe any methods used to explore possible causes of heterogeneity among study results (e.g. subgroup analysis, meta-regression). | Line 149-151 |
|  | 13f | Describe any sensitivity analyses conducted to assess robustness of the synthesized results. | Line 139-148,  Appendix 3.1 |
| Reporting bias assessment | 14 | Describe any methods used to assess risk of bias due to missing results in a synthesis (arising from reporting biases). | Line 110-114,  Appendix 3.2 |
| Certainty assessment | 15 | Describe any methods used to assess certainty (or confidence) in the body of evidence for an outcome. | Line 154-166,  Appendix 3.5 |
| **RESULTS** | | |  |
| Study selection | 16a | Describe the results of the search and selection process, from the number of records identified in the search to the number of studies included in the review, ideally using a flow diagram. | Line169-172, 426-427 |
|  | 16b | Cite studies that might appear to meet the inclusion criteria, but which were excluded, and explain why they were excluded. | Line 426-427 |
| Study characteristics | 17 | Cite each included study and present its characteristics. | Line 172-175,  Appendix 4 |
| Risk of bias in studies | 18 | Present assessments of risk of bias for each included study. | Line 176-180,  Appendix 5 |
| Results of individual studies | 19 | For all outcomes, present, for each study: (a) summary statistics for each group (where appropriate) and (b) an effect estimate and its precision (e.g. confidence/credible interval), ideally using structured tables or plots. | Line 181-232,  Appendix 6 |
| Results of syntheses | 20a | For each synthesis, briefly summarise the characteristics and risk of bias among contributing studies. | Line 172-211,  Appendix 5 |
|  | 20b | Present results of all statistical syntheses conducted. If meta-analysis was done, present for each the summary estimate and its precision (e.g. confidence/credible interval) and measures of statistical heterogeneity. If comparing groups, describe the direction of the effect. | Line 182-267,  Appendix 6 |
|  | 20c | Present results of all investigations of possible causes of heterogeneity among study results. | Line 268-274,  Appendix 6.5, 9 |
|  | 20d | Present results of all sensitivity analyses conducted to assess the robustness of the synthesized results. | Line 274-276,  Appendix 10 |
| Reporting biases | 21 | Present assessments of risk of bias due to missing results (arising from reporting biases) for each synthesis assessed. | Appendix 5 |
| Certainty of evidence | 22 | Present assessments of certainty (or confidence) in the body of evidence for each outcome assessed. | Line 190-232,  Appendix 8 |
| **DISCUSSION** | | |  |
| Discussion | 23a | Provide a general interpretation of the results in the context of other evidence. | Line 278-374 |
|  | 23b | Discuss any limitations of the evidence included in the review. | Line 375-382 |
|  | 23c | Discuss any limitations of the review processes used. | Line 375-382 |
|  | 23d | Discuss implications of the results for practice, policy, and future research. | Line 278-286, 368-374 |
| **OTHER INFORMATION** | | |  |
| Registration and protocol | 24a | Provide registration information for the review, including register name and registration number, or state that the review was not registered. | Line 74-77 |
|  | 24b | Indicate where the review protocol can be accessed, or state that a protocol was not prepared. | Non protocol |
|  | 24c | Describe and explain any amendments to information provided at registration or in the protocol. | Non protocol |
| Support | 25 | Describe sources of financial or non-financial support for the review, and the role of the funders or sponsors in the review. | Line 395-397 |
| Competing interests | 26 | Declare any competing interests of review authors. | Line 393-394 |
| Availability of data, code and other materials | 27 | Report which of the following are publicly available and where they can be found: template data collection forms; data extracted from included studies; data used for all analyses; analytic code; any other materials used in the review. | Line 391-392 |

### Appendix 2: Search strategy

**PubMed**

#1"Obesity"[Mesh]

249,138 results

#2"Weight Loss"[Mesh]

48,064 results

#3"Weight Loss"[MeSH] OR "loss weight"[Title/Abstract] OR "losses weight"[Title/Abstract] OR "weight losses"[Title/Abstract] OR "weight reduction"[Title/Abstract] OR "reduction weight"[Title/Abstract] OR "reductions weight"[Title/Abstract] OR "weight reductions"[Title/Abstract]

56,321 results

#4Overweight[Mesh]

260,037 results

#5"obes*"[Title/Abstract] OR "body mass ind*"[Title/Abstract] OR "adiposity"[Title/Abstract] OR "overweight"[Title/Abstract] OR "over weight"[Title/Abstract] OR "overload syndrome*"[Title/Abstract] OR "over eat*"[Title/Abstract] OR "overfeed*"[Title/Abstract] OR "over feed*"[Title/Abstract] OR "overfed"[Title/Abstract] OR "over fed"[Title/Abstract] OR "weight cycling"[Title/Abstract] OR "skinfold thickness"[Title/Abstract] OR "antiobesity"[Title/Abstract] OR "anti-obesity"[Title/Abstract] OR "obesitas"[Title/Abstract] OR "bodyweight"[Title/Abstract] OR "body weight"[Title/Abstract]

729,745 results

#6 #1 OR #3 OR #4 OR #5

789,952 results

#7 Orlistat[MeSH]

1,342 results

#8"Orlistat"[MeSH] OR "Alli"[Title/Abstract] OR "orlipastat"[Title/Abstract] OR "Orlistat"[Title/Abstract] OR "Ro-18-0647"[Title/Abstract] OR "Ro 18 0647"[Title/Abstract] OR "Tetrahydrolipstatin"[Title/Abstract] OR "Tetrahydrolipastatin"[Title/Abstract] OR "THLP"[Title/Abstract] OR "Xenical"[Title/Abstract] OR "Xenical"[Title/Abstract]

2,450 results

#9"lorcaserin"[Title/Abstract] OR "lorqess"[Title/Abstract] OR "Belviq"[Title/Abstract] OR "APD356"[Title/Abstract] OR "APD-356"[Title/Abstract] OR "AR-10A"[Title/Abstract]

449 results

#10("phentermine"[Title/Abstract] AND "topiramate"[Title/Abstract]) OR "phentermine-topiramate"[Title/Abstract] OR"phenterminetopiramate"[Title/Abstract] OR "qnexa"[Title/Abstract] OR "qsiva"[Title/Abstract] OR "Qsymia"[Title/Abstract] OR "topiramatephentermine"[Title/Abstract] OR "phentermine/topiramate"[Title/Abstract]

292 results

#11"bupropion-naltrexone"[Title/Abstract] OR "Contrave"[Title/Abstract] OR "bupropion/naltrexone"[Title/Abstract] OR ("amfebutamone"[Title/Abstract] AND "naltrexone"[Title/Abstract]) OR ("bupropion"[Title/Abstract] AND "naltrexone"[Title/Abstract])

325 results

#12"Glucagon-Like Peptide 1"[MeSH] OR "Glucagon Like Peptide 1"[Title/Abstract] OR "GLP-1"[Title/Abstract] OR "GLP-1"[Title/Abstract] OR "Glucagon-Like Peptide 1"[Title/Abstract]

20,086 results

#13"Liraglutide"[MeSH] OR "liraglutide"[Title/Abstract] OR "nn 2211"[Title/Abstract] OR "nn2211"[Title/Abstract] OR "nnc 90 1170"[Title/Abstract] OR "Saxenda"[Title/Abstract] OR "victoza"[Title/Abstract]

3759 results

#14"albiglutide"[Title/Abstract] OR "Tanzeum"[Title/Abstract] OR "dulaglutide"[Title/Abstract] OR "Trulicity"[Title/Abstract] OR "exenatide"[Title/Abstract] OR "Byetta"[Title/Abstract] OR "Extended-release exenatide"[Title/Abstract] OR "Bydureon"[Title/Abstract] OR "lixisenatide"[Title/Abstract] OR "Adlyxin"[Title/Abstract] OR "semaglutide"[Title/Abstract] OR "Ozempic"[Title/Abstract] OR "Rybelsus"[Title/Abstract]

3,664 results

#15 "Sodium-Glucose Transporter 2 Inhibitors"[Mesh] OR (Sodium Glucose Transporter 2 Inhibitors[Title/Abstract]) OR (SGLT-2 Inhibitors[Title/Abstract] OR (SGLT 2 Inhibitors[Title/Abstract]) OR (SGLT2 Inhibitors[Title/Abstract] OR (Sodium-Glucose Transporter 2 Inhibitor[Title/Abstract]) OR (Sodium Glucose Transporter 2 Inhibitor[Title/Abstract]) OR (SGLT2 Inhibitor[Title/Abstract]) OR (Inhibitor, SGLT2[Title/Abstract]) OR (Gliflozins[Title/Abstract]) OR (Gliflozin[Title/Abstract]) OR (SGLT-2 Inhibitor[Title/Abstract]) OR (Inhibitor, SGLT-2[Title/Abstract]) OR (SGLT 2 Inhibitor[Title/Abstract])

7,186 results

#16 ertugliflozin[Title/Abstract] OR Steglatro[Title/Abstract] OR canagliflozin[Title/Abstract] OR Invokana[Title/Abstract] OR empagliflozin[Title/Abstract] OR Jardiance[Title/Abstract] OR dapagliflozin[Title/Abstract] OR Farxiga[Title/Abstract] OR ipragliflozin[Title/Abstract] OR luseogliflozin[Title/Abstract] OR "remogliflozin etabonate"[Title/Abstract] OR (remogliflozin[Title/Abstract] AND etabonate[Title/Abstract]) OR "sergliflozin etabonatem"[Title/Abstract] OR (sergliflozin[Title/Abstract] AND etabonatem[Title/Abstract]) OR tofogliflozin[Title/Abstract]

5,082 results

#17 "Metformin"[Mesh] OR metformin[Title/Abstract] OR Glumetza[Title/Abstract] OR "Glucophage XR"[Title/Abstract] OR Fortamet[Title/Abstract] OR Glucophage[Title/Abstract] OR Riomet[Title/Abstract] OR "metformin ER"[Title/Abstract] OR "metformin IR"[Title/Abstract] ) OR Dimethylbiguanidine[Title/Abstract] OR Dimethylguanylguanidine[Title/Abstract]

27,777 results

#18 phentermine[Title/Abstract] OR Adipex-P[Title/Abstract] OR Lomaira[Title/Abstract] OR Suprenza[Title/Abstract] OR phendimetrazine[Title/Abstract] OR Bontril[Title/Abstract] OR Melfiat[Title/Abstract] OR benzphetamine[Title/Abstract] OR Didrex[Title/Abstract] OR Regimex[Title/Abstract] OR diethylpropion[Title/Abstract] OR Tenuate[Title/Abstract] OR "Tenuate Dospan"[Title/Abstract]

1,851 results

#19 "Carnitine"[Mesh] OR carnitine[Title/Abstract] OR levocarnitine[Title/Abstract] OR l-carnitine[Title/Abstract] OR "(L-)carnitine"[Title/Abstract] OR "L Carnitine"[Title/Abstract] OR Levocarnitine[Title/Abstract] OR Vitamin BT[Title/Abstract] OR L-Carnitine[Title/Abstract] OR Bicarnesine[Title/Abstract]

19,155 results

#20 pramlintide[Title/Abstract] OR symlin[Title/Abstract] OR “AC 0137” [Title/Abstract] OR “pramlintide acetate” [Title/Abstract]

394 results

#21 "Pediatric Obesity/drug therapy"[Mesh]

103 results

#22 #8 OR #9 OR #10 OR #11 OR #12 OR #13 OR #14 OR #15 OR #16 OR #17 OR #18 OR #19 OR #20 OR #21

77,205 results

#23 "Randomized Controlled Trial"[Publication Type]

578,364 results

#24 "Randomized Controlled Trials as Topic"[MeSH]

161,499 results

#25 "randomized controlled study"[Title/Abstract] OR "randomized controlled trial"[Title/Abstract] OR "randomized study"[Title/Abstract] OR "randomized trial"[Title/Abstract] OR "randomized placebo-controlled study"[Title/Abstract] OR "randomized placebo-controlled trial"[Title/Abstract] OR "randomized placebo controlled"[Title/Abstract] OR "randomized placebo-controlled"[Title/Abstract] OR "randomized double-blin*"[Title/Abstract] OR "randomized double blin*"[Title/Abstract] OR (randomized[Title/Abstract] AND double-blin*[Title/Abstract]) OR (randomized[Title/Abstract] AND placebo-controlled[Title/Abstract])

278,408 results

#26 #23 OR #24 OR #25

797,526 results

#27 ("Child"[Mesh]) OR (Children[Title/Abstract])

2,431,226 results

#28 "Adolescent"[MeSH] OR "Adolescents"[Title/Abstract] OR "Adolescence"[Title/Abstract] OR "Teens"[Title/Abstract] OR "Teen"[Title/Abstract] OR "Teenagers"[Title/Abstract] OR "Teenager"[Title/Abstract] OR "Youth"[Title/Abstract] OR "Youths"[Title/Abstract] OR "adolescents female"[Title/Abstract] OR "adolescent female"[Title/Abstract] OR "female adolescent"[Title/Abstract] OR "female adolescents"[Title/Abstract] OR "adolescents male"[Title/Abstract] OR "adolescent male"[Title/Abstract] OR "male adolescent"[Title/Abstract] OR "male adolescent"[Title/Abstract]

2,276,681 results

#29 #27 OR #28

3,646,506 results

#30 #6 AND #22 AND #26 AND #29

629 results + 25 results

**Embase**

#1 pediatric obesity.mp. or childhood obesity/	21562

#2 adolescent obesity/	3578

#3 pediatric.mp. or pedlatrics/	609316

#4 child. mp. or child/	2554555

#5 children. mp. 1489403

#6 adalescent/	1696622

7# obesity/ or obesity.mp. 646457

#8 childhoad obesity intervention	113

#9 overwelghtmp. or obesity/	521948

#10 drug therapy/	880140

#11 orlistat.mp. or tetrahydrolipstatin/	7357

#12 phentermine plus topiramate.mp. or phentermine plus topiramate/	806

#13 Glucagon-Like Peptide 1 receptor agonists.mp. or glucagon like peptide 1 receptor agonist/	8908

#14 liraglutide/ or liraglutide.mp. 11621

#15 semaglutide.mp. or semaglutlde/	2725

#16 albiglutide.mp. or albiglutide/	1290

#17 dulaglutide.mp. or dulaglutide/	2300

#18 exenatide.mp. or exendin 4/	11794

#19 lixisenatide/ or lixisenatide.mp. 2069

#20 Sodium-Glucose Transporter 2 Inhibitors. mp. or sodium glucose cotransporter 2 inhibitor/	9350

#21 ertugliflozin/ or ertugliflozin.mp. 913

#22 canagliflozin.mp. or canagliflozin/	4856

#23 empaglifiozin.mp. or empagliflozin/	6718

#24 dapagliflozin.mp. or dapagliflozin/	6639

#25 ipragliflozin.mp. or ipraglifiozin/	828

#26 luseogliflozin.mp. or luseoglifiozin/	517

#27 remogliflozin etabonate.mp. or remogliflozin etabonate/	204

#28 sergliflozin etabonate.mp. or sergliflozin etabonate/	99

#29 tofogliflozin.mp. or tofogliflozin/	488

#30 metformin.mp. or metformin/	81970

#31 pramlintide.mp. or pramlintide /	1634

#32 carnitine.mp. or carnitine/	31227

#33 Randomized Controlled Trial.mp. or randomized controlled trial/	979664

#34 double blind procedure.mp. or double blind procedure/	199236

#35 ("randomized controlled study" or "randomized controlled trial" or " randomized study " or " randomized trial " or " randomized placebo-controlled study or " randomized placebo-controlled trial " or " randomized placebo controlled " or " randomized placebo-controlled " or " randomized double-blin " or " randomized double blin* " or (randomized and double-blin*) or (randomized and placebo-controlled)). ti, ab. 385073

#36 3 or 4 or 5 or 6	3749880

#37 7 or 9	656851

#38 36 or 37	119209

#39 1 or 2 or 8 or 38	121239

#40 10 or 11 or 12 or 13 or 14 or 15 or 16 or 17 or 18 or 19 or 20 or 21 or 22 or 23 or 24 or 25 or 26 or 27 or 28 or 29 or 30 or 31 or 32	1014700

#41 33 or 34 or 35	1100074

#42 39 and 40 and 41	552

552 results +25 results = 577 results

**The Cochrane library (CENTRAL)**

ID Search Hits

#1 MeSH descriptor: [Pediatric Obesity] explode all trees	1536

#2 adolescent	142445

#3 child	187088

#4 MeSH descriptor: [Obesity] explode all trees	16001

#5 MeSH descriptor: [Child] explode all trees	61856

#6 MeSH descriptor: [Adolescent] explode all trees	110535

#7 MeSH descriptor: [Overweight] explode all trees	19130

#8 obesity	46545

#9 overweight	19902

#10 #2 OR #3 OR #5 OR #6	276539

#11 #4 OR #7 OR #8 OR #9	51988

#12 #10 AND #11	11202

#13 #1 OR #12	11289

#14 MeSH descriptor: [Orlistat] explode all trees	295

#15 orlistat	610

#16 metformin	12296

#17 MeSH descriptor: [Metformin] explode all trees	4526

#18 phentermine	242

#19 MeSH descriptor: [Phentermine] explode all trees	125

#20 MeSH descriptor: [Drug Therapy] explode all trees	148318

#21 phentermine and topiramate	114

#22 MeSH descriptor: [Glucagon-Like Peptide 1] explode all trees	1970

#23 MeSH descriptor: [Liraglutide] explode all trees	809

#24 liraglutide	2170

#25 semaglutide	749

#26 albiglutide	145

#27 dulaglutide	470

#28 exenatide	1298

#29 MeSH descriptor: [Exenatide] explode all trees	590

#30 lixisenatide	345

#31 MeSH descriptor: [Sodium-Glucose Transporter 2 Inhibitors] explode all trees	536

#32 ertugliflozin	174

#33 canagliflozin	680

#34 MeSH descriptor: [Canagliflozin] explode all trees	264

#35 empagliflozin	1436

#36 dapagliflozin	1554

#37 ipragliflozin	163

#38 luseogliflozin	100

#39 remogliflozin etabonate	24

#40 tofogliflozin	94

#41 pramlintide	164

#42 carnitine	1966

#43 levocarnitine	108

#44 #14 OR #15 OR #16 OR #17 OR #18 OR #19 OR #20 OR #21 OR #22 OR #23 OR #24 OR #25 OR #26 OR #27 OR #28 OR #29 OR #30 OR #31 OR #32 OR #33 OR #34 OR #35 OR #36 OR #37 OR #38 OR #39 OR #40 OR #41 OR #42 OR #43	168043

#45 “randomized controlled study” OR “randomized controlled trial” OR “randomized study” OR “randomized trial” OR “randomized placebo-controlled study” OR “randomized placebo-controlled trial” OR “randomized placebo controlled” OR “randomized placebo-controlled” OR “randomized double-blin*” OR “randomized double blin*”	1042978

#46 MeSH descriptor: [Randomized Controlled Trials as Topic] explode all trees	15211

#47 #45 OR #46	1044622

#48 #13 AND #44 and #47	in Trials 658

658 result + 69 results=727 results

### Appendix 3: Supplementary of methods

#### 3.1 Details of the sensitivity analyses

We conducted multiple sensitivity analyses to assess the robustness of the final results, including:

1) Estimating all outcomes by the Bayesian method. Outcomes were performed in a Bayesian framework using the Markov Chain Monte Carlo method with the random-effect consistency model. Three Markov chains were established for generating 250,000 sample iterations with an initial burn-in period of 50,000 iterations to obtain aposterior distribution. The Brooks-Gelman-Rubin method was used to test the convergence of the iterations [1]. The interventions were ranked according to their posteriori probabilities and the surface under the Cumulative Ranking (SUCRA) values were calculated.

2) Exclusion of studies with fewer than 40 participants.

3) Exclusion of studies without a placebo-control.

4) Exclusion of studies that did not report BMI change from baseline.

5) Exclusion of studies with treatment duration < 3 months.

6) Exclusion of the high risk of bias studies.

#### 3.2 Details of risk of bias sssessment

Risk of bias (ROB) for all included randomised controlled trials was assessed by two independent assessors (LL and SZ) with the Cochrane Risk of Bias 2 (ROB-2) tool [2], disagreements were resolved by discussion with a third assessor. This tool evaluates 5 domains of bias: D1, the randomization process; D2, deviations from intended interventions; D3, missing outcome data; D4, measurement of the outcome; D5, selection of the reported result. Each question in the 5 domains was answered by yes, probably yes, no, probably no, and information. Based on these answers, the ROB of each domain and overall were judged as low, some concerns, or high [3].

#### 3.3 Details of absolute effect calculation

The baseline risk (P_0_) is the event rate in the control group. We simulated the baseline risk by using the pooled effect of the control group (lifestlye modification alone group) through the random-effect single-arm meta-analysis [4]. The absolute risk difference (RD) of the intervention versus lifestlye modification alone was estimated from the relative effect and the estimate of baseline risk, with the following formula [5]:

$$RD=\frac{(OR-1)(P_{0}-1)P_{0}}{P_{0}*OR-P_{0}+1}$$

$$P_{1}=\frac{P_{0}*OR}{P_{0}(OR-1)+1}$$

Where *P_0_* represents baseline risk, *P_1_* represents the risk after intervention, and *OR* represents relative effect.

#### 3.4 Details of minimal important difference

The minimum important difference (MID) is the smallest value of change that patients consider beneficial and can lead to modifications in patient management plans. We identified the MID of the primary continuous outcomes, including change in BMI from baseline and change in weight from baseline, by searching through previous research studies. The estimated values of the lifestyle intervention group were used as MID for weight loss, which was -4 kg, and the MID for BMI change, which was -1.4 kg/m^2^[6].

We consider that achieving a 5% weight loss is equivalent in difficulty to reducing BMI by 5%. Therefore, we followed the FDA-defined weight loss criteria to define the MID threshold for the proportion of participants achieving BMI reduction of at least 5% (i.e., achieving a reduction in BMI ≥ 5% being approximately double the proportion in the control group)[7].

#### 3.5 Details of GRADE Minimal Contextualized Framework and the presentation tool

The primary outcomes were assessed according to the previously determined minimal important differences to determine the level of classification. We established four categories of effect class: “among the best”, “intermediate-possibly better”, “intermediate-possibly worse”, and “among the worst”.

For the outcome of change in BMI from baseline and change in weight from baseline, the “among the best” effct class was defined if the upper limit of the 95% confidence interval for the effect estimate was smaller than the MID. Conversely, if the lower limit of the confidence interval was greater than the MID, it was defined as “among the worst”. When the 95% confidence interval intersected with the MID, and the estimated value was to the left of the MID, it was defined as “intermediate-possibly better”. If the estimated value was to the right of the MID, it was defined as “intermediate-possibly worse”.

For the proportion of participants achieving BMI reduction of at least 5%, the “among the best” effct class was defined if the lower limit of the 95% confidence interval for the absolute effect estimate was greater than the MID. If the upper limit of the confidence interval was smallerr than the MID, it was defined as “among the worst”. When the 95% confidence interval intersected with the MID, and the estimate of absolute effect was to the right of the MID, it was defined as “intermediate-possibly better”. If the estimated value was to the left of the MID, it was defined as “intermediate-possibly worse”.

For the outcome of discontinuation due to adverse events and total gastrointestinal events, we set three categories of effect class: “among the worst”, “intermediate”, “among the best”.

If there was no difference compared with the control group, it was defined as “among the best”. The “intermediate” effct class was defined when the effect was worse than the control group but no worse than other drugs. If the effect was significantly worse compared to the control group and any of the other drugs, it is defined as "among the worst".

### Appendix 4: Characteristics of the included studies

#### 4.1 Summary of included studies

| Study | Register number | Location | Intervention | Sample size | Follow-up |
| --- | --- | --- | --- | --- | --- |
| Ozkan 2004 | - | Turkey | Orlistat (120 mg/tid) | 22 | 11.7±3.7 months |
|  |  |  | Lifestyle modification alone | 20 | 10.2±3.7 months |
| Chanoine 2005 | - | United States, Canada | Orlistat (120 mg/tid) | 352 | 52 weeks |
|  |  |  | Placebo | 181 |  |
| Yu 2013 | CUHK_CCT00135 | Hong kong | Diet+Orlistat (120 mg/tid)+Exercise | 23 | 10 weeks |
|  |  |  | Diet+Orlistat (120 mg/tid) | 21 |  |
|  |  |  | Diet alone | 20 |  |
| Maahs 2006 | - | United States | Orlistat (120 mg/tid) | 20 | 6 months |
|  |  |  | Placebo | 20 |  |
| Shankar 2022 | NCT01485614 | 26 countries | Sitagliptin (100 mg/day) | 95 | 54 weeks |
|  |  |  | Placebo/Metformin | 95 |  |
| Fox 2016 | NCT01859013 | United States | Topiramate (75 mg/day) | 16 | 24 weeks |
|  |  |  | Placebo | 14 |  |
| Barrientos 2022 | NCT02803918 | United States, Mauritius, Mexico, South Africa, Spain, Turkey | Lixisenatide (5~20 μg /day) | 18 | 6 weeks |
|  |  |  | Placebo | 5 |  |
| Hsia 2020 | NCT02714062 | United States | Phentermine/Topiramate 15 mg/92 mg/day | 12 | 8 weeks |
|  |  |  | Phentermine/Topiramate 7.5 mg/46 mg/day | 15 |  |
|  |  |  | Placebo | 13 |  |
| Arslanian 2022 | NCT02963766 | United States, Brazil, France, Germany, Hungary, India, Mexico, Puerto Rico, Saudi Arabia, Turkey, United Kingdom | Dulaglutide 1.5 mg/once weekly | 52 | 26 weeks(trial)+26 weeks(follow-up after trial |
|  |  |  | Dulaglutide 0.75 mg/once weekly | 51 |  |
|  |  |  | Placebo | 51 |  |
| Kelly 2020 | NCT02918279 | United States, Sweden, Russian Federation, Mexico, Belgium | Liraglutide (3.0 mg/day) | 125 | 56 weeks(trial)+26 weeks(follow-up after trial) |
|  |  |  | Placebo | 126 |  |
| Mastrandrea 2019 | NCT02696148 | United States | Liraglutide (0.3~3.0 mg/day) | 16 | 7~8 weeks |
|  |  |  | Placebo | 8 |  |
| Danne 2017 | NCT01789086 | Germany | Liraglutide（0.6 mg~3.0mg/day） | 14 | 5 weeks |
|  |  |  | Placebo | 7 |  |
| Tamborlane 2019 | NCT01541215 | Global | Liraglutide (0.6~1.8 mg/day)+Metformin | 66 | 26 weeks(trial)+26 weeks(follow-up after trial) |
|  |  |  | Placebo+Metformin | 68 |  |
| Fox 2022 | NCT02496611 | United States | Exenatide (2mg/once weekly) | 33 | 52 weeks |
|  |  |  | Placebo | 33 |  |
| Kelly 2012 | NCT00886626 | United States | Exenatide (5mcg~10mcg/bid) | 5 | 3 months |
|  |  |  | Placebo | 6 |  |
| Weghuber 2020 | 2015-001628-45 | Sweden, Austria | Exenatide (2mg/once weekly) | 22 | 6 months |
|  |  |  | Placebo | 22 |  |
| Atabek 2008 | - | Turkey | Metformin (500mg/bid) | 90 | 6 months |
|  |  |  | Placebo | 30 |  |
| Kelsey 2021 | NCT01775813 | United States | Metformin(1000mg/bid) | 20 | 2.4±0.6 years |
|  |  |  | Placebo | 24 |  |
| Nadeau 2009 | - | United States | Metformin (850mg bid) | 37 | 6 months |
|  |  |  | Placebo | 13 |  |
| Evia-Viscarra 2012 | NCT01410604 | Mexico | Metformin (1g/day) | 12 | 3 months |
|  |  |  | Placebo | 14 |  |
| Burgert 2008 | NCT00667498 | United States | Metformin (1500 mg/day) | 15 | 4 months |
|  |  |  | Placebo | 13 |  |
| Mauras 2012 | NCT00139477 | United States | Metformin (500-1000 mg/bid) | 35 | 6 months |
|  |  |  | Lifestyle modification alone | 31 |  |
| Warnakulasuriya 2018 | NCT02274948 | Sri Lanka | Metformin (500-1000 mg/bid) | 82 | 12 months |
|  |  |  | Placebo | 68 |  |
| Kendall 2013 | ISRCTN 19517475 | Britain | Metformin (1500 mg/day) | 74 | 6 months |
|  |  |  | Placebo | 77 |  |
| Clarson 2009 | - | Canada | Metformin (1500 mg/day) | 11 | 6 months |
|  |  |  | Lifestyle modification alone | 14 |  |
| Pastor-Villaescusa 2017 | - | Spain | Metformin (Prepubertal)500mg/bid | 40 | 6 months |
|  |  |  | Placebo | 40 |  |
|  |  |  | Metformin (Pubertal) | 40 |  |
|  |  |  | Placebo | 40 |  |
| Wilson 2010 | NCT00120146 | United States | Metformin (750 mg/bid) | 39 | 48 weeks(trial)+48 weeks(follow-up after trial) |
|  |  |  | Placebo | 38 |  |
| Wiegand 2010 | Eudra-CT-Nr 2004-003816-47 | Germany, Switzerland | Metformin (500 mg/bid) | 36 | 6 months |
|  |  |  | Placebo | 34 |  |
| MP van der Aa 2016 | NCT01487993 | Netherlands | Metformin (2000 mg/day) | 23 | 18 months |
|  |  |  | Placebo | 19 |  |
| Rynders 2012 | NCT00139477 | United States | Metformin (1000 mg/day) | 7 | 6 months |
|  |  |  | Lifestyle modification alone | 9 |  |
| Yanovski 2011 | NCT00005669 | United States | Metformin (500~1000 mg bid) | 53 | 6 months(trial)+6 months(follow-up after trial) |
|  |  |  | Placebo | 47 |  |
| Bassols 2019 | Eudra-CT-Nr r: 2010-024414-61 | Spain | Metformin (850 mg/day) | 9 | 24 months |
|  |  |  | Placebo | 9 |  |
| Garibay-Nieto 2017 | NCT02063802 | Mexico | Metformin (1000 mg/day) | 14 | 4 months |
|  |  |  | Placebo | 17 |  |
|  |  |  | Conjugated Linoleic Acid(3g/day) | 17 |  |
| Kay 2001 | - | United States | Metformin (850 mg bid) | 12 | 8 weeks |
|  |  |  | Placebo | 12 |  |
| Rezvanian 2010 | - | Iran | Metformin (1500 mg/day) | 45 | 12 weeks(trial)+12 weeks(with drug follow-up after trial) |
|  |  |  | fluoxetine (20 mg/day) | 45 |  |
|  |  |  | Combination | 45 |  |
|  |  |  | Placebo | 45 |  |
| Li 2019 | - | China | Metformin (250mg/500mg/tid) | 42 | 6 months |
|  |  |  | Lifestyle modification alone | 42 |  |
| Vendrell 2006 | - | Canada | Metformin (75mg/bid) | 11 | 12 weeks |
|  |  |  | Placebo | 11 |  |
| Daniel 2022 | NCT04102189 | United States, Austria, Belgium, Croatia, Ireland, Mexico, Russian Federation, United Kingdom | Semaglutide (2.4mg/once weekly) | 134 | 68 weeks+7 weeks  (follow-up after trial) |
|  |  |  | Placebo | 67 |  |
| Kelly 2022 | NCT03922945 | United States | Phentermine/Topiramate 15 mg/92 mg/day | 113 | 56 weeks |
|  |  |  | Phentermine/Topiramate 7.5 mg/46 mg/day | 54 |  |
|  |  |  | Placebo | 56 |  |
| Diene 2022 | NCT02527200 | United States, Australia, Canada, France, Italy, Netherlands, New Zealand, Turkey | Liraglutide (0.6 mg~3mg) (Adolescents) | 19 | 52 weeks(16 weeks trial + 36 weeks open-label period + 2-week off-drug follow-up) |
|  |  |  | Placebo | 12 |  |
|  |  |  | Liraglutide (0.6 mg~3mg) (Children) | 17 |  |
|  |  |  | Placebo | 7 |  |
| Yanovski 2012 | NCT00001723 | United States | Orlistat (120 mg/tid) | 100 | 6 months |
|  |  |  | Placebo | 100 |  |
| Hoeger 2008 | NCT00283816 | United States | Metformin (2000 mg/day) | 18 | 24 weeks |
|  |  |  | Placebo | 18 |  |

#### 4.2 Baseline characteristics of included studies

| Study | Intervention | Sample size | Age (y) | Male(n, %) | BMI (kg/m^2^) | Weight (kg) |
| --- | --- | --- | --- | --- | --- | --- |
| Ozkan 2004 | Orlistat | 22 | 12.9±2.4 | 5(33%) | 32.5 | 82.1±20.9 |
|  | Lifestyle modification alone | 20 | 12.5±2.2 | 5(33%) | 31.2 | 73.9±15.3 |
| Chanoine 2005 | Orlistat | 352 | 13.6±1.3 | 124(35%) | 35.7±4.2 | 97.7±15.0 |
|  | Placebo | 181 | 13.5±1.2 | 52(29%) | 35.4±4.1 | 95.1±14.2 |
| Yu 2013 | Diet+Orlistat+Exercise | 23 | 11~18 yr | 17(73.9%) | 33.2 (30.0 to 35.4) | 90.2 (82.4 to 99.2) |
|  | Diet+Orlistat | 21 |  | 14(66.7%) | 31.4 (27.8 to 34.0) | 81.7 (75.1 to 90.6) |
|  | Diet alone | 20 |  | 14(70%) | 30.4 (27.9 to 34.3) | 81.3 (73.2 to 97.8) |
| Maahs 2006 | Orlistat | 20 | 15.8±1.5 | 12(60%) | 39.2±5.3 | 111.1±22.9 |
|  | Placebo | 20 | 15.8±1.4 | 15(75%) | 41.7±11.7 | 114.3±38.4 |
| Shankar 2022 | Sitagliptin | 95 | 14.3±2.0 | 41(43.2%) | 33.3±7.7 | 89.1±25.3 |
|  | Placebo/Metformin | 95 | 13.7±1.9 | 34(35.8%) | 31.2±7.7 | 81.9±24.8 |
| Fox 2016 | Topiramate | 16 | 14.9±1.6 | 6(37.5%) | 41.0±5.0 | 117±23.5 |
|  | Placebo | 14 | 15.7±1.8 | 5(35.7%) | 39.5±4.0 | 112±15.3 |
| Barrientos 2022 | Lixisenatide | 18 | 15.6±1.0 | 5(27.8%) | 33.2±4.8 | 91.3±18.8 |
|  | Placebo | 5 | 15.4±1.5 | 2(40%) | 37.4±3.6 | 98.0±14.7 |
| Hsia 2020 | Phentermine/Topiramate 15 mg/92 mg/day | 12 | 12~17 yr | - | - | 99.4±22.6 |
|  | Phentermine/Topiramate 7.5 mg/46 mg/day | 15 |  | - | - | 97.8±25.1 |
|  | Placebo | 13 |  | - | - | 111.3±23.4 |
| Arslanian 2022 | Dulaglutide 1.5 mg/once weekly | 52 | 14.7±1.8 | 18(35%) | 34.3±7.0 | 92.6±21.6 |
|  | Dulaglutide 0.75 mg/once weekly | 51 | 14.7±2.2 | 16(31%) | 33.6±9.0 | 90.0±28.3 |
|  | Placebo | 51 | 14.2±2.1 | 10(20%) | 34.3±10.2 | 88.9±29.4 |
| Kelly 2020 | Liraglutide | 125 | 14.6±1.6 | 54(43.2%) | 35.3±5.1 | 99.3±19.7 |
|  | Placebo | 126 | 14.5±1.6 | 48(38.1%) | 35.8±5.7 | 102.2±21.6 |
| Mastrandrea 2019 | Liraglutide | 16 | 9.7±1.1 | 8(50.0%) | - | 66.6±12.6 |
|  | Placebo | 8 | 10.4±1.1 | 7(87.5%) | - | 81.4±16.6 |
| Danne 2017 | Liraglutide | 14 | 15.1±0.9 | 3(21.4%) | 36.5±3.7 | 103.5±12.8 |
|  | Placebo | 7 | 14.4±1.8 | 4(57.1%) | 35.7±5.4 | 109.6±30.8 |
| Tamborlane 2019 | Liraglutide+Metformin | 66 | 14.6±1.7 | 25(37.9%) | 34.6±10.9 | 93.3±31.0 |
|  | placebo+Metformin | 68 | 14.6±1.7 | 26(38.2%) | 33.3±7.4 | 89.8±22.1 |
| Fox 2022 | Exenatide | 33 | 15.9±1.6 | 15(45%) | 36.5±4.3 | 105.6±17.7 |
|  | Placebo | 33 | 16.1±1.5 | 20(61%) | 37.3±4.6 | 111.4±17.2 |
| Kelly 2012 | Exenatide | 5 | 13.0±1.87 | 0 | 37.4±4.18 | 94.4±17.2 |
|  | Placebo | 6 | 12.5±2.43 | 2(33.3%) | 36.2±5.59 | 93.3±24.7 |
| Weghuber 2020 | Exenatide | 22 | 14.5±2.3 | 9(41%) | 36.0±4.8 | 106.2±19.7 |
|  | Placebo | 22 | 13.5±2.3 | 13(59%) | 36.2±5.0 | 102.5±24.5 |
| Atabek 2008 | Metformin | 90 | 11.83±2.8 | 45(50%) | 28.5±3.4 | 67.16±16.8 |
|  | Placebo | 30 | 11.6±2.7 | 15(50%) | 28.0±3.4 | 66.27±16.9 |
| Kelsey 2021 | Metformin | 20 | 11.9±1.6 | 11(55.0%) | - | - |
|  | Placebo | 24 | 11.1±1.1 | 7(29.2%) | - | - |
| Nadeau 2009 | Metformin | 37 | 15.1 (range 12-18 yr) | 12(32%) | 39.6±0.98 | - |
|  | Placebo | 13 |  | 5(38%) | 40.2±1.8 | - |
| Evia-Viscarra 2012 | Metformin | 12 | 12.65±1.98 | 3(25%) | 33.44±5.82 | 81.21±18.74 |
|  | Placebo | 14 | 14.12±1.21 | 6(42.9%) | 32.82±6.37 | 86.26±26.02 |
| Burgert 2008 | Metformin | 15 | 15±2 | 5(33.3%) | 41±6 | 115±19 |
|  | Placebo | 13 | 15±1 | 4(30.8%) | 40±6 | 106±15 |
| Mauras 2012 | Metformin | 35 | 12.3±0.5 | 15(42.9%) | 32.0±1.0 | - |
|  | Lifestyle modification alone | 31 | 12.0±0.4 | 15(48.4%) | 33.2±0.7 | - |
| Warnakulasuriya 2018 | Metformin | 82 | 12.25±2.27 | 44(53.7%) | 27.44±2.7 | 63.44±15.5 |
|  | Placebo | 68 | 11.94±2.17 | 40(58.8%) | 27.44±2.96 | 63.51±14.37 |
| Kendall 2013 | Metformin | 74 | 13.68±2.3 | 25(33.8%) | 37.10±6.35 | 100.3±24.1 |
|  | Placebo | 77 | 13.64±2.2 | 24(31.2%) | 35.95±6.32 | 96.4±21.8 |
| Clarson 2009 | Metformin | 11 | 13.1 (range 10.1-16.1 yr) | 4(36.4%) | 36.4±1.8 | - |
|  | Lifestyle modification alone | 14 |  | 9(64.3%) | 33.9±1.1 | - |
| Pastor-Villaescusa 2017 | Metformin（Prepubertal） | 40 | 6.8 ~ 15.3 yr | 40(50%) | 28.2±0.6 | 55.8±2.1 |
|  | Placebo | 40 |  |  | 29.2±0.6 | 59.9±2.0 |
|  | Metformin（Pubertal） | 40 |  | 40(50%) | 29.4±0.5 | 76.9±2.4 |
|  | Placebo | 40 |  |  | 30.6±0.5 | 80.5±2.4 |
| Wilson 2010 | Metformin | 39 | 14.8±1.3 | 26(67%) | 35.9±5.7 | 95.9±16.6 |
|  | Placebo | 38 | 15.0±1.5 | 25(66%) | 35.9±4.7 | 101.8±15.7 |
| Wiegand 2010 | Metformin | 36 | 15.1 | 10(28%) | 34.25±4.95 | - |
|  | Placebo | 34 | 15 | 13(38%) | 35.47±5.77 | - |
| MP van der Aa 2016 | Metformin | 23 | median(interquartilerange)  13.6 (12.6 to 15.3) | 6(26.1%) | median (interquartilerange)  29.8 (28.1 to 34.5) | 82.2 (75.4 to 92.7) |
|  | Placebo | 19 | median(interquartilerange)  12.0 (11.3 to 14.0) | 8(42.1%) | median (interquartilerange) 30.5 (28.7 to 38.6) | 86.1 (74.0 to 103.0) |
| Rynders 2012 | Metformin | 7 | 15.0±2.0 | 4(57.1%) | 33.6±7.2 | 90.2±21.8 |
|  | Lifestyle modification alone | 9 | 13.4±1.7 | 3(33.3%) | 33.6±3.4 | 93.9±13.8 |
| Yanovski 2011 | Metformin | 53 | 10.1±1.6 | 23(43%) | 34.2±6.8 | 76.4±23.1 |
|  | Placebo | 47 | 10.4±1.4 | 17(36%) | 34.6±6.2 | 80.1±20.5 |
| Bassols 2019 | Metformin | 9 | 8.8±0.6 | 6(67%) | - | - |
|  | Placebo | 9 | 10.0±0.5 | 5(56%) | - | - |
| Garibay-Nieto 2017 | Metformin | 14 | 11.43±2.1 | - | 28.54±2.8 | 63.25±12.6 |
|  | Placebo | 17 | 12.59±2.62 |  | 28.79±2.8 | 70.15±13.11 |
|  | Conjugated Linoleic Acid | 17 | 11.41±2.71 |  | 27.48±3.7 | 62.11±18.47 |
| Kay 2001 | Metformin | 12 | 15.6±0.4 | 5(41.7%) | 41.2±1.8 | 116.0±5.1 |
|  | Placebo | 12 | 15.7±0.5 | 4(33.3%) | 40.8±1.4 | 113.4±5.0 |
| Rezvanian 2010 | Metformin | 45 | 13.1±1.4 | - | 26.4±0.5 | - |
|  | fluoxetine | 45 | 13.5±1.2 | - | 26.5±0.7 | - |
|  | Combination | 45 | 13.7±1.1 | - | 26.6±0.8 | - |
|  | Placebo | 45 | 13.4±1.4 | - | 26.2±0.6 | - |
| Li 2019 | Metformin | 42 | 12.27±1.64 | 26(61.9%) | 31.76±2.46 | - |
|  | Lifestyle modification alone | 42 | 11.99±1.45 | 28(66.7%) | 30.76±2.45 | - |
| Vendrell 2006 | Metformin | 11 | 16.07±0.97 | - | 33.6±5.6 | - |
|  | Placebo | 11 | 16.08±1.39 | - | 30.81±3.0 | - |
| Daniel 2022 | Semaglutide | 134 | 15.5±1.5 | 50(37%) | 37.7±6.7 | 109.9±25.2 |
|  | Placebo | 67 | 15.3±1.6 | 26(39%) | 35.7±5.4 | 102.6±22.3 |
| Kelly 2022 | Phentermine/Topiramate 15 mg/92 mg/day | 113 | 13.9±1.36 | 50(44.2%) | 39.0±7.4 | 108.5±25.0 |
|  | Phentermine/Topiramate 7.5 mg/46 mg/day | 54 | 14.1±1.28 | 26(48.1%) | 36.9±6.8 | 105.2±22.4 |
|  | Placebo | 56 | 14.0±1.41 | 26(46.4%) | 36.4±6.4 | 102.2±21.8 |
| Diene 2022 | Liraglutide (Adolescents) | 19 | 14.4±1.9 | 9(47.4%) | 36.3±6.5 | - |
|  | Placebo | 12 | 14.1±1.9 | 7(58.3%) | 40.2±10.7 | - |
|  | Liraglutide (Children) | 17 | 8.1±1.7 | 6(35.3%) | 32.4±7.5 | - |
|  | Placebo | 7 | 9.4±1.6 | 5(71.4%) | 30.3±5.5 | - |
| Yanovski 2012 | Orlistat | 100 | 14.65±1.38 | 35(35%) | - | - |
|  | Placebo | 100 | 14.52±1.46 | 34(34%) | - | - |
| Hoeger 2008 | Metformin | 18 | 14.7±1.6 | 0(0) | - | - |
|  | Placebo | 18 | 15.8±1.6 | 0(0) | - | - |

### Appendix 5: Risk of bias assessments

**Note:** D1: Risk of bias arising from the randomization process; D2: Risk of bias due to deviations from the intended interventions; D3: Risk of bias due to missing outcome data; D4: Risk of bias in measurement of the outcome; D5: Risk of bias in selection of the reported result; Overall: Overall risk of bias

| **Study** | **D1** | **D2** | **D3** | **D4** | **D5** | **Overall** |
| --- | --- | --- | --- | --- | --- | --- |
| **Change in BMI from baseline** | | | | | | |
| Ozkan 2004 | High | Some concerns | Low | Low | Low | High |
| Yu 2013 | Some concerns | Low | Low | Low | Low | Some concerns |
| Maahs 2006 | Low | Low | Low | Low | Low | Low |
| Shankar 2022 | Some concerns | Some concerns | Low | Low | Low | Some concerns |
| Fox 2016 | Low | Low | Low | Low | Low | Low |
| Barrientos 2022 | Some concerns | Low | Low | Low | Low | Some concerns |
| Kelly 2020 | Low | Low | Low | Low | Low | Low |
| Fox 2022 | Low | Low | Low | Low | Low | Low |
| Kelly 2012 | Low | Some concerns | Low | Low | Low | Some concerns |
| Weghuber 2020 | Low | Low | Low | Low | Low | Low |
| Atabek 2008 | Some concerns | Low | Low | Low | Low | Some concerns |
| Nadeau 2009 | Some concerns | Some concerns | Low | Low | Low | Some concerns |
| Evia-Viscarra 2012 | Low | Some concerns | Some concerns | Low | Low | Some concerns |
| Burgert 2008 | Some concerns | Some concerns | Low | Low | Low | Some concerns |
| Mauras 2012 | Some concerns | Some concerns | Low | Low | Low | Some concerns |
| Warnakulasuriya 2018 | Low | Some concerns | Low | Low | Low | Some concerns |
| Kendall 2013 | Low | Some concerns | Some concerns | Low | Low | Some concerns |
| Clarson 2009 | Low | Some concerns | Low | Low | Low | Some concerns |
| Pastor-Villaescusa 2017 | Some concerns | Some concerns | Low | Low | Low | Some concerns |
| Wilson 2010 | Low | Low | Low | Low | Low | Low |
| Wiegand 2010 | Some concerns | Some concerns | Some concerns | Low | Low | Some concerns |
| MP van der Aa 2016 | Low | Some concerns | Some concerns | Low | Low | Some concerns |
| Rynders 2012 | Some concerns | Low | Low | Low | Low | Some concerns |
| Yanovski 2011 | Low | Low | Low | Low | Low | Low |
| Garibay-Nieto 2017 | Some concerns | Some concerns | Low | Low | Low | Low |
| Rezvanian 2010 | Some concerns | Low | Low | Low | Low | Some concerns |
| Li 2019 | Some concerns | Some concerns | Low | Low | Low | Some concerns |
| Kelly 2022 | Low | Low | Low | Low | Low | Low |
| **Change in weight from baseline** | | | | | | |
| Ozkan 2004 | High | Some concerns | Low | Low | Low | High |
| Yu 2013 | Some concerns | Low | Low | Low | Low | Some concerns |
| Maahs 2006 | Low | Low | Low | Low | Low | Low |
| Shankar 2022 | Some concerns | Some concerns | Low | Low | Low | Some concerns |
| Fox 2016 | Low | Low | Low | Low | Low | Low |
| Barrientos 2022 | Some concerns | Low | Low | Low | Low | Some concerns |
| Kelly 2020 | Low | Low | Low | Low | Low | Low |
| Fox 2022 | Low | Low | Low | Low | Low | Low |
| Kelly 2012 | Low | Some concerns | Low | Low | Low | Some concerns |
| Weghuber 2020 | Low | Low | Low | Low | Low | Low |
| Atabek 2008 | Some concerns | Low | Low | Low | Low | Some concerns |
| Evia-Viscarra 2012 | Low | Some concerns | Some concerns | Low | Low | Some concerns |
| Burgert 2008 | Some concerns | Some concerns | Low | Low | Low | Some concerns |
| Mauras 2012 | Some concerns | Some concerns | Low | Low | Low | Some concerns |
| Warnakulasuriya 2018 | Low | Some concerns | Low | Low | Low | Some concerns |
| Kendall 2013 | Low | Some concerns | Some concerns | Low | Low | Some concerns |
| Pastor-Villaescusa 2017 | Some concerns | Some concerns | Low | Low | Low | Some concerns |
| MP van der Aa 2016 | Low | Some concerns | Some concerns | Low | Low | Some concerns |
| Rynders 2012 | Some concerns | Low | Low | Low | Low | Some concerns |
| Yanovski 2011 | Low | Low | Low | Low | Low | Low |
| Garibay-Nieto 2017 | Some concerns | Some concerns | Low | Low | Low | Low |
| Kay 2001 | Some concerns | Low | Low | Low | Low | Some concerns |
| Kelly 2022 | Low | Low | Low | Low | Low | Low |
| **Percentage of participants achieving BMI reduction of at least 5%** | | | | | | |
| Chanoine­ 2005 | Low | Low | Low | Low | Low | Low |
| Fox 2016 | Low | Low | Low | Low | Low | Low |
| Kelly 2020 | Low | Low | Low | Low | Low | Low |
| Daniel 2022 | Low | Low | Low | Low | Low | Low |
| Kelly 2022 | Low | Low | Low | Low | Low | Low |
| Diene 2022 | Some concerns | Low | Low | Low | Low | Some concerns |
| **Percentage of participants achieving BMI reduction of at least 10%** | | | | | | |
| Chanoine­ 2005 | Low | Low | Low | Low | Low | Low |
| Fox 2016 | Low | Low | Low | Low | Low | Low |
| Kelly 2020 | Low | Low | Low | Low | Low | Low |
| Daniel 2022 | Low | Low | Low | Low | Low | Low |
| Kelly 2022 | Low | Low | Low | Low | Low | Low |
| Diene 2022 | Some concerns | Low | Low | Low | Low | Some concerns |
| **Change in BMI z-score from baseline** | | | | | | |
| Yu 2013 | Some concerns | Low | Low | Low | Low | Some concerns |
| Fox 2016 | Low | Low | Low | Low | Low | Low |
| Kelsey 2021 | Low | Low | Some concerns | Low | Some concerns | Some concerns |
| Clarson 2009 | Low | Some concerns | Low | Low | Low | Some concerns |
| Pastor-Villaescusa 2017 | Some concerns | Some concerns | Low | Low | Low | Some concerns |
| Wilson 2010 | Low | Low | Low | Low | Low | Low |
| **Change in BMI SDS from baseline** | | | | | | |
| Kelly 2020 | Low | Low | Low | Low | Low | Low |
| Weghuber 2020 | Low | Low | Low | Low | Low | Low |
| Warnakulasuriya 2018 | Low | Some concerns | Low | Low | Low | Some concerns |
| Kendall 2013 | Low | Some concerns | Some concerns | Low | Low | Some concerns |
| Wiegand 2010 | Some concerns | Some concerns | Some concerns | Low | Low | Some concerns |
| MP van der Aa 2016 | Low | Some concerns | Some concerns | Low | Low | Some concerns |
| Yanovski 2011 | Low | Low | Low | Low | Low | Low |
| Bassols 2019 | Low | Low | Low | Low | Low | Low |
| Diene 2022 | Some concerns | Low | Low | Low | Low | Some concerns |
| **Total gastrointestinal adverse events** | | | | | | |
| Ozkan 2004 | High | Some concerns | Low | Low | Low | High |
| Yu 2013 | Some concerns | Low | Low | Low | Low | Some concerns |
| Barrientos 2022 | Some concerns | Low | Low | Low | Low | Some concerns |
| Hsia 2020 | Low | Low | Low | Low | Low | Low |
| Kelly 2020 | Low | Low | Low | Low | Low | Low |
| Mastrandrea 2019 | Low | Low | Low | Low | Low | Low |
| Danne 2017 | Some concerns | Low | Low | Low | Low | Some concerns |
| Weghuber 2020 | Low | Low | Low | Low | Low | Low |
| Atabek 2008 | Some concerns | Low | Low | Low | Low | Some concerns |
| Kelsey 2021 | Low | Low | Some concerns | Low | Some concerns | Some concerns |
| Nadeau 2009 | Some concerns | Some concerns | Low | Low | Low | Some concerns |
| Evia-Viscarra 2012 | Low | Some concerns | Some concerns | Low | Low | Some concerns |
| Warnakulasuriya 2018 | Low | Some concerns | Low | Low | Low | Some concerns |
| Pastor-Villaescusa 2017 | Some concerns | Some concerns | Low | Low | Low | Some concerns |
| Wiegand 2010 | Some concerns | Some concerns | Some concerns | Low | Low | Some concerns |
| Vendrell 2006 | Some concerns | Low | Low | Low | Low | Some concerns |
| Daniel 2022 | Low | Low | Low | Low | Low | Low |
| Kelly 2022 | Low | Low | Low | Low | Low | Low |
| Diene 2022 | Some concerns | Low | Low | Low | Low | Some concerns |
| **Discontinuation due to adverse events** | | | | | | |
| Ozkan 2004 | High | Some concerns | Low | Low | Low | High |
| Chanoine­ 2005 | Low | Low | Low | Low | Low | Low |
| Maahs 2006 | Low | Low | Low | Low | Low | Low |
| Shankar 2022 | Some concerns | Some concerns | Low | Low | Low | Some concerns |
| Hsia 2020 | Low | Low | Low | Low | Low | Low |
| Arslanian 2022 | Low | Low | Low | Low | Low | Low |
| Kelly 2020 | Low | Low | Low | Low | Low | Low |
| Tamborlane 2019 | Low | Low | Low | Low | Low | Low |
| Weghuber 2020 | Low | Low | Low | Low | Low | Low |
| Evia-Viscarra 2012 | Low | Some concerns | Some concerns | Low | Low | Some concerns |
| Wilson 2010 | Low | Low | Low | Low | Low | Low |
| Wiegand 2010 | Some concerns | Some concerns | Some concerns | Low | Low | Some concerns |
| MP van der Aa 2016 | Low | Some concerns | Some concerns | Low | Low | Some concerns |
| Yanovski 2011 | Low | Low | Low | Low | Low | Low |
| Vendrell 2006 | Some concerns | Low | Low | Low | Low | Some concerns |
| Daniel 2022 | Low | Low | Low | Low | Low | Low |
| Kelly 2022 | Low | Low | Low | Low | Low | Low |
| Diene 2022 | Some concerns | Low | Low | Low | Low | Some concerns |
| **Serious adverse events** | | | | | | |
| Chanoine­ 2005 | Low | Low | Low | Low | Low | Low |
| Shankar 2022 | Some concerns | Some concerns | Low | Low | Low | Some concerns |
| Barrientos 2022 | Some concerns | Low | Low | Low | Low | Some concerns |
| Hsia 2020 | Low | Low | Low | Low | Low | Low |
| Arslanian 2022 | Low | Low | Low | Low | Low | Low |
| Kelly 2020 | Low | Low | Low | Low | Low | Low |
| Tamborlane 2019 | Low | Low | Low | Low | Low | Low |
| Fox 2022 | Low | Low | Low | Low | Low | Low |
| Wilson 2010 | Low | Low | Low | Low | Low | Low |
| Daniel 2022 | Low | Low | Low | Low | Low | Low |
| Kelly 2022 | Low | Low | Low | Low | Low | Low |
| Diene 2022 | Some concerns | Low | Low | Low | Low | Some concerns |
| **Nausea events** | | | | | | |
| Chanoine­ 2005 | Low | Low | Low | Low | Low | Low |
| Shankar 2022 | Some concerns | Some concerns | Low | Low | Low | Some concerns |
| Barrientos 2022 | Some concerns | Low | Low | Low | Low | Some concerns |
| Arslanian 2022 | Low | Low | Low | Low | Low | Low |
| Kelly 2020 | Low | Low | Low | Low | Low | Low |
| Mastrandrea 2019 | Low | Low | Low | Low | Low | Low |
| Danne 2017 | Some concerns | Low | Low | Low | Low | Some concerns |
| Tamborlane 2019 | Low | Low | Low | Low | Low | Low |
| Fox 2022 | Low | Low | Low | Low | Low | Low |
| Nadeau 2009 | Some concerns | Some concerns | Low | Low | Low | Some concerns |
| Wilson 2010 | Low | Low | Low | Low | Low | Low |
| MP van der Aa 2016 | Low | Some concerns | Some concerns | Low | Low | Some concerns |
| Kay 2001 | Some concerns | Low | Low | Low | Low | Some concerns |
| Daniel 2022 | Low | Low | Low | Low | Low | Low |
| Kelly 2022 | Low | Low | Low | Low | Low | Low |
| **Vomiting events** | | | | | | |
| Shankar 2022 | Some concerns | Some concerns | Low | Low | Low | Some concerns |
| Barrientos 2022 | Some concerns | Low | Low | Low | Low | Some concerns |
| Arslanian 2022 | Low | Low | Low | Low | Low | Low |
| Kelly 2020 | Low | Low | Low | Low | Low | Low |
| Mastrandrea 2019 | Low | Low | Low | Low | Low | Low |
| Danne 2017 | Some concerns | Low | Low | Low | Low | Some concerns |
| Tamborlane 2019 | Low | Low | Low | Low | Low | Low |
| Fox 2022 | Low | Low | Low | Low | Low | Low |
| Wilson 2010 | Low | Low | Low | Low | Low | Low |
| Daniel 2022 | Low | Low | Low | Low | Low | Low |
| Diene 2022 | Some concerns | Low | Low | Low | Low | Some concerns |
| **Diarrhea events** | | | | | | |
| Shankar 2022 | Some concerns | Some concerns | Low | Low | Low | Some concerns |
| Arslanian 2022 | Low | Low | Low | Low | Low | Low |
| Kelly 2020 | Low | Low | Low | Low | Low | Low |
| Mastrandrea 2019 | Low | Low | Low | Low | Low | Low |
| Danne 2017 | Some concerns | Low | Low | Low | Low | Some concerns |
| Tamborlane 2019 | Low | Low | Low | Low | Low | Low |
| Fox 2022 | Low | Low | Low | Low | Low | Low |
| Atabek 2008 | Some concerns | Low | Low | Low | Low | Some concerns |
| Nadeau 2009 | Some concerns | Some concerns | Low | Low | Low | Some concerns |
| Pastor-Villaescusa 2017 | Some concerns | Some concerns | Low | Low | Low | Some concerns |
| MP van der Aa 2016 | Low | Some concerns | Some concerns | Low | Low | Some concerns |
| Daniel 2022 | Low | Low | Low | Low | Low | Low |
| Diene 2022 | Some concerns | Low | Low | Low | Low | Some concerns |

### Appendix 6: Other outcomes

#### 6.1 Network plots

**Notes:** LMA, Lifestyle modification alone; GLP-1, glucagon-like peptide-1.

**Outcome: percentage of participants achieving BMI reduction of at least 10%**

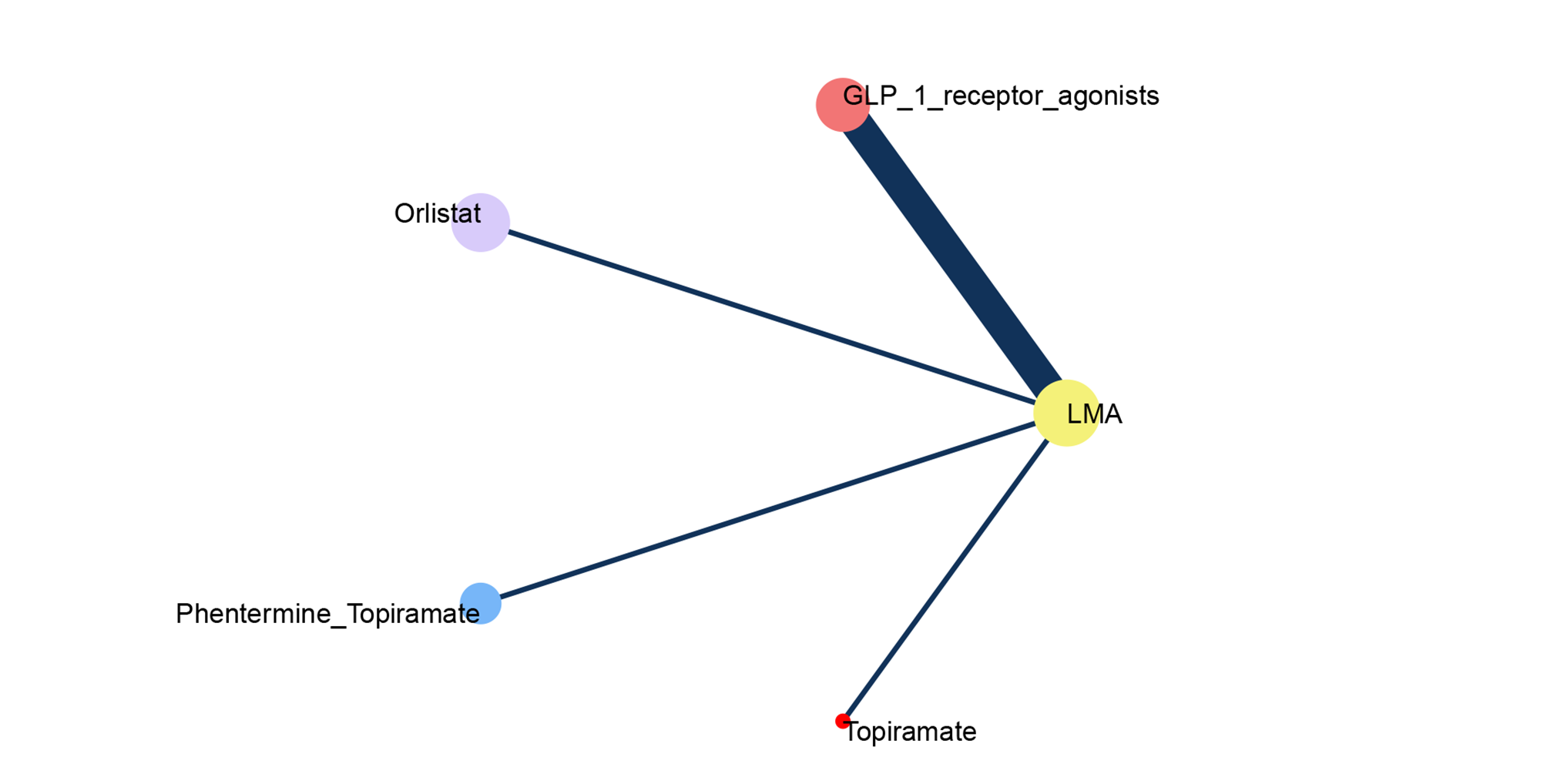

**Outcome: change in BMI z-score from baseline**

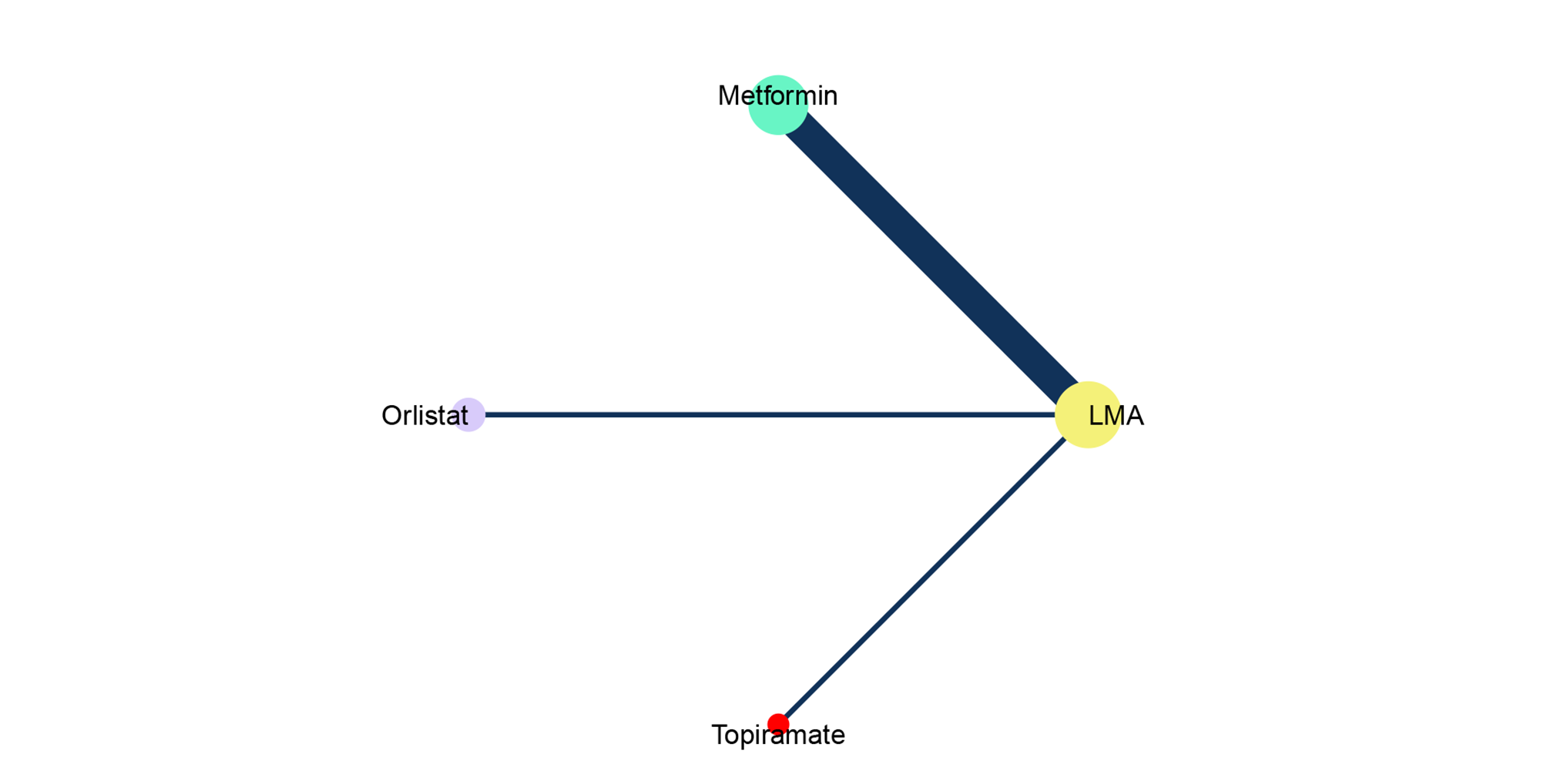

**Outcome: change in BMI SDS from baseline**

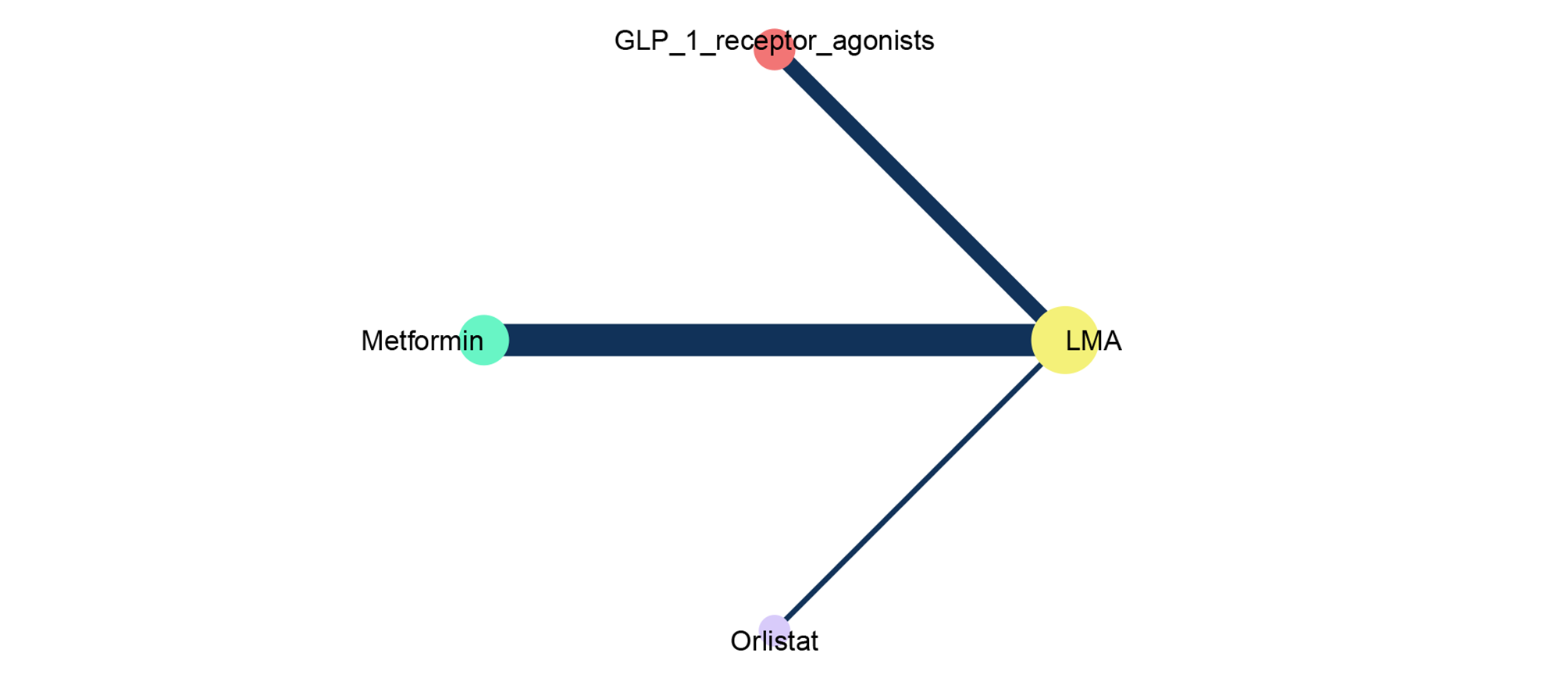

**Outcome: total gastrointestinal adverse events**

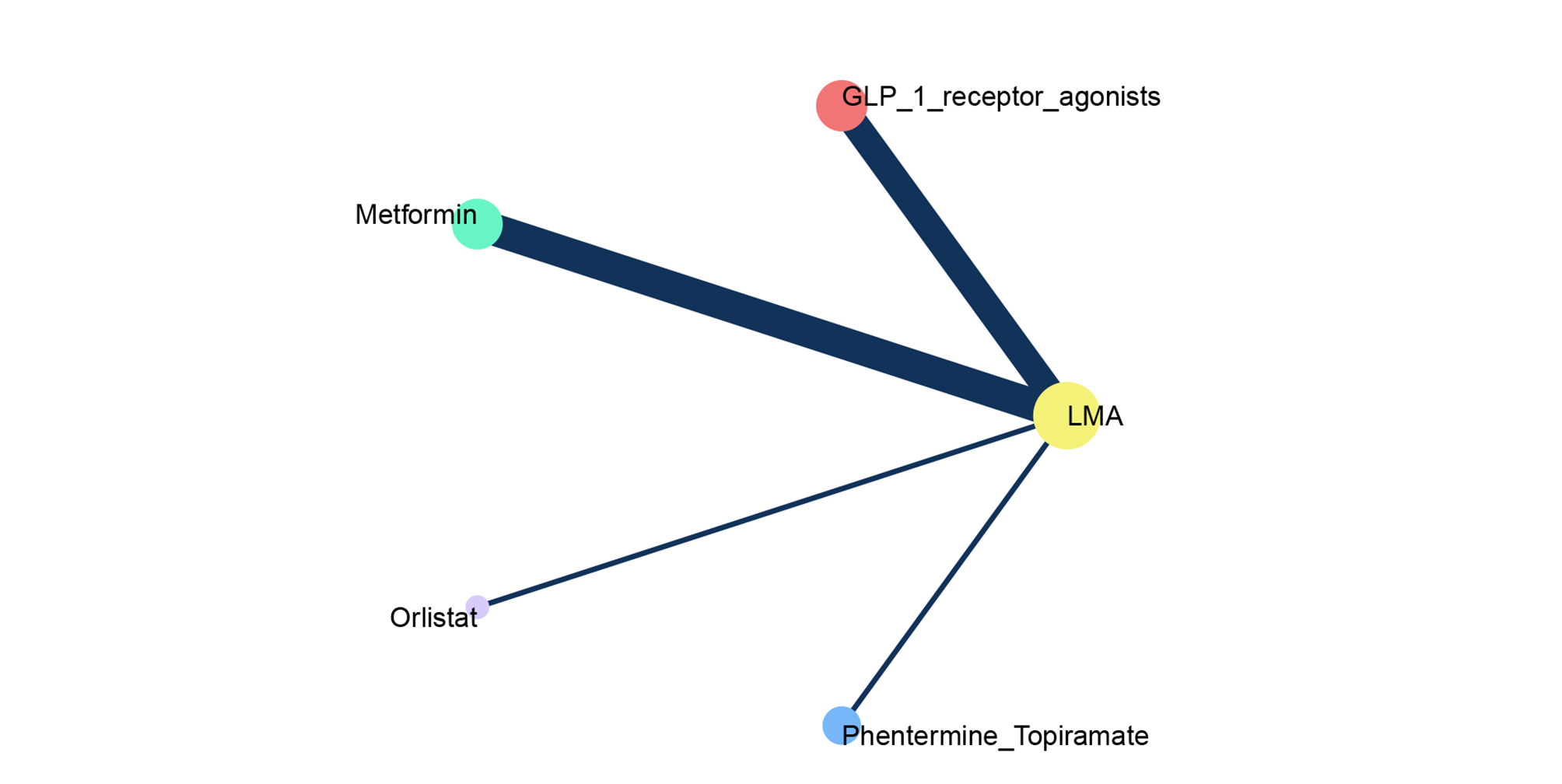

**Outcome: discontinuation due to any adverse event**

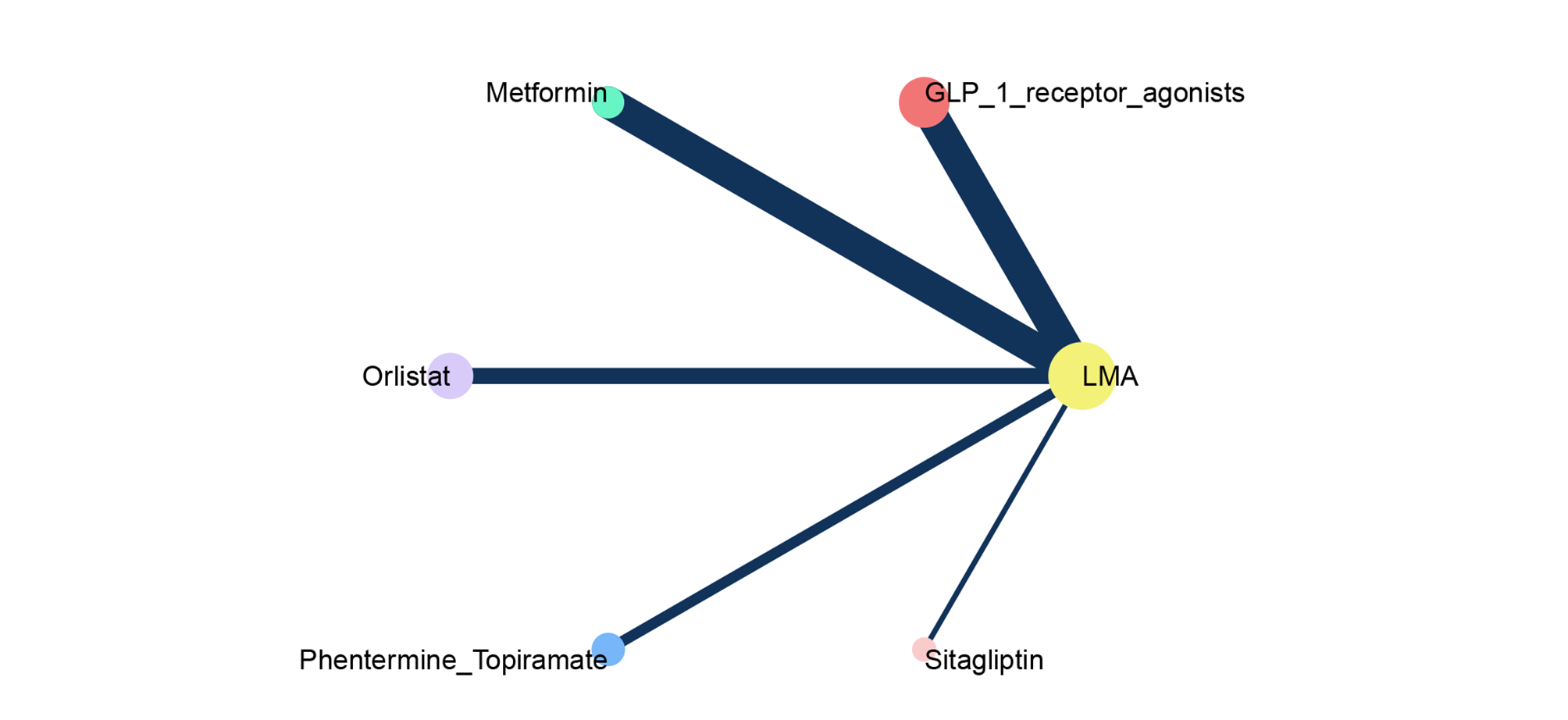

**Outcome: serious adverse events**

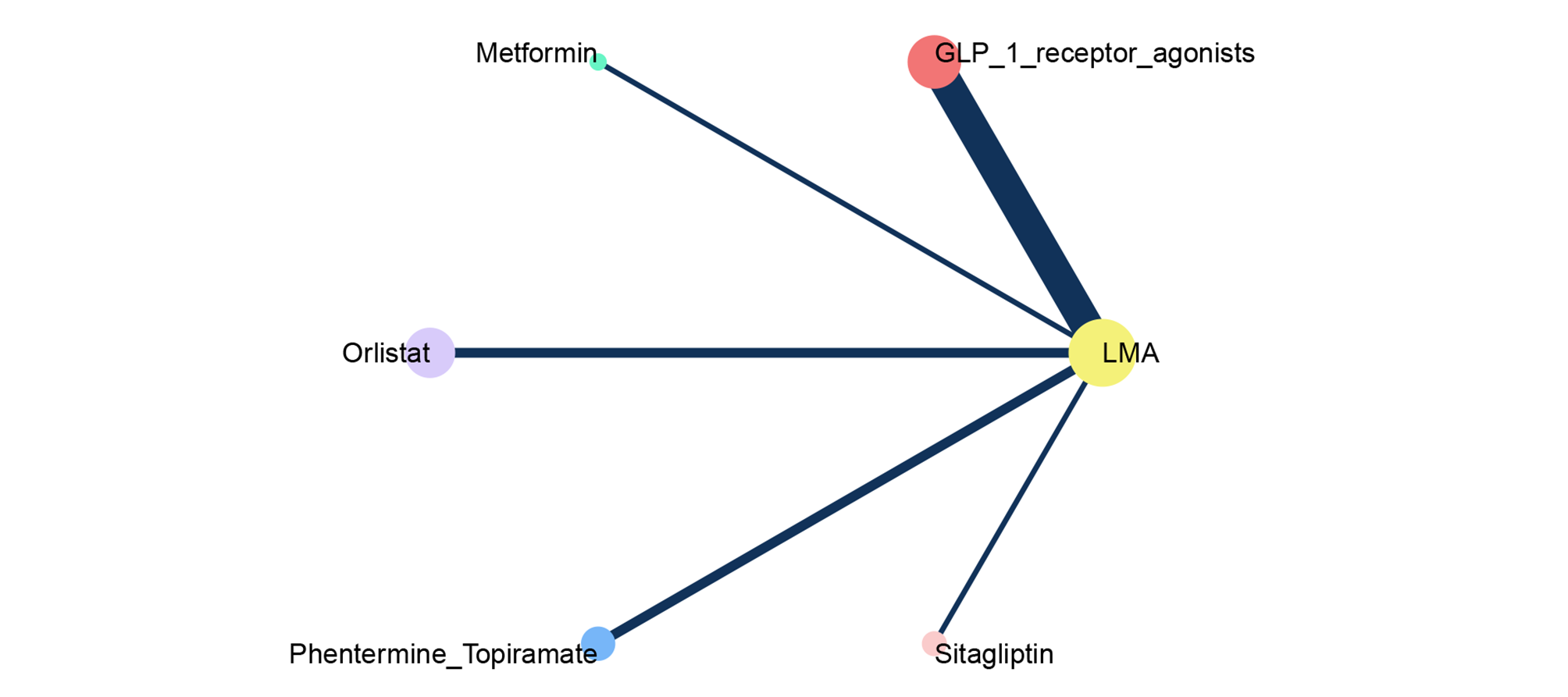

**Outcome: nausea events**

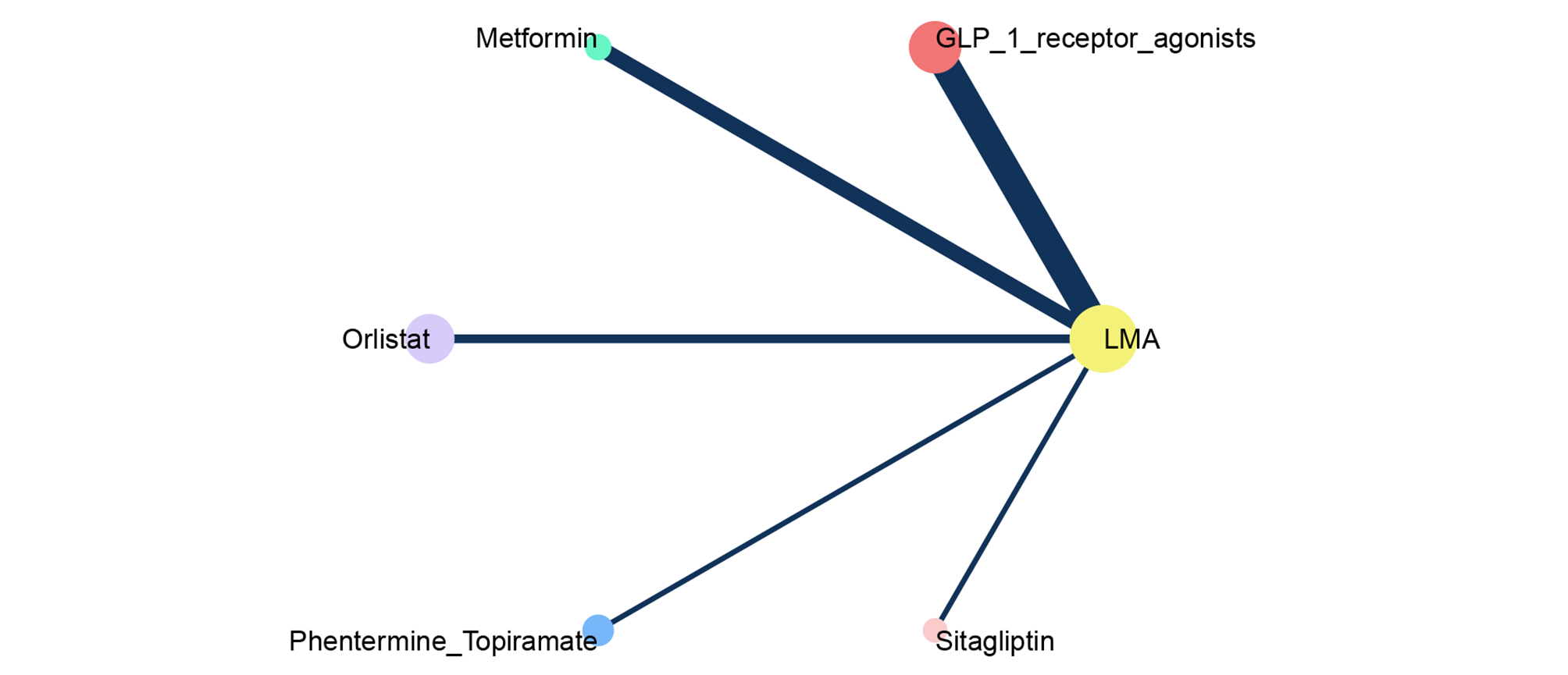

**Outcome: vomiting events**

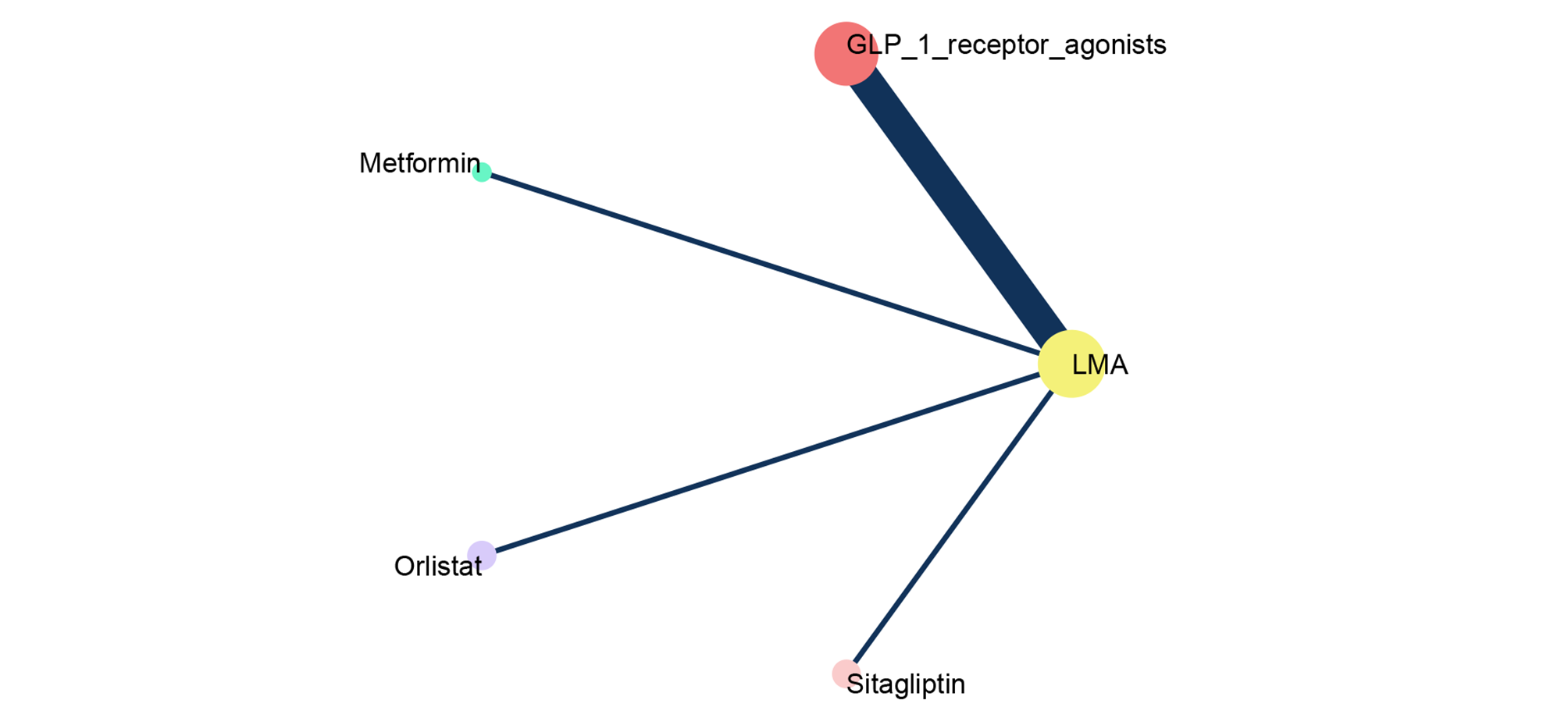

**Outcome: diarrhea events**

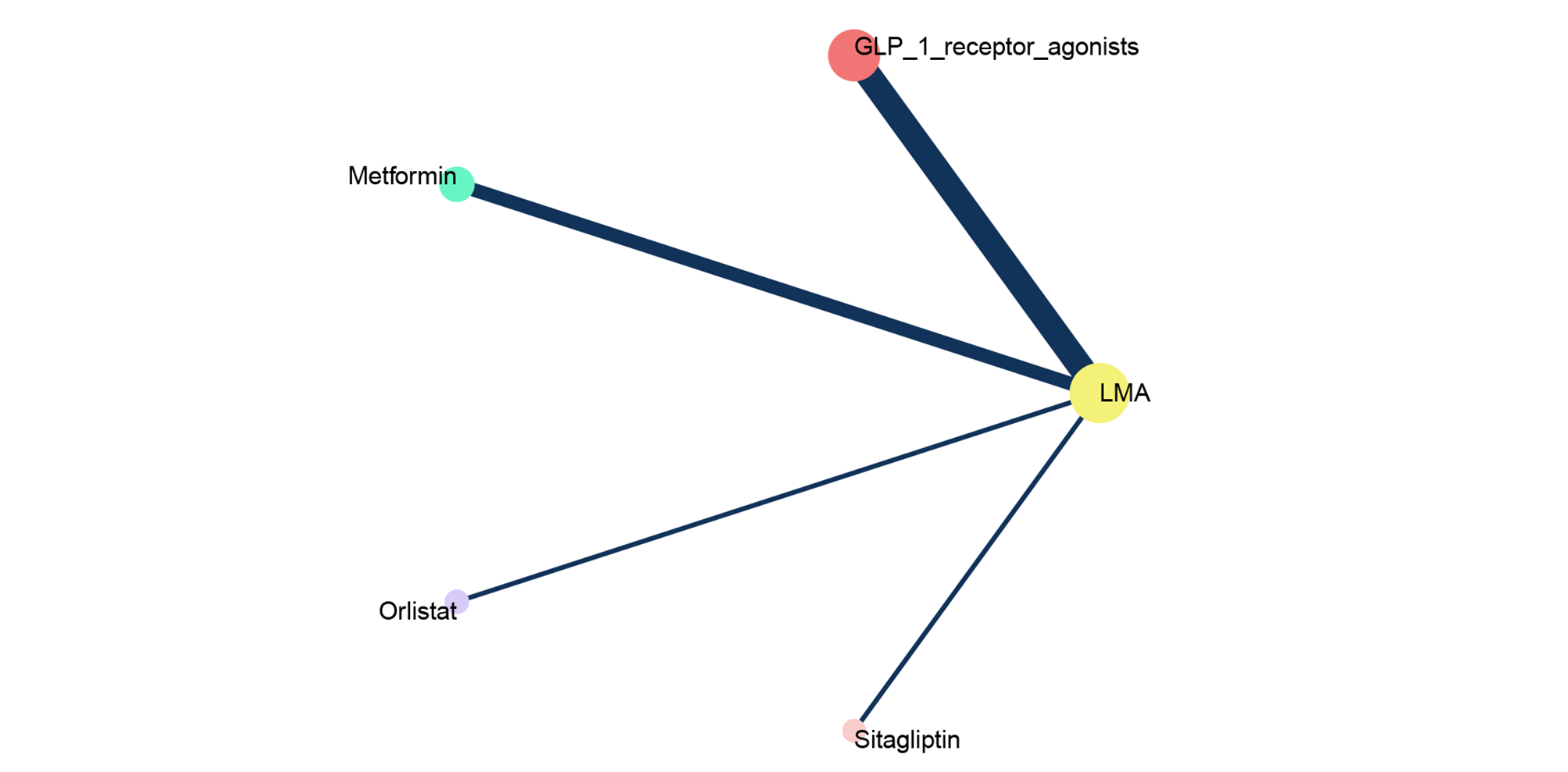

#### 6.2 Network estimates (league tables)

**Outcome: percentage of participants achieving BMI reduction of at least 10% (odds ratio; 95% confidence interval)**

High

Moderate

Low

Very low

Certainty of evidence

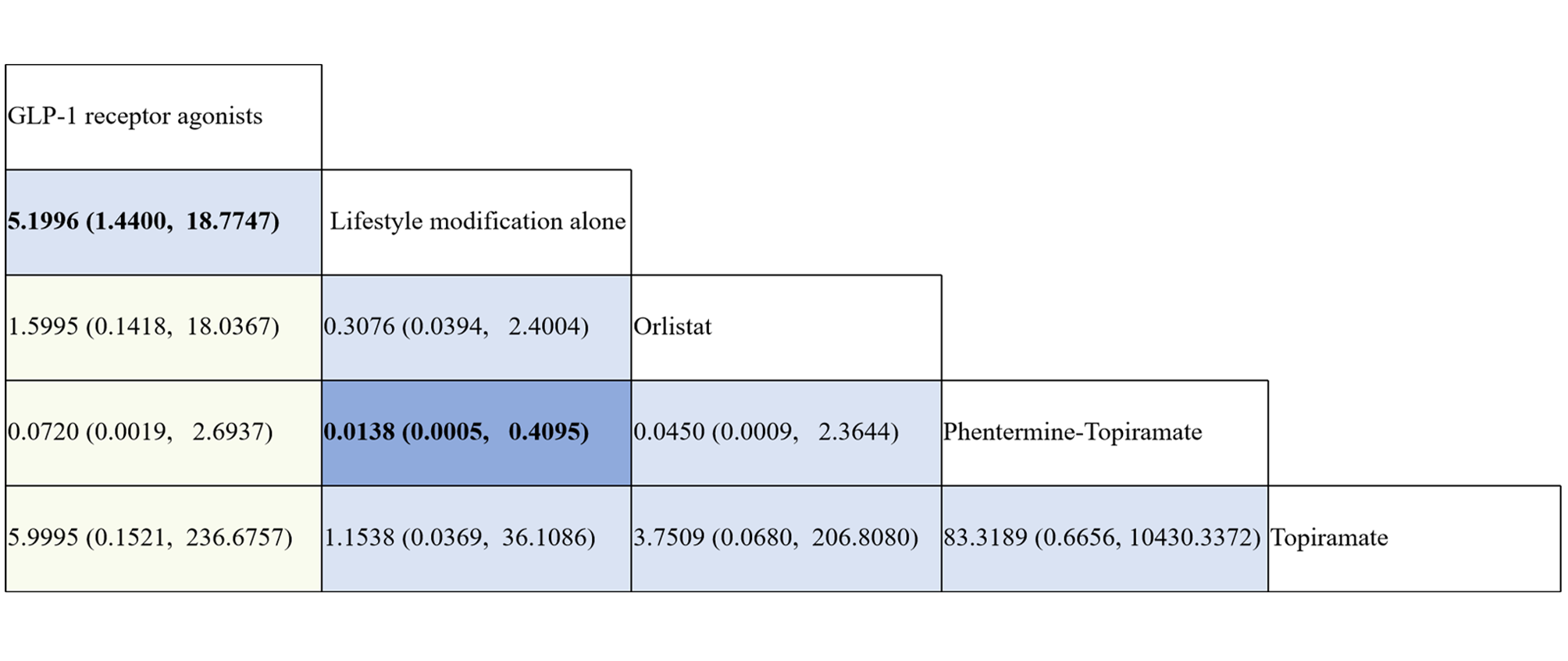

**Outcome: change in BMI z-score from baseline (mean difference; 95% confidence interval)**

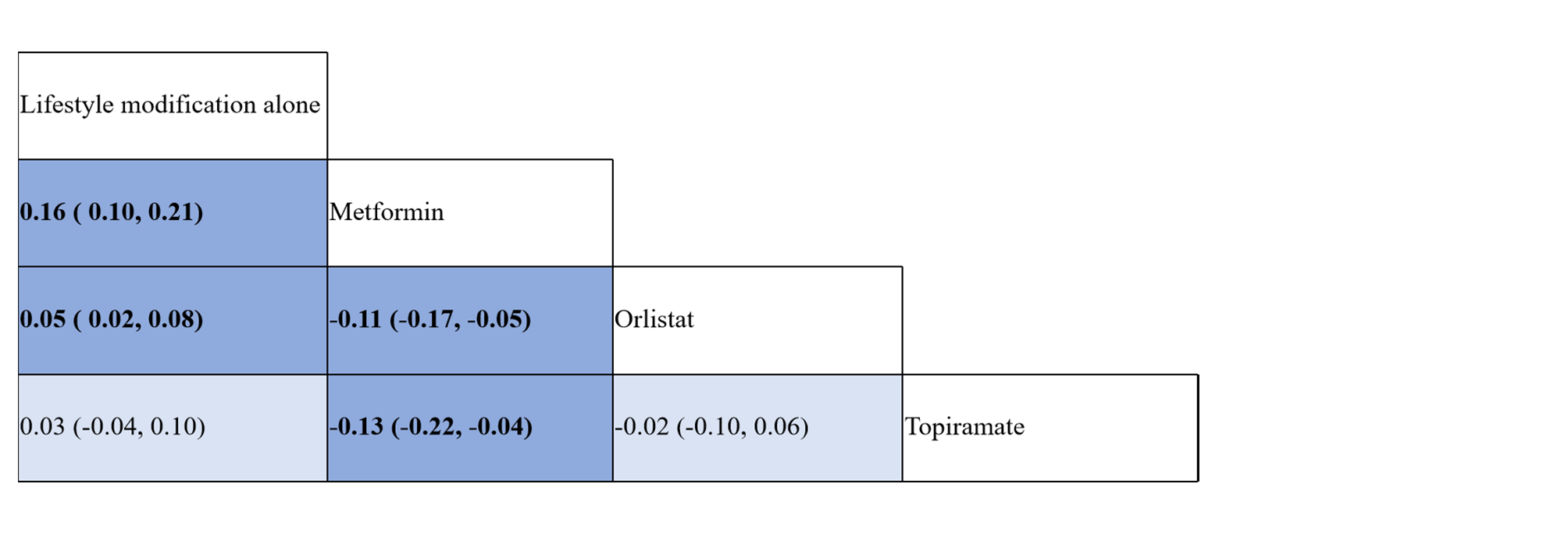

**Outcome: change in BMI SDS from baseline (mean difference; 95% confidence interval)**

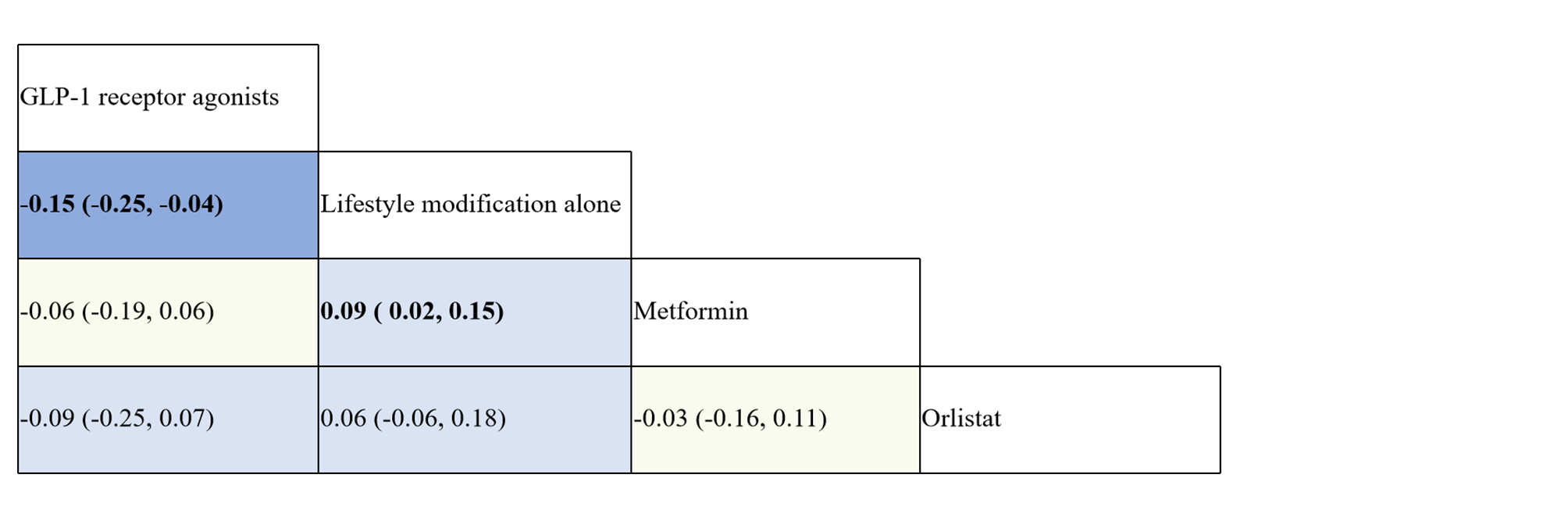

**Outcome: total gastrointestinal adverse events (odds ratio; 95% confidence interval)**

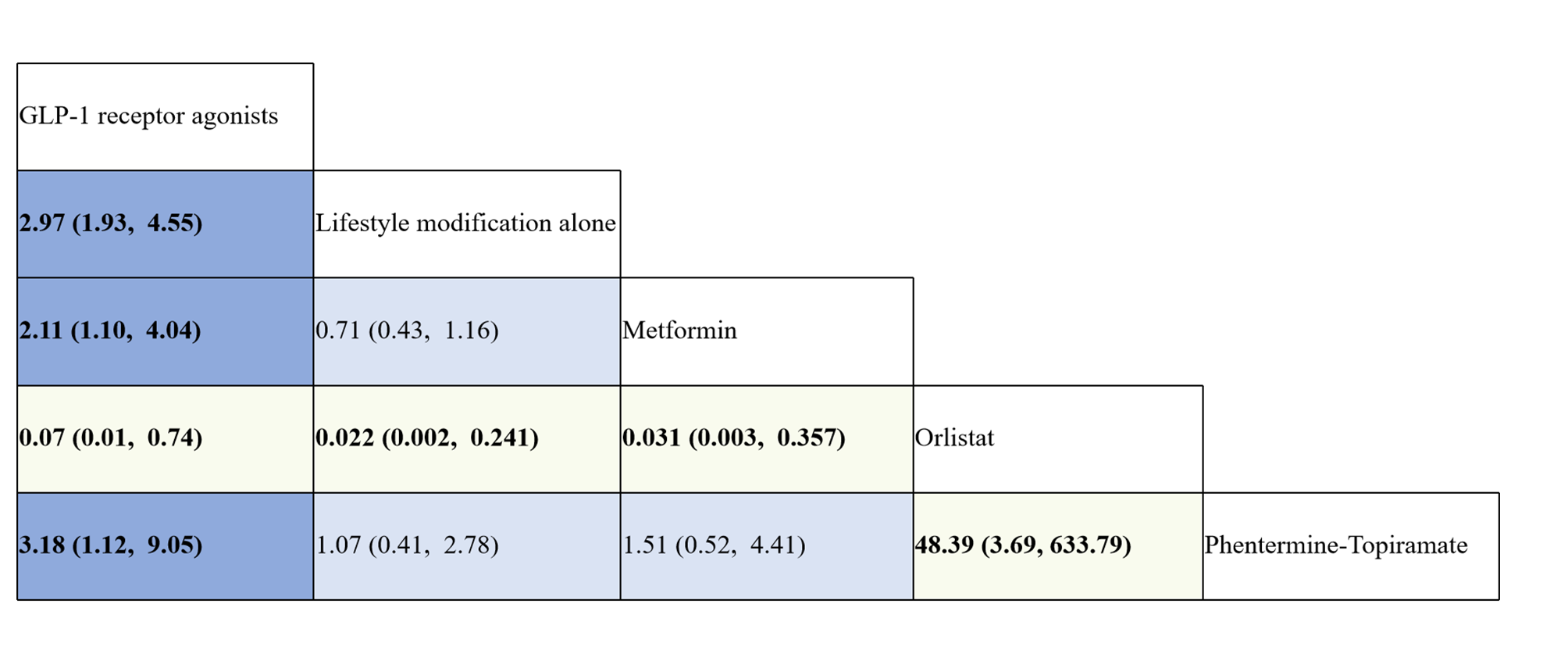

**Outcome: discontinuation due to any adverse event (odds ratio; 95% confidence interval)**

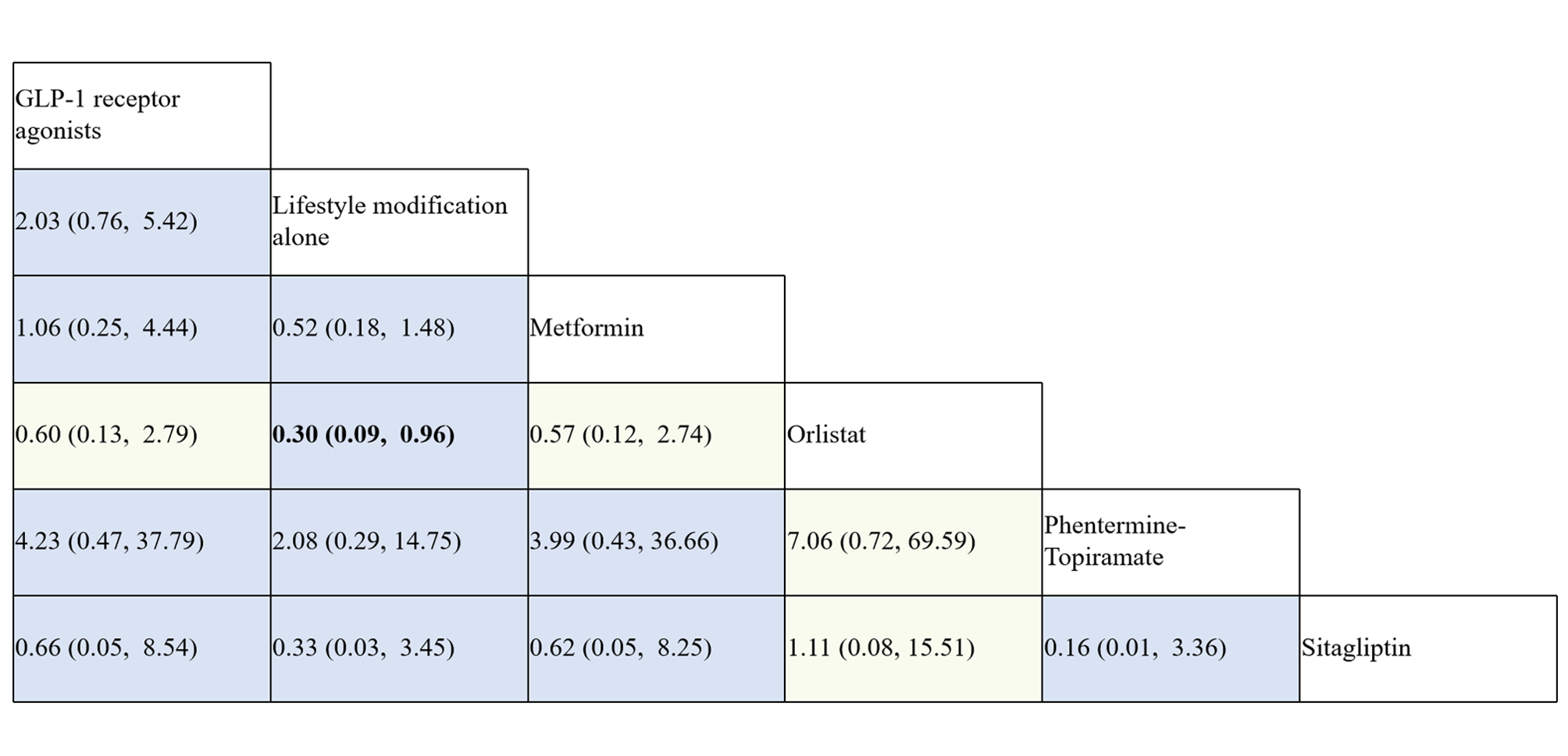

**Outcome: serious adverse events (odds ratio; 95% confidence interval)**

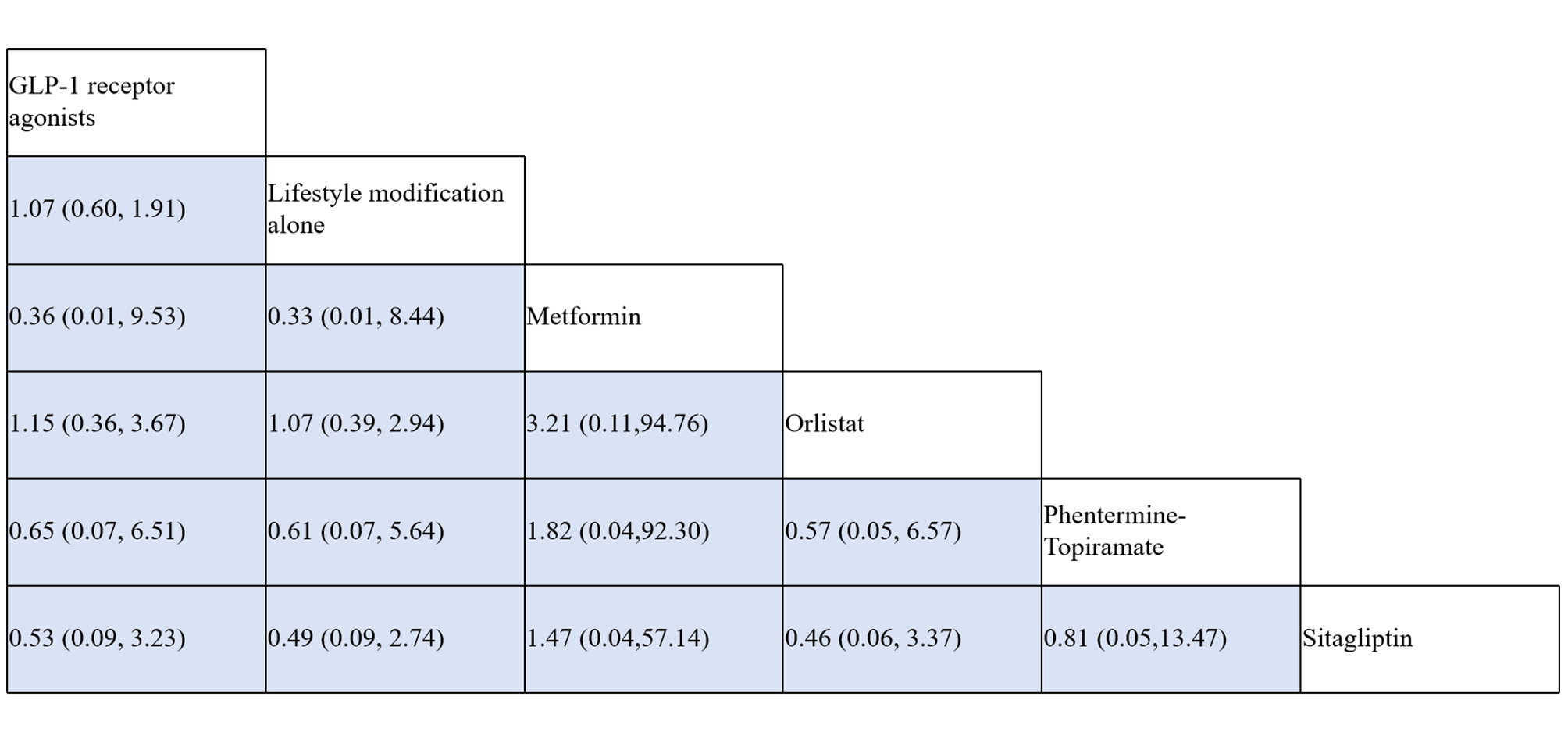

**Outcome: nausea events (odds ratio; 95% confidence interval)**

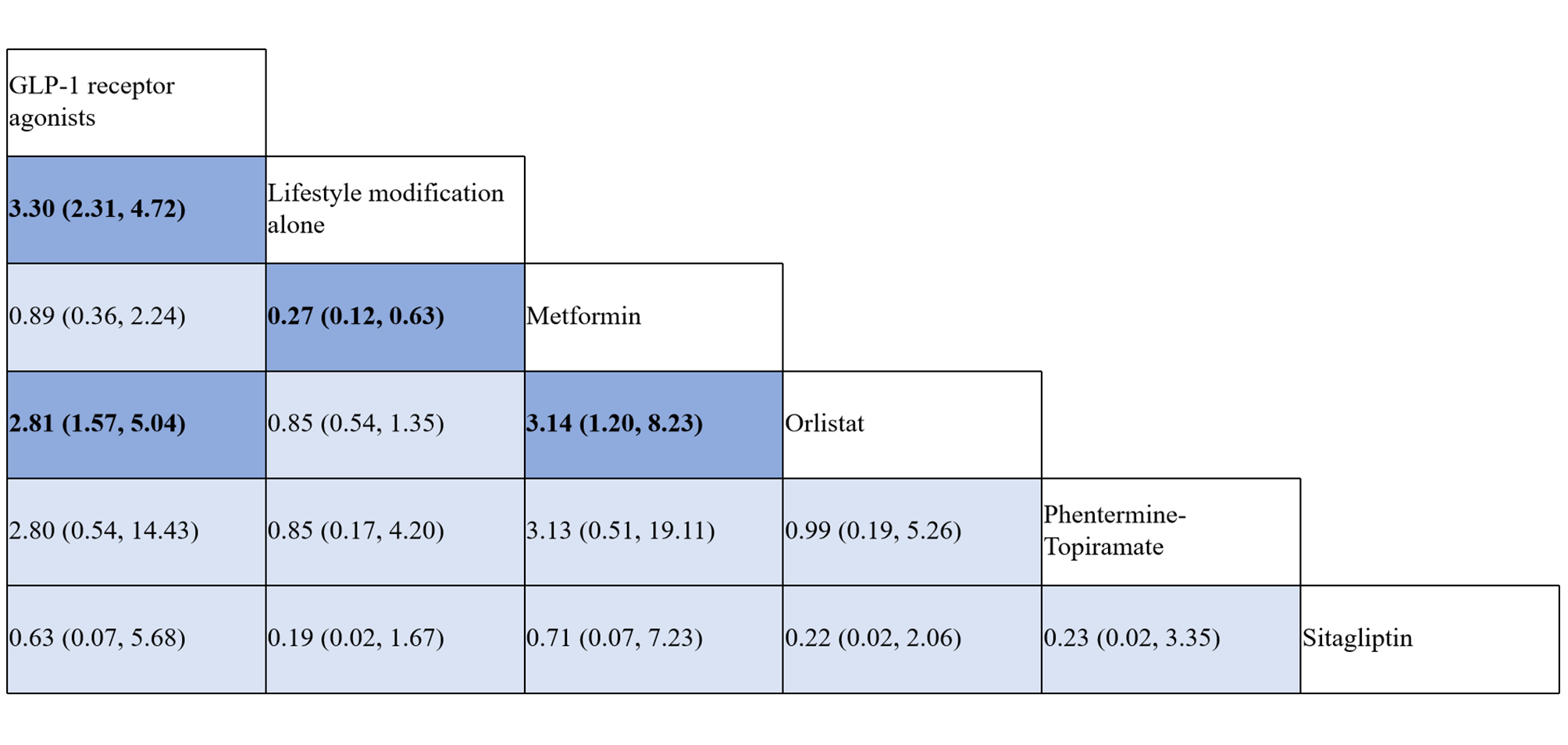

**Outcome: vomiting events** **(odds ratio; 95% confidence interval)**

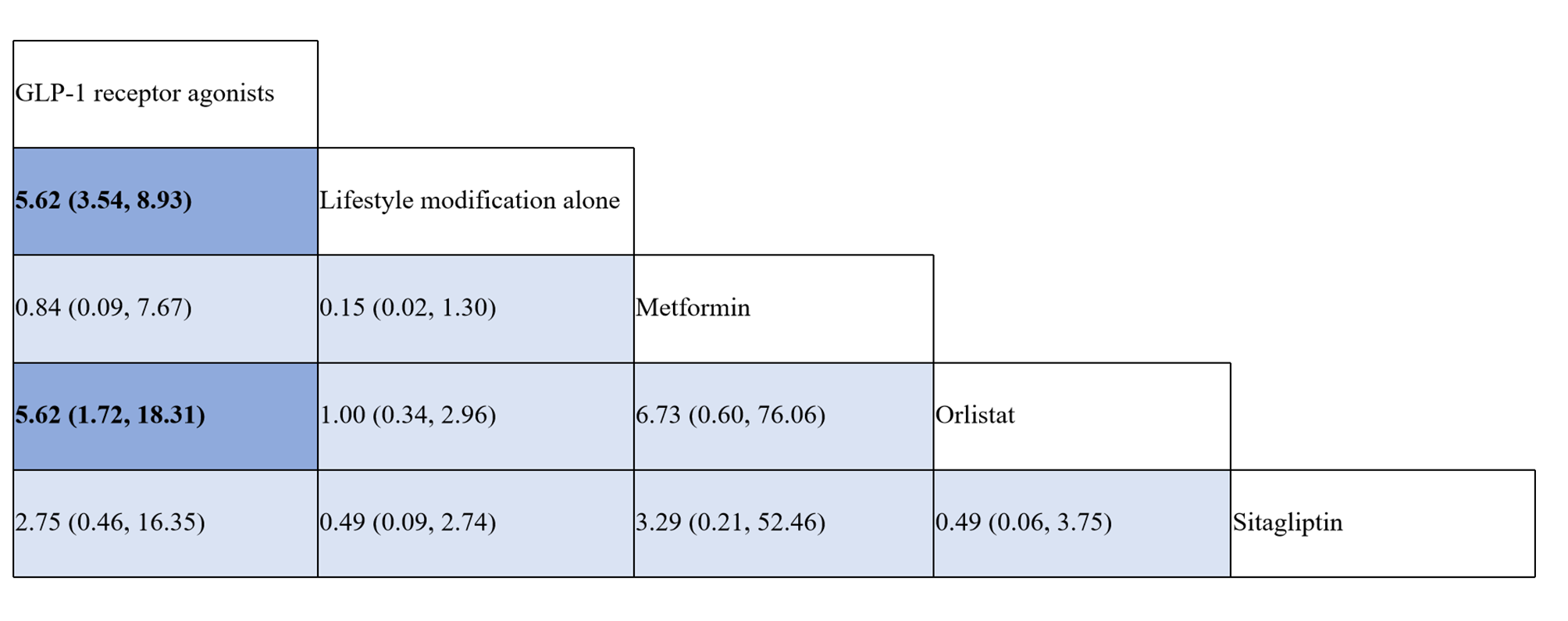

**Outcome: diarrhea events (odds ratio;** **95% confidence interval)**

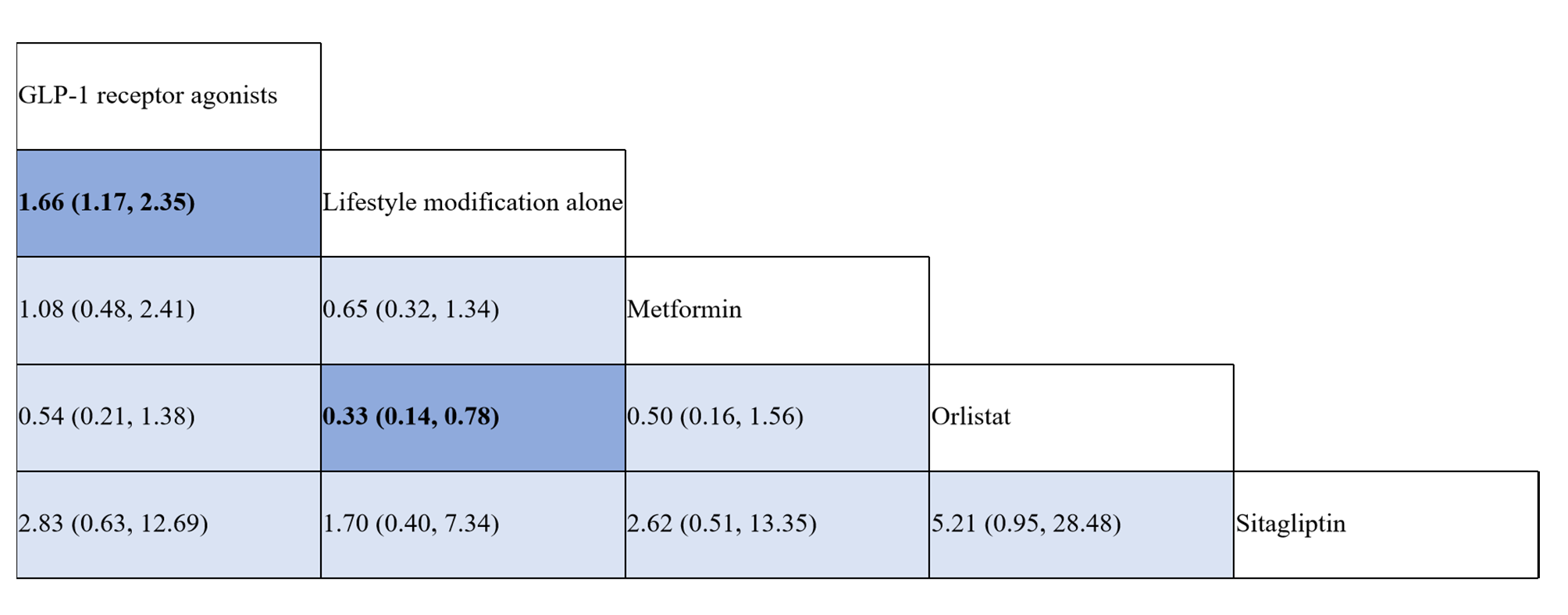

#### 6.3 Absolute effects

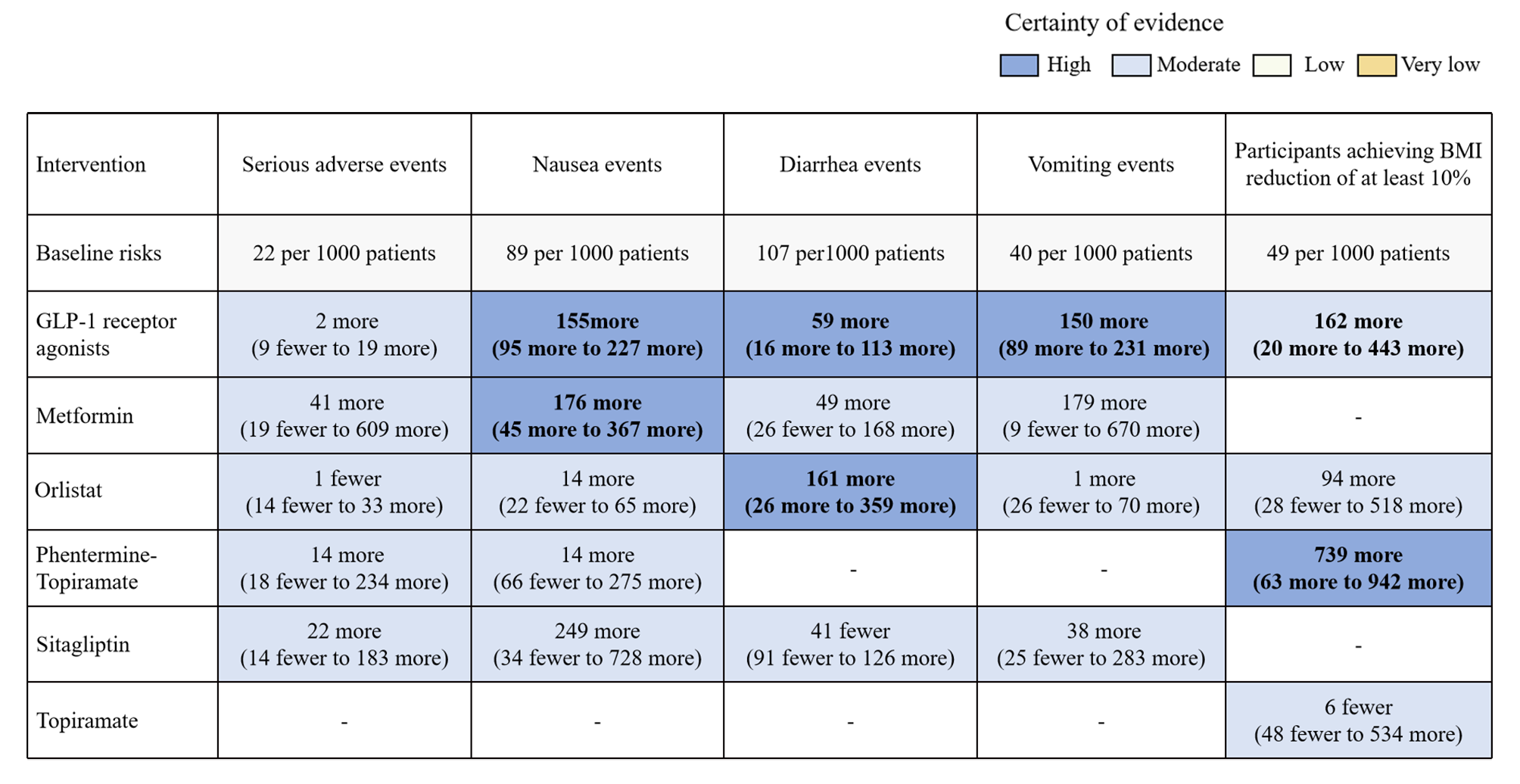

#### 6.4 Forest plots

**Notes:** LMA, Lifestyle modification alone; GLP-1, glucagon-like peptide-1; Metformin_Fluoxetine, Metformin combined with Fluoxetine; MD, mean difference; OR, odds ratio; 95%-CI, 95% confidence interval; “other” means drugs.

**Outcome:** **change in BMI from baseline**

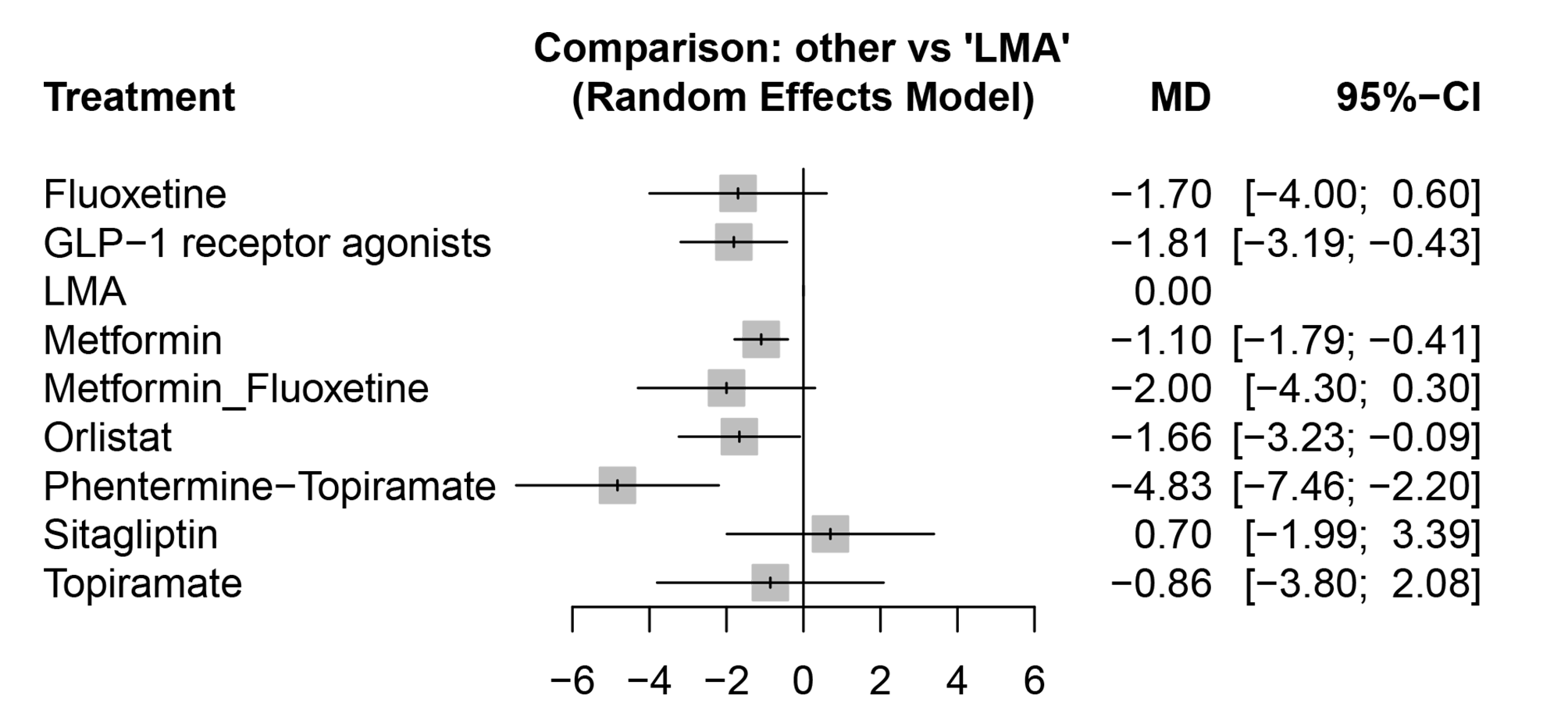

**Outcome: change in weight from baseline**

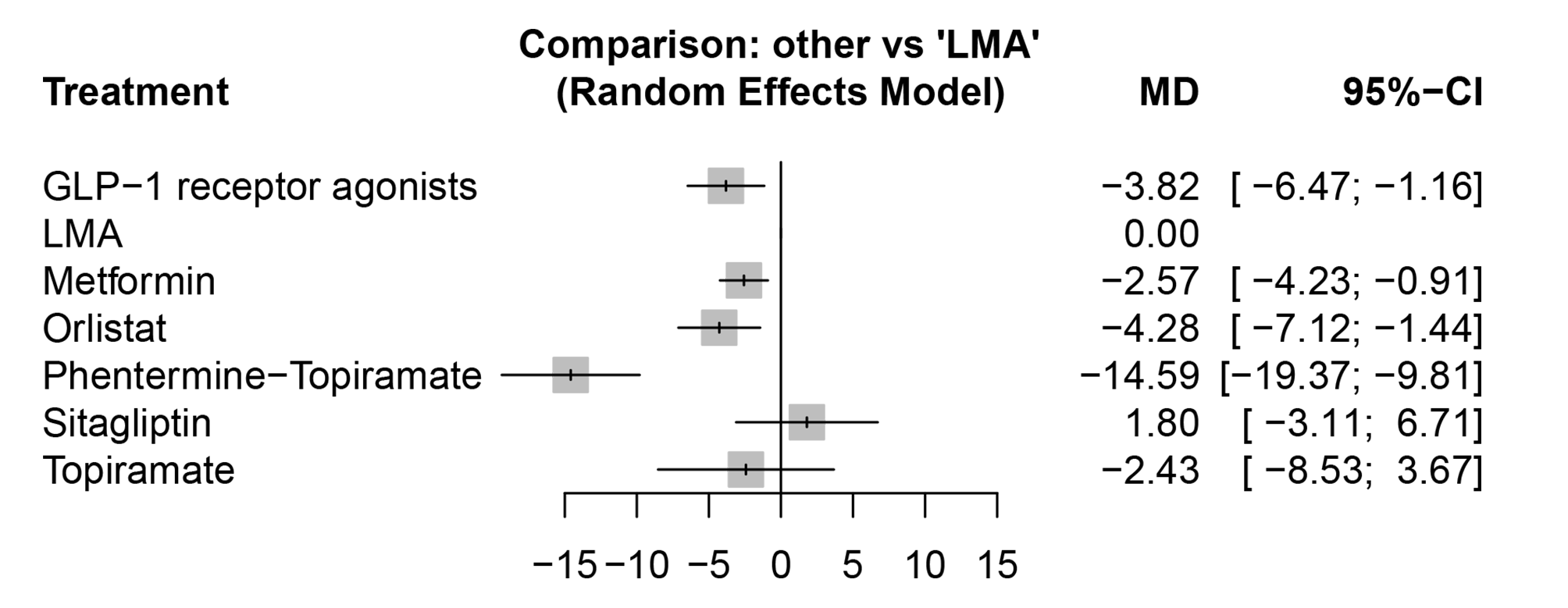

**Outcome: percentage of participants achieving BMI reduction of at least 5%**

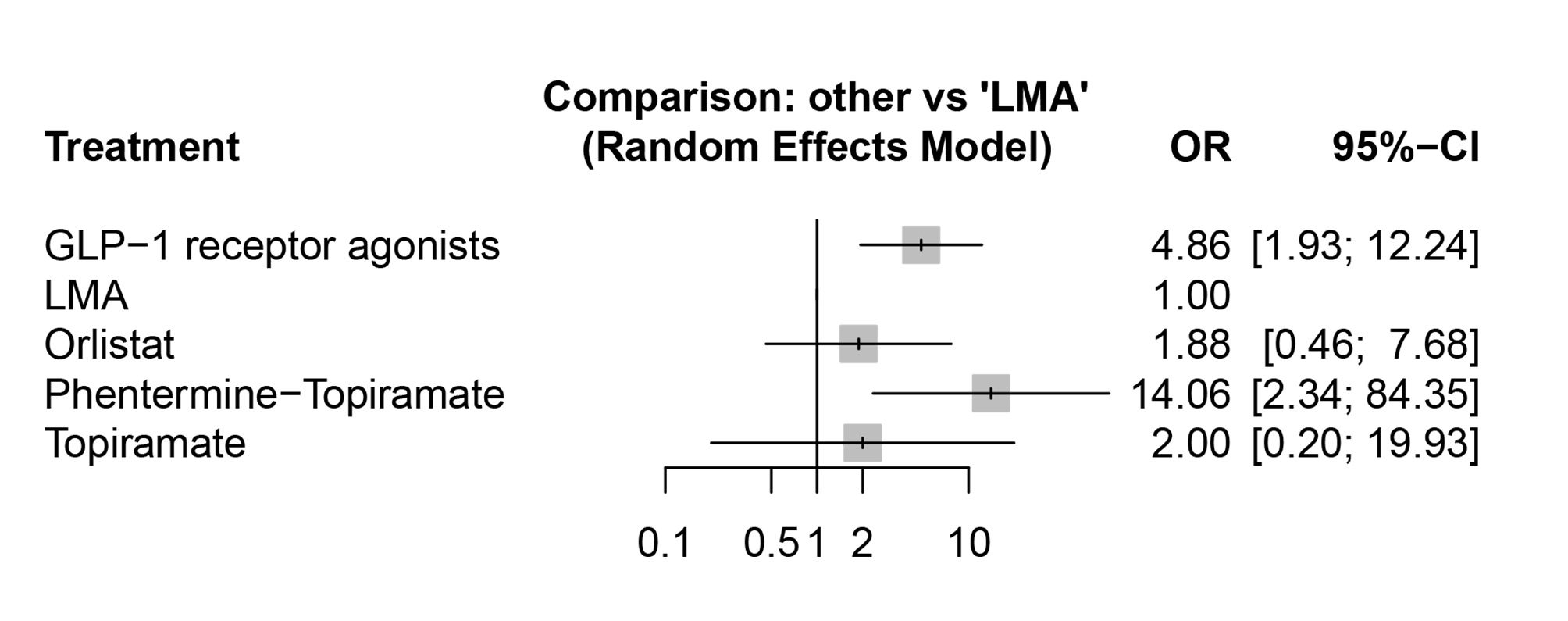

**Outcome:** **percentage of participants achieving BMI reduction of at least 10%**
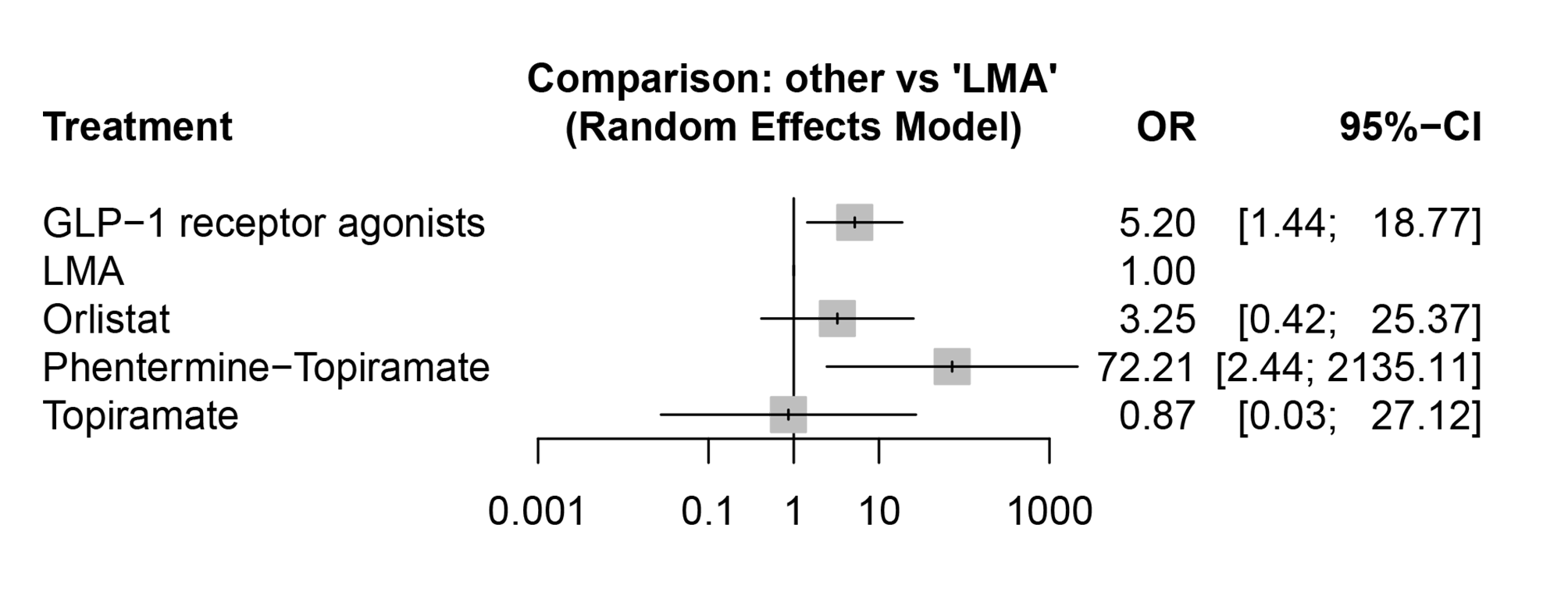

**Outcome: change in BMI z-score from baseline**

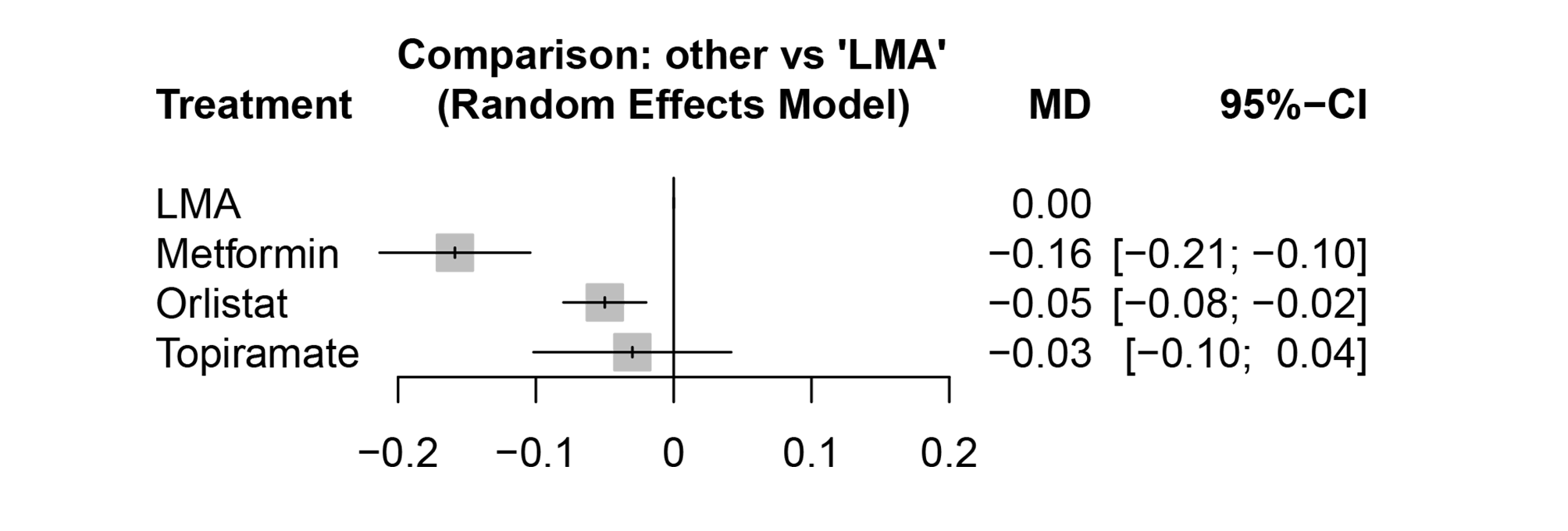

**Outcome: change in BMI SDS from baseline**

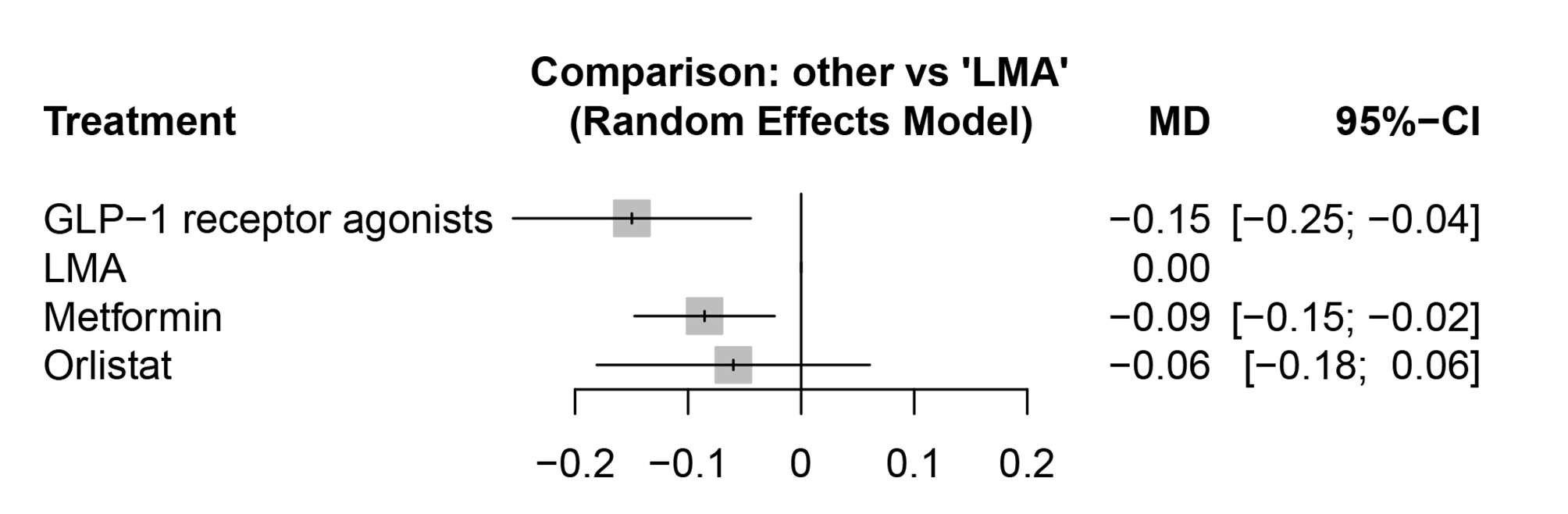

**Outcome: total gastrointestinal adverse events**

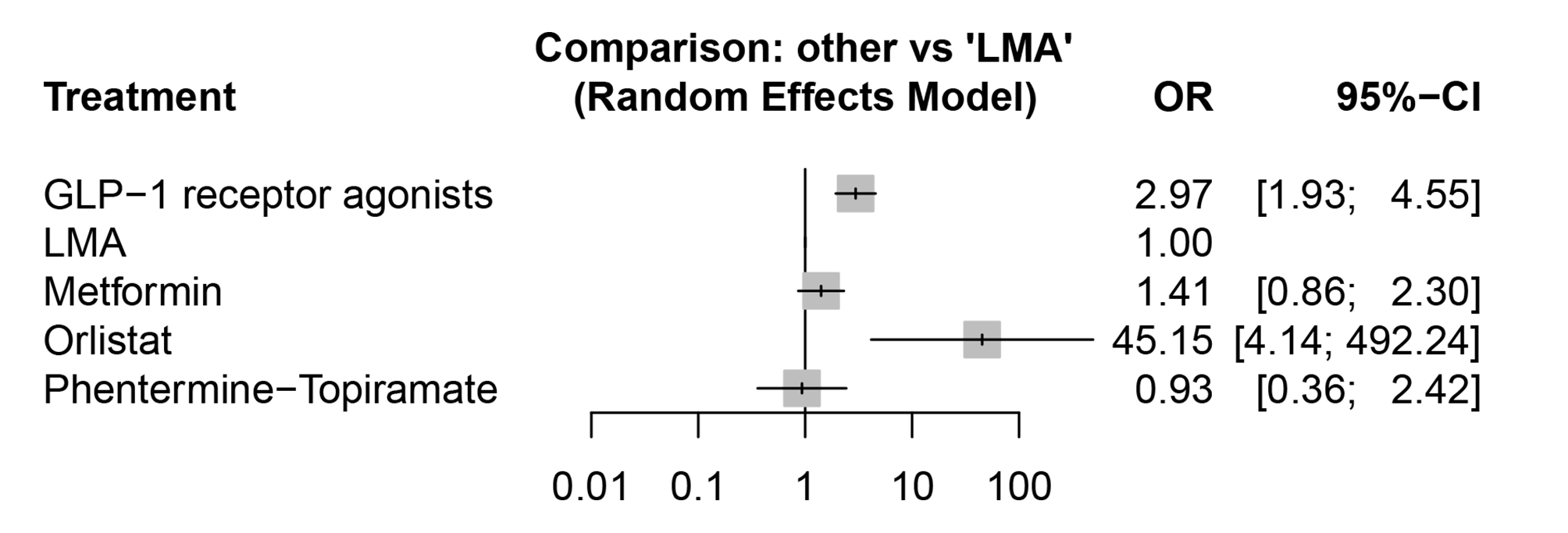

**Outcome: discontinuation due to any adverse event**

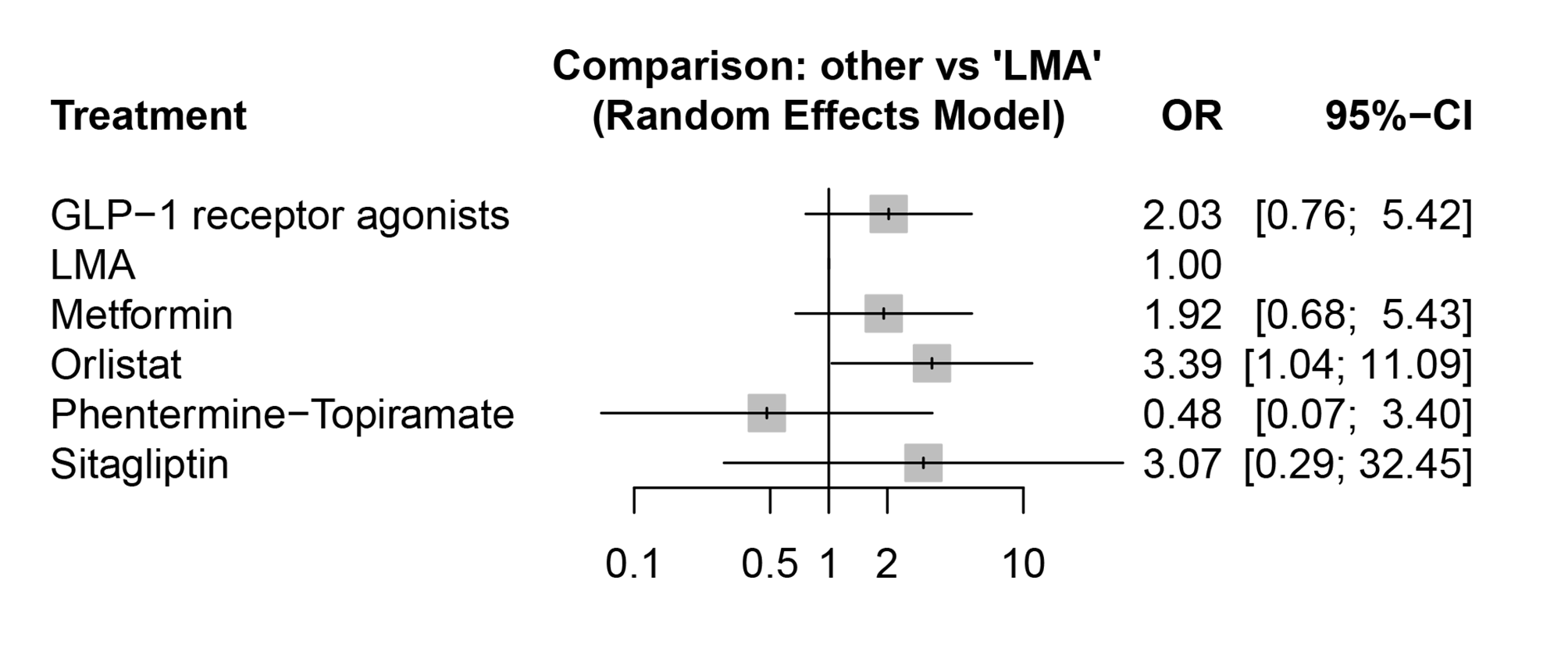

**Outcome: serious adverse events**

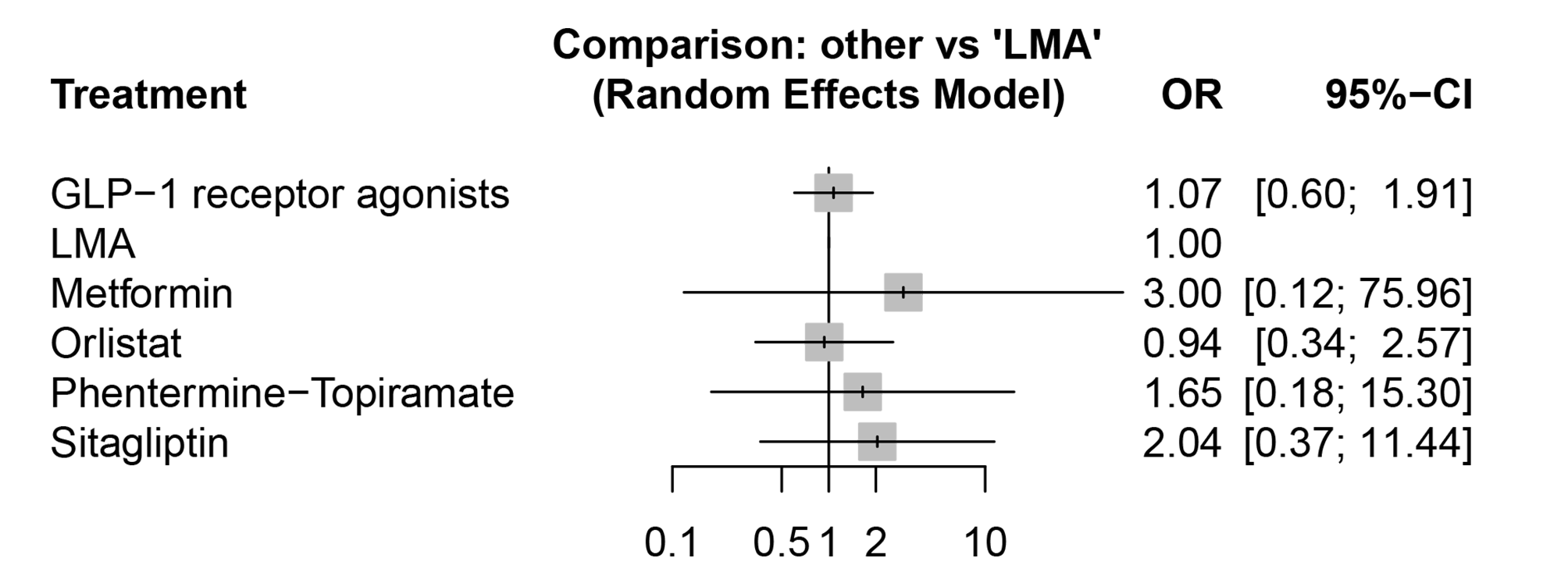

**Outcome: nausea events**

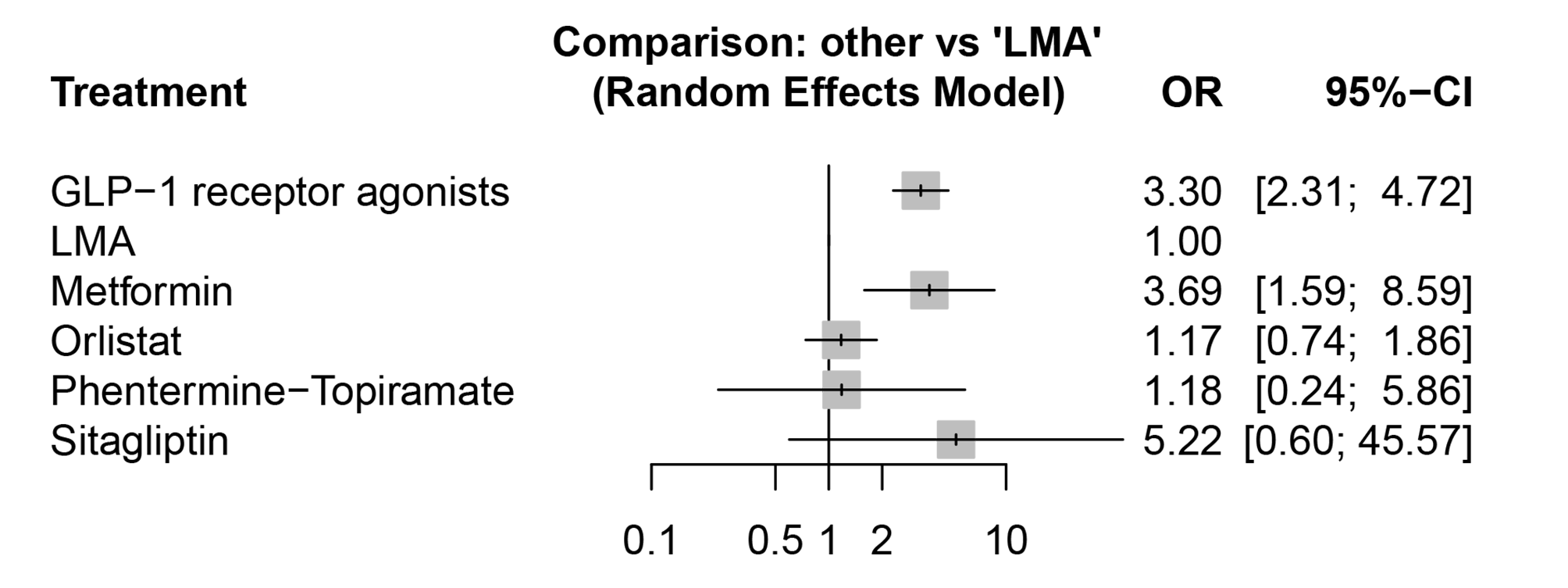

**Outcome: vomiting events**

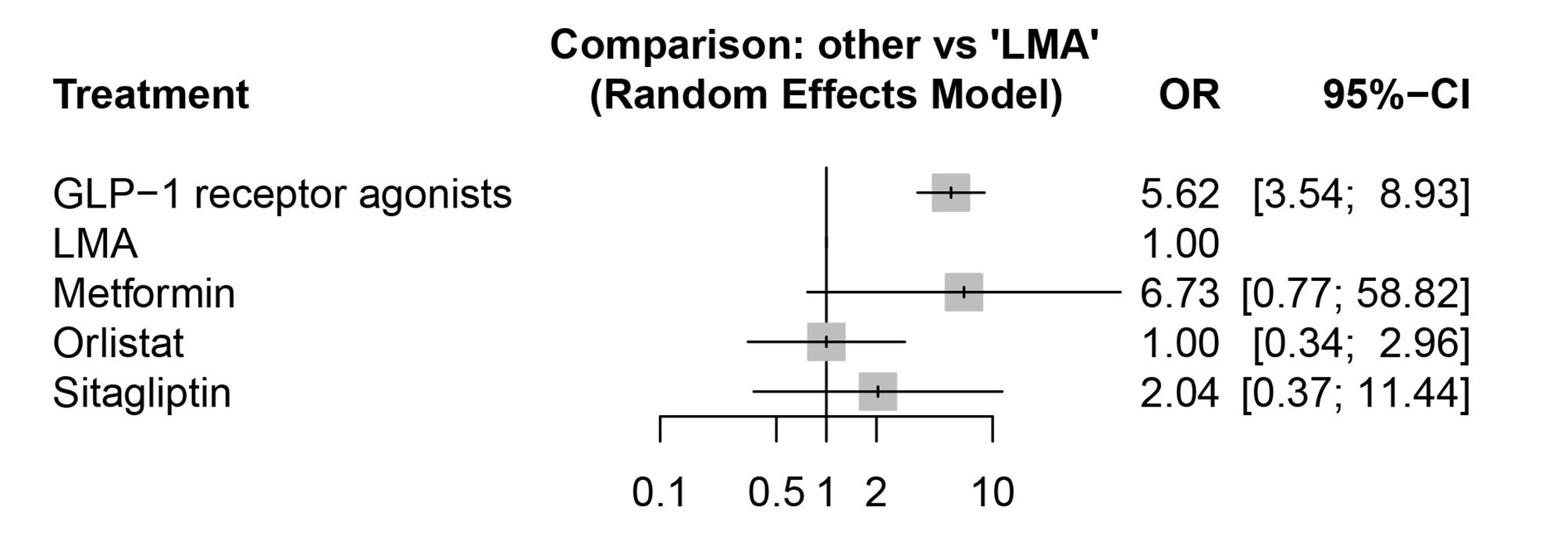

**Outcome: diarrhea events**

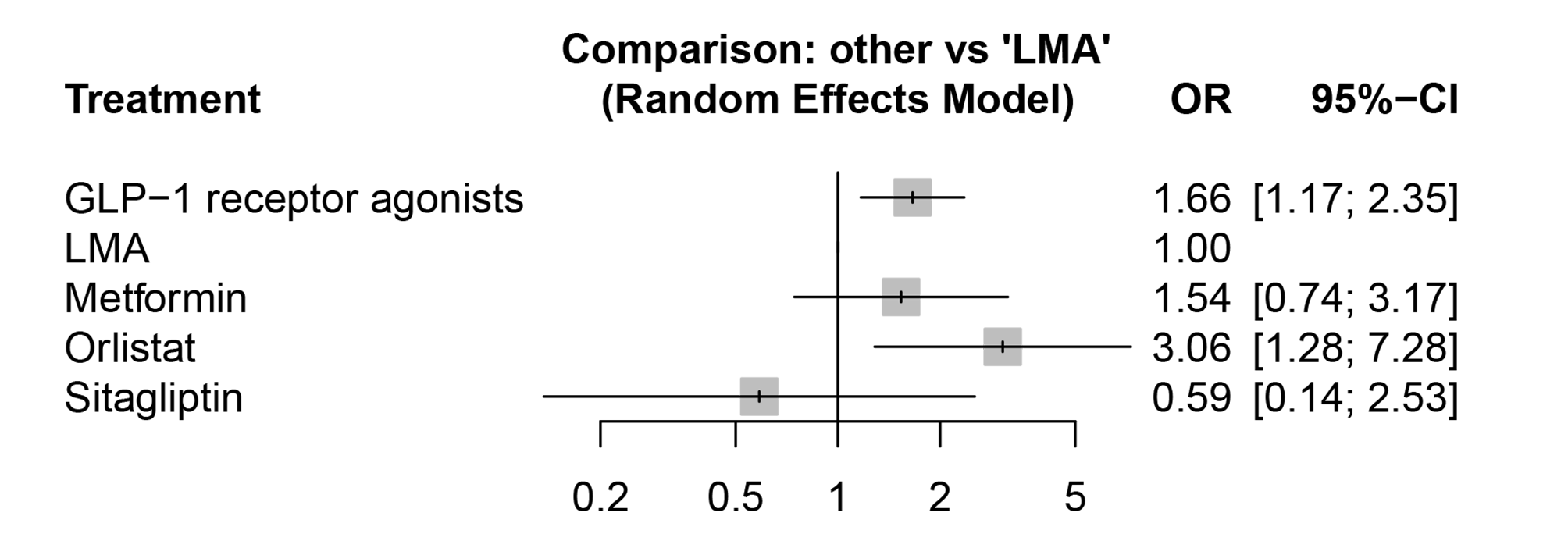

#### 6.5 Heterogeneity assessments

| **Outcomes** | **Design-based Q statistic, comparisons, and overall statement** | **Qstatistic** | **Degree of freedom** | **P value** |
| --- | --- | --- | --- | --- |
| Change in BMI from baseline | **Design-specific decomposition of within-designs Q statistic** | | | |
|  | Lifestyle modification alone vs Metformin | 173.06 | 16 | < 0.0001 |
|  | Lifestyle modification alone vs Orlistat | 18.17 | 3 | 0.0004 |
|  | GLP-1 receptor agonists vs Lifestyle modification alone | 8.13 | 4 | 0.0870 |
|  | **Between-designs Q statistic after detaching of single designs** | | | |
|  | Lifestyle modification alone vs Metformin | 0.00 | 0 | -- |
|  | Q statistic to assess consistency under the assumption of a full design-by-treatment interaction random effects model: between designs Q statistic, 4.17; degree of freedom, 1; p value, 0.0411; tau.within, 0.8920; tau^2^.within, 0.7956. | | | |
| Change in weight from baseline | **Design-specific decomposition of within-designs Q statistic** | | | |
|  | Lifestyle modification alone vs Metformin | 87.08 | 11 | < 0.0001 |
|  | Lifestyle modification alone vs Orlistat | 19.63 | 3 | 0.0002 |
|  | GLP-1 receptor agonists vs Lifestyle modification alone | 2.75 | 4 | 0.5998 |
|  | Q statistic to assess consistency under the assumption of a full design-by-treatment interaction random effects model: between designs Q statistic, 0.00; degree of freedom, 0; p value, --; tau.within, 2.4292; tau^2^.within, 5.9008. | | | |
| percentage of participants achieving BMI reduction of at least 5% | **Design-specific decomposition of within-designs Q statistic** | | | |
|  | GLP-1 receptor agonists vs Lifestyle modification alone | 7.31 | 2 | 0.0258 |
|  | Q statistic to assess consistency under the assumption of a full design-by-treatment interaction random effects model: between designs Q statistic, 0.00; degree of freedom, 0; p value, --; tau.within, 0.6777; tau^2^.within, 0.4593. | | | |
| percentage of participants achieving BMI reduction of at least 10% | **Design-specific decomposition of within-designs Q statistic** | | | |
|  | GLP-1 receptor agonists vs Lifestyle modification alone | 8.55 | 2 | 0.0139 |
|  | Q statistic to assess consistency under the assumption of a full design-by-treatment interaction random effects model: between designs Q statistic, 0.00; degree of freedom, 0; p value, --; tau.within, 0.9711; tau^2^.within, 0.9431. | | | |
| Change in BMI z-score from baseline | **Design-specific decomposition of within-designs Q statistic** | | | |
|  | Lifestyle modification alone vs Metformin | 3.122 | 3 | 0.3732 |
|  | Q statistic to assess consistency under the assumption of a full design-by-treatment interaction random effects model: between designs Q statistic, 0.00; degree of freedom, 0; p value, --; tau.within, 0.0130; tau^2^.within, 0.0002. | | | |
| Change in BMI SDS from baseline | **Design-specific decomposition of within-designs Q statistic** | | | |
|  | Lifestyle modification alone vs Metformin | 13.56 | 5 | 0.0186 |
|  | GLP-1 receptor agonists vs Lifestyle modification alone | 2.45 | 2 | 0.2939 |
|  | Q statistic to assess consistency under the assumption of a full design-by-treatment interaction random effects model: between designs Q statistic, 0.00; degree of freedom, 0; p value, --; tau.within, 0.0547; tau^2^.within, 0.0030. | | | |
| Nausea events | **Design-specific decomposition of within-designs Q statistic** | | | |
|  | Lifestyle modification alone vs Metformin | 1.72 | 3 | 0.6323 |
|  | GLP-1 receptor agonists vs Lifestyle modification alone | 3.31 | 7 | 0.8553 |
|  | Lifestyle modification alone vs Orlistat | 0.01 | 1 | 0.9162 |
|  | Q statistic to assess consistency under the assumption of a full design-by-treatment interaction random effects model: between designs Q statistic, 0.00; degree of freedom, 0; p value, --; tau.within, 0; tau^2^.within, 0. | | | |
| Vomiting events | **Design-specific decomposition of within-designs Q statistic** | | | |
|  | GLP-1 receptor agonists vs Lifestyle modification alone | 4.29 | 8 | 0.8296 |
|  | Q statistic to assess consistency under the assumption of a full design-by-treatment interaction random effects model: between designs Q statistic, 0.00; degree of freedom, 0; p value, --; tau.within, 0; tau^2^.within, 0. | | | |
| Diarrhea events | **Design-specific decomposition of within-designs Q statistic** | | | |
|  | GLP-1 receptor agonists vs Lifestyle modification alone | 3.77 | 7 | 0.8062 |
|  | Lifestyle modification alone vs Metformin | 0.10 | 3 | 0.9916 |
|  | Q statistic to assess consistency under the assumption of a full design-by-treatment interaction random effects model: between designs Q statistic, 0.00; degree of freedom, 0; p value, --; tau.within, 0; tau^2^.within, 0. | | | |
| Total gastrointestinal  adverse events | **Design-specific decomposition of within-designs Q statistic** | | | |
|  | Lifestyle modification alone vs Orlistat | 5.26 | 1 | 0.0218 |
|  | Lifestyle modification alone vs Phentermine-Topiramate | 1.47 | 1 | 0.2248 |
|  | Lifestyle modification alone vs Metformin | 5.88 | 7 | 0.5534 |
|  | GLP-1 receptor agonists vs Lifestyle modification alone | 4.59 | 6 | 0.5969 |
|  | Q statistic to assess consistency under the assumption of a full design-by-treatment interaction random effects model: between designs Q statistic, 0.00; degree of freedom, 0; p value, --; tau.within, 0.2404; tau^2^.within, 0.0578. | | | |
| Serious adverse events | **Design-specific decomposition of within-designs Q statistic** | | | |
|  | Lifestyle modification alone vs Orlistat | 1.13 | 1 | 0.2868 |
|  | GLP-1 receptor agonists vs Lifestyle modification alone | 5.14 | 6 | 0.5255 |
|  | Lifestyle modification alone vs Phentermine-Topiramate | 0.00 | 1 | 0.9734 |
|  | Q statistic to assess consistency under the assumption of a full design-by-treatment interaction random effects model: between designs Q statistic, 0.00; degree of freedom, 0; p value, --; tau.within, 0; tau^2^.within, 0. | | | |
| Discontinuation due to any adverse event | **Design-specific decomposition of within-designs Q statistic** | | | |
|  | Lifestyle modification alone vs Phentermine-Topiramate | 1.98 | 1 | 0.1598 |
|  | Lifestyle modification alone vs Orlistat | 2.04 | 2 | 0.3600 |
|  | GLP-1 receptor agonists vs Lifestyle modification alone | 4.98 | 5 | 0.4182 |
|  | Lifestyle modification alone vs Metformin | 4.78 | 5 | 0.4438 |
|  | Q statistic to assess consistency under the assumption of a full design-by-treatment interaction random effects model: between designs Q statistic, 0.00; degree of freedom, 0; p value, --; tau.within, 0.3077; tau^2^.within, 0.0947. | | | |

#### 6.6 Transitivity assessments

Transitivity covers the validity of logical inferences [8] and represents a crucial condition for ensuring the accuracy of the network estimate results. It is appropriate to use network meta-analysis if the assumption of transitivity can be satisfied [9]. To assess the transitivity assumption, we compared the distribution of the possible effect modifiers across intervention comparisons, such like mean age at baseline, mean BMI at baseline and the proportion of males.

In addition, we considered for the effect of lifestyle modification alone groups in all outcomes to assess transitivity (i.e. change in BMI and weight from baseline, percentage of participants achieving BMI reduction of at least 5%, percentage of participants achieving BMI reduction of at least 10%, change in BMI z-score and BMI SDS from baseline, proportion of total gastrointestinal adverse events, serious adverse events and discontinuation due to any adverse event, the incidence of adverse events such as nausea, vomiting and diarrhea.)

**Note:** LMA, Lifestyle modification alone; GLP-1, glucagon-like peptide-1; Metformin_Fluoxetine, Metformin combined with Fluoxetine.

**Effect of lifestyle modification alone on change in BMI from baseline**

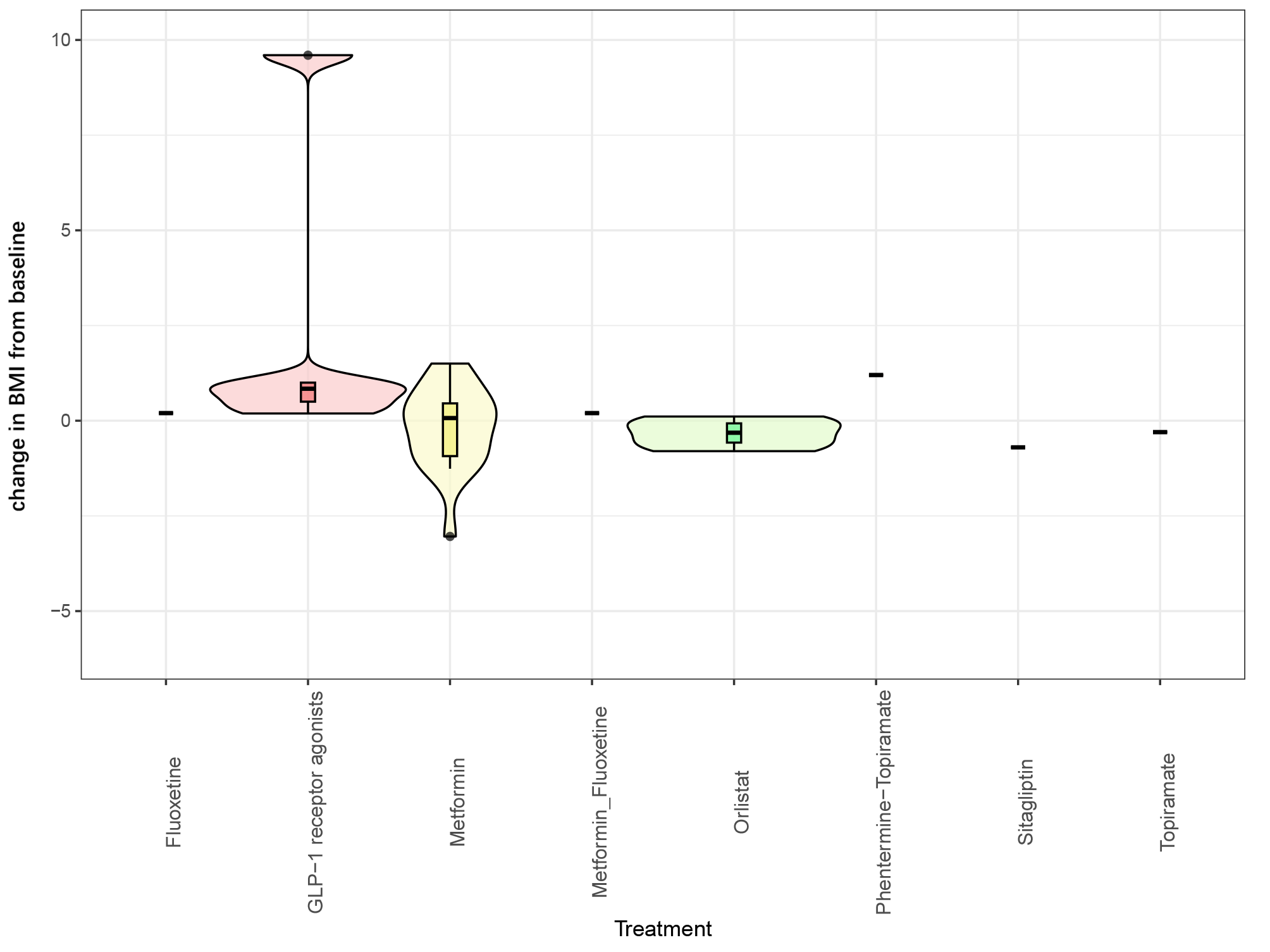

**Effect of lifestyle modification alone on change in weight from baseline**

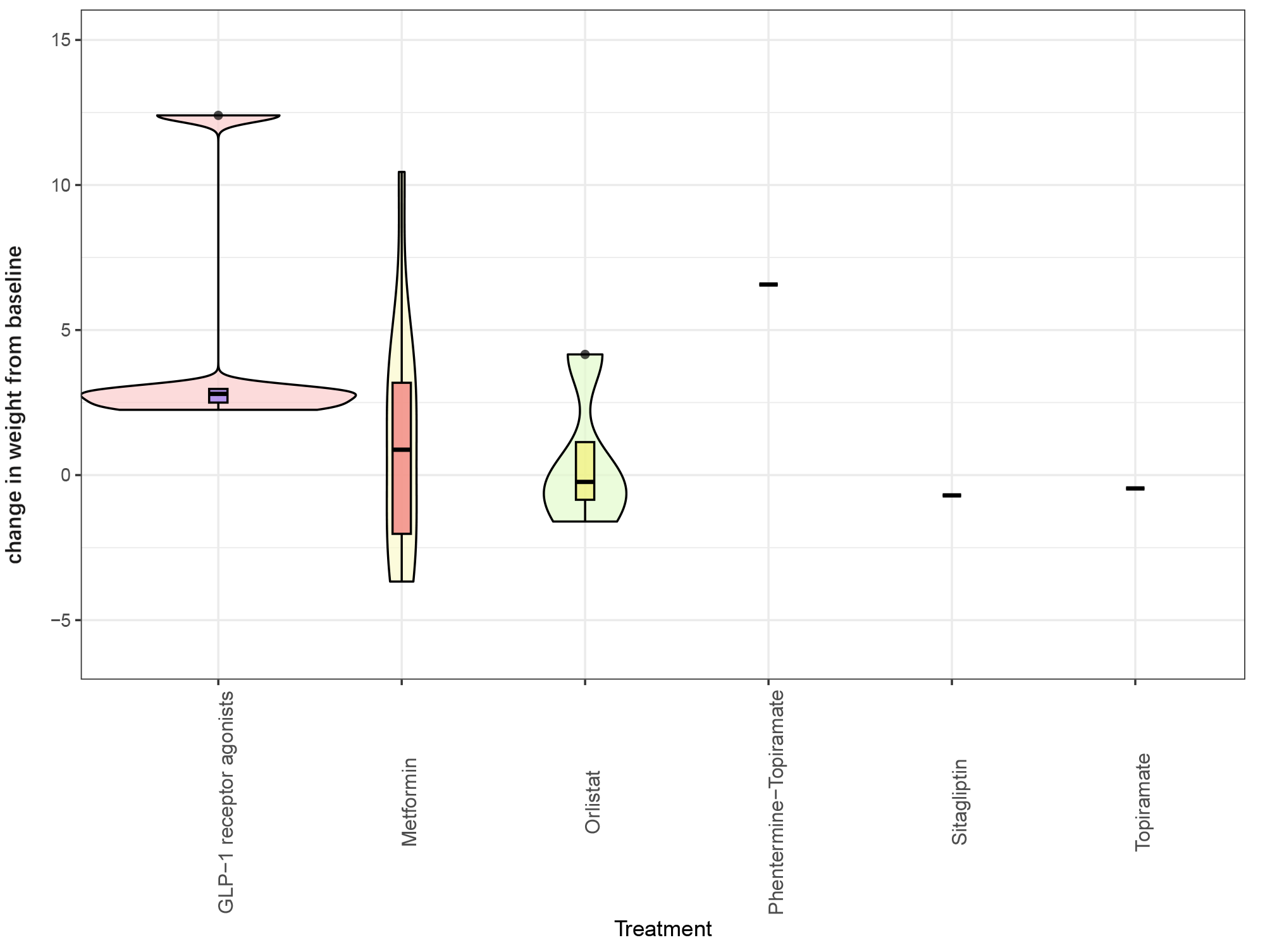

**Effect of lifestyle modification alone on percentage of participants achieving BMI reduction of at least 5%**

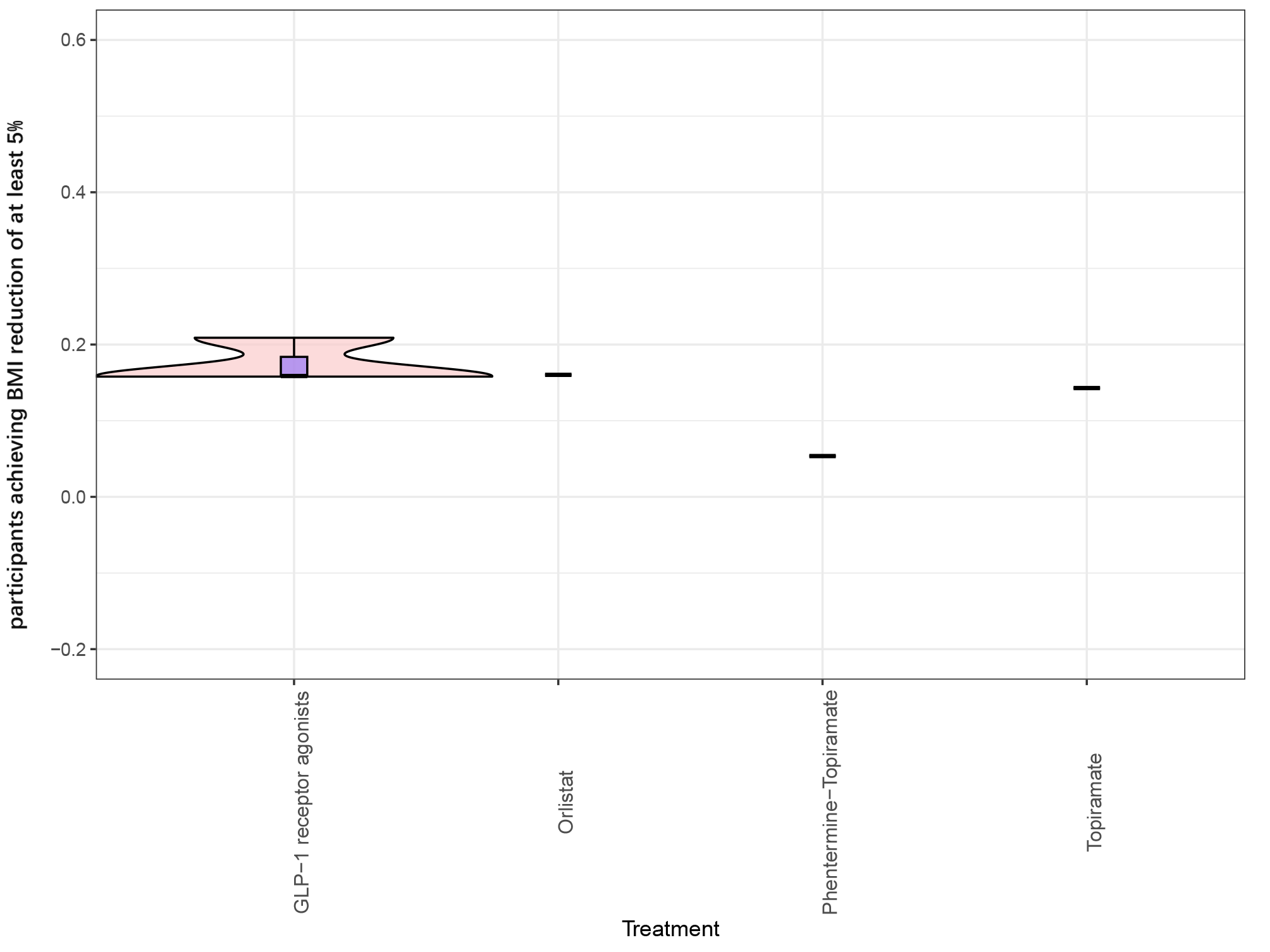

**Effect of lifestyle modification alone on percentage of participants achieving BMI reduction of at least 10%**

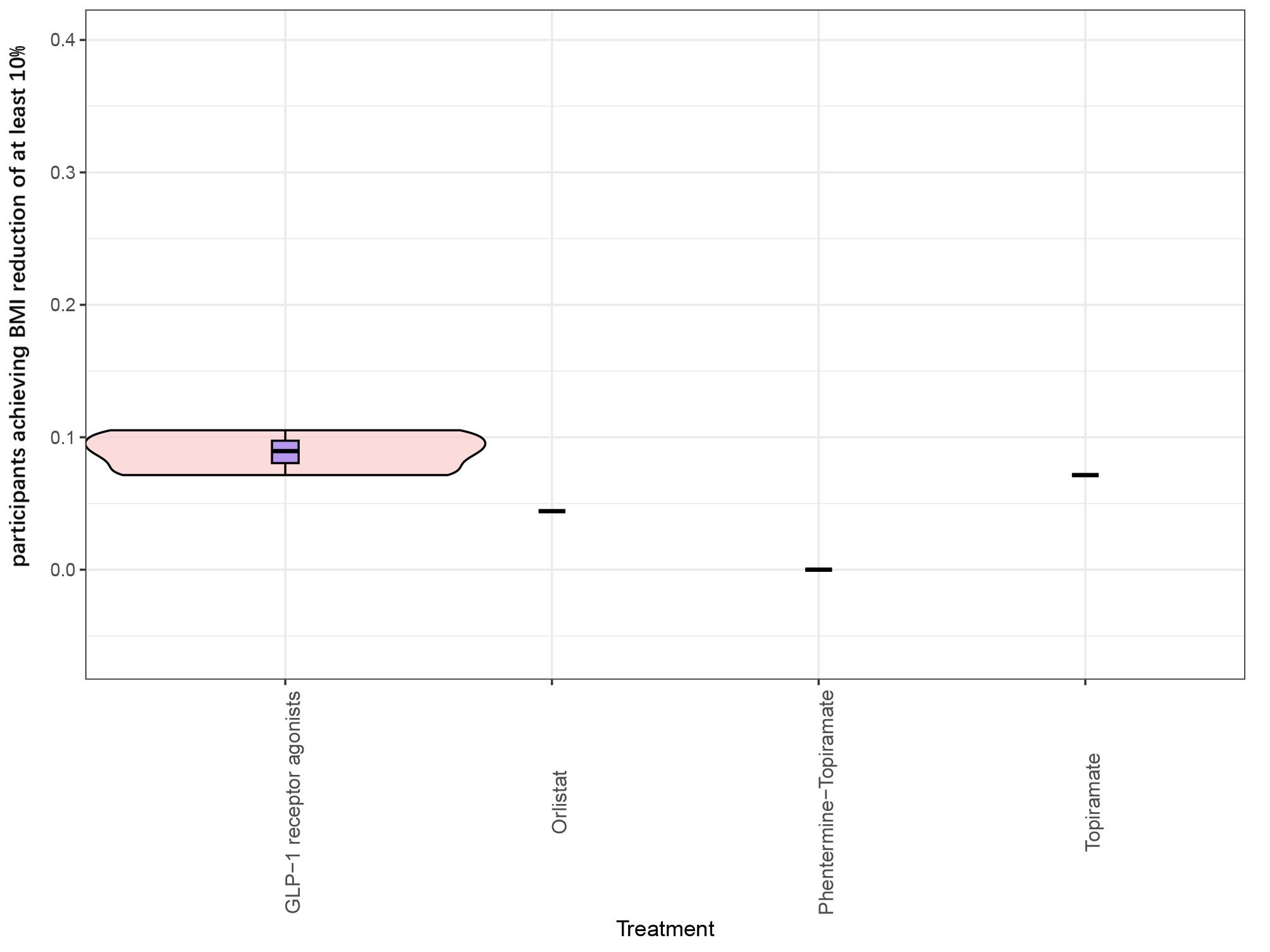

**Effect of lifestyle modification alone on change in BMI z-score from baseline**

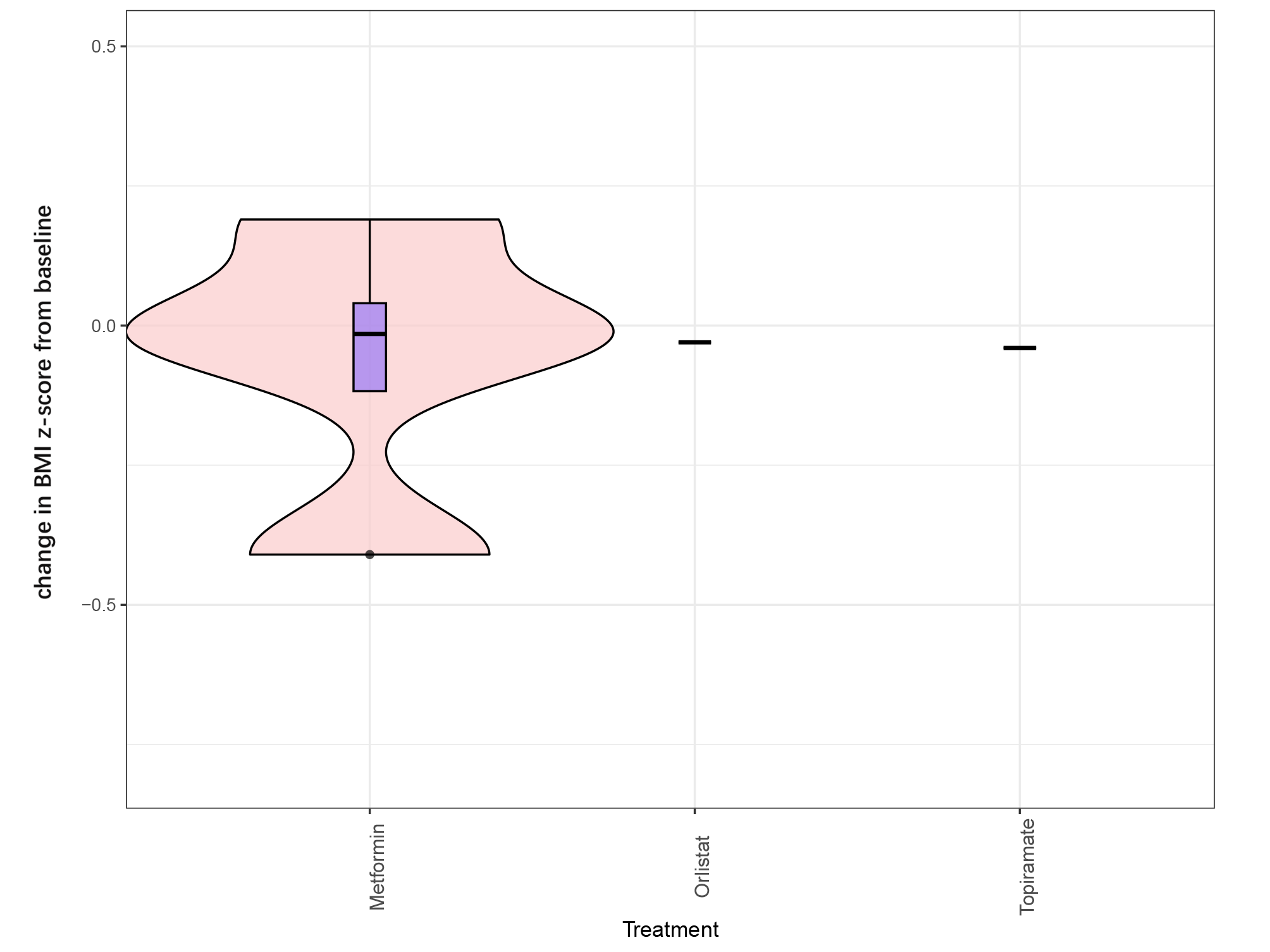

**Effect of lifestyle modification alone on change in BMI SDS from baseline**

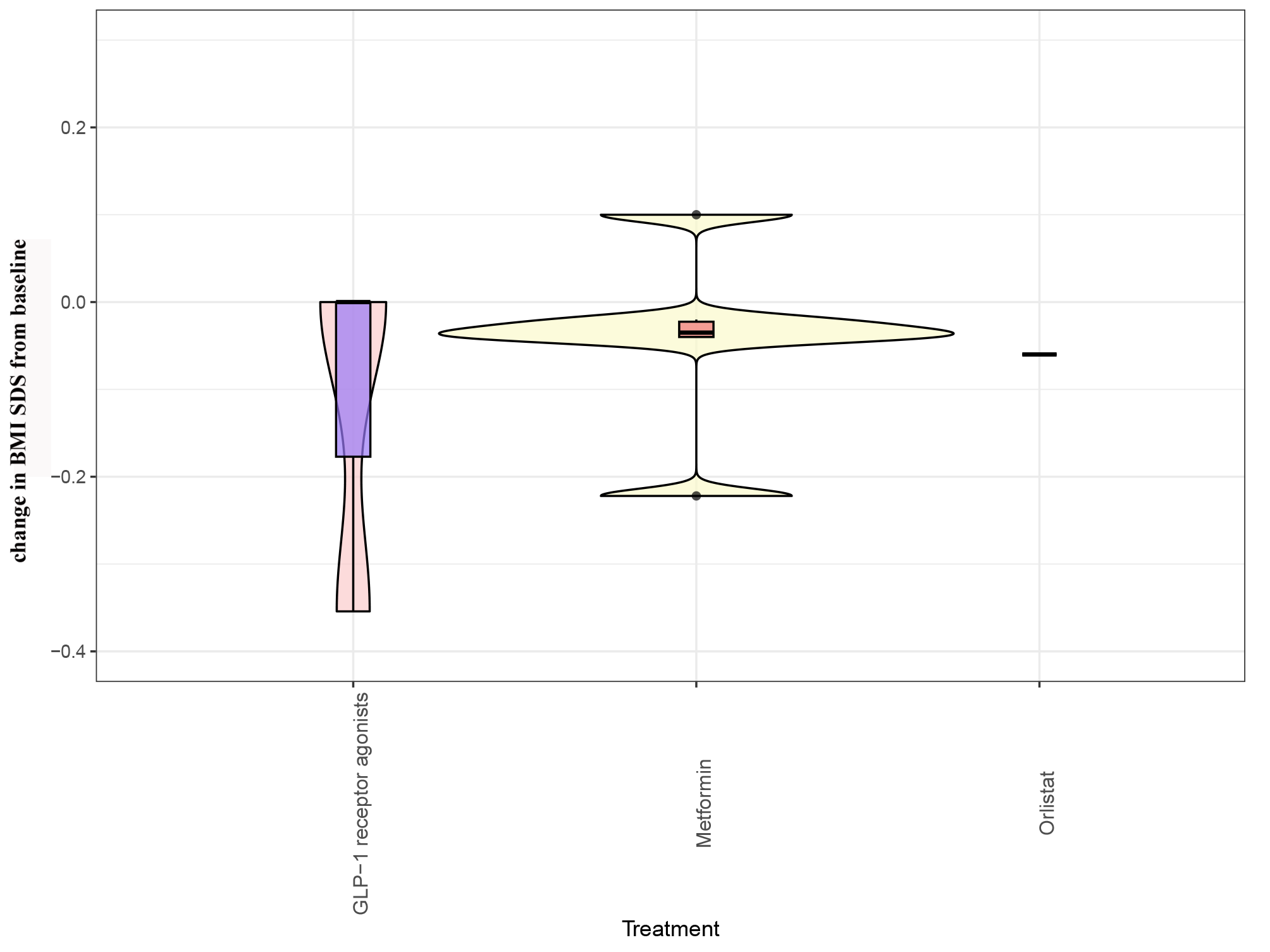

**Effect of lifestyle modification alone on total gastrointestinal adverse events**

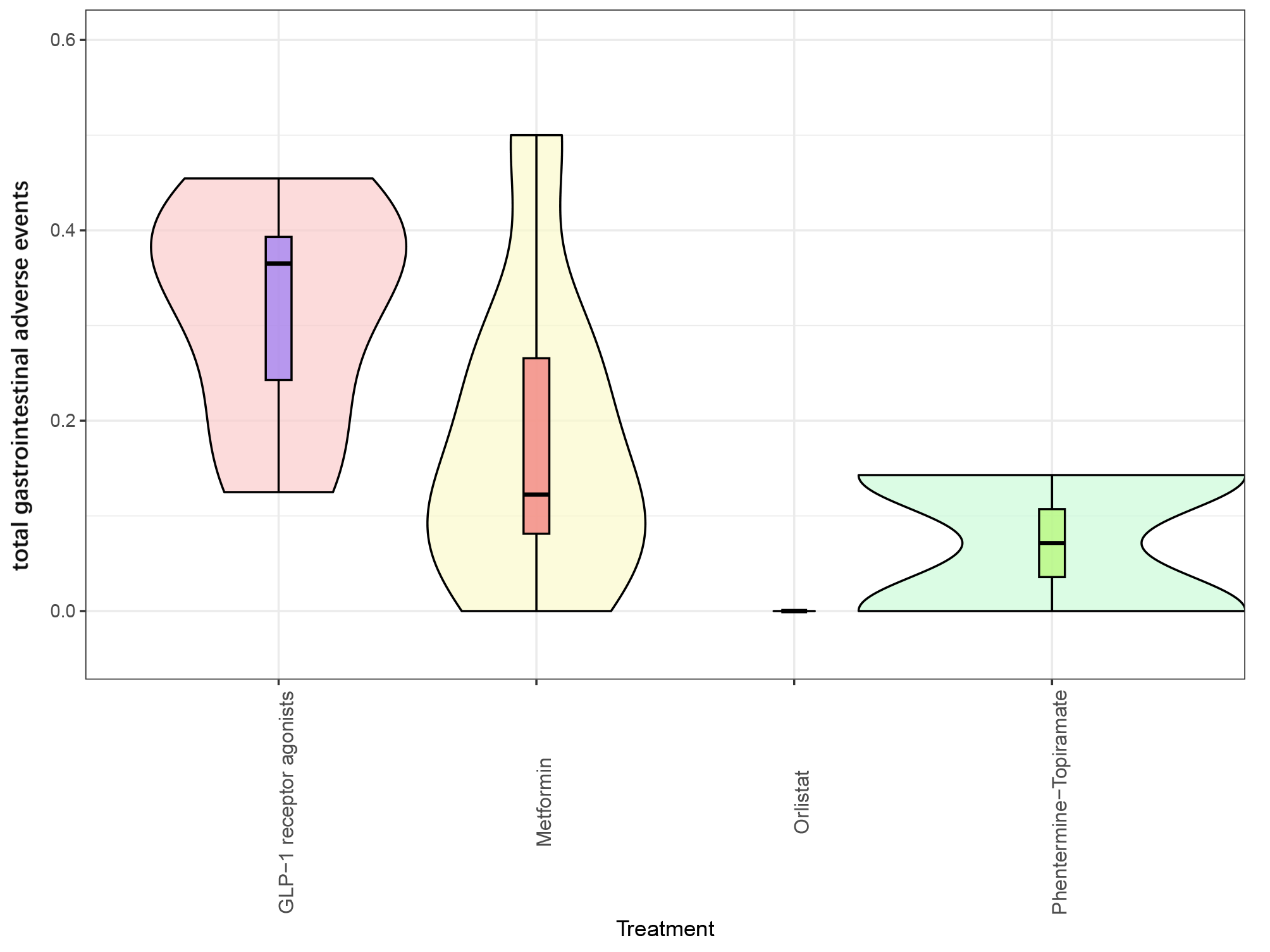

**Effect of lifestyle modification alone on discontinuation due to any adverse event**

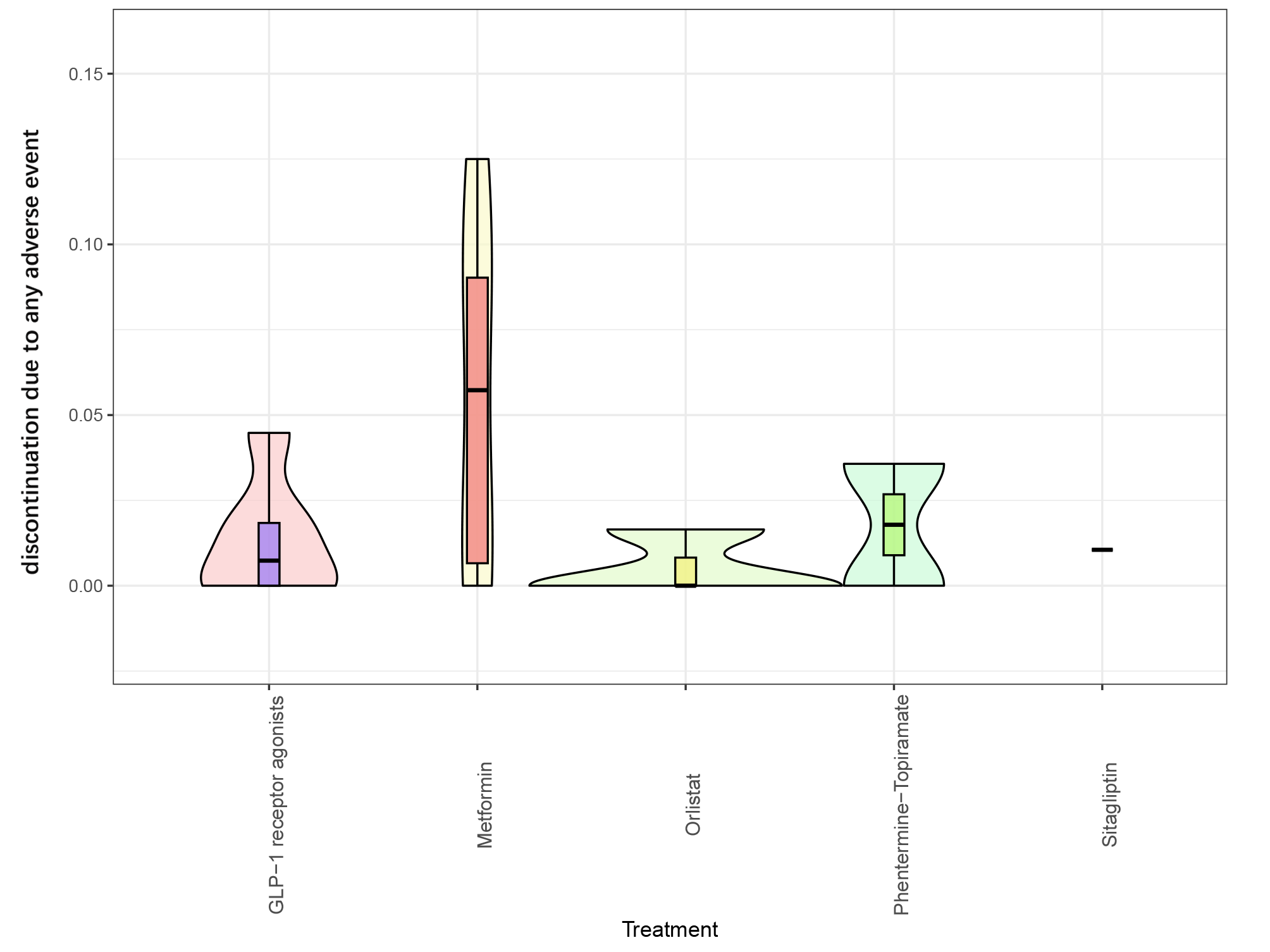

**Effect of lifestyle modification alone on serious adverse events**

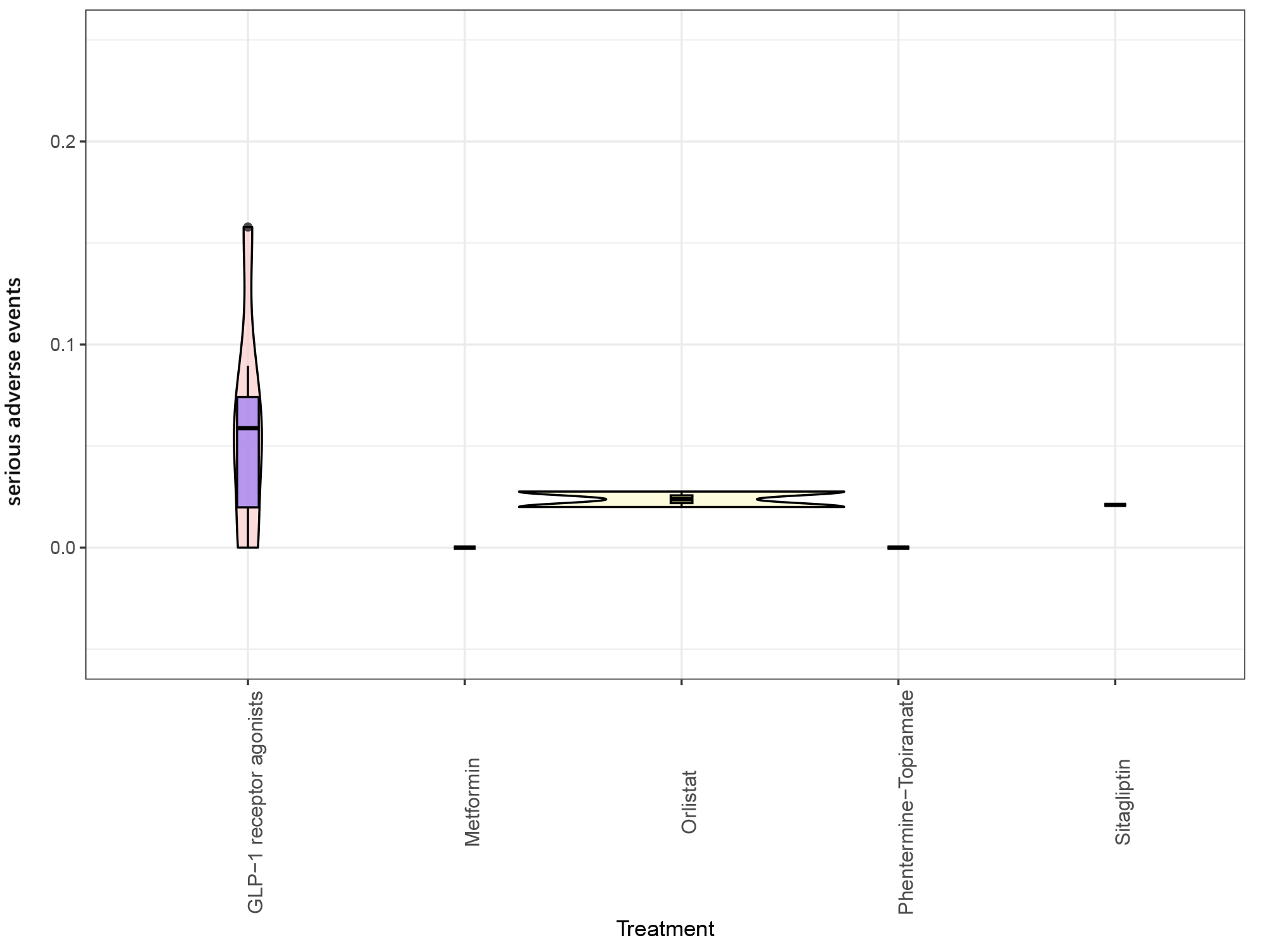

**Effect of lifestyle modification alone on nausea events**

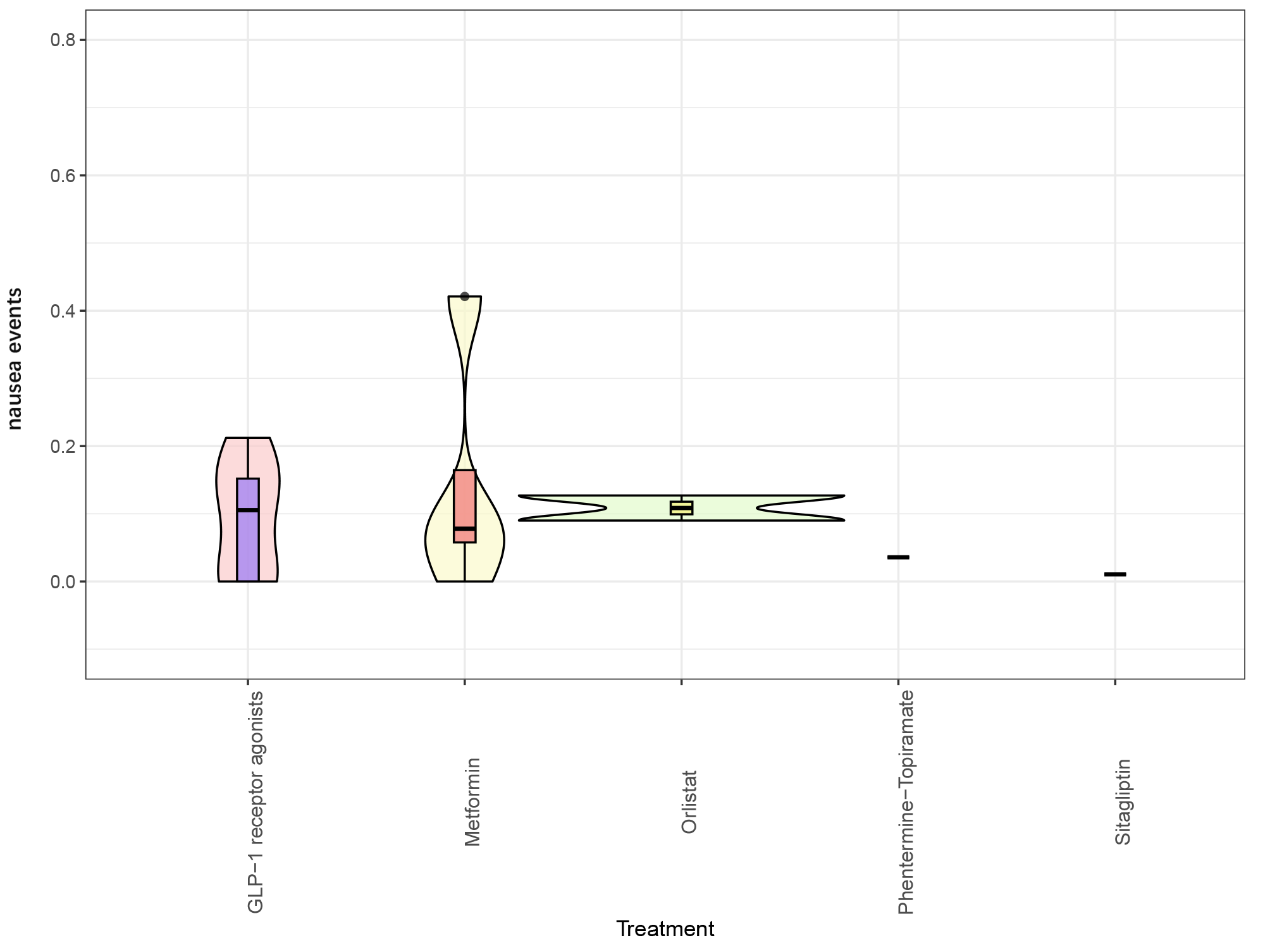

**Effect of lifestyle modification alone on vomiting events**

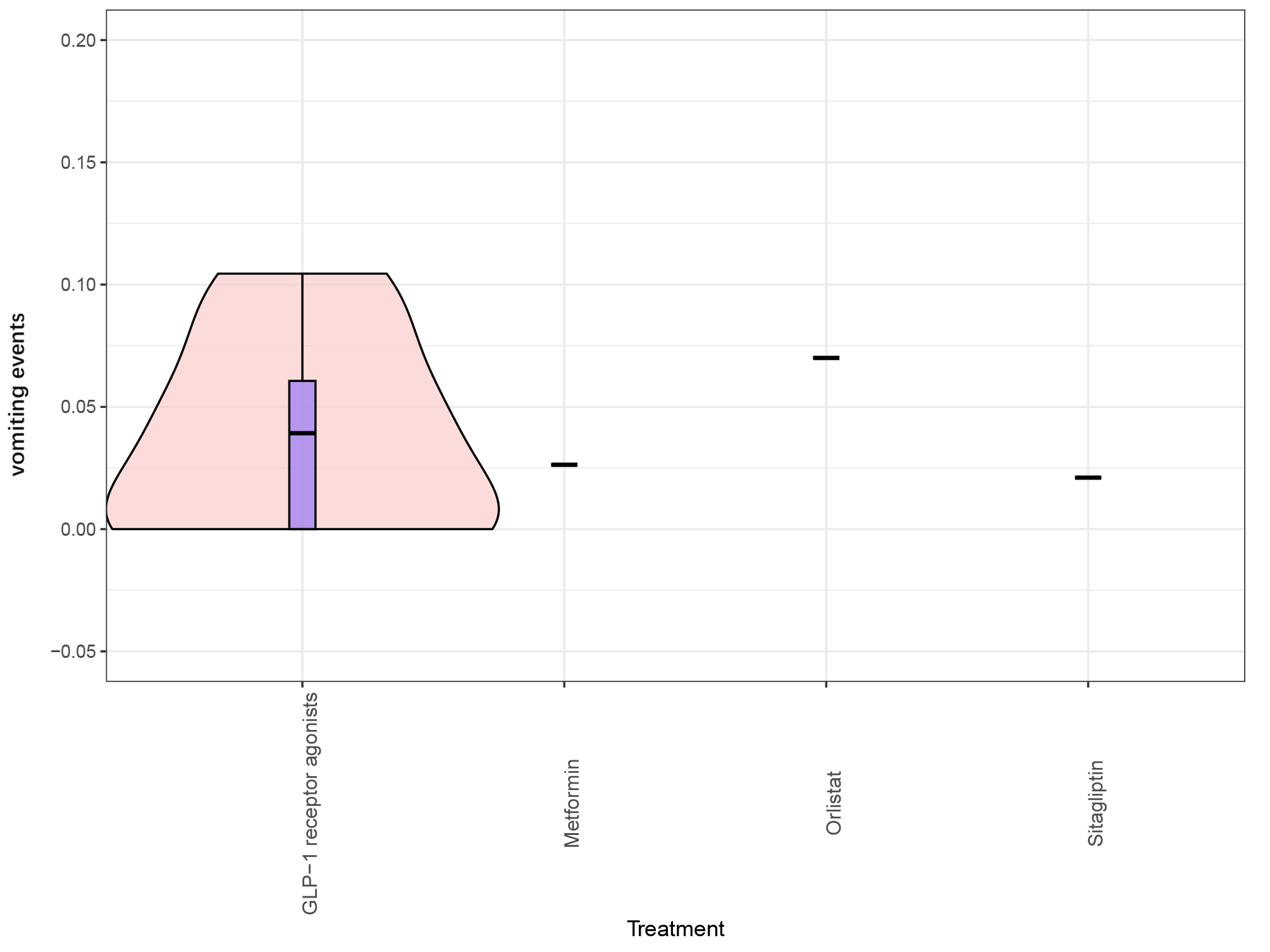

**Effect of lifestyle modification alone on diarrhea events**

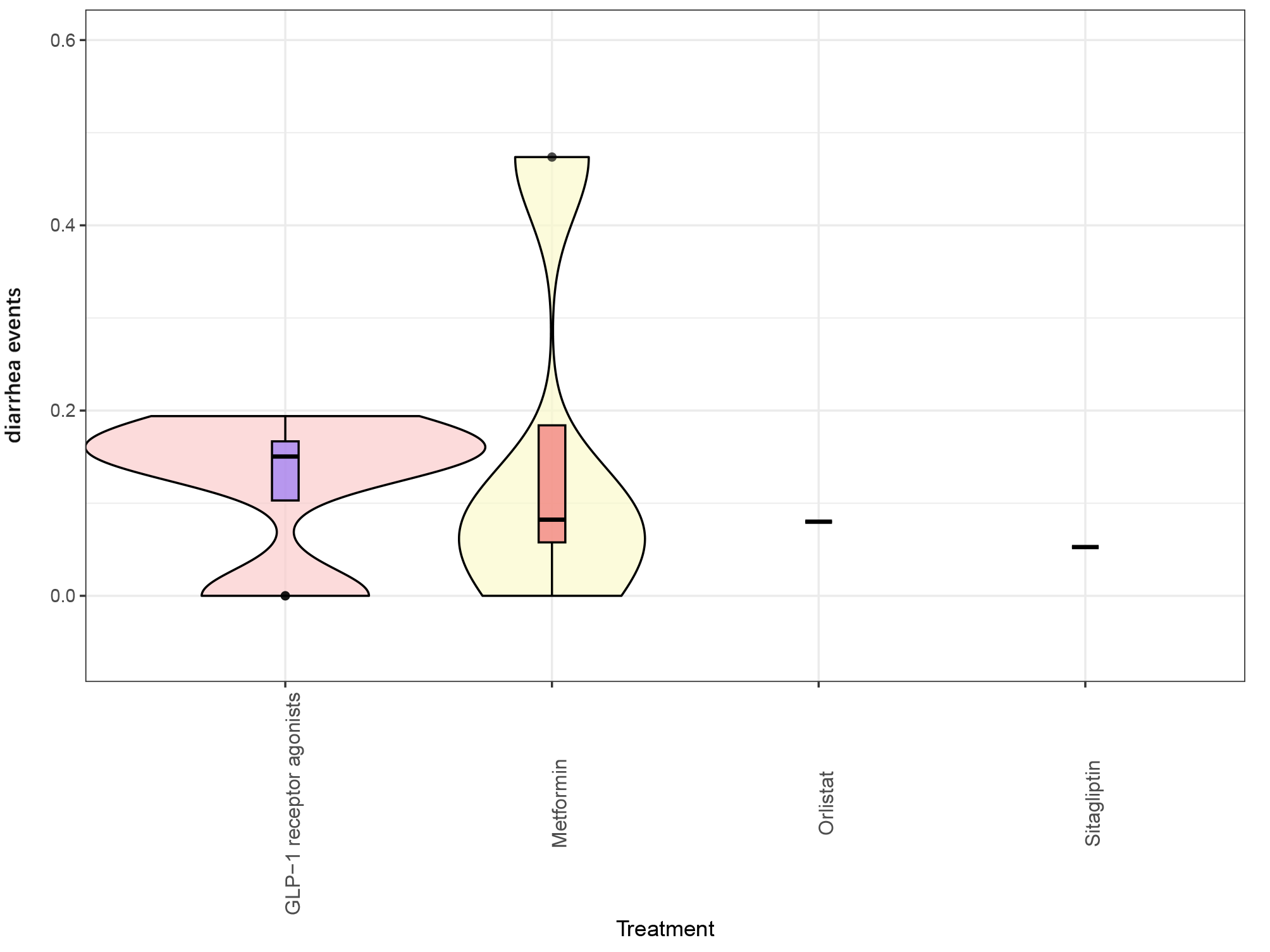

**Mean age at baseline**

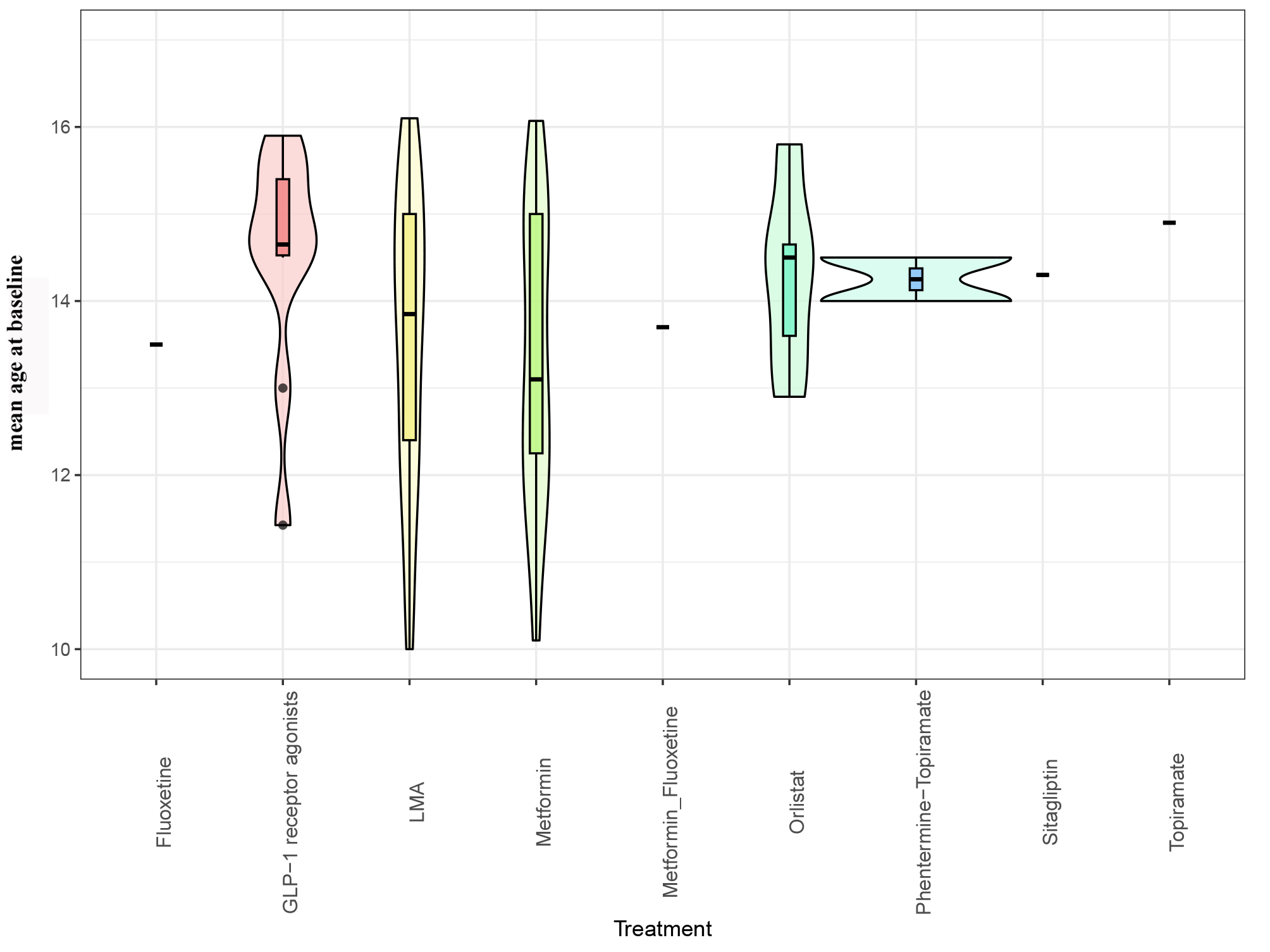

**Mean BMI at baseline**

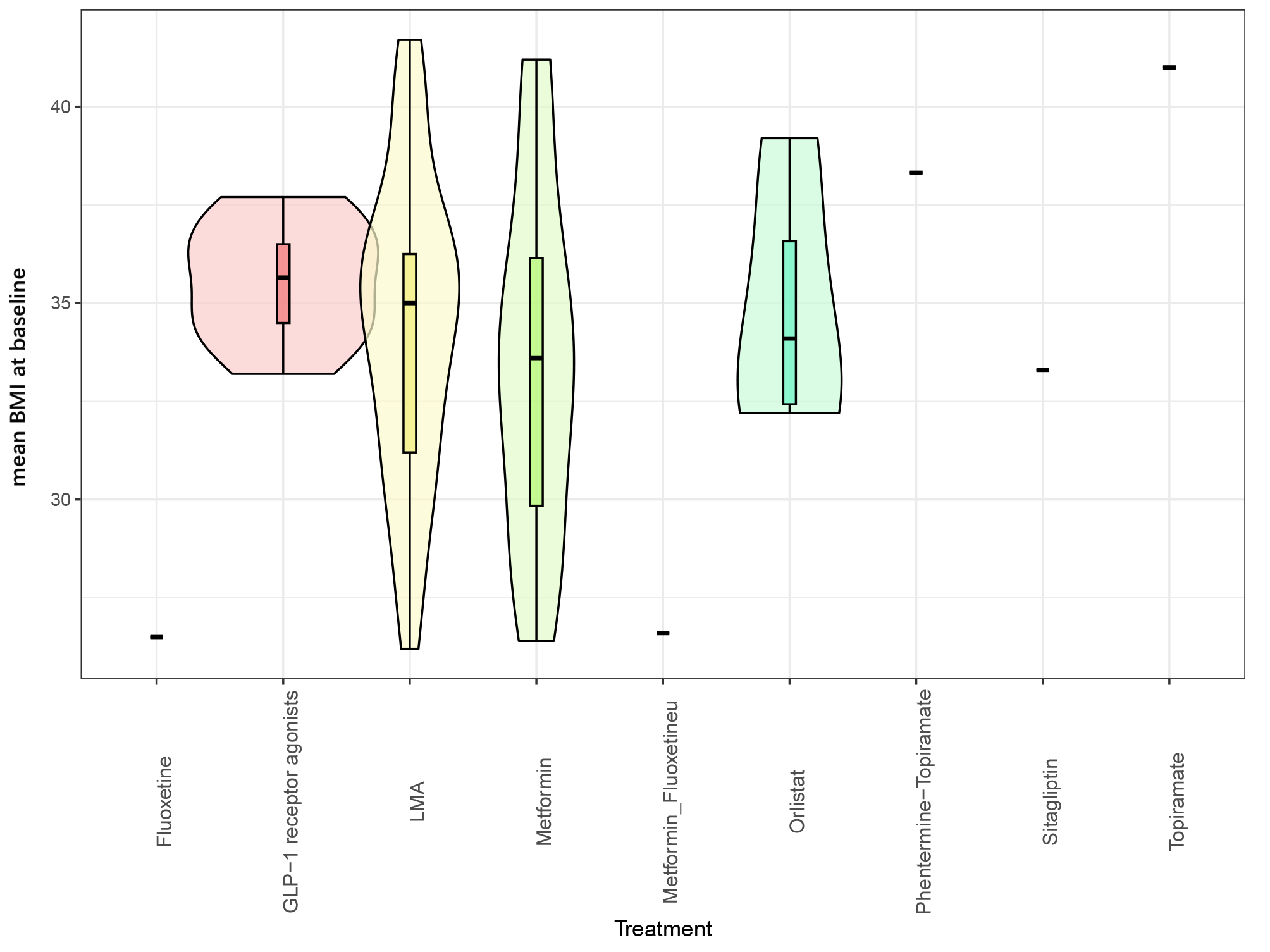

**Proportion of males**

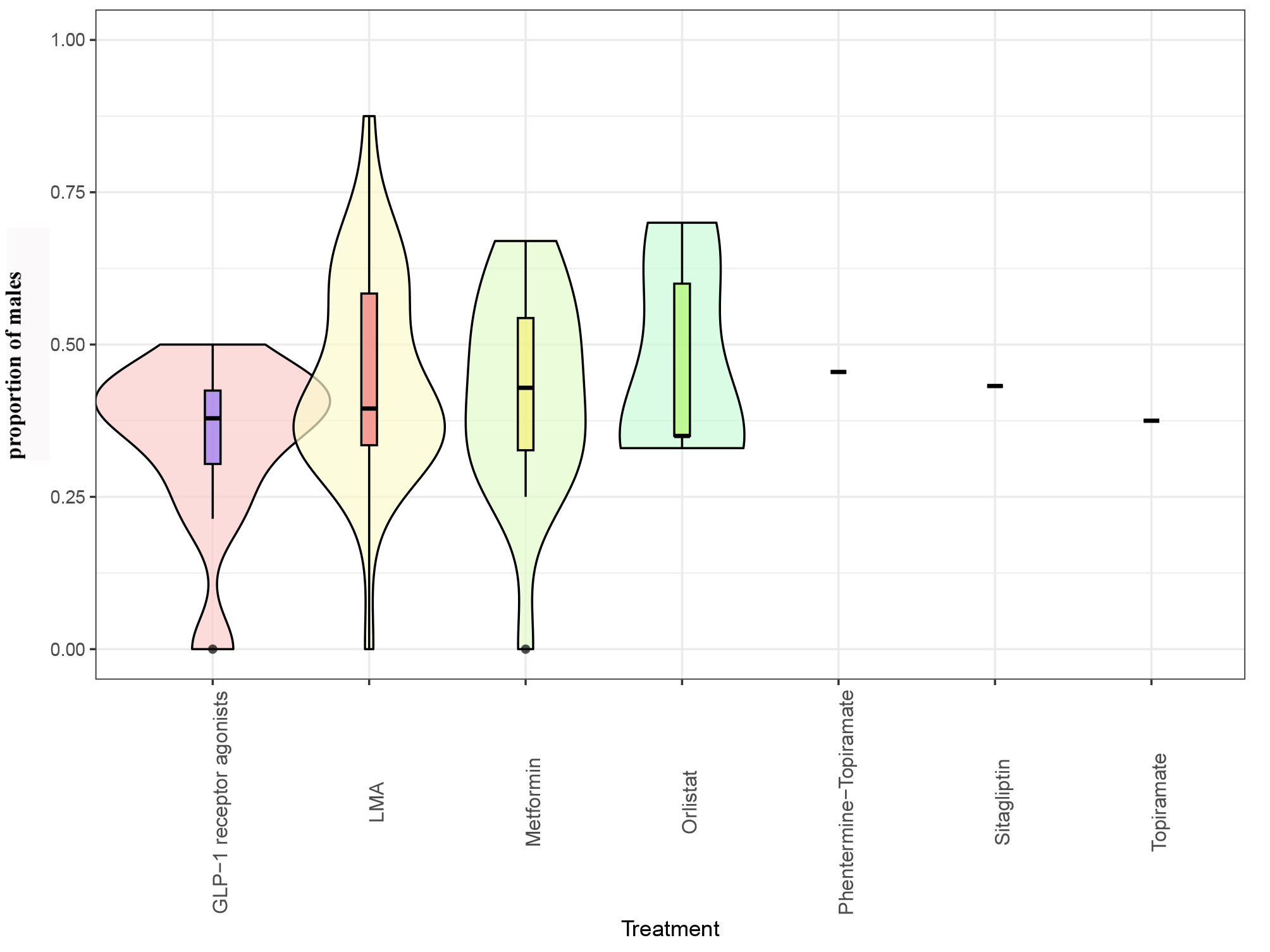

#### 6.7 Inconsistency assessments

**change in BMI from baseline**

| **Comparison** | **Direct estimate** | **Indirect estimate** | **Network estimate** | **P-value** |
| --- | --- | --- | --- | --- |
| Fluoxetine vs Lifestyle modification alone | -0.80 (-3.42, 1.82) | -4.66 (-9.43, 0.10) | -1.70 (-4.00, 0.60) | 0.77 |
| Fluoxetine vs Metformin | -1.5 (-4.13, 1.13) | 2.36 (-2.40, 7.13) | -0.60 (-2.90, 1.70) | 0.77 |
| Metformin vs Metformin combined with Fluoxetine | 1.80 (-0.82, 4.42) | -2.06 (-6.83, 2.70) | 0.90 (-1.40, 3.20) | 0.77 |
| Metformin combined with Fluoxetine vs Lifestyle modification alone | -1.10 (-3.72, 1.52) | -4.96 (-9.73, -0.20) | -2.00 (-4.30, 0.30) | 0.77 |

#### 6.8 Funnel plots

**Note:** LMA, Lifestyle modification alone; GLP-1, glucagon-like peptide-1; Metformin_Fluoxetine, Metformin combined with Fluoxetine.

**Outcome: change in BMI from baseline**

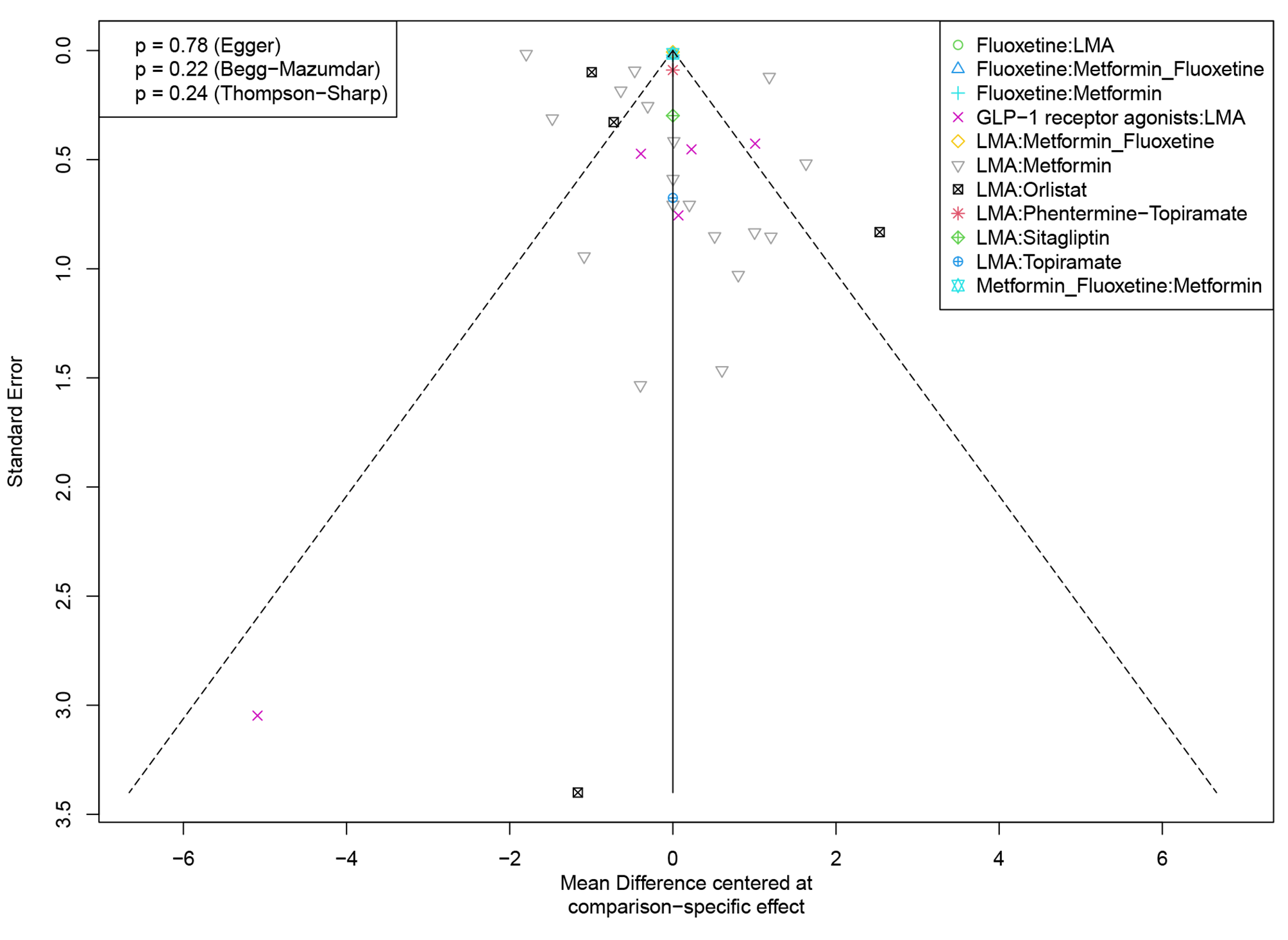

**Outcome: change in weight from baseline**

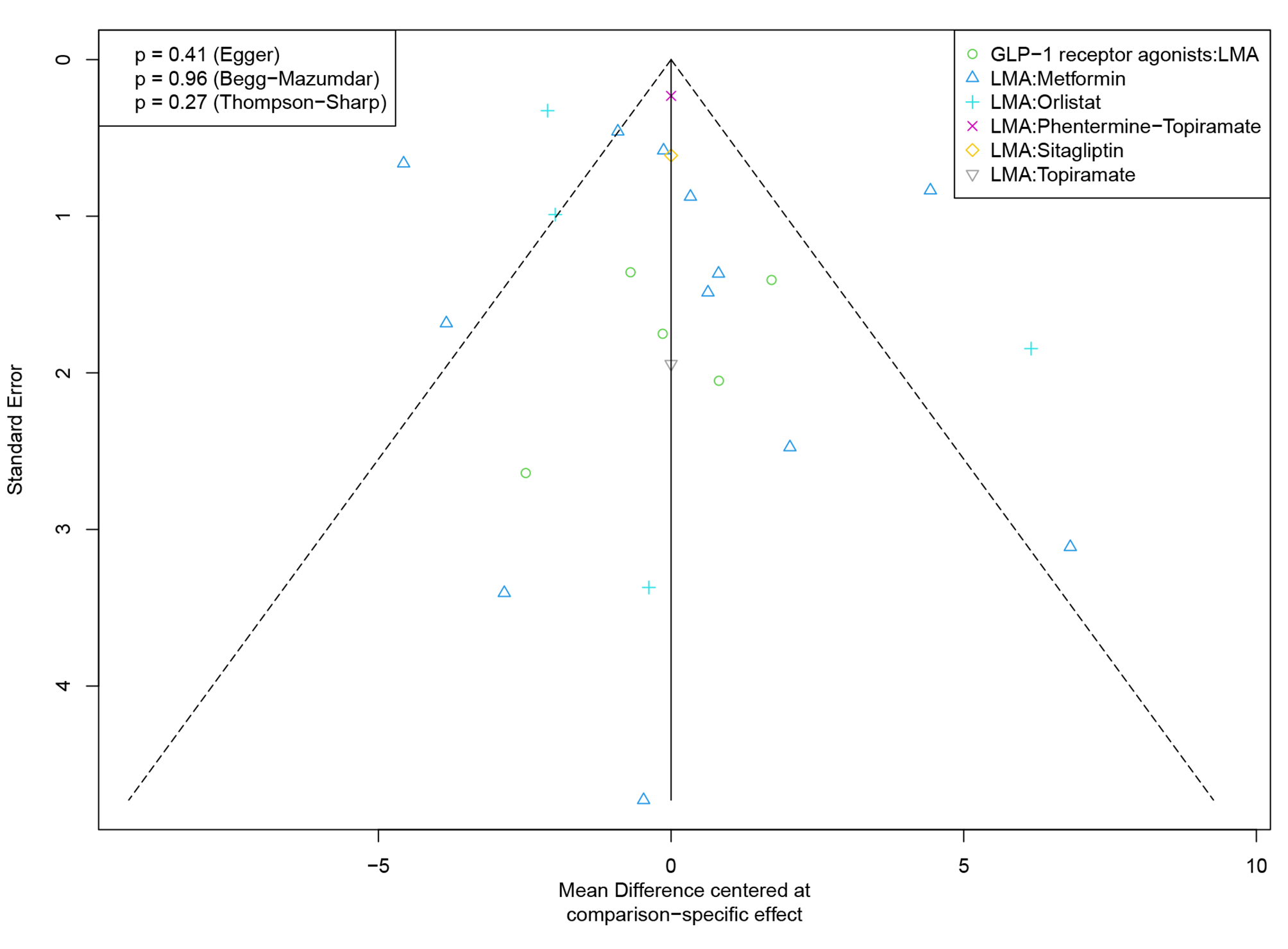

**Outcome:** **percentage of participants achieving BMI reduction of at least 5%**

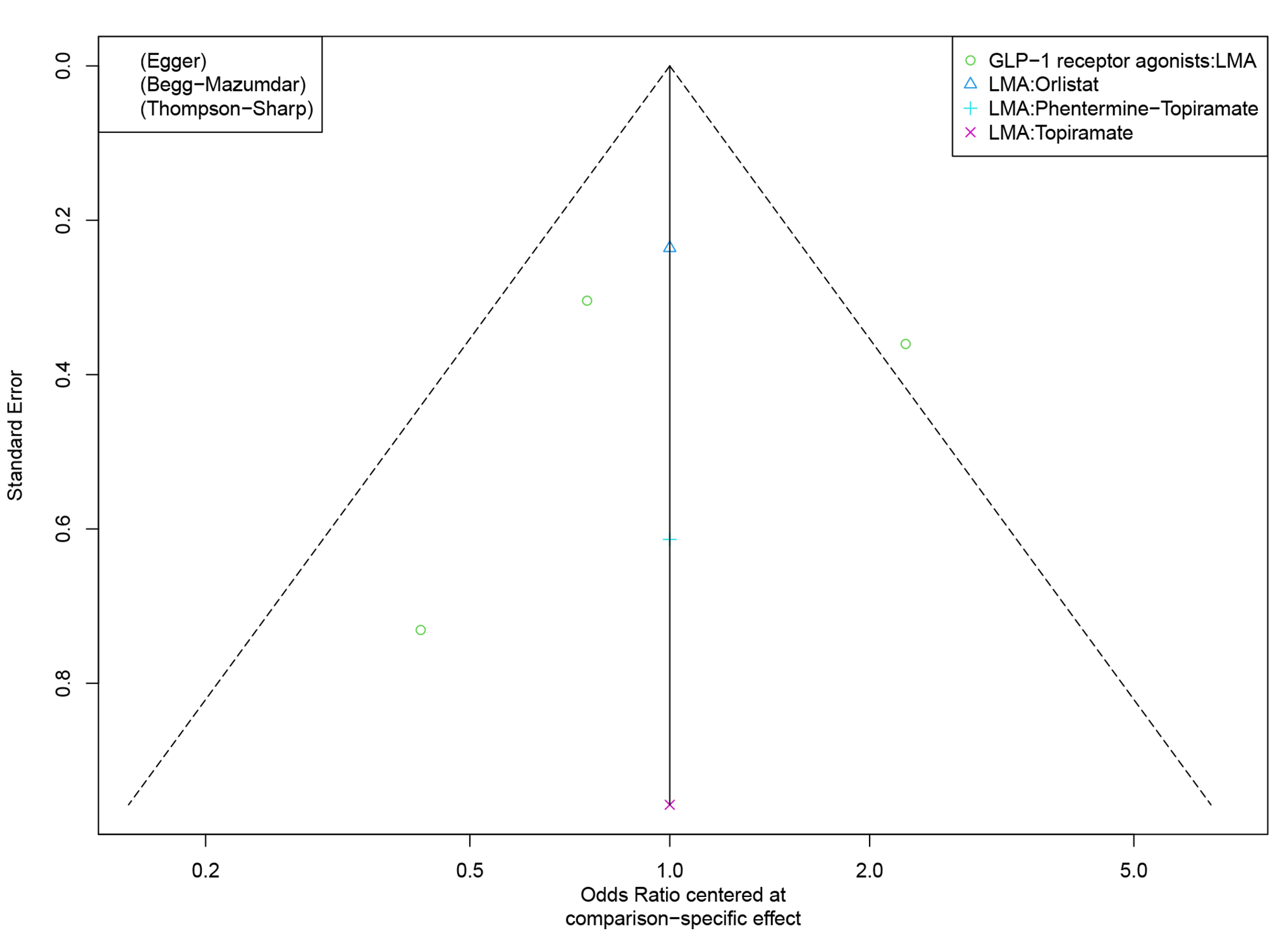

**Outcome: percentage of participants achieving BMI reduction of at least 10%**

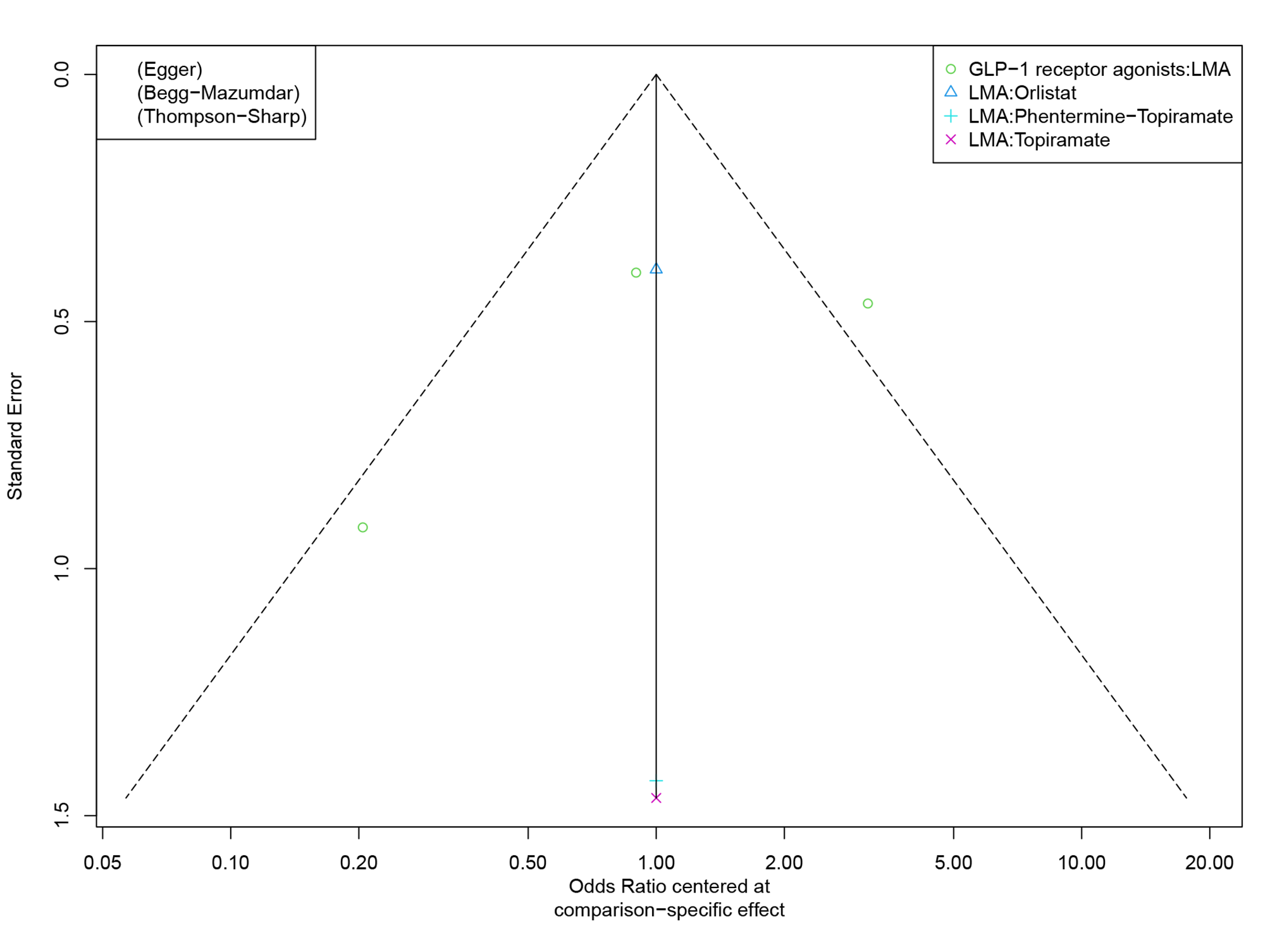

**Outcome: change in BMI z-score from baseline**

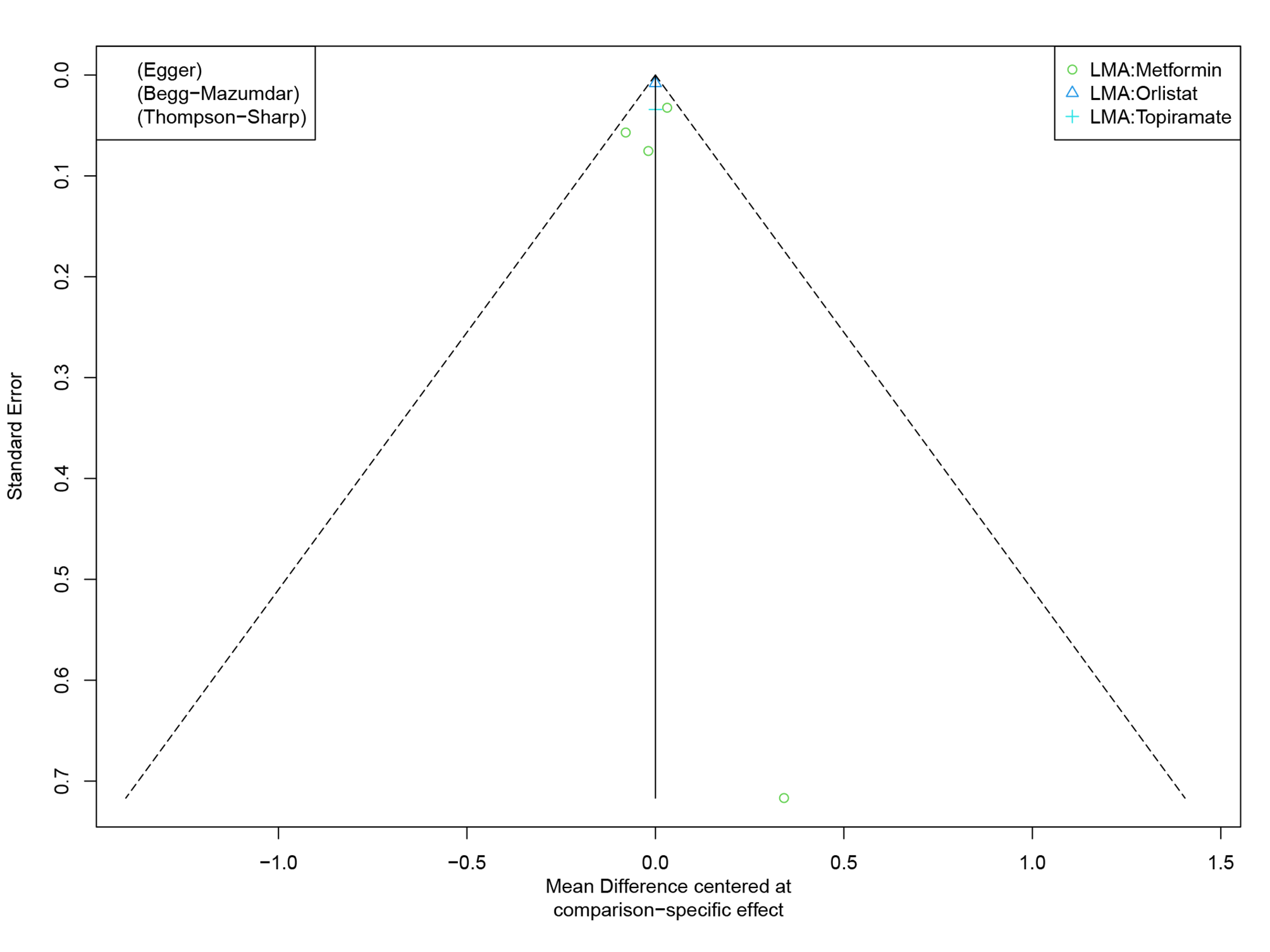

**Outcome: change in BMI SDS from baseline**

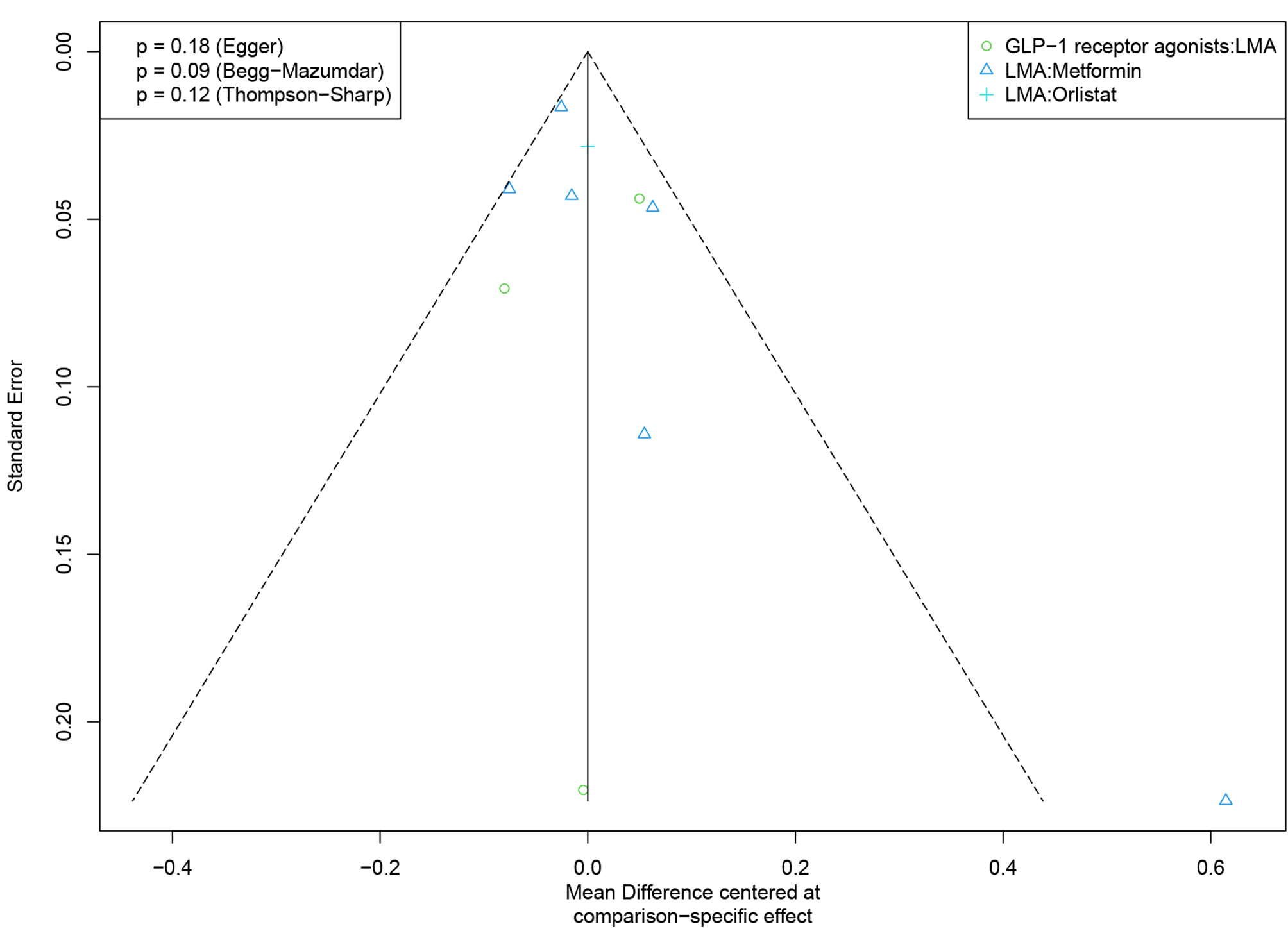

**Outcome: total gastrointestinal adverse events**

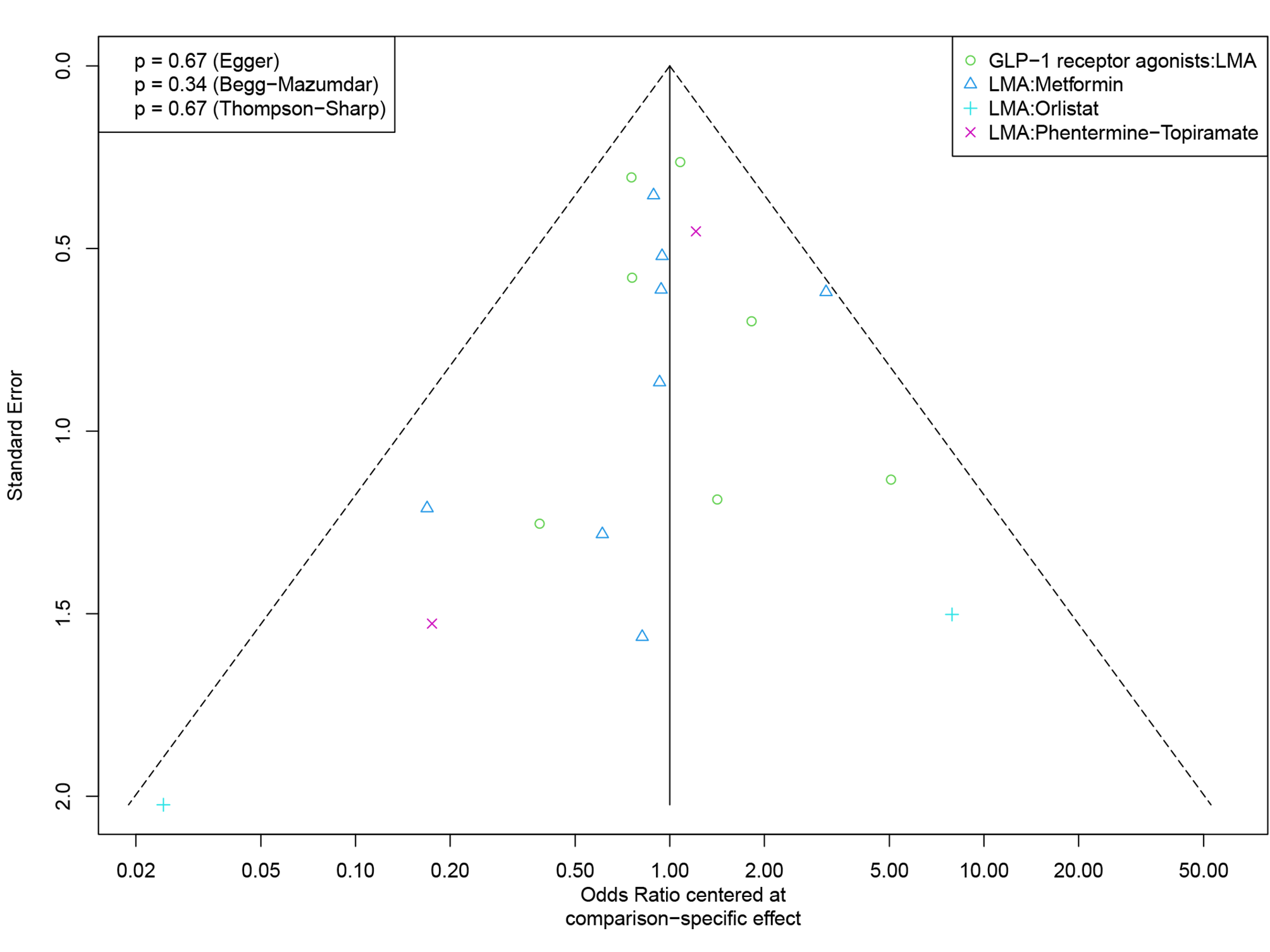

**Outcome: discontinuation due to any adverse event**

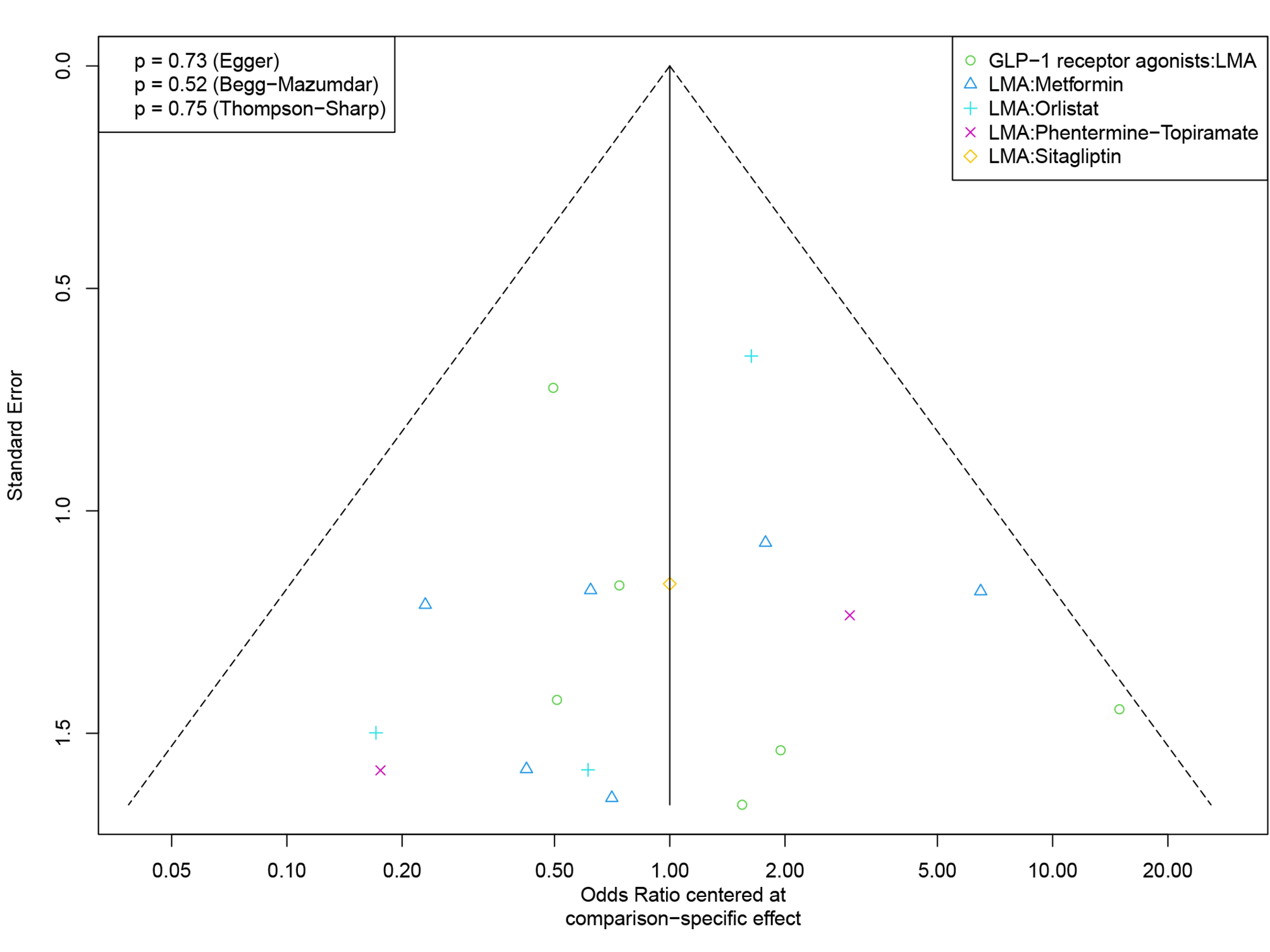

**Outcome: serious adverse events**

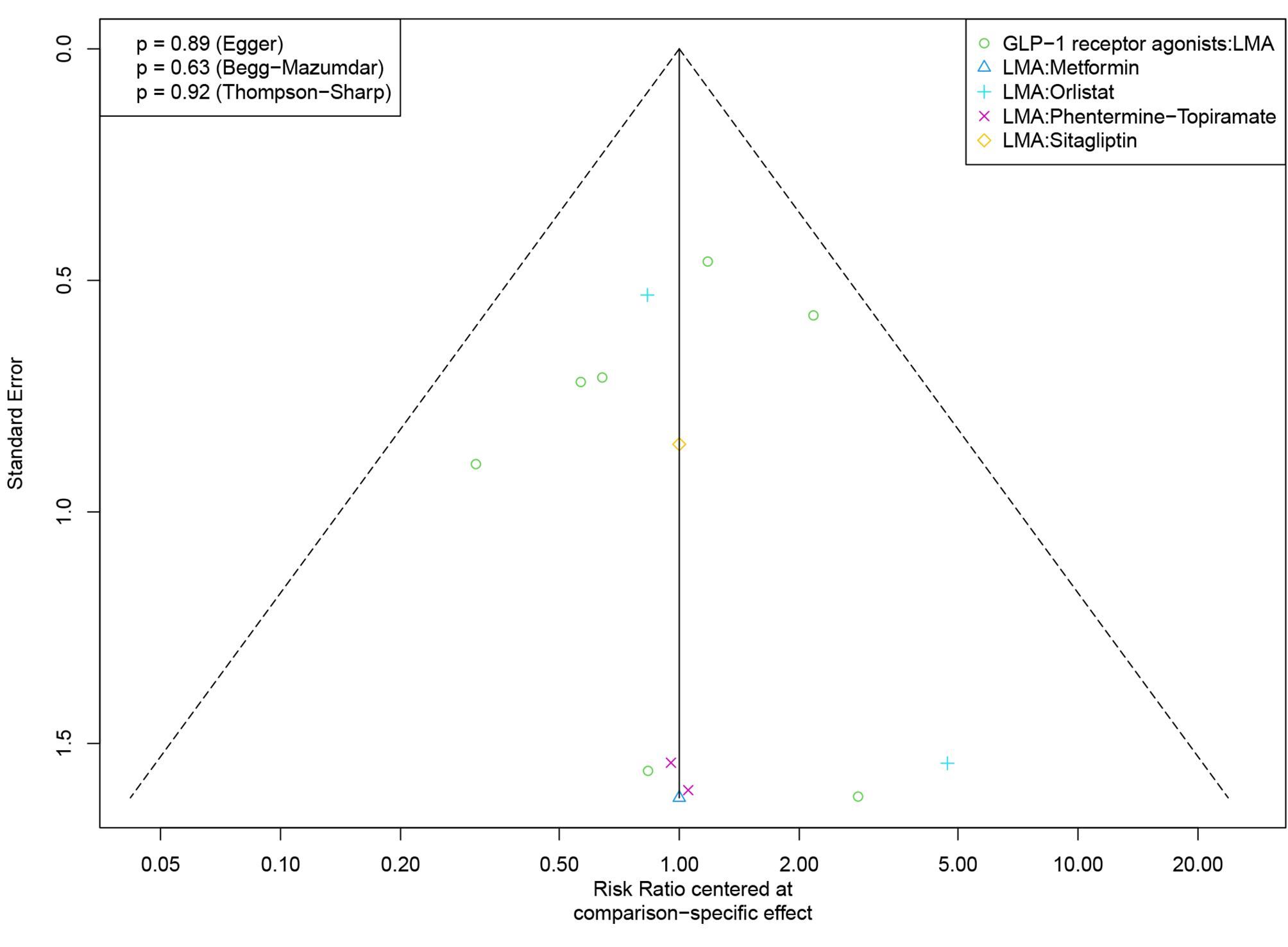

**Outcome: nausea events**

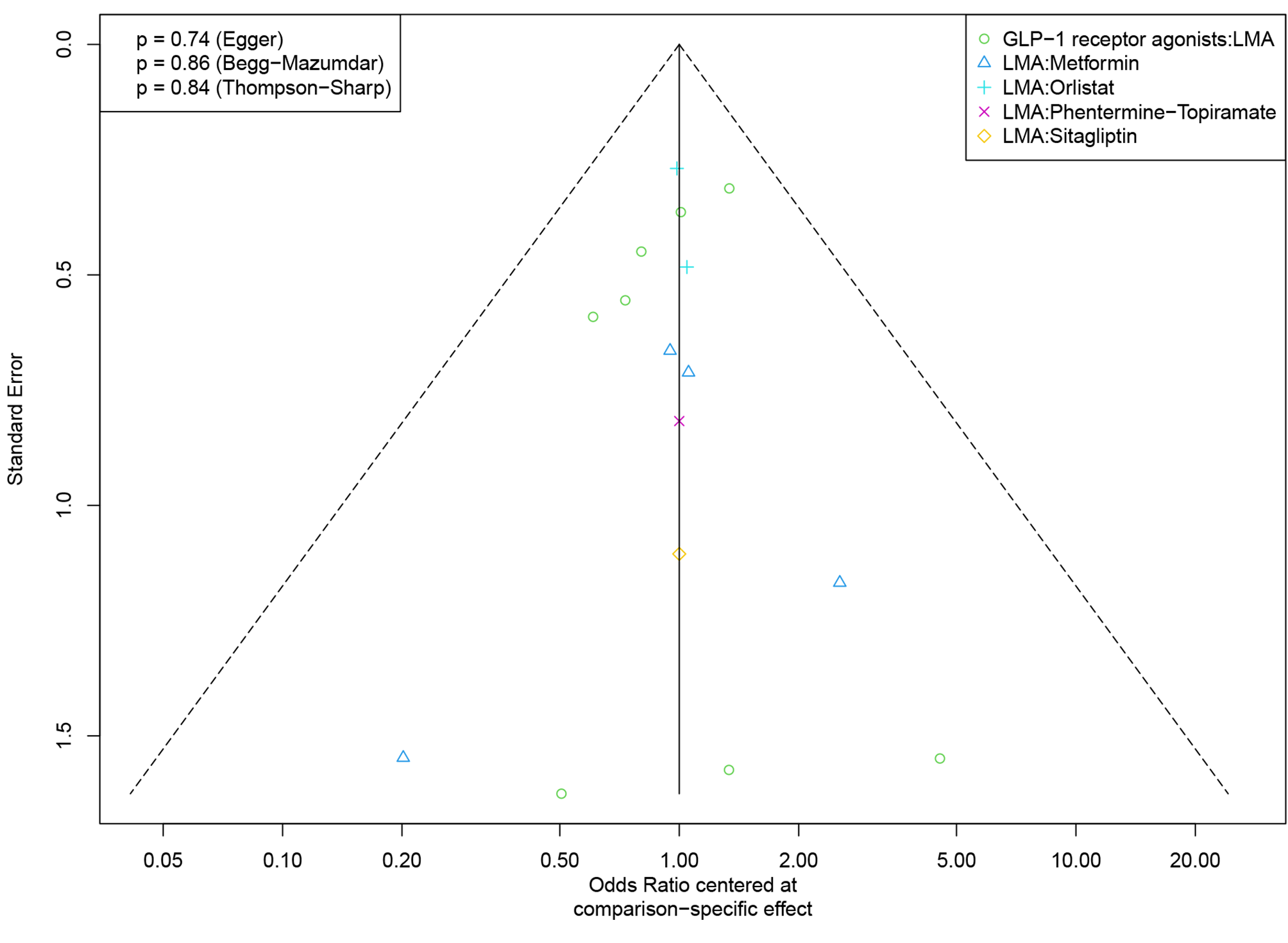

**Outcome: vomiting events**

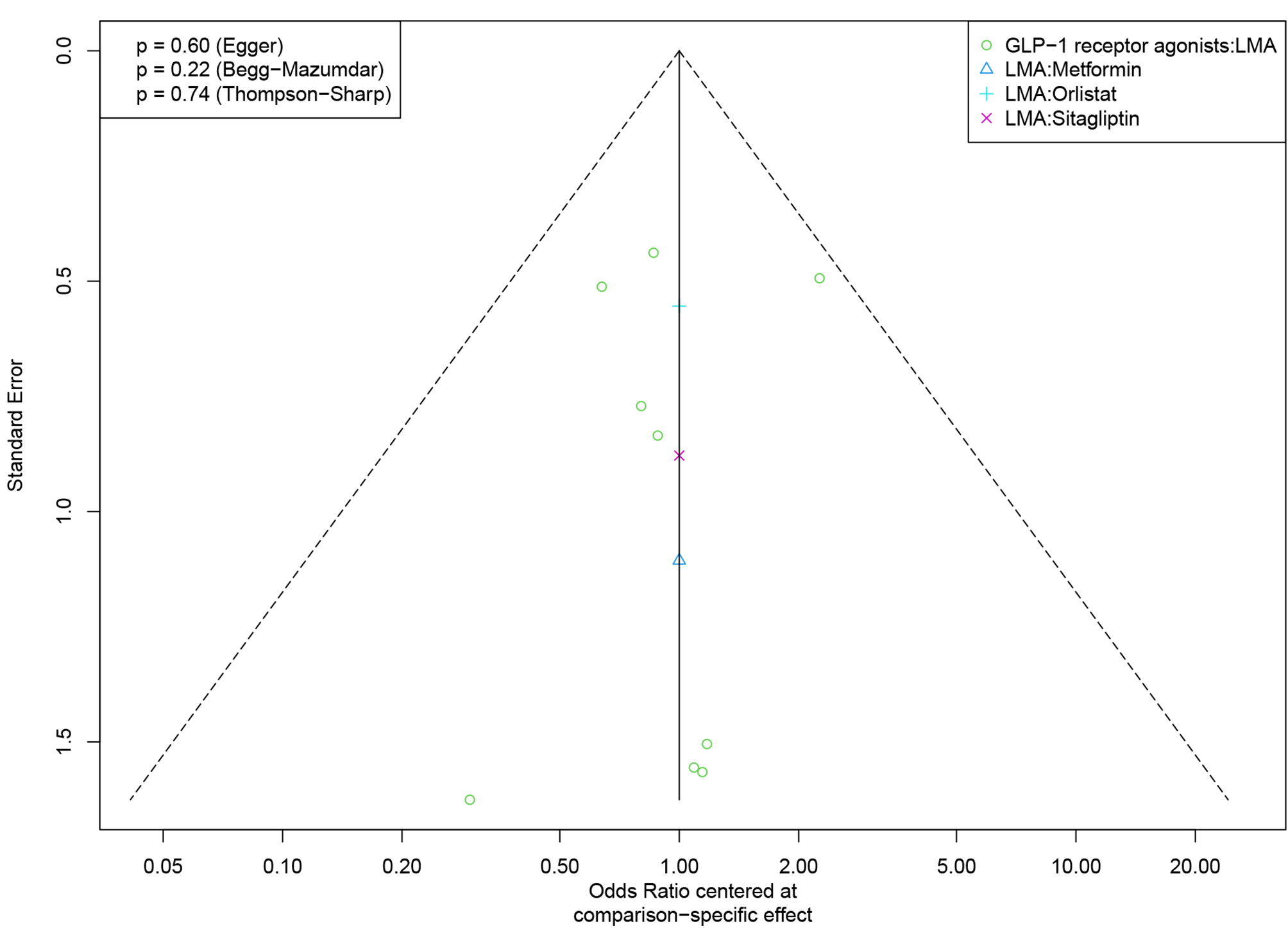

**Outcome: diarrhea events**

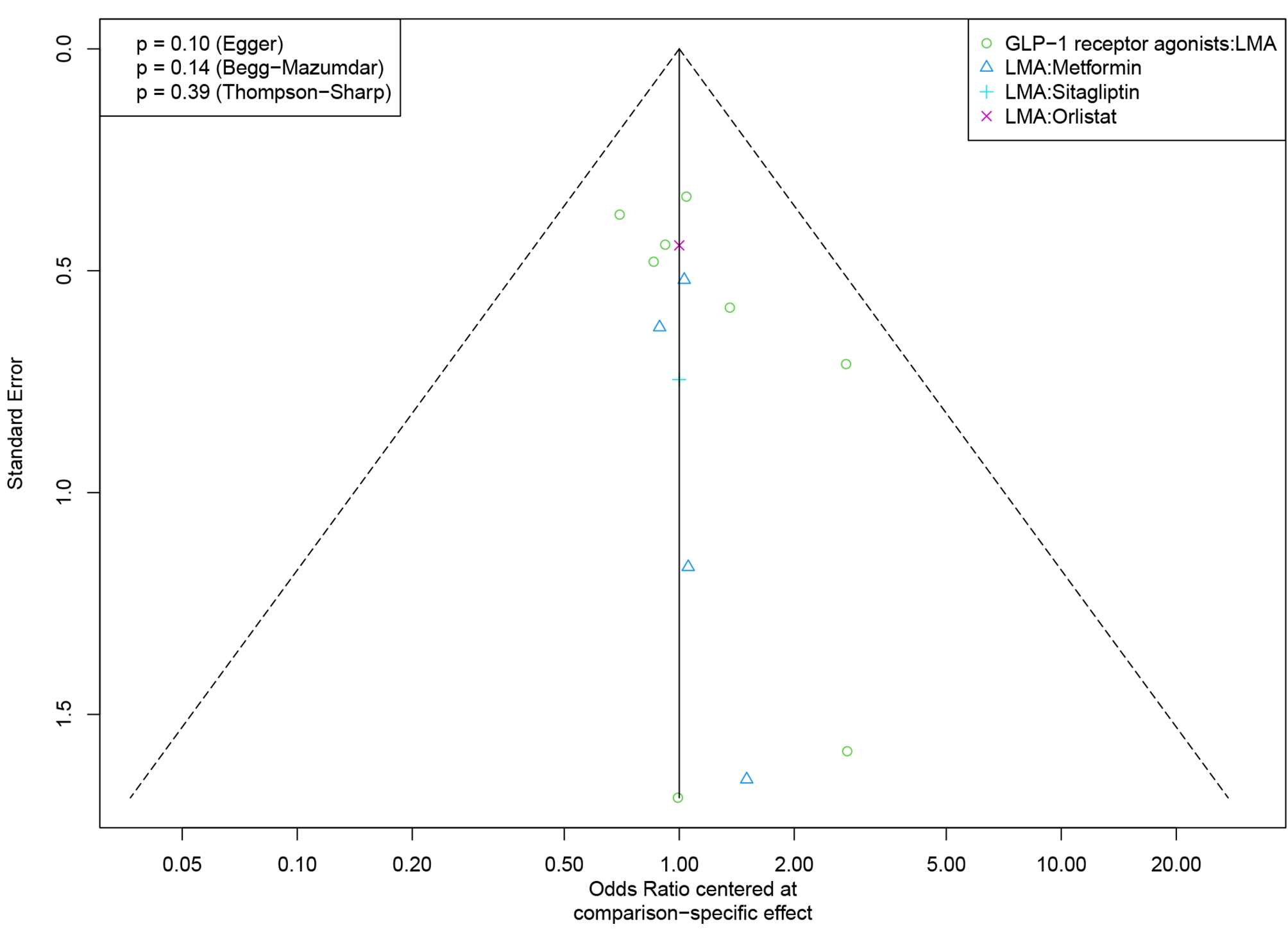

#### 6.9 Estimation of the baseline risk

**Effect of lifestyle modification alone on percentage of participants achieving BMI reduction of at least 5%**
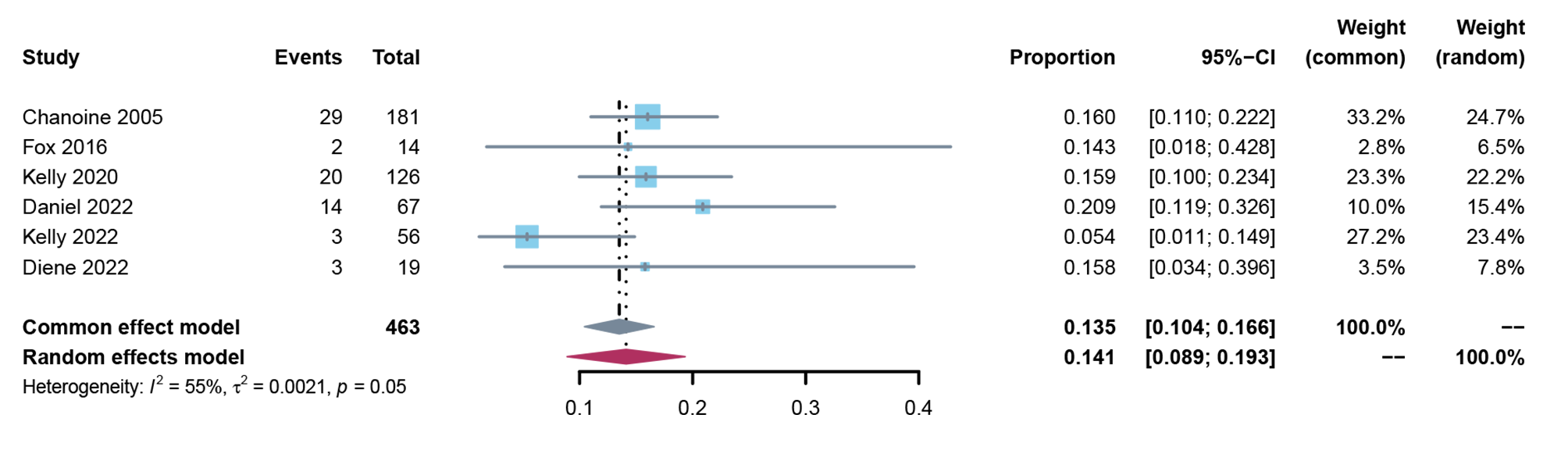

**Effect of lifestyle modification alone on percentage of participants achieving BMI reduction of at least 10%**
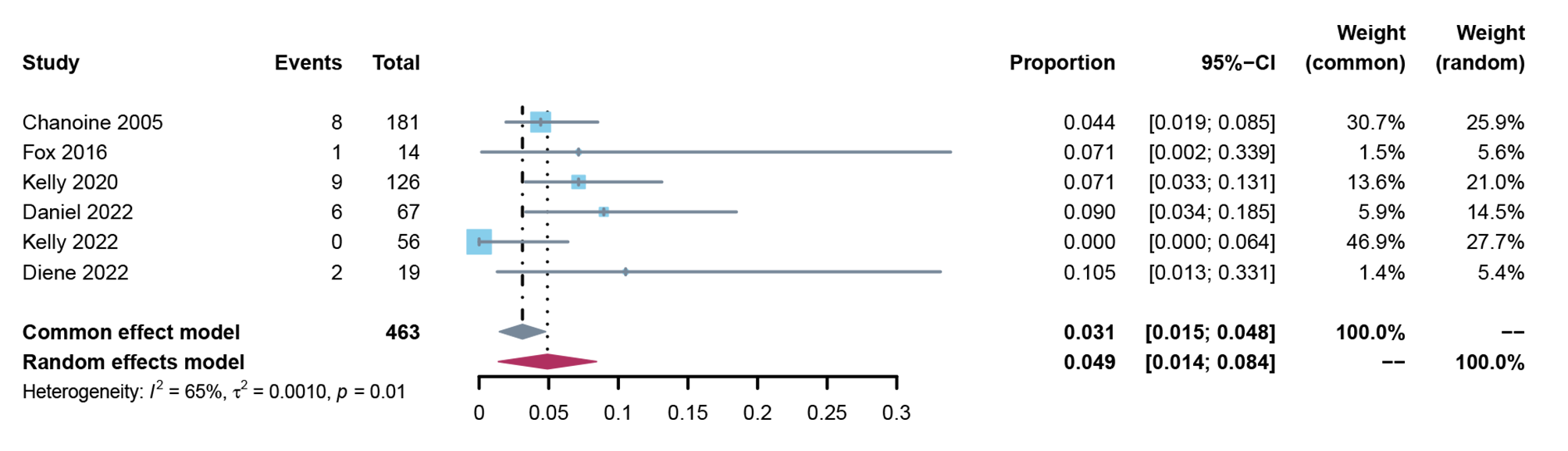

**Effect of lifestyle modification alone on total gastrointestinal adverse events**
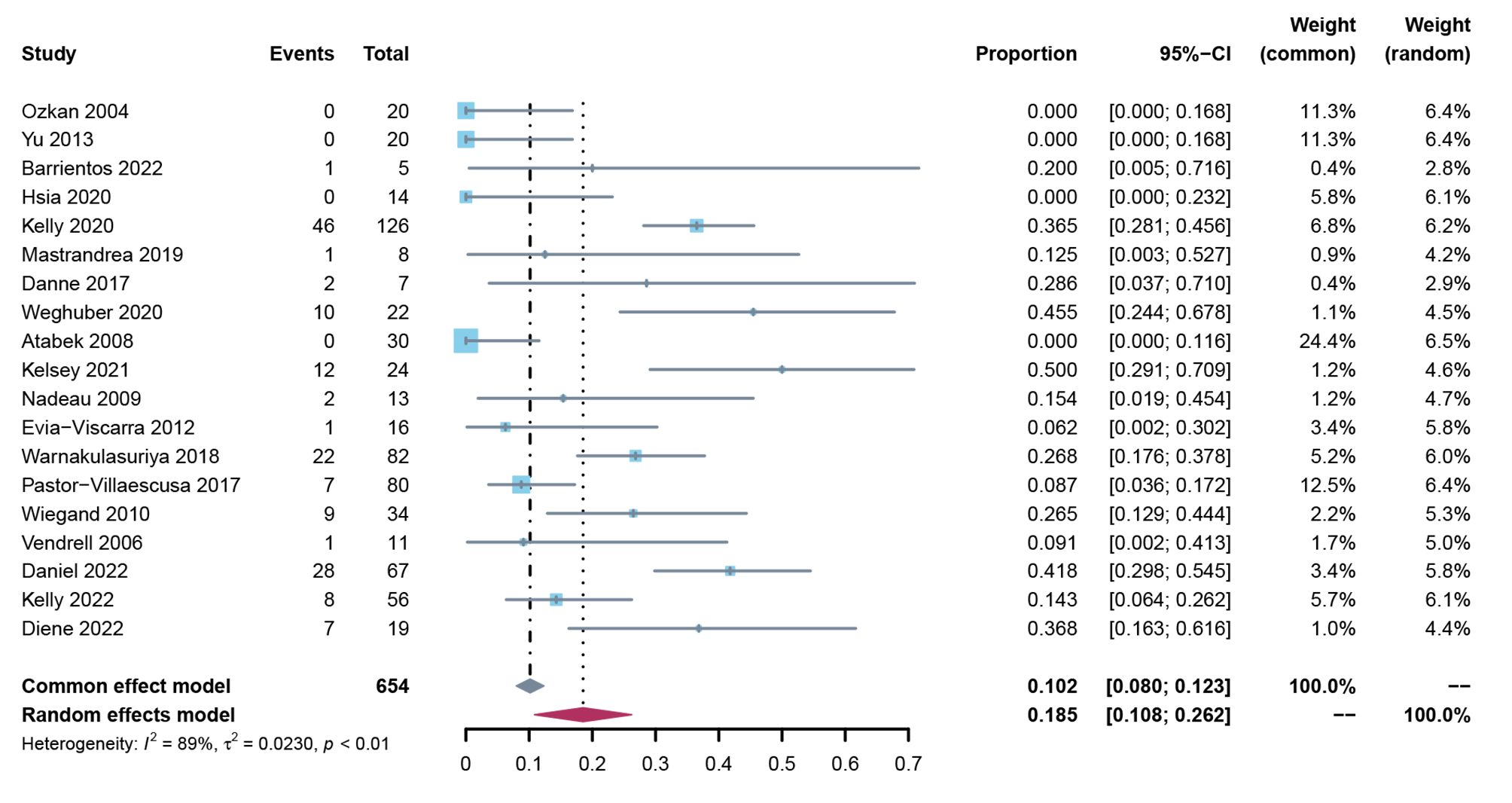

**Effect of lifestyle modification alone on discontinuation due to any adverse event**

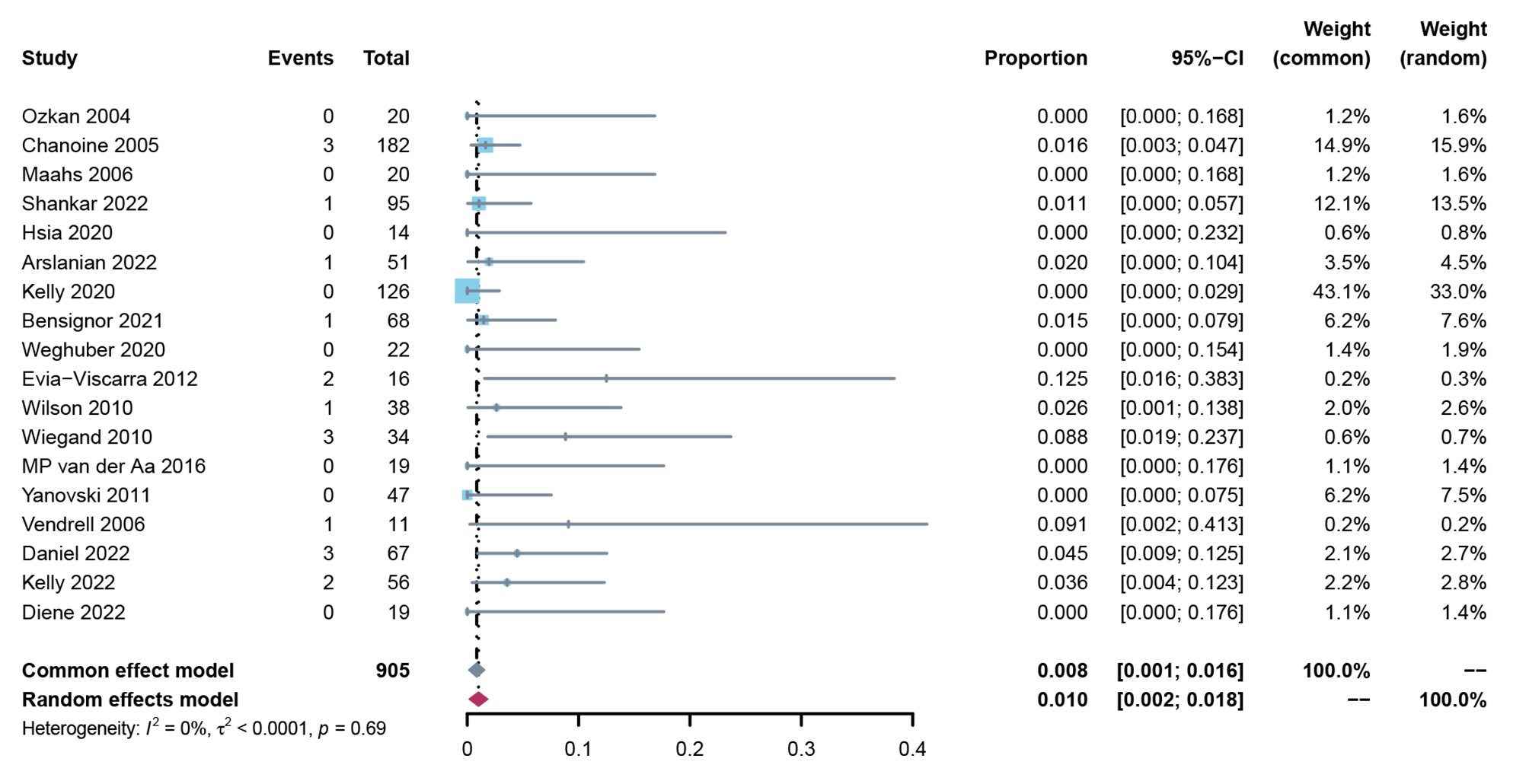

**Effect of lifestyle modification alone on serious adverse events**

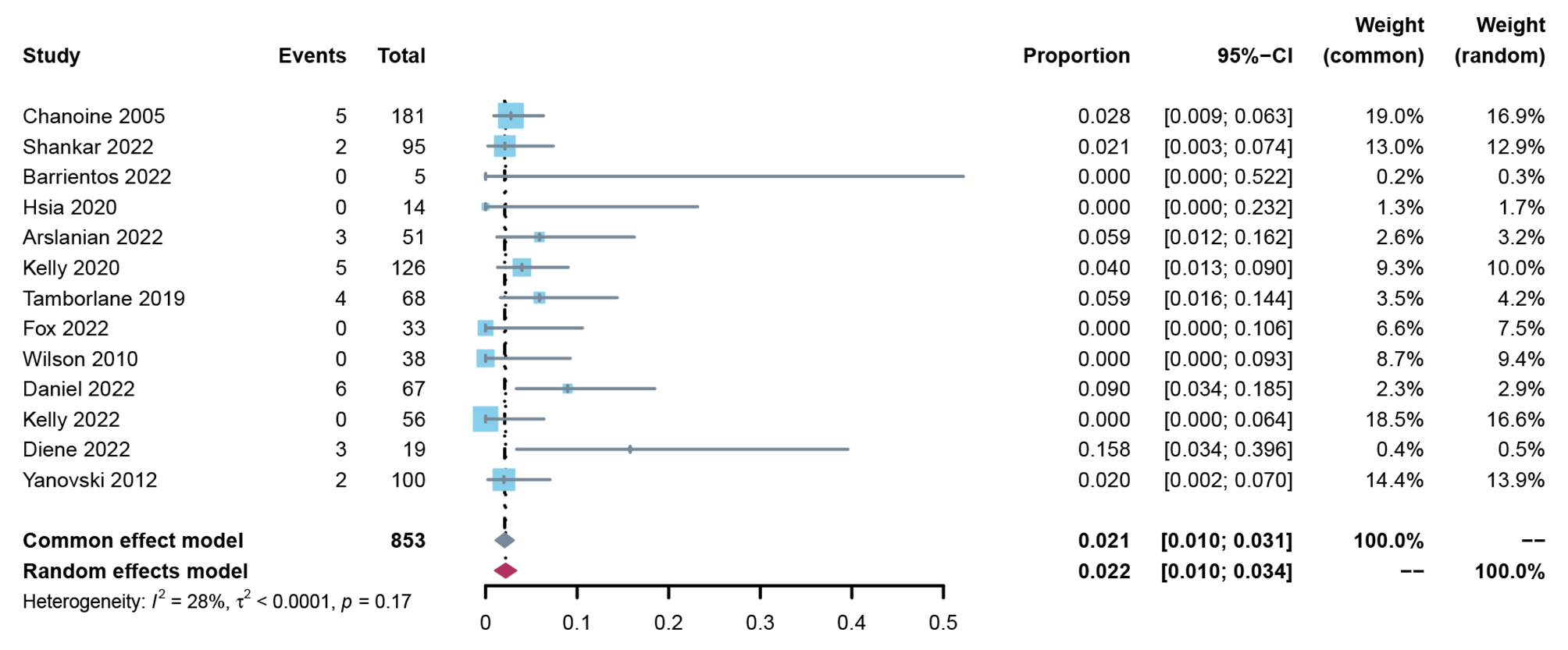

**Effect of lifestyle modification alone on nausea events**

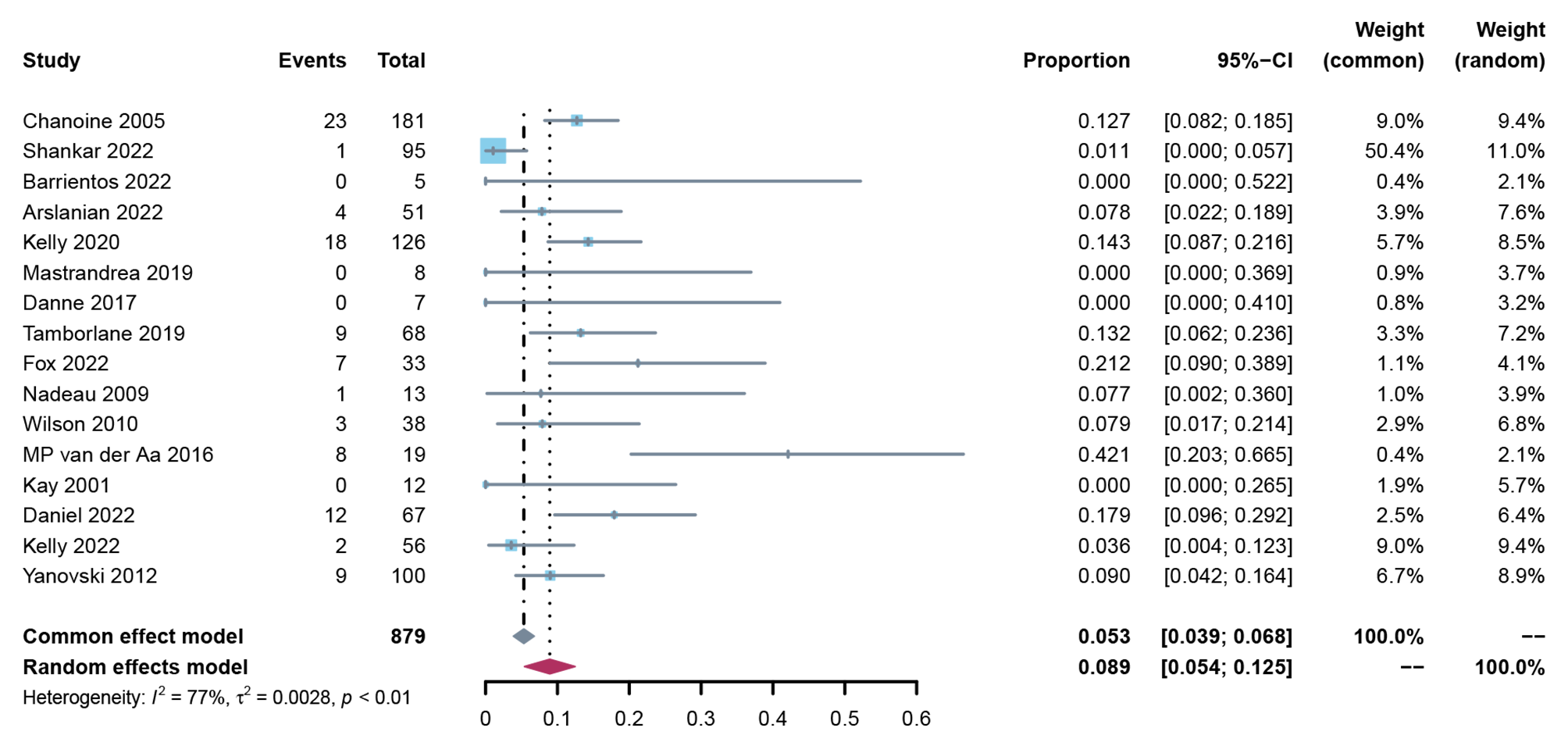

**Effect of lifestyle modification alone on vomiting events**

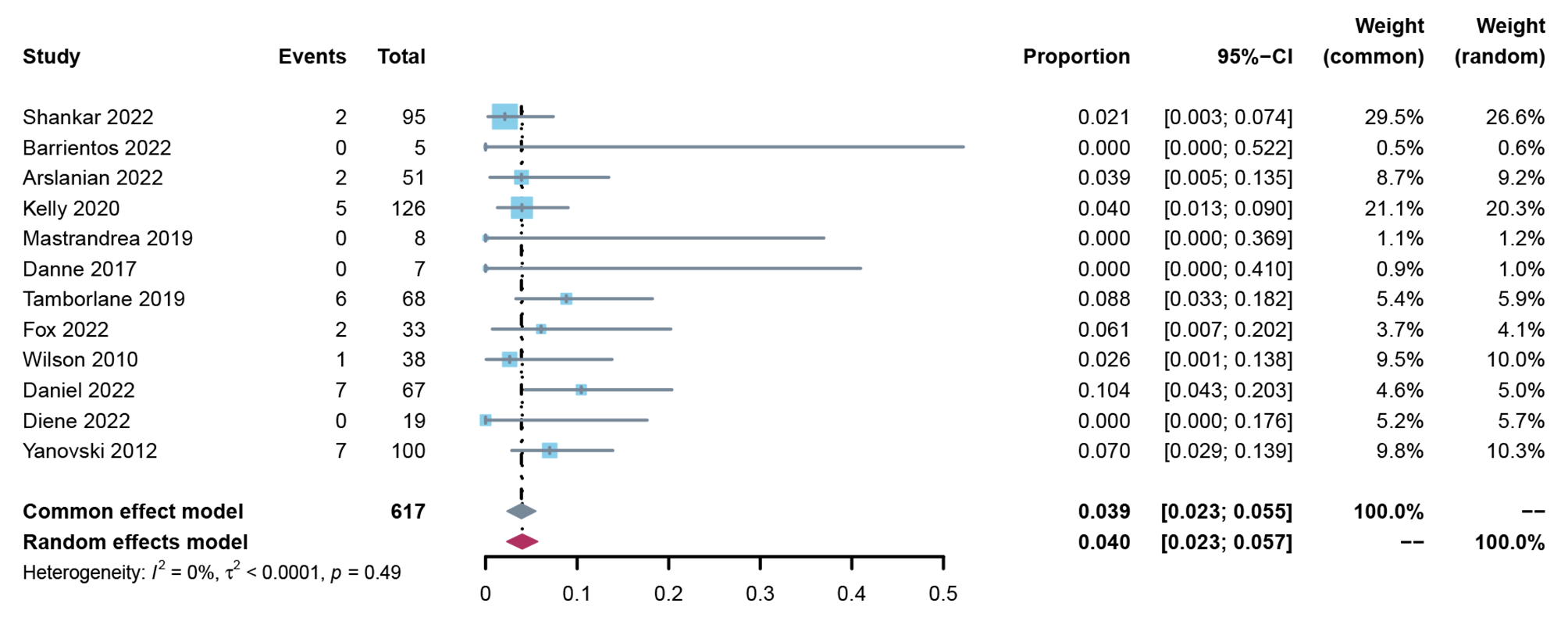

**Effect of lifestyle modification alone on diarrhea events**

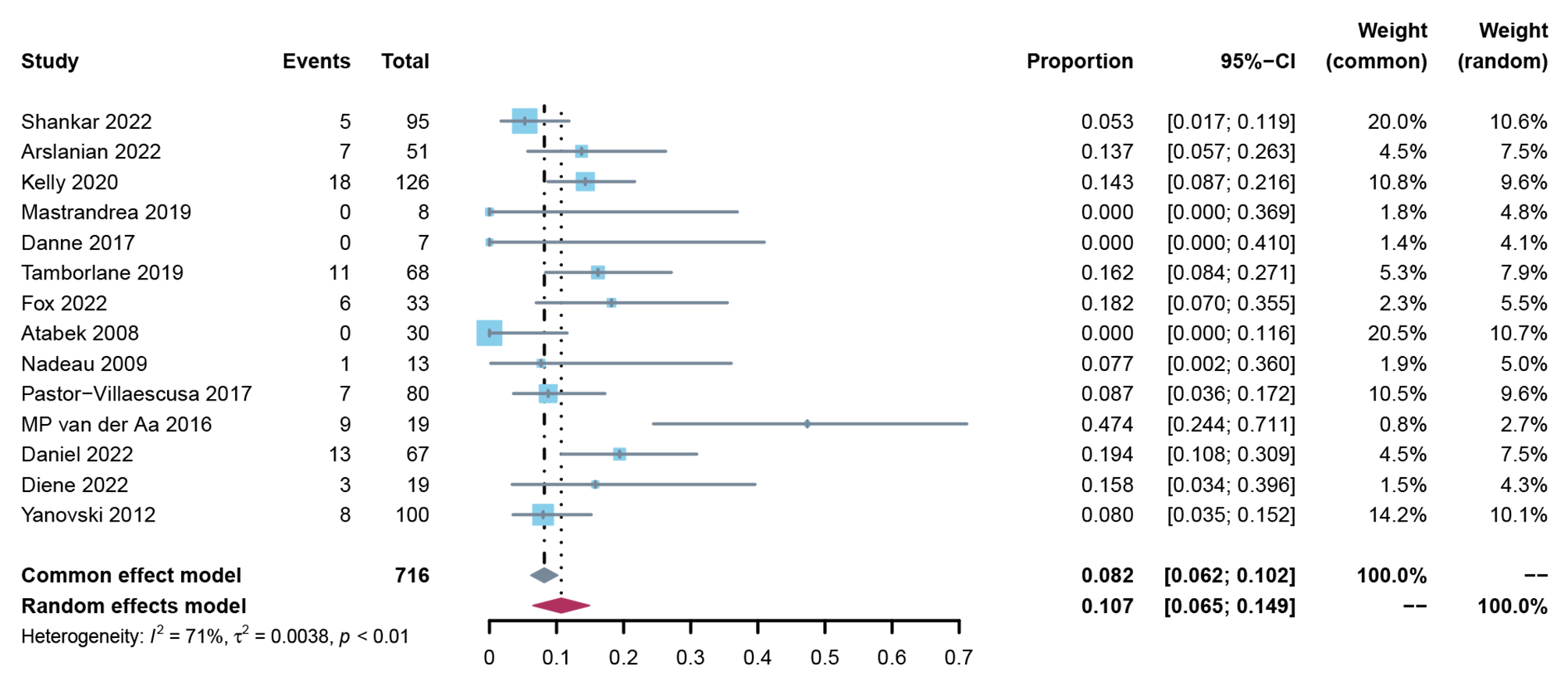

#### 6.10 Minimally contextualized framework

| **Classification** | **Drugs** | **Compared with lifestyle modification alone** | **Pscore** |
| --- | --- | --- | --- |
| **Change in BMI from baseline (mean difference; 95% confidence interval)** | | | |
| **Moderate to high certainty evidence** | | | |
| Among the best | Phentermine-Topiramate | -4.83 (-7.46, -2.20) | 0.9787 |
| Intermediate—possibly better | GLP-1 receptor agonists | -1.81 (-3.19, -0.43) | 0.6286 |
| Intermediate—possibly worse | Metformin | -1.10 (-1.79, -0.41) | 0.4287 |
| Among the worst | Metformin combined with Fluoxetine | -2.00 (-4.30, 0.30) | 0.6482 |
|  | Fluoxetine | -1.70 (-4.00, 0.60) | 0.5786 |
|  | Topiramate | -0.86 (-3.80, 2.08) | 0.3956 |
|  | Sitagliptin | 0.70 (-1.99, 3.39) | 0.1138 |
| **Low to very low certainty evidence** | | | |
| Intermediate—possibly better | Orlistat | -1.66 (-3.23, -0.09) | 0.5877 |
| **Change in weight from baseline (mean difference; 95% confidence interval)** | | | |
| **Moderate to high certainty evidence** | | | |
| Among the best | Phentermine-Topiramate | -14.59 (-19.37, -9.81) | 0.9998 |
| Intermediate—possibly worse | GLP-1 receptor agonists | -3.82 (-6.47, -1.16) | 0.6369 |
|  | Metformin | -2.57 ( -4.23, -0.91) | 0.4731 |
| Among the worst | Topiramate | -2.43 (-8.53, 3.67) | 0.4597 |
|  | Sitagliptin | 1.80 (-3.11, 6.71) | 0.0787 |
| **Low to very low certainty evidence** | | | |
| Intermediate—possibly better | Orlistat | -4.28 ( -7.12, -1.44) | 0.6874 |
| **Percentage of participants achieving BMI reduction of at least 5% (the absolute risk difference)** | | | |
| **Moderate to high certainty evidence** | | | |
| Among the best | Phentermine-Topiramate | 557 more (137 more to 729 more) | 0.9276 |
| Intermediate—possibly better | GLP-1 receptor agonists | 303 more (100 more to 527 more) | 0.6939 |
| Among the worst | Topiramate | 106 more (109 fewer to 625 more) | 0.3940 |
|  | Orlistat | 95 more (71 fewer to 417 more) | 0.3673 |
|  | Lifestyle modification alone | 141 per 1000 patients | 0.1172 |
| **Total gastrointestinal adverse events ((the absolute risk difference))** | | | |
| **Moderate to high certainty evidence** | | | |
| Among the least harmful | Lifestyle modification alone | 185 per 1000 patients | 0.8392 |
|  | Phentermine-Topiramate | 11 fewer (109 fewer to 170 more) | 0.8285 |
|  | Metformin | 57 more (22 fewer to 158 more) | 0.5742 |
| Among the most harmful | GLP-1 receptor agonists | 218 more (120 more to 323 more) | 0.2534 |
| **Low to very low certainty evidence** | | | |
| Among the most harmful | Orlistat | 726 more (299 more to 806 more) | 0.0048 |
| **Discontinuation due to adverse events ((the absolute risk difference)** | | | |
| **Moderate to high certainty evidence** | | | |
| Among the least harmful | Phentermine-Topiramate | 5 fewer (9 fewer to 23 more) | 0.8788 |
|  | Lifestyle modification alone | 10 per 1000 patients | 0.7689 |
|  | Metformin | 9 more (3 fewer to 42 more) | 0.4308 |
|  | GLP-1 receptor agonists | 10 more (2 fewer to 42 more) | 0.4023 |
|  | Sitagliptin | 20 more (7 fewer to 237 more) | 0.3121 |
| More harmful than lifestyle modification alone, but no worse than other interventions | Orlistat | 23 more (1 more to 91 more) | 0.2071 |

### Appendix 7: Contribution matrices

**Note:** LMA, Lifestyle modification alone; GLP-1, glucagon-like peptide-1; Metformin_Fluoxetine, Metformin combined with Fluoxetine.

**Outcome: change in BMI from baseline**

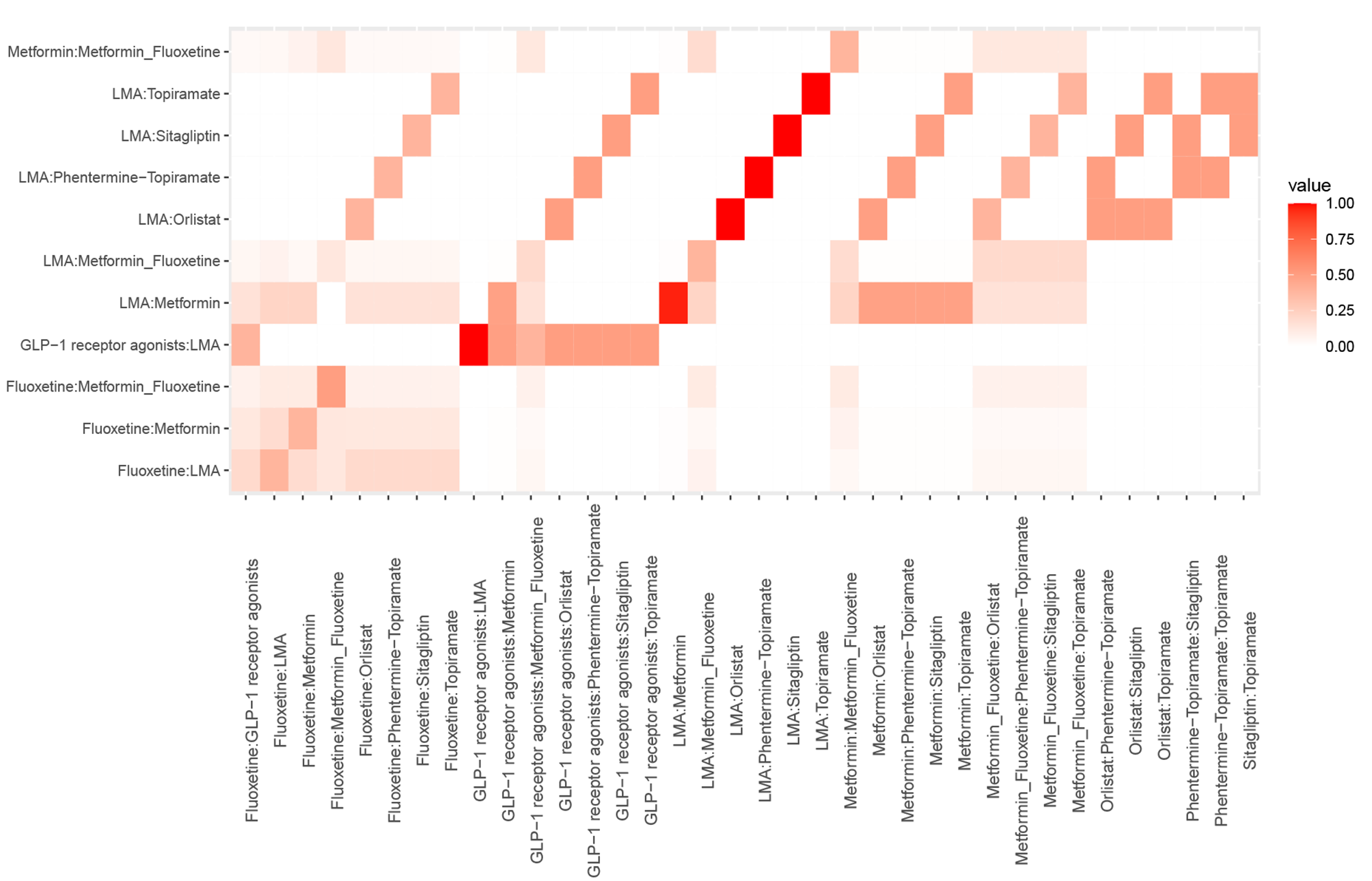

**Outcome: change in weight from baseline**

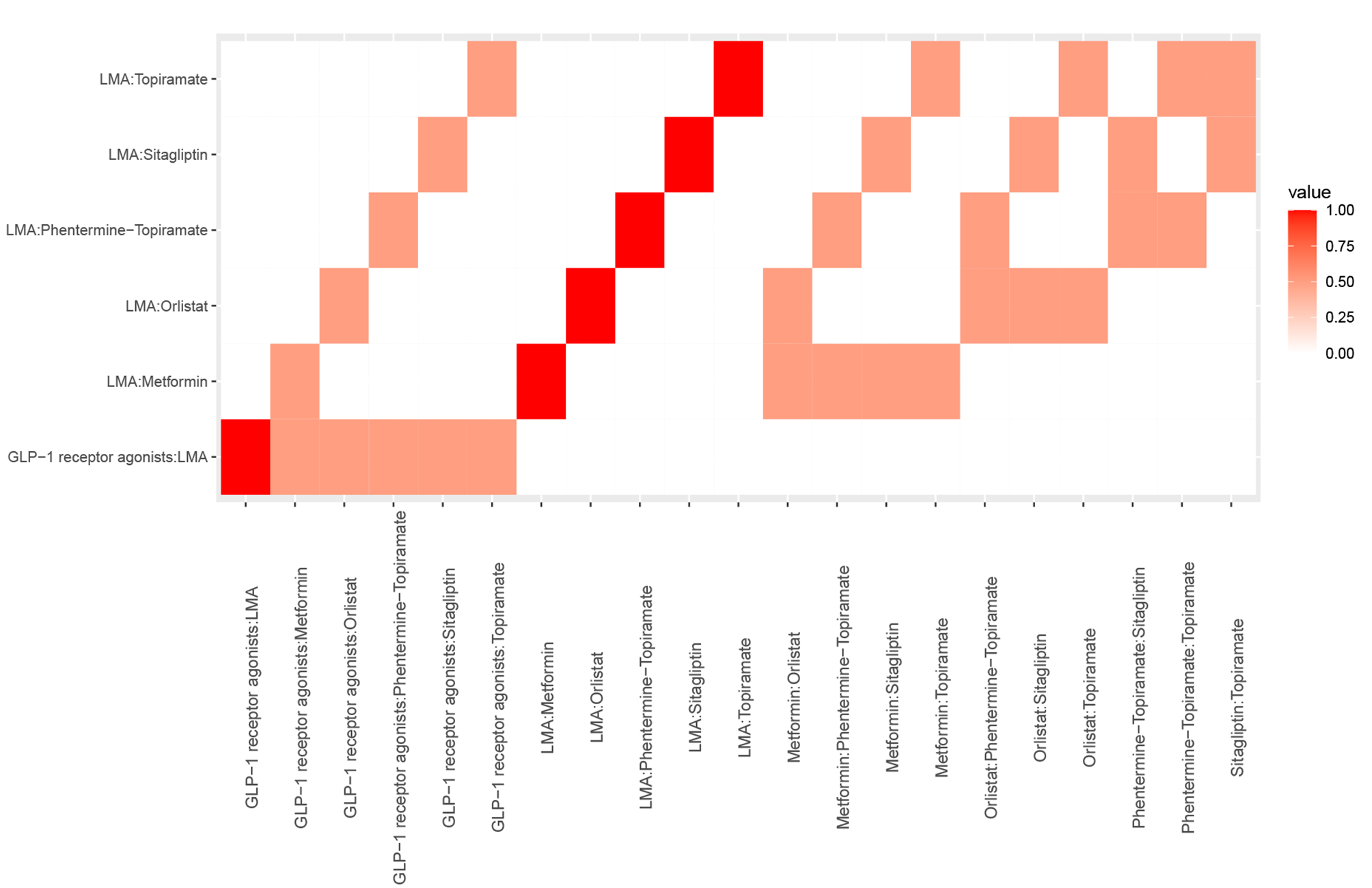

**Outcome: percentage of participants achieving BMI reduction of at least 5%**

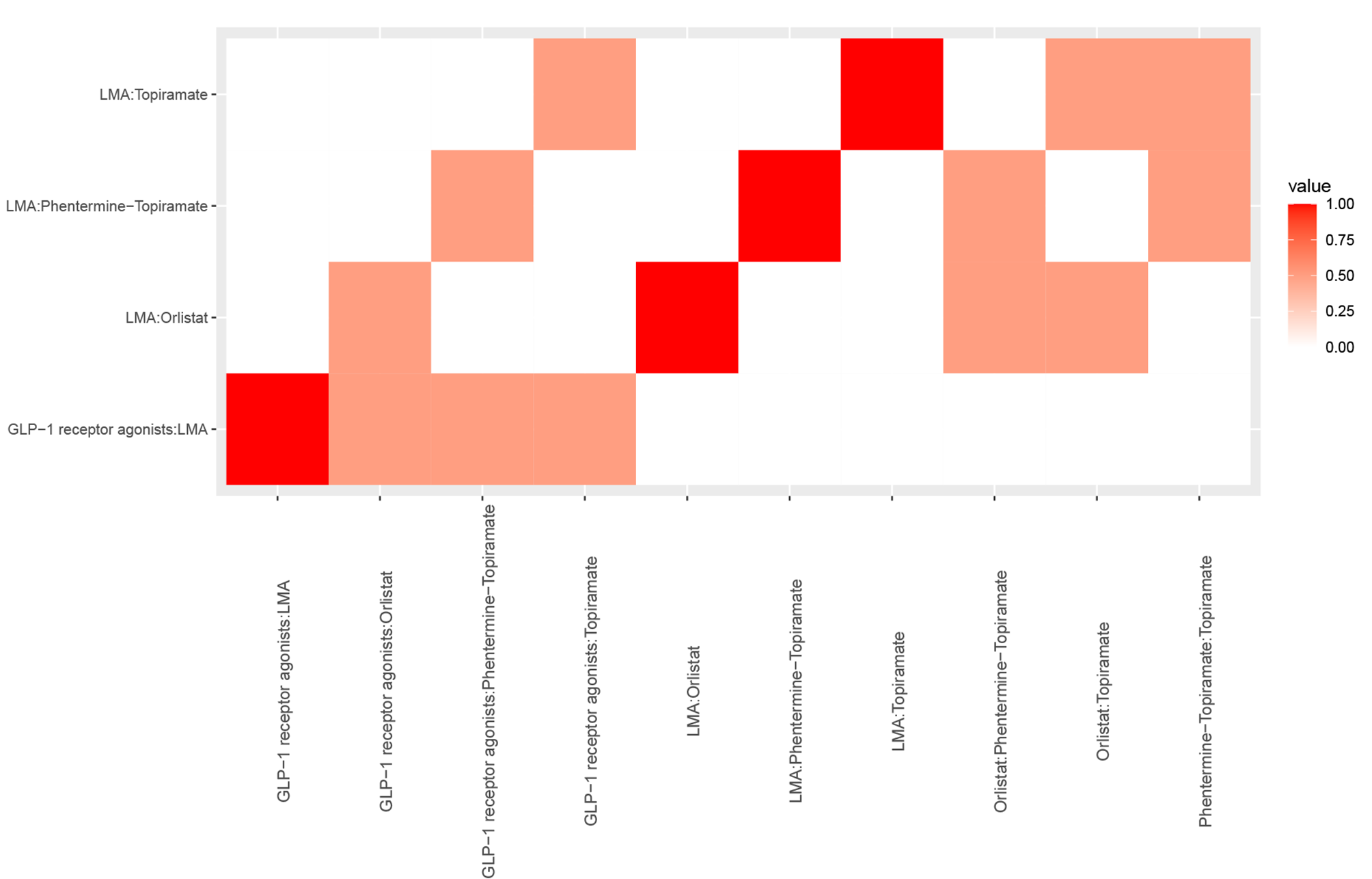

**Outcome: percentage of participants achieving BMI reduction of at least 10%**

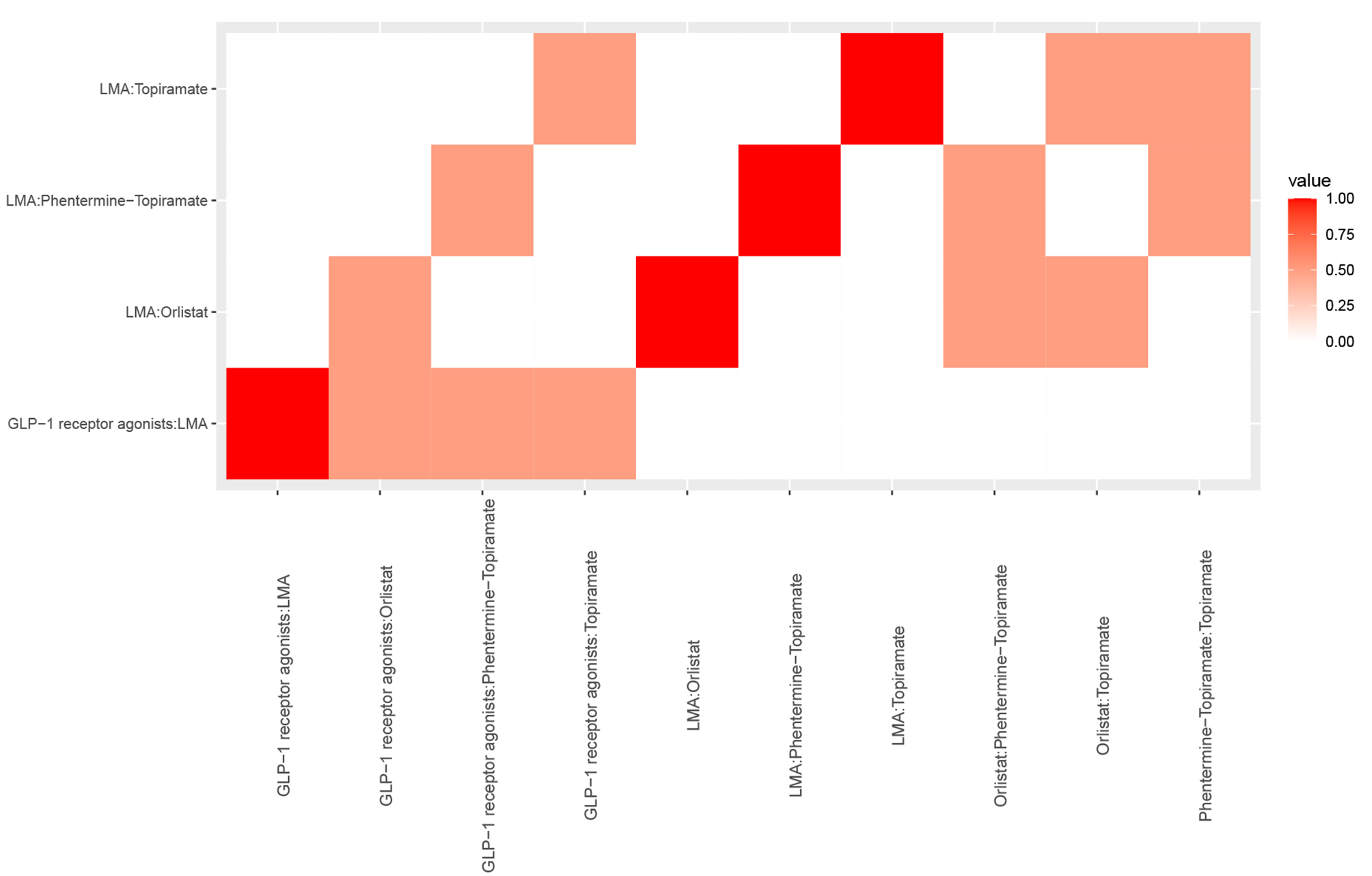

**Outcome: change in BMI z-score from baseline**

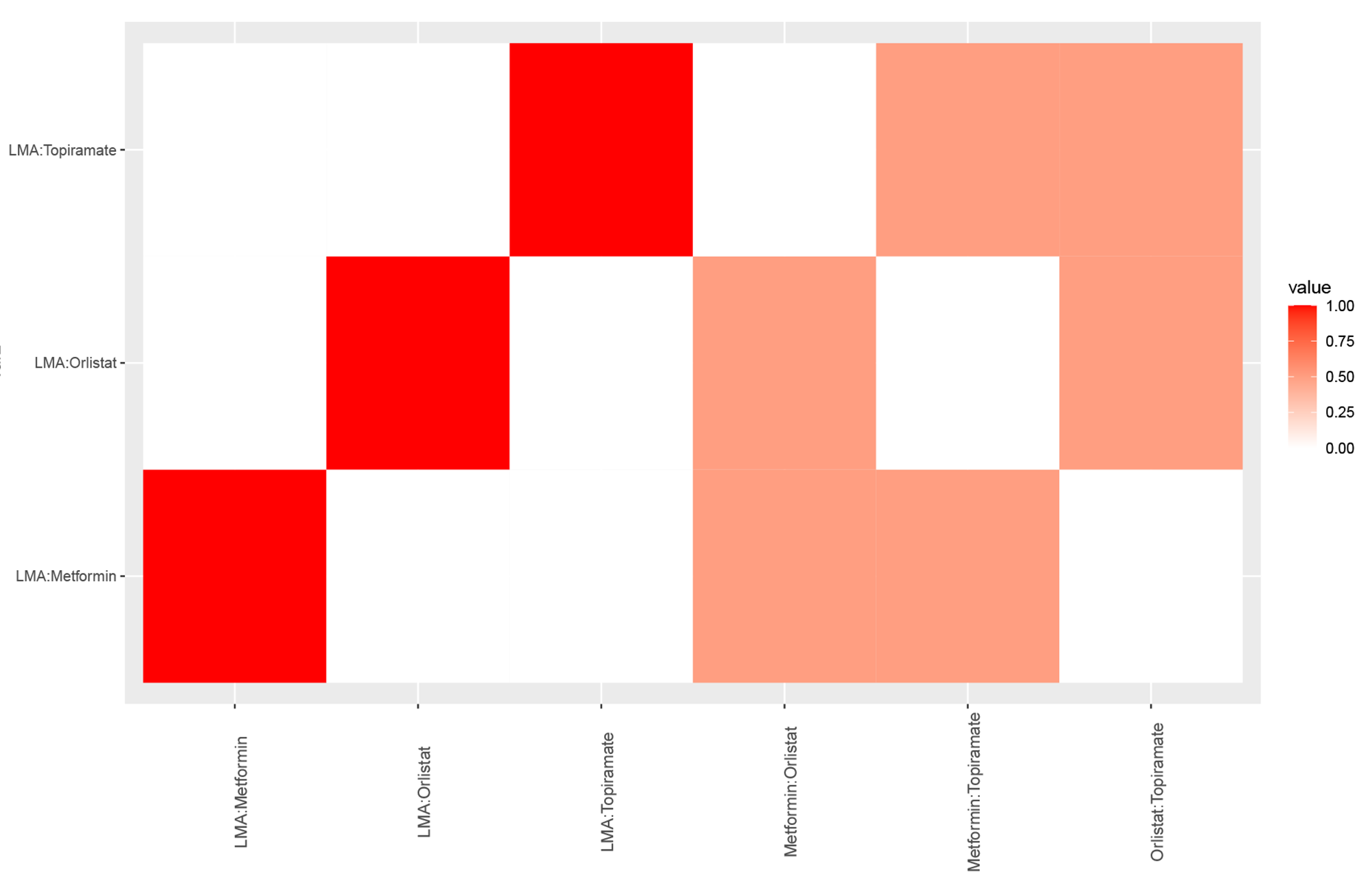

**Outcome: change in BMI SDS from baseline**

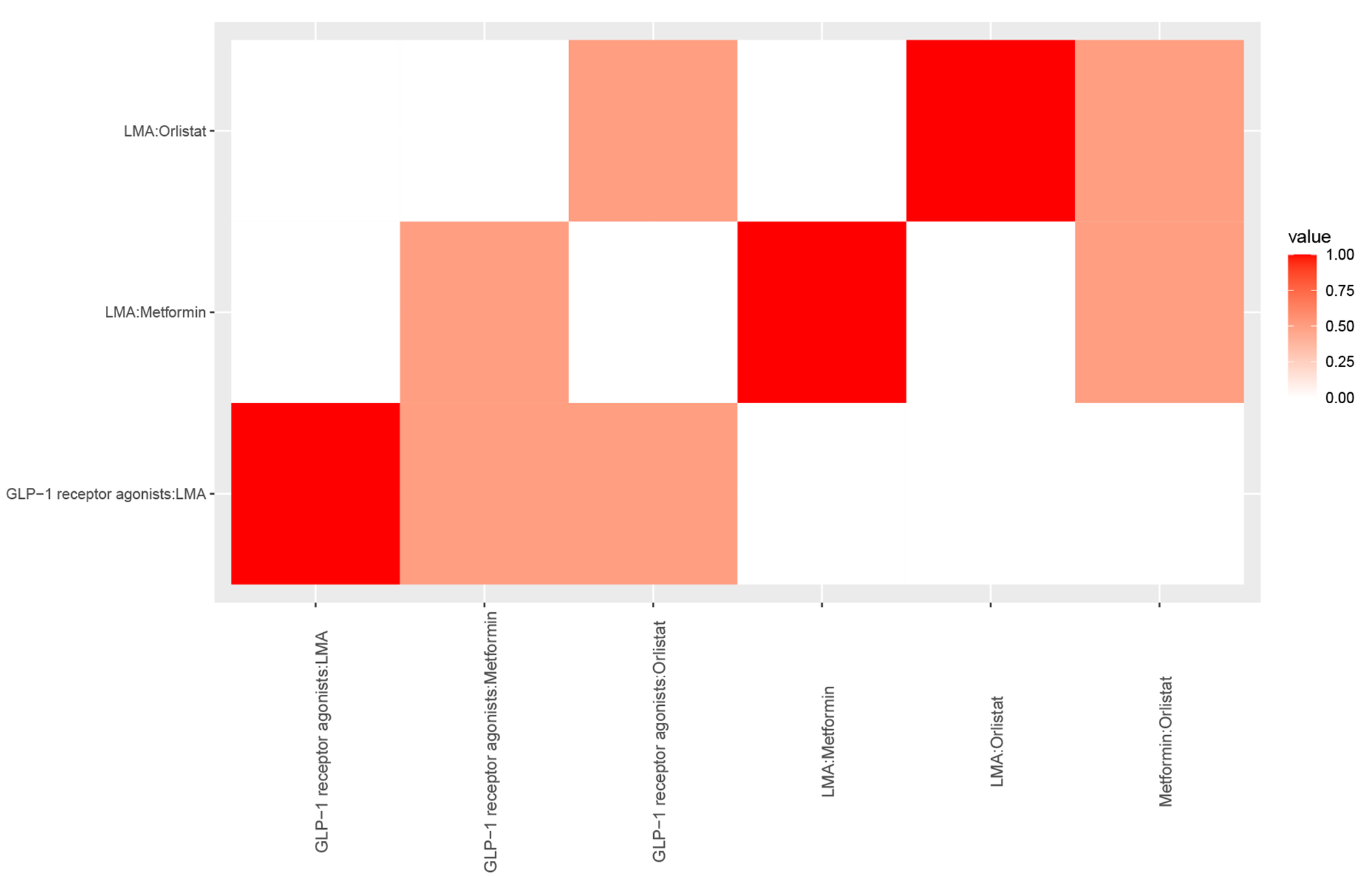

**Outcome: total gastrointestinal adverse events**

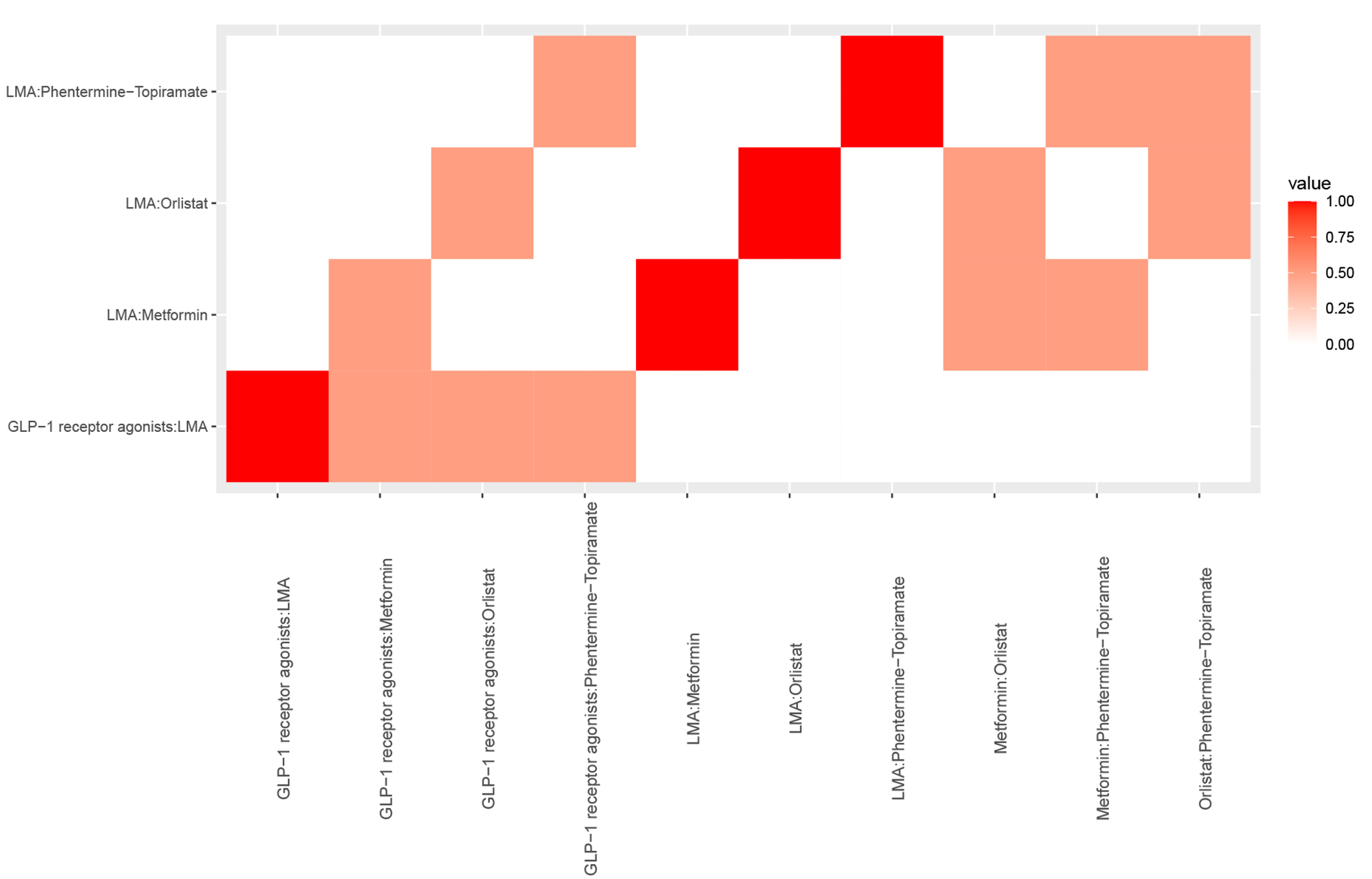

**Outcome: discontinuation due to any adverse event**

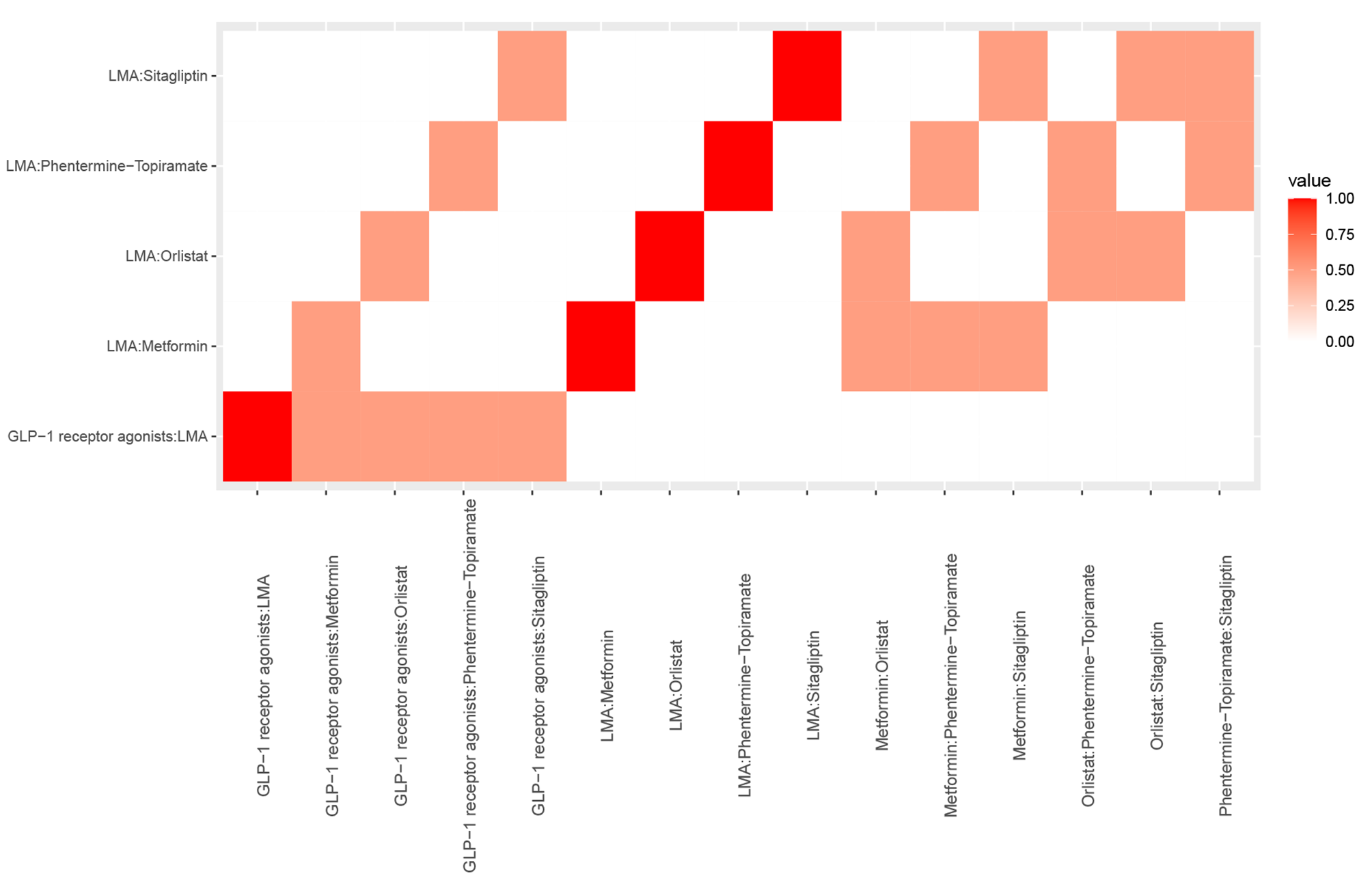

**Outcome: serious adverse events**

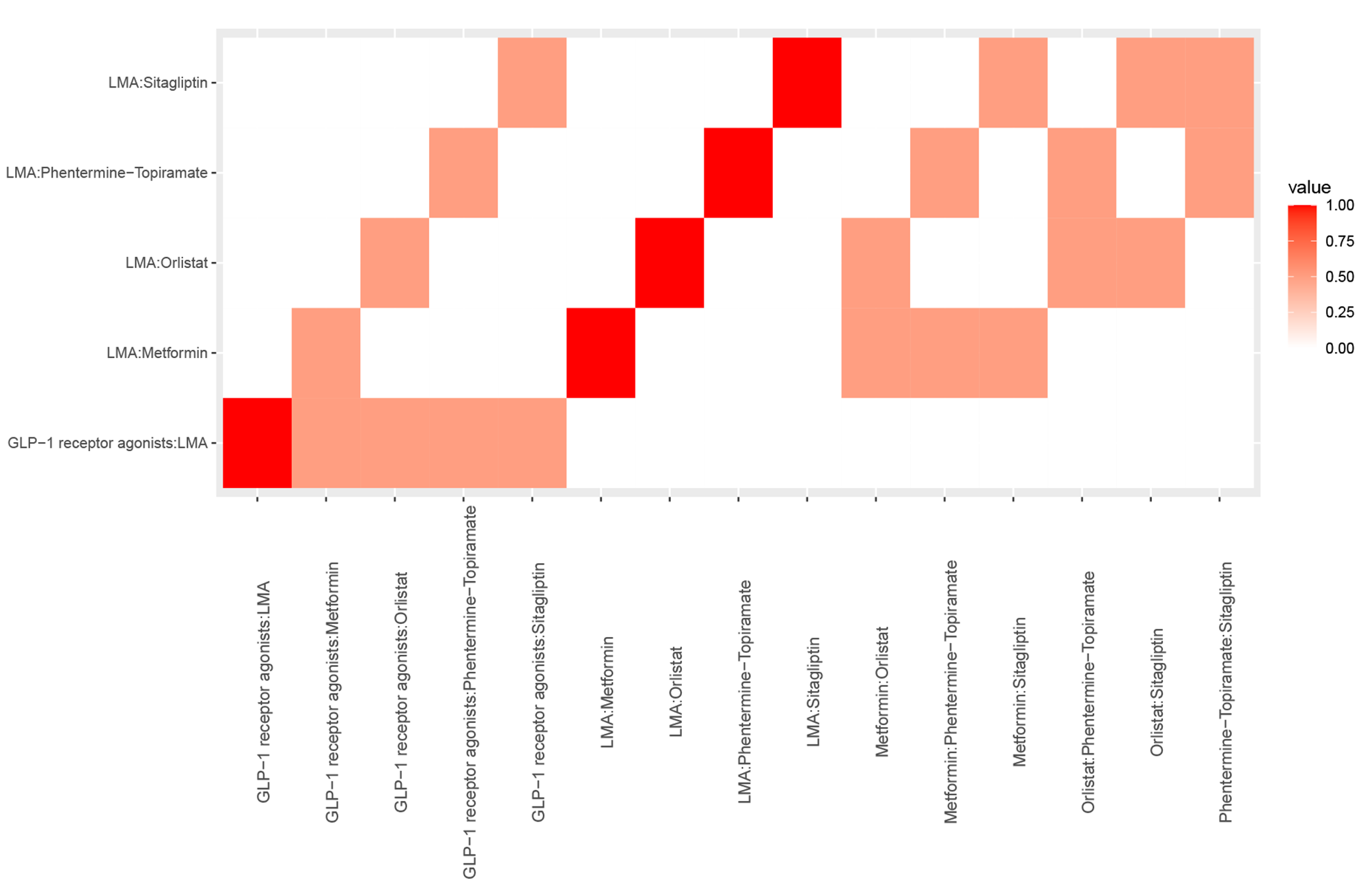

**Outcome: nausea events**

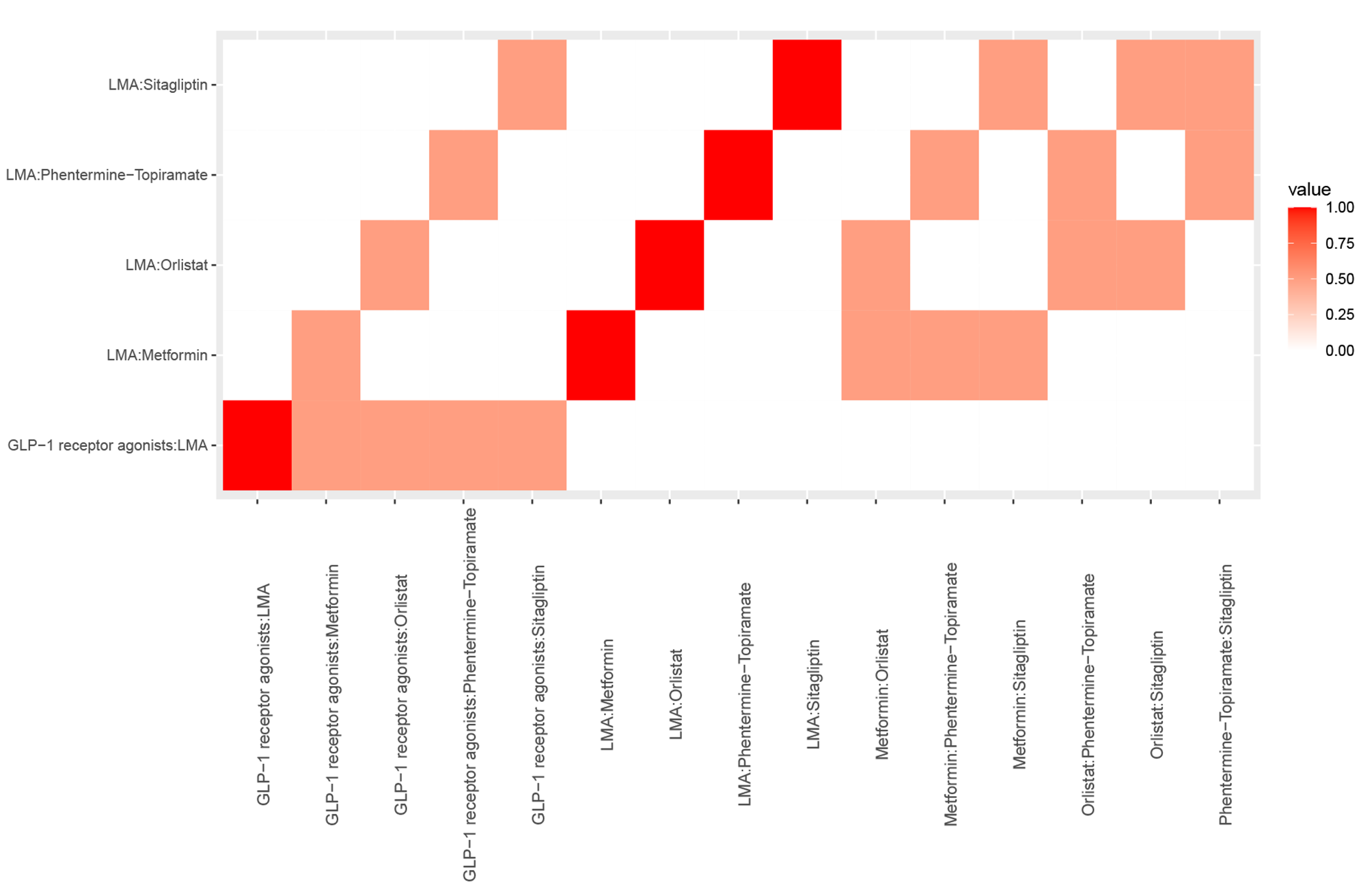

**Outcome: vomiting events**

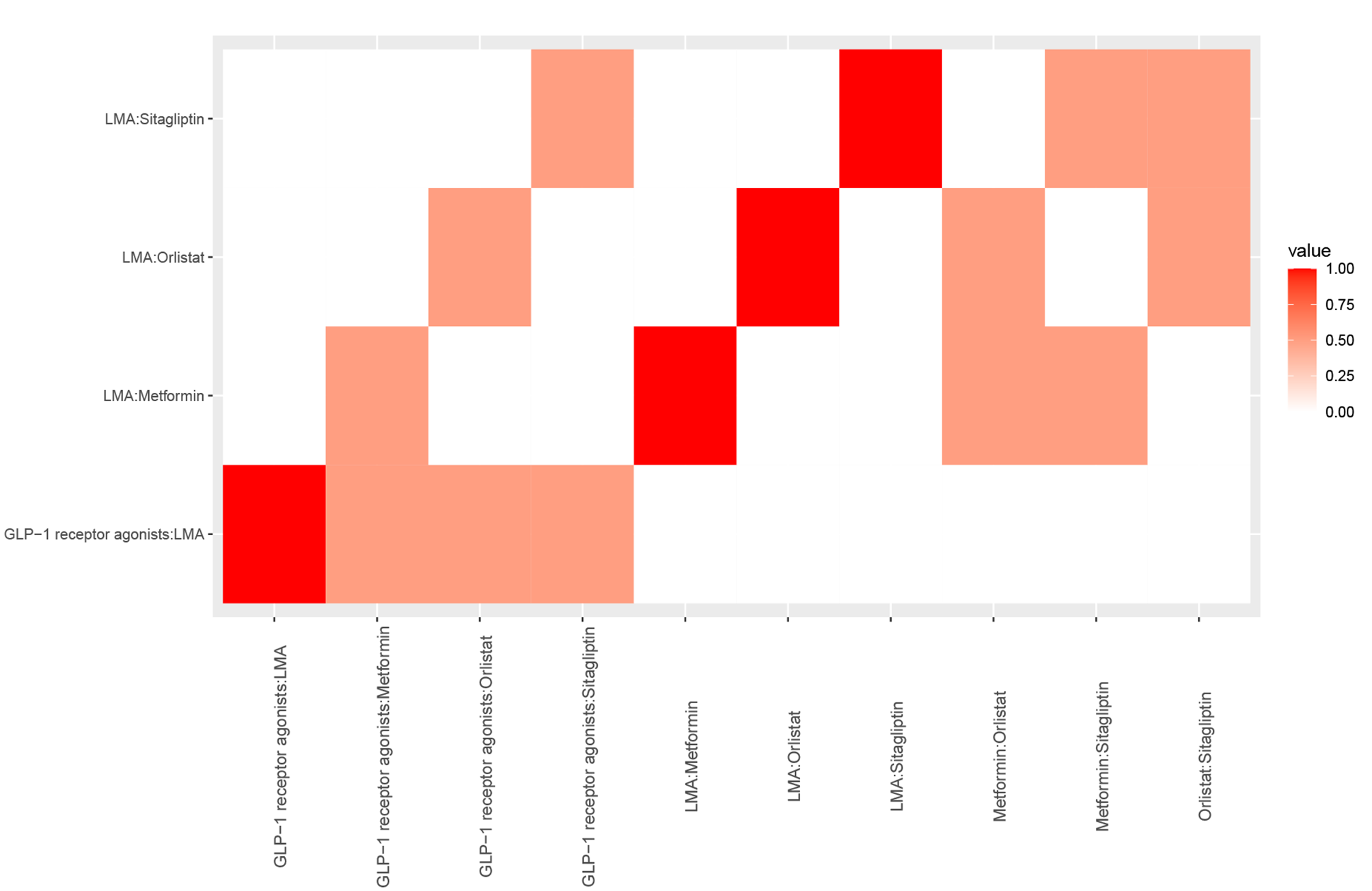

**Outcome: diarrhea events**

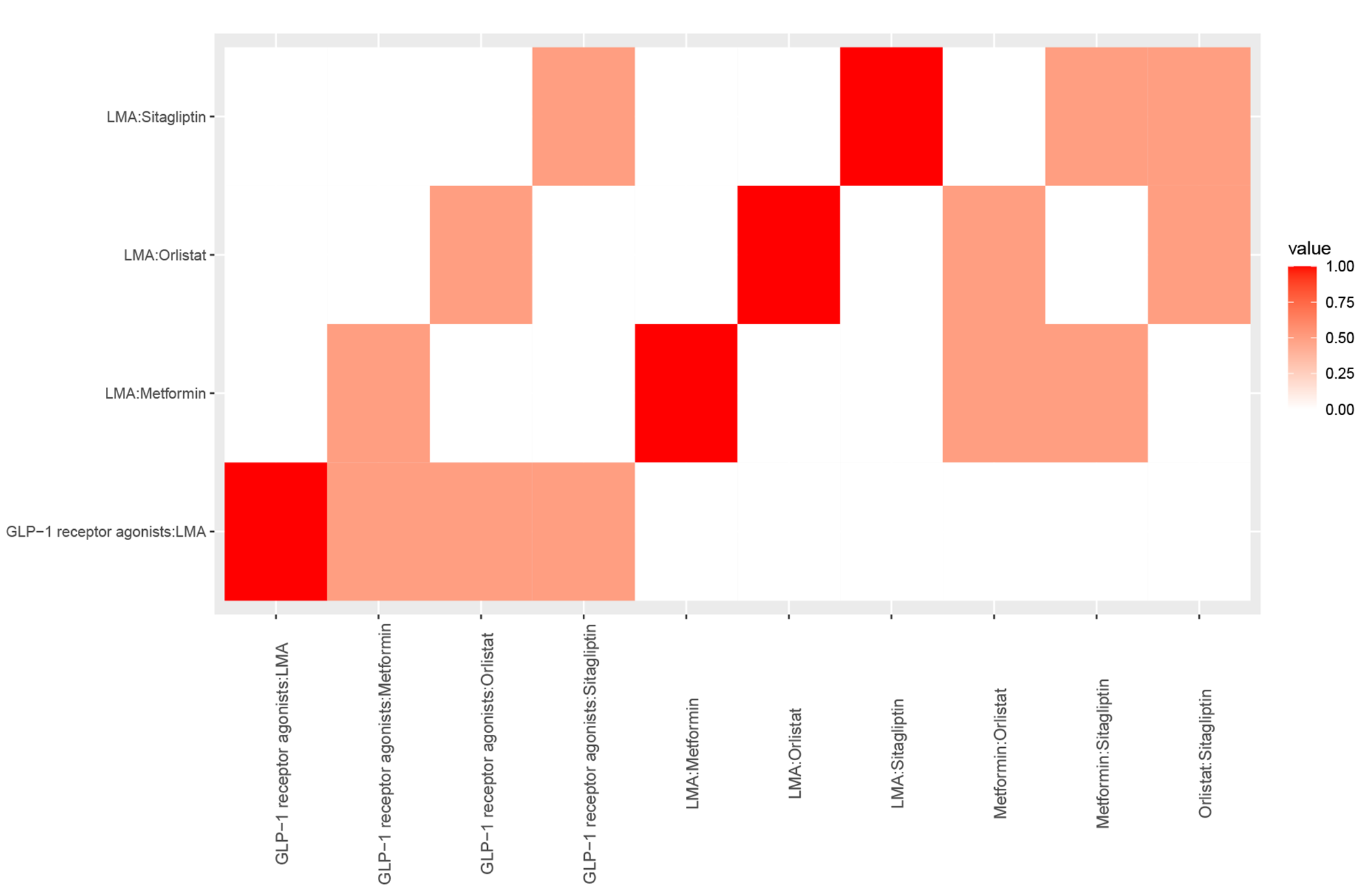

### Appendix 8: GRADE assessments

**Notes:** a, Risk of bias; b, Heterogeneity; c, Publication bias; d, Contributing direct evidence of moderate quality; e, Contributing direct evidence of low quality; f, Contributing direct evidence of very low quality; g, Inconsistency; h, Imprecision.

**Outcome:** **change in BMI from baseline**

| **Comparison** | **Direct estimate** | **Certainty** | **Indirect estimate** | **Certainty** | **Network estimate** | **Certainty** |
| --- | --- | --- | --- | --- | --- | --- |
| Fluoxetine vs GLP-1 receptor agonists | - | - | 0.11 (-2.57, 2.79) | Moderate^d^ | 0.11 (-2.57, 2.79) | Low^h^ |
| Fluoxetine vs Lifestyle modification alone | -0.80 (-3.42, 1.82) | High | -4.66 (-9.43, 0.10) | Moderate^d^ | -1.70 (-4.00, 0.60) | Moderate^h^ |
| Fluoxetine vs Metformin | -1.5 (-4.13, 1.13) | High | 2.36 (-2.40, 7.13) | Moderate^d^ | -0.60 (-2.90, 1.70) | Moderate^h^ |
| Fluoxetine vs Metformin combined with Fluoxetine | 0.30 (-2.32, 2.92) | High | - | - | 0.30 (-2.32, 2.92) | Moderate^h^ |
| Fluoxetine vs Orlistat | - | - | -0.03 (-2.82, 2.75) | Low^de^ | -0.03 (-2.82, 2.75) | Very Low^h^ |
| Fluoxetine vs Phentermine-Topiramate | - | - | 3.13 (-0.36, 6.63) | Moderate^d^ | 3.13 (-0.36, 6.63) | Low^h^ |
| Fluoxetine vs Sitagliptin | - | - | -2.40 (-5.94, 1.14) | Moderate^d^ | -2.40 (-5.94, 1.14) | Low^h^ |
| Fluoxetine vs Topiramate | - | - | -0.84 (-4.57, 2.89) | Moderate^d^ | -0.84 (-4.57, 2.89) | Low^h^ |
| GLP-1 receptor agonists vs Lifestyle modification alone | -1.81 (-3.19, -0.43) | High | - | - | -1.81 (-3.19, -0.43) | High |
| GLP-1 receptor agonists vs Metformin | - | - | -0.71 (-2.25, 0.83) | Moderate^d^ | -0.71 (-2.25, 0.83) | Low^h^ |
| GLP-1 receptor agonists vs Metformin combined with Fluoxetine | - | - | 0.19 (-2.49, 2.87) | Moderate^d^ | 0.19 (-2.49, 2.87) | Low^h^ |
| GLP-1 receptor agonists vs Orlistat | - | - | -0.14 (-2.23, 1.95) | Low^e^ | -0.14 (-2.23, 1.95) | Very Low^h^ |
| GLP-1 receptor agonists vs Phentermine-Topiramate | - | - | 3.02 ( 0.05, 5.99) | High | 3.02 ( 0.05, 5.99) | High |
| GLP-1 receptor agonists vs Sitagliptin | - | - | -2.51 (-5.53, 0.51) | High | -2.51 (-5.53, 0.51) | Moderate^h^ |
| GLP-1 receptor agonists vs Topiramate | - | - | -0.95 (-4.20, 2.30) | High | -0.95 (-4.20, 2.30) | Moderate^h^ |
| Metformin vs Lifestyle modification alone | -1.10 (-1.79, -0.41) | Moderate^b^ | - | - | -1.10 (-1.79, -0.41) | Moderate |
| Metformin vs Metformin combined with Fluoxetine | 1.80 (-0.82, 4.42) | High | -2.06 (-6.83, 2.70) | Moderate^d^ | 0.90 (-1.40, 3.20) | Moderate^h^ |
| Metformin vs Orlistat | - | - | 0.57 (-1.15, 2.28) | Low^de^ | 0.57 (-1.15, 2.28) | Very Low^h^ |
| Metformin vs Phentermine-Topiramate | - | - | 3.73 ( 1.01, 6.45) | Moderate^d^ | 3.73 ( 1.01, 6.45) | Moderate |
| Metformin vs Sitagliptin | - | - | -1.80 (-4.57, 0.98) | Moderate^d^ | -1.80 (-4.57, 0.98) | Low^h^ |
| Metformin vs Topiramate | - | - | -0.24 (-3.26, 2.78) | Moderate^d^ | -0.24 (-3.26, 2.78) | Low^h^ |
| Metformin combined with Fluoxetine vs Lifestyle modification alone | -1.10(-3.72, 1.52) | High | -4.96 (-9.73, -0.20) | Moderate^d^ | -2.00 (-4.30, 0.30) | Moderate^h^ |
| Metformin combined with Fluoxetine vs Orlistat | - | - | -0.33 (-3.12, 2.45) | Low^de^ | -0.33 (-3.12, 2.45) | Very Low^h^ |
| Metformin combined with Fluoxetine vs Phentermine-Topiramate | - | - | 2.83 (-0.66, 6.33) | Moderate^d^ | 2.83 (-0.66, 6.33) | Low^h^ |
| Metformin combined with Fluoxetine vs Sitagliptin | - | - | -2.70 (-6.24, 0.84) | Moderate^d^ | -2.70 (-6.24, 0.84) | Low^h^ |
| Metformin combined with Fluoxetine vs Topiramate | - | - | -1.14 (-4.87, 2.59) | Moderate^d^ | -1.14 (-4.87, 2.59) | Low^h^ |
| Orlistat vs Lifestyle modification alone | -1.66 (-3.23, -0.09) | Low^ab^ | - | - | -1.66 (-3.23, -0.09) | Low |
| Orlistat vs Phentermine-Topiramate | - | - | 3.17 ( 0.10, 6.23) | Low^e^ | 3.17 ( 0.10, 6.23) | Low |
| Orlistat vs Sitagliptin | - | - | -2.36 (-5.48, 0.75) | Low^e^ | -2.36 (-5.48, 0.75) | Very Low^h^ |
| Orlistat vs Topiramate | - | - | -0.80 (-4.14, 2.53) | Low^e^ | -0.80 (-4.14, 2.53) | Very Low^h^ |
| Phentermine-Topiramate vs Lifestyle modification alone | -4.83 (-7.46, -2.20) | High | - | - | -4.83 (-7.46, -2.20) | High |
| Phentermine-Topiramate vs Sitagliptin | - | - | -5.53 (-9.29, -1.77) | High | -5.53 (-9.29, -1.77) | High |
| Phentermine-Topiramate vs Topiramate | - | - | -3.97 (-7.91, -0.03) | High | -3.97 (-7.91, -0.03) | High |
| Sitagliptin vs Lifestyle modification alone | 0.70 (-1.99, 3.39) | High | - | - | 0.70 (-1.99, 3.39) | Moderate^h^ |
| Sitagliptin vs Topiramate | - | - | 1.56 (-2.42, 5.54) | High | 1.56 (-2.42, 5.54) | Moderate^h^ |
| Topiramate vs Lifestyle modification alone | -0.86 (-3.80, 2.08) | High | - | - | -0.86 (-3.80, 2.08) | Moderate^h^ |

**Outcome: change in weight from baseline**

| **Comparison** | **Direct estimate** | **Certainty** | **Indirect estimate** | **Certainty** | **Network estimate** | **Certainty** |
| --- | --- | --- | --- | --- | --- | --- |
| GLP-1 receptor agonists vs Lifestyle modification alone | -3.82 (-6.47, -1.16) | High | - | - | -3.82 (-6.47, -1.16) | High |
| GLP-1 receptor agonists vs Metformin | - | - | -1.25 (-4.38, 1.89) | Moderate^d^ | -1.25 (-4.38, 1.89) | Low^h^ |
| GLP-1 receptor agonists vs Orlistat | - | - | 0.46 (-3.42, 4.35) | Low^e^ | 0.46 (-3.42, 4.35) | Very Low^h^ |
| GLP-1 receptor agonists vs Phentermine-Topiramate | - | - | 10.77 (5.30, 16.24) | High | 10.77 (5.30, 16.24) | High |
| GLP-1 receptor agonists vs Sitagliptin | - | - | -5.62 (-11.20, -0.03) | High | -5.62 (-11.20, -0.03) | High |
| GLP-1 receptor agonists vs Topiramate | - | - | -1.39 ( -8.04, 5.26) | High | -1.39 ( -8.04, 5.26) | Moderate^h^ |
| Metformin vs Lifestyle modification alone | -2.57 ( -4.23, -0.91) | Moderate^b^ | - | - | -2.57 ( -4.23, -0.91) | Moderate |
| Metformin vs Orlistat | - | - | 1.71 (-1.58, 5.00) | Low^de^ | 1.71 (-1.58, 5.00) | Very Low^h^ |
| Metformin vs Phentermine-Topiramate | - | - | 12.02 (6.96, 17.08) | Moderate^d^ | 12.02 (6.96, 17.08) | Moderate |
| Metformin vs Sitagliptin | - | - | -4.37 ( -9.55, 0.81) | Moderate^d^ | -4.37 ( -9.55, 0.81) | Low^h^ |
| Metformin vs Topiramate | - | - | -0.14 ( -6.46, 6.18) | Moderate^d^ | -0.14 ( -6.46, 6.18) | Low^h^ |
| Orlistat vs Lifestyle modification alone | -4.28 ( -7.12, -1.44) | Low^ab^ | - | - | -4.28 ( -7.12, -1.44) | Low |
| Orlistat vs Phentermine-Topiramate | - | - | 10.31 (4.75, 15.87) | Low^e^ | 10.31 (4.75, 15.87) | Low |
| Orlistat vs Sitagliptin | - | - | -6.08 (-11.75, -0.41) | Low^e^ | -6.08 (-11.75, -0.41) | Low |
| Orlistat vs Topiramate | - | - | -1.85 ( -8.57, 4.88) | Low^e^ | -1.85 ( -8.57, 4.88) | Very Low^h^ |
| Phentermine-Topiramate vs Lifestyle modification alone | -14.59 (-19.37, -9.81) | High | - | - | -14.59 (-19.37, -9.81) | High |
| Phentermine-Topiramate vs Sitagliptin |  |  | -16.39 (-23.24, -9.54) | High | -16.39 (-23.24, -9.54) | High |
| Phentermine-Topiramate vs Topiramate | - | - | -12.16 (-19.91, -4.41) | High | -12.16 (-19.91, -4.41) | High |
| Sitagliptin vs Lifestyle modification alone | 1.80 (-3.11, 6.71) | High | - | - | 1.80 (-3.11, 6.71) | Moderate^h^ |
| Sitagliptin vs Topiramate | - | - | 4.23 (-3.60, 12.06) | High | 4.23 (-3.60, 12.06) | Moderate^h^ |
| Topiramate vs Lifestyle modification alone | -2.43 (-8.53, 3.67) | High | - | - | -2.43 (-8.53, 3.67) | Moderate^h^ |

**Outcome: percentage of participants achieving BMI reduction of at least 5%**

| **Comparison** | **Direct estimate** | **Certainty** | **Indirect estimate** | **Certainty** | **Network estimate** | **Certainty** |
| --- | --- | --- | --- | --- | --- | --- |
| GLP-1 receptor agonists vs Lifestyle modification alone | 4.86 (1.93, 12.24) | Moderate^b^ | - | - | 4.86 (1.93, 12.24) | Moderate |
| GLP-1 receptor agonists vs Orlistat | - | - | 2.58 (0.48, 13.90) | Moderate^d^ | 2.58 (0.48, 13.90) | Low^h^ |
| GLP-1 receptor agonists vs Phentermine-Topiramate | - | - | 0.35 (0.05, 2.60) | Moderate^d^ | 0.35 (0.05, 2.60) | Low^h^ |
| GLP-1 receptor agonists vs Topiramate | - | - | 2.43 (0.20, 28.96) | Moderate^d^ | 2.43 (0.20, 28.96) | Low^h^ |
| Orlistat vs Lifestyle modification alone | 1.88 (0.46, 7.68) | High | - | - | 1.88 (0.46, 7.68) | Moderate^h^ |
| Orlistat vs Phentermine-Topiramate | - | - | 0.13 (0.01, 1.31) | High | 0.13 (0.01, 1.31) | Moderate^h^ |
| Orlistat vs Topiramate | - | - | 0.94 (0.06, 13.94) | High | 0.94 (0.06, 13.94) | Moderate^h^ |
| Phentermine-Topiramate vs Lifestyle modification alone | 14.06 (2.34, 84.35) | High | - | - | 14.06 (2.34, 84.35) | High |
| Phentermine-Topiramate vs Topiramate | - | - | 7.03 (0.38, 129.65) | High | 7.03 (0.38, 129.65) | Moderate^h^ |
| Topiramate vs Lifestyle modification alone | 2.00 (0.20, 19.93) | High | - | - | 2.00 (0.20, 19.93) | Moderate^h^ |

**Outcome: percentage of participants achieving BMI reduction of at least 10%**

| **Comparison** | **Direct estimate** | **Certainty** | **Indirect estimate** | **Certainty** | **Network estimate** | **Certainty** |
| --- | --- | --- | --- | --- | --- | --- |
| GLP-1 receptor agonists vs Lifestyle modification alone | 5.1996(1.4400, 18.7747) | Moderate^b^ | - | - | 5.1996(1.4400, 18.7747) | Moderate |
| GLP-1 receptor agonists vs Orlistat | - | - | 1.5995(0.1418, 18.0367) | Moderate^d^ | 1.5995(0.1418, 18.0367) | Low^h^ |
| GLP-1 receptor agonists vs Phentermine-Topiramate | - | - | 0.0720(0.0019, 2.6937) | Moderate^d^ | 0.0720(0.0019, 2.6937) | Low^h^ |
| GLP-1 receptor agonists vs Topiramate | - | - | 5.9995(0.1521, 236.6757) | Moderate^d^ | 5.9995(0.1521, 236.6757) | Low^h^ |
| Orlistat vs Lifestyle modification alone | 3.2508(0.4166, 25.3670) | High | - | - | 3.2508(0.4166, 25.3670) | Moderate^h^ |
| Orlistat vs Phentermine-Topiramate | - | - | 0.0450(0.0009, 2.3644) | High | 0.0450(0.0009, 2.3644) | Moderate^h^ |
| Orlistat vs Topiramate | - | - | 3.7509(0.0680, 206.8080) | High | 3.7509(0.0680, 206.8080) | Moderate^h^ |
| Phentermine-Topiramate vs Lifestyle modification alone | 72.2098(2.4421, 2135.1130) | High | - | - | 72.2098(2.4421, 2135.1130) | High |
| Phentermine-Topiramate vs Topiramate | - | - | 83.3189(0.6656, 10430.3372) | High | 83.3189(0.6656, 10430.3372) | Moderate^h^ |
| Topiramate vs Lifestyle modification alone | 0.8667(0.0277, 27.1216) | High | - | - | 0.8667(0.0277, 27.1216) | Moderate^h^ |

**Outcome: change in BMI z-score from baseline**

| **Comparison** | **Direct estimate** | **Certainty** | **Indirect estimate** | **Certainty** | **Network estimate** | **Certainty** |
| --- | --- | --- | --- | --- | --- | --- |
| Metformin vs Lifestyle modification alone | -0.16 (-0.21, -0.10) | High | - | - | -0.16 (-0.21, -0.10) | High |
| Metformin vs Orlistat | - | - | -0.11 (-0.17, -0.05) | High | -0.11 (-0.17, -0.05) | High |
| Metforminvs Topiramate | - | - | -0.13 (-0.22, -0.04) | High | -0.13 (-0.22, -0.04) | High |
| Orlistat vs Lifestyle modification alone | -0.05 (-0.08, -0.02) | High | - | - | -0.05 (-0.08, -0.02) | High |
| Orlistat vs Topiramate | - | - | -0.02 (-0.10, 0.06) | High | -0.02 (-0.10, 0.06) | Moderate^h^ |
| Topiramate vs Lifestyle modification alone | -0.03 (-0.10, 0.04) | High | - | - | -0.03 (-0.10, 0.04) | Moderate^h^ |

**Outcome: change in BMI SDS from baseline**

| **Comparison** | **Direct estimate** | **Certainty** | **Indirect estimate** | **Certainty** | **Network estimate** | **Certainty** |
| --- | --- | --- | --- | --- | --- | --- |
| GLP-1 receptor agonists vs Lifestyle modification alone | -0.15 (-0.25, -0.04) | High | - | - | -0.15 (-0.25, -0.04) | High |
| GLP-1 receptor agonists vs Metformin | - | - | -0.06 (-0.19, 0.06) | Moderate^d^ | -0.06 (-0.19, 0.06) | Low^h^ |
| GLP-1 receptor agonists vs Orlistat | - | - | -0.09 (-0.25, 0.07) | High | -0.09 (-0.25, 0.07) | Moderate^h^ |
| Metformin vs Lifestyle modification alone | -0.09 (-0.15, -0.02) | Moderate^b^ | - | - | -0.09 (-0.15, -0.02) | Moderate |
| Metformin vs Orlistat |  |  | -0.03 (-0.16, 0.11) | Moderate^d^ | -0.03 (-0.16, 0.11) | Low^h^ |
| Orlistat vs Lifestyle modification alone | -0.06 (-0.18, 0.06) | High | - | - | -0.06 (-0.18, 0.06) | Moderate^h^ |

**Outcome: total gastrointestinal adverse events**

| **Comparison** | **Direct estimate** | **Certainty** | **Indirect estimate** | **Certainty** | **Network estimate** | **Certainty** |
| --- | --- | --- | --- | --- | --- | --- |
| GLP-1 receptor agonists vs Lifestyle modification alone | 2.97 (1.93, 4.55) | High | - | - | 2.97 (1.93, 4.55) | High |
| GLP-1 receptor agonists vs Metformin | - | - | 2.11 (1.10, 4.04) | High | 2.11 (1.10, 4.04) | High |
| GLP-1 receptor agonists vs Orlistat | - | - | 0.07 (0.01, 0.74) | Low^e^ | 0.07 (0.01, 0.74) | Low |
| GLP-1 receptor agonists vs Phentermine-Topiramate | - | - | 3.18 (1.12, 9.05) | High | 3.18 (1.12, 9.05) | High |
| Metformin vs Lifestyle modification alone | 1.41 (0.86, 2.30) | High | - | - | 1.41 (0.86, 2.30) | Moderate^h^ |
| Metformin vs Orlistat | - | - | 0.031(0.003, 0.357) | Low^e^ | 0.031(0.003, 0.357) | Low |
| Metformin vs Phentermine-Topiramate | - | - | 1.51 (0.52, 4.41) | High | 1.51 (0.52, 4.41) | Moderate^h^ |
| Orlistat vs Lifestyle modification alone | 45.15 (4.14, 492.24) | Low^ab^ | - | - | 45.15 (4.14, 492.24) | Low |
| Orlistat vs Phentermine-Topiramate | - | - | 48.39 (3.69, 633.79) | Low^e^ | 48.39 (3.69, 633.79) | Low |
| Phentermine-Topiramate vs Lifestyle modification alone | 0.93 (0.36, 2.42) | High | - | - | 0.93 (0.36, 2.42) | Moderate^h^ |

**Outcome: discontinuation due to adverse events**

| **Comparison** | **Direct estimate** | **Certainty** | **Indirect estimate** | **Certainty** | **Network estimate** | **Certainty** |
| --- | --- | --- | --- | --- | --- | --- |
| GLP-1 receptor agonists vs Lifestyle modification alone | 2.03 (0.76, 5.42) | High | - | - | 2.03 (0.76, 5.42) | Moderate^h^ |
| GLP-1 receptor agonists vs Metformin | - | - | 1.06 (0.25, 4.44) | High | 1.06 (0.25, 4.44) | Moderate^h^ |
| GLP-1 receptor agonists vs Orlistat | - | - | 0.60 (0.13, 2.79) | Moderate^d^ | 0.60 (0.13, 2.79) | Low^h^ |
| GLP-1 receptor agonists vs Phentermine-Topiramate | - | - | 4.23 (0.47, 37.79) | High | 4.23 (0.47, 37.79) | Moderate^h^ |
| GLP-1 receptor agonists vs Sitagliptin | - | - | 0.66 (0.05, 8.54) | High | 0.66 (0.05, 8.54) | Moderate^h^ |
| Metformin vs Lifestyle modification alone | 1.92 (0.68, 5.43) | High | - | - | 1.92 (0.68, 5.43) | Moderate^h^ |
| Metformin vs Orlistat | - | - | 0.57 (0.12, 2.74) | Moderate^d^ | 0.57 (0.12, 2.74) | Low^h^ |
| Metformin vs Phentermine-Topiramate | - | - | 3.99 (0.43, 36.66) | High | 3.99 (0.43, 36.66) | Moderate^h^ |
| Metformin vs Sitagliptin | - | - | 0.62 (0.05, 8.25) | High | 0.62 (0.05, 8.25) | Moderate^h^ |
| Orlistat vs Lifestyle modification alone | 3.39 (1.04, 11.09) | Moderate^a^ | - | - | 3.39 (1.04, 11.09) | Moderate |
| Orlistat vs Phentermine-Topiramate | - | - | 7.06 (0.72, 69.59) | Moderate^d^ | 7.06 (0.72, 69.59) | Low^h^ |
| Orlistat vs Sitagliptin | - | - | 1.11 (0.08, 15.51) | Moderate^d^ | 1.11 (0.08, 15.51) | Low^h^ |
| Phentermine-Topiramate vs Lifestyle modification alone | 0.48 (0.07, 3.40) | High | - | - | 0.48 (0.07, 3.40) | Moderate^h^ |
| Phentermine-Topiramate vs Sitagliptin | - | - | 0.16 (0.01, 3.36) | High | 0.16 (0.01, 3.36) | Moderate^h^ |
| Sitagliptin vs Lifestyle modification alone | 3.07 (0.29, 32.45) | High | - | - | 3.07 (0.29, 32.45) | Moderate^h^ |

**Outcome: serious adverse events**

| **Comparison** | **Direct estimate** | **Certainty** | **Indirect estimate** | **Certainty** | **Network estimate** | **Certainty** |
| --- | --- | --- | --- | --- | --- | --- |
| GLP-1 receptor agonists vs Lifestyle modification alone | 1.07 (0.60, 1.91) | High | - | - | 1.07 (0.60, 1.91) | Moderate^h^ |
| GLP-1 receptor agonists vs Metformin | - | - | 0.36 (0.01, 9.53) | High | 0.36 (0.01, 9.53) | Moderate^h^ |
| GLP-1 receptor agonists vs Orlistat | - | - | 1.15 (0.36, 3.67) | High | 1.15 (0.36, 3.67) | Moderate^h^ |
| GLP-1 receptor agonists vs Phentermine-Topiramate | - | - | 0.65 (0.07, 6.51) | High | 0.65 (0.07, 6.51) | Moderate^h^ |
| GLP-1 receptor agonists vs Sitagliptin | - | - | 0.53 (0.09, 3.23) | High | 0.53 (0.09, 3.23) | Moderate^h^ |
| Metformin vs Lifestyle modification alone | 3.00 (0.12, 75.96) | High | - | - | 3.00 (0.12, 75.96) | Moderate^h^ |
| Metformin vs Orlistat | - | - | 3.21 (0.11, 94.76) | High | 3.21 (0.11, 94.76) | Moderate^h^ |
| Metformin vs Phentermine-Topiramate | - | - | 1.82 (0.04, 92.30) | High | 1.82 (0.04, 92.30) | Moderate^h^ |
| Metformin vs Sitagliptin | - | - | 1.47 (0.04, 57.14) | High | 1.47 (0.04, 57.14) | Moderate^h^ |
| Orlistat vs Lifestyle modification alone | 0.94 (0.34, 2.57) | High | - | - | 0.94 (0.34, 2.57) | Moderate^h^ |
| Orlistat vs Phentermine-Topiramate | - | - | 0.57 (0.05, 6.57) | High | 0.57 (0.05, 6.57) | Moderate^h^ |
| Orlistat vs Sitagliptin | - | - | 0.46 (0.06, 3.37) | High | 0.46 (0.06, 3.37) | Moderate^h^ |
| Phentermine-Topiramate vs Lifestyle modification alone | 1.65 (0.18, 15.30) | High | - | - | 1.65 (0.18, 15.30) | Moderate^h^ |
| Phentermine-Topiramate vs Sitagliptin | - | - | 0.81 (0.05, 13.47) | High | 0.81 (0.05, 13.47) | Moderate^h^ |
| Sitagliptin vs Lifestyle modification alone | 2.04 (0.37, 11.44) | High | - | - | 2.04 (0.37, 11.44) | Moderate^h^ |

**Outcome: nausea events**

| **Comparison** | **Direct estimate** | **Certainty** | **Indirect estimate** | **Certainty** | **Network estimate** | **Certainty** |
| --- | --- | --- | --- | --- | --- | --- |
| GLP-1 receptor agonists vs Lifestyle modification alone | 3.30 (2.31, 4.72) | High | - | - | 3.30 (2.31, 4.72) | High |
| GLP-1 receptor agonists vs Metformin | - | - | 0.89 (0.36, 2.24) | High | 0.89 (0.36, 2.24) | Moderate^h^ |
| GLP-1 receptor agonists vs Orlistat | - | - | 2.81 (1.57, 5.04) | High | 2.81 (1.57, 5.04) | High |
| GLP-1 receptor agonists vs Phentermine-Topiramate | - | - | 2.80 (0.54, 14.43) | High | 2.80 (0.54, 14.43) | Moderate^h^ |
| GLP-1 receptor agonists vs Sitagliptin | - | - | 0.63 (0.07, 5.68) | High | 0.63 (0.07, 5.68) | Moderate^h^ |
| Metformin vs Lifestyle modification alone | 3.69 (1.59, 8.59) | High | - | - | 3.69 (1.59, 8.59) | High |
| Metformin vs Orlistat | - | - | 3.14 (1.20, 8.23) | High | 3.14 (1.20, 8.23) | High |
| Metformin vs Phentermine-Topiramate | - | - | 3.13 (0.51, 19.11) | High | 3.13 (0.51, 19.11) | Moderate^h^ |
| Metformin vs Sitagliptin | - | - | 0.71 (0.07, 7.23) | High | 0.71 (0.07, 7.23) | Moderate^h^ |
| Orlistat vs Lifestyle modification alone | 1.17 (0.74, 1.86) | High | - | - | 1.17 (0.74, 1.86) | Moderate^h^ |
| Orlistat vs Phentermine-Topiramate | - | - | 0.99 (0.19, 5.26) | High | 0.99 (0.19, 5.26) | Moderate^h^ |
| Orlistat vs Sitagliptin | - | - | 0.22 (0.02, 2.06) | High | 0.22 (0.02, 2.06) | Moderate^h^ |
| Phentermine-Topiramate vs Lifestyle modification alone | 1.18 (0.24, 5.86) | High | - | - | 1.18 (0.24, 5.86) | Moderate^h^ |
| Phentermine-Topiramate vs Sitagliptin | - | - | 0.23 (0.02, 3.35) | High | 0.23 (0.02, 3.35) | Moderate^h^ |
| Sitagliptin vs Lifestyle modification alone | 5.22 (0.60, 45.57) | High | - | - | 5.22 (0.60, 45.57) | Moderate^h^ |

**Outcome: vomiting events**

| **Comparison** | **Direct estimate** | **Certainty** | **Indirect estimate** | **Certainty** | **Network estimate** | **Certainty** |
| --- | --- | --- | --- | --- | --- | --- |
| GLP-1 receptor agonists vs Lifestyle modification alone | 5.62 (3.54, 8.93) | High | - | - | 5.62 (3.54, 8.93) | High |
| GLP-1 receptor agonists vs Metformin | - | - | 0.84 (0.09, 7.67) | High | 0.84 (0.09, 7.67) | Moderate^h^ |
| GLP-1 receptor agonists vs Orlistat | - | - | 5.62 (1.72, 18.31) | High | 5.62 (1.72, 18.31) | High |
| GLP-1 receptor agonists vs Sitagliptin | - | - | 2.75 (0.46, 16.35) | High | 2.75 (0.46, 16.35) | Moderate^h^ |
| Metformin vs Lifestyle modification alone | 6.73 (0.77, 58.82) | High | - | - | 6.73 (0.77, 58.82) | Moderate^h^ |
| Metformin vs Orlistat | - | - | 6.73 (0.60, 76.06) | High | 6.73 (0.60, 76.06) | Moderate^h^ |
| Metformin vs Sitagliptin | - | - | 3.29 (0.21, 52.46) | High | 3.29 (0.21, 52.46) | Moderate^h^ |
| Orlistat vs Lifestyle modification alone | 1.00 (0.34, 2.96) | High | - | - | 1.00 (0.34, 2.96) | Moderate^h^ |
| Orlistat vs Sitagliptin | - | - | 0.49 (0.06, 3.75) | High | 0.49 (0.06, 3.75) | Moderate^h^ |
| Sitagliptin vs Lifestyle modification alone | 2.04 (0.37, 11.44) | High | - | - | 2.04 (0.37, 11.44) | Moderate^h^ |

**Outcome: diarrhea events**

| **Comparison** | **Direct estimate** | **Certainty** | **Indirect estimate** | **Certainty** | **Network estimate** | **Certainty** |
| --- | --- | --- | --- | --- | --- | --- |
| GLP-1 receptor agonists vs Lifestyle modification alone | 1.66 (1.17, 2.35) | High | - | - | 1.66 (1.17, 2.35) | High |
| GLP-1 receptor agonists vs Metformin | - | - | 1.08 (0.48, 2.41) | High | 1.08 (0.48, 2.41) | Moderate^h^ |
| GLP-1 receptor agonists vs Orlistat | - | - | 0.54 (0.21, 1.38) | High | 0.54 (0.21, 1.38) | Moderate^h^ |
| GLP-1 receptor agonists vs Sitagliptin | - | - | 2.83 (0.63, 12.69) | High | 2.83 (0.63, 12.69) | Moderate^h^ |
| Metformin vs Lifestyle modification alone | 1.54 (0.74, 3.17) | High | - | - | 1.54 (0.74, 3.17) | Moderate^h^ |
| Metformin vs Orlistat | - | - | 0.50 (0.16, 1.56) | High | 0.50 (0.16, 1.56) | Moderate^h^ |
| Metformin vs Sitagliptin | - | - | 2.62 (0.51, 13.35) | High | 2.62 (0.51, 13.35) | Moderate^h^ |
| Orlistat vs Lifestyle modification alone | 3.06 (1.28, 7.28) | High | - | - | 3.06 (1.28, 7.28) | High |
| Orlistat vs Sitagliptin | - | - | 5.21 (0.95, 28.48) | High | 5.21 (0.95, 28.48) | Moderate^h^ |
| Sitagliptin vs Lifestyle modification alone | 0.59 (0.14, 2.53) | High | - | - | 0.59 (0.14, 2.53) | Moderate^h^ |

### Appendix 9: Meta-regression analyses

| **Change in BMI from baseline (mean age at baseline)** | | | | | | | | | | | | | | | | | | | | | | | | | | | | | | | | | | |
| --- | --- | --- | --- | --- | --- | --- | --- | --- | --- | --- | --- | --- | --- | --- | --- | --- | --- | --- | --- | --- | --- | --- | --- | --- | --- | --- | --- | --- | --- | --- | --- | --- | --- | --- |
| Mixed-Effects Model (k = 66; tau^2 estimator: REML) | | | | | | | | | | | | | | | | | | | | | | | | | | | | | | | | | | |
| logLik | | | | | | | | | | | deviance | | | | | AIC | | | | | | | | | BIC | | | | | | AICc | | | |
| -127.0242 | | | | | | | | | | | 254.0484 | | | | | 260.0484 | | | | | | | | | 266.5250 | | | | | | 260.4484 | | | |
| tau^2 (estimated amount of residual heterogeneity): 2.1007 (SE = 0.5481); tau (square root of estimated tau^2 value): 1.4494;  I^2 (residual heterogeneity / unaccounted variability): 67.75%; H^2 (unaccounted variability / sampling variability): 3.10; R^2 (amount of heterogeneity accounted for): 12.35% | | | | | | | | | | | | | | | | | | | | | | | | | | | | | | | | | | |
| Test for Residual Heterogeneity: QE(df = 64) = 198.4449, p-val < .0001 | | | | | | | | | | | | | | | | | | | | | | | | | | | | | | | | | | |
| Test of Moderators (coefficient 2): QM(df = 1) = 7.2028, p-val = 0.0073 | | | | | | | | | | | | | | | | | | | | | | | | | | | | | | | | | | |
| Model Results: | | | | | | | | | | | | | | | | | | | | | | | | | | | | | | | | | | |
|  | | | | | | | | estimate | | | | | | se | | | | | zval | | | pval | | | | | | ci.lb | | | | | ci.ub | |
| intrcpt | | | | | | | | -6.5677 | | | | | | 2.2902 | | | | | -2.8677 | | | 0.0041 | | | | | | -11.0565 | | | | | -2.0790 | |
| Mean age | | | | | | | | 0.4455 | | | | | | 0.1660 | | | | | 2.6838 | | | 0.0073 | | | | | | 0.1201 | | | | | 0.7708 | |
| **Change in BMI from baseline (mean BMI at baseline)** | | | | | | | | | | | | | | | | | | | | | | | | | | | | | | | | | | |
| Mixed-Effects Model (k = 66; tau^2 estimator: REML) | | | | | | | | | | | | | | | | | | | | | | | | | | | | | | | | | | |
| logLik | | | | | | | | | | | deviance | | | | | AIC | | | | | | | | | BIC | | | | | | AICc | | | |
| -129.5404 | | | | | | | | | | | 259.0808 | | | | | 265.0808 | | | | | | | | | 271.5574 | | | | | | 265.4808 | | | |
| tau^2 (estimated amount of residual heterogeneity): 2.3544 (SE = 0.5930); tau (square root of estimated tau^2 value): 1.5344;  I^2 (residual heterogeneity / unaccounted variability): 70.19%; H^2 (unaccounted variability / sampling variability): 3.35; R^2 (amount of heterogeneity accounted for): 0.00% | | | | | | | | | | | | | | | | | | | | | | | | | | | | | | | | | | |
| Test for Residual Heterogeneity: QE(df = 64) = 214.6788, p-val < .0001 | | | | | | | | | | | | | | | | | | | | | | | | | | | | | | | | | | |
| Test of Moderators (coefficient 2): QM(df = 1) = 0.9859, p-val = 0.3207 | | | | | | | | | | | | | | | | | | | | | | | | | | | | | | | | | | |
| Model Results: | | | | | | | | | | | | | | | | | | | | | | | | | | | | | | | | | | |
|  | | | | | | | | estimate | | | | | | se | | | | | zval | | | pval | | | | | | ci.lb | | | | | ci.ub | |
| intrcpt | | | | | | | | -2.0576 | | | | | | 1.6691 | | | | | -1.2327 | | | 0.2177 | | | | | | -5.3290 | | | | | 1.2138 | |
| Mean BMI at baseline | | | | | | | | 0.0499 | | | | | | 0.0503 | | | | | 0.9929 | | | 0.3207 | | | | | | -0.0486 | | | | | 0.1485 | |
| **Change in BMI from baseline (mean weight at baseline)** | | | | | | | | | | | | | | | | | | | | | | | | | | | | | | | | | | |
| Mixed-Effects Model (k = 44; tau^2 estimator: REML) | | | | | | | | | | | | | | | | | | | | | | | | | | | | | | | | | | |
| logLik | | | | | | | | | | | deviance | | | | | AIC | | | | | | | | | BIC | | | | | | AICc | | | |
| -87.4129 | | | | | | | | | | | 174.8258 | | | | | 180.8258 | | | | | | | | | 186.0388 | | | | | | 181.4573 | | | |
| tau^2 (estimated amount of residual heterogeneity): 2.7608 (SE = 0.8207); tau (square root of estimated tau^2 value): 1.6616;  I^2 (residual heterogeneity / unaccounted variability): 73.41%; H^2 (unaccounted variability / sampling variability): 3.76; R^2 (amount of heterogeneity accounted for): 5.33% | | | | | | | | | | | | | | | | | | | | | | | | | | | | | | | | | | |
| Test for Residual Heterogeneity: QE(df = 42) = 157.9551, p-val < .0001 | | | | | | | | | | | | | | | | | | | | | | | | | | | | | | | | | | |
| Test of Moderators (coefficient 2): QM(df = 1) = 2.7770, p-val = 0.0956 | | | | | | | | | | | | | | | | | | | | | | | | | | | | | | | | | | |
| Model Results: | | | | | | | | | | | | | | | | | | | | | | | | | | | | | | | | | | |
|  | | | | | | | | estimate | | | | | | se | | | | | zval | | | pval | | | | | | ci.lb | | | | | ci.ub | |
| intrcpt | | | | | | | | -3.1901 | | | | | | 1.7491 | | | | | -1.8239 | | | 0.0682 | | | | | | -6.6182 | | | | | 0.2381 | |
| Mean weight at baseline | | | | | | | | 0.0317 | | | | | | 0.0190 | | | | | 1.6664 | | | 0.0956 | | | | | | -0.0056 | | | | | 0.0690 | |
| **Change in BMI from baseline (proportion of males)** | | | | | | | | | | | | | | | | | | | | | | | | | | | | | | | | | | |
| Mixed-Effects Model (k = 56; tau^2 estimator: REML) | | | | | | | | | | | | | | | | | | | | | | | | | | | | | | | | | | |
| logLik | | | | | | | | | deviance | | | | | | AIC | | | | | | | | | BIC | | | | | | AICc | | | | |
| -113.2466 | | | | | | | | | 226.4931 | | | | | | 232.4931 | | | | | | | | | 238.4601 | | | | | | 232.9731 | | | | |
| tau^2 (estimated amount of residual heterogeneity): 2.8823 (SE = 0.7472); tau (square root of estimated tau^2 value): 1.6977;  I^2 (residual heterogeneity / unaccounted variability): 74.24%; H^2 (unaccounted variability / sampling variability): 3.88; R^2 (amount of heterogeneity accounted for): 0.00% | | | | | | | | | | | | | | | | | | | | | | | | | | | | | | | | | | |
| Test for Residual Heterogeneity: QE(df = 54) = 209.6467, p-val < .0001 | | | | | | | | | | | | | | | | | | | | | | | | | | | | | | | | | | |
| Test of Moderators (coefficient 2): QM(df = 1) = 0.0535, p-val = 0.8171 | | | | | | | | | | | | | | | | | | | | | | | | | | | | | | | | | | |
| Model Results: | | | | | | | | | | | | | | | | | | | | | | | | | | | | | | | | | | |
|  | estimate | | | | | | | | | | | se | | | | | | zval | | | pval | | | | | | ci.lb | | | | | | ci.ub | |
| intrcpt | -0.3214 | | | | | | | | | | | 0.7891 | | | | | | -0.4073 | | | 0.6838 | | | | | | -1.8680 | | | | | | 1.2252 | |
| Proportion of males | -0.4060 | | | | | | | | | | | 1.7558 | | | | | | -0.2312 | | | 0.8171 | | | | | | -3.8474 | | | | | | 3.0354 | |
| **Change in BMI from baseline** **(trial’s follow-up)** | | | | | | | | | | | | | | | | | | | | | | | | | | | | | | | | | | |
| Mixed-Effects Model (k = 70; tau^2 estimator: REML) | | | | | | | | | | | | | | | | | | | | | | | | | | | | | | | | | | |
| logLik | | | | | | | | | deviance | | | | | | AIC | | | | | | | | | BIC | | | | | | AICc | | | | |
| -135.6954 | | | | | | | | | 271.3909 | | | | | | 277.3909 | | | | | | | | | 284.0494 | | | | | | 277.7659 | | | | |
| tau^2 (estimated amount of residual heterogeneity): 2.1682 (SE = 0.5433); tau (square root of estimated tau^2 value): 1.4725;  I^2 (residual heterogeneity / unaccounted variability): 68.44%; H^2 (unaccounted variability / sampling variability): 3.17; R^2 (amount of heterogeneity accounted for): 2.03% | | | | | | | | | | | | | | | | | | | | | | | | | | | | | | | | | | |
| Test for Residual Heterogeneity: QE(df = 68) = 215.4379, p-val < .0001 | | | | | | | | | | | | | | | | | | | | | | | | | | | | | | | | | | |
| Test of Moderators (coefficient 2): QM(df = 1) = 1.9762, p-val = 0.1598 | | | | | | | | | | | | | | | | | | | | | | | | | | | | | | | | | | |
| Model Results: | | | | | | | | | | | | | | | | | | | | | | | | | | | | | | | | | | |
|  | estimate | | | | | | | | | | | se | | | | | | zval | | | pval | | | | | | ci.lb | | | | | | ci.ub | |
| intrcpt | -1.0452 | | | | | | | | | | | 0.4651 | | | | | | -2.2471 | | | 0.0246 | | | | | | -1.9569 | | | | | | -0.1335 | |
| trial’s follow-up | 0.0838 | | | | | | | | | | | 0.0596 | | | | | | 1.4058 | | | 0.1598 | | | | | | -0.0330 | | | | | | 0.2005 | |
| **Change in weight from baseline (mean age at baseline)** | | | | | | | | | | | | | | | | | | | | | | | | | | | | | | | | | | |
| Mixed-Effects Model (k = 44; tau^2 estimator: REML) | | | | | | | | | | | | | | | | | | | | | | | | | | | | | | | | | | |
| logLik | | | | | | | | | deviance | | | | | | AIC | | | | | | | | | BIC | | | | | | AICc | | | | |
| -120.3588 | | | | | | | | | 240.7175 | | | | | | 246.7175 | | | | | | | | | 251.9305 | | | | | | 247.3491 | | | | |
| tau^2 (estimated amount of residual heterogeneity): 17.0563 (SE = 3.9402); tau (square root of estimated tau^2 value): 4.1299;  I^2 (residual heterogeneity / unaccounted variability): 94.46%; H^2 (unaccounted variability / sampling variability): 18.06; R^2 (amount of heterogeneity accounted for): 0.00% | | | | | | | | | | | | | | | | | | | | | | | | | | | | | | | | | | |
| Test for Residual Heterogeneity: QE(df = 42) = 758.3627, p-val < .0001 | | | | | | | | | | | | | | | | | | | | | | | | | | | | | | | | | | |
| Test of Moderators (coefficient 2): QM(df = 1) = 0.0325, p-val = 0.8570 | | | | | | | | | | | | | | | | | | | | | | | | | | | | | | | | | | |
| Model Results: | | | | | | | | | | | | | | | | | | | | | | | | | | | | | | | | | | |
|  | estimate | | | | | | | | | | | se | | | | | | zval | | | pval | | | | | | ci.lb | | | | | | ci.ub | |
| intrcpt | 1.1382 | | | | | | | | | | | 5.9552 | | | | | | 0.1911 | | | 0.8484 | | | | | | -10.5337 | | | | | | 12.8101 | |
| mean age at baseline | -0.0776 | | | | | | | | | | | 0.4304 | | | | | | -0.1802 | | | 0.8570 | | | | | | -0.9211 | | | | | | 0.7660 | |
| **Change in weight from baseline (mean BMI at baseline)** | | | | | | | | | | | | | | | | | | | | | | | | | | | | | | | | | | |
| Mixed-Effects Model (k = 46; tau^2 estimator: REML) | | | | | | | | | | | | | | | | | | | | | | | | | | | | | | | | | | |
| logLik | | | | | | | | | deviance | | | | | | AIC | | | | | | | | | BIC | | | | | | AICc | | | | |
| -124.8506 | | | | | | | | | 249.7013 | | | | | | 255.7013 | | | | | | | | | 261.0538 | | | | | | 256.3013 | | | | |
| tau^2 (estimated amount of residual heterogeneity): 16.0671 (SE = 3.6387); tau (square root of estimated tau^2 value): 4.0084;  I^2 (residual heterogeneity / unaccounted variability): 94.14%; H^2 (unaccounted variability / sampling variability): 17.07; R^2 (amount of heterogeneity accounted for): 0.00% | | | | | | | | | | | | | | | | | | | | | | | | | | | | | | | | | | |
| Test for Residual Heterogeneity: QE(df = 44) = 750.9518, p-val < .0001 | | | | | | | | | | | | | | | | | | | | | | | | | | | | | | | | | | |
| Test of Moderators (coefficient 2): QM(df = 1) = 0.2828, p-val = 0.5949 | | | | | | | | | | | | | | | | | | | | | | | | | | | | | | | | | | |
| Model Results: | | | | | | | | | | | | | | | | | | | | | | | | | | | | | | | | | | |
|  | estimate | | | | | | | | | | | se | | | | | | zval | | | pval | | | | | | ci.lb | | | | | | ci.ub | |
| intrcpt | 2.9454 | | | | | | | | | | | 5.3384 | | | | | | 0.5517 | | | 0.5811 | | | | | | -7.5176 | | | | | | 13.4085 | |
| mean BMI at baseline | -0.0820 | | | | | | | | | | | 0.1542 | | | | | | -0.5318 | | | 0.5949 | | | | | | -0.3842 | | | | | | 0.2202 | |
| **Change in weight from baseline (mean weight at baseline)** | | | | | | | | | | | | | | | | | | | | | | | | | | | | | | | | | | |
| Mixed-Effects Model (k = 44; tau^2 estimator: REML) | | | | | | | | | | | | | | | | | | | | | | | | | | | | | | | | | | |
| logLik | | | | | | | | | deviance | | | | | | AIC | | | | | | | | | BIC | | | | | | AICc | | | | |
| -119.3882 | | | | | | | | | 238.7764 | | | | | | 244.7764 | | | | | | | | | 249.9895 | | | | | | 245.4080 | | | | |
| tau^2 (estimated amount of residual heterogeneity): 16.2408 (SE = 3.7622); tau (square root of estimated tau^2 value): 4.0300  I^2 (residual heterogeneity / unaccounted variability): 94.20%; H^2 (unaccounted variability / sampling variability): 17.24; R^2 (amount of heterogeneity accounted for): 0.00% | | | | | | | | | | | | | | | | | | | | | | | | | | | | | | | | | | |
| Test for Residual Heterogeneity: QE(df = 42) = 724.1120, p-val < .0001 | | | | | | | | | | | | | | | | | | | | | | | | | | | | | | | | | | |
| Test of Moderators (coefficient 2): QM(df = 1) = 0.1169, p-val = 0.7324 | | | | | | | | | | | | | | | | | | | | | | | | | | | | | | | | | | |
| Model Results: | | | | | | | | | | | | | | | | | | | | | | | | | | | | | | | | | | |
|  | | | | | | estimate | | | | | | | se | | | | | | | zval | | | pval | | | | | | ci.lb | | | | | ci.ub |
| intrcpt | | | | | | 1.4964 | | | | | | | 3.6093 | | | | | | | 0.4146 | | | 0.6784 | | | | | | -5.5777 | | | | | 8.5706 |
| mean weight at baseline | | | | | | -0.0133 | | | | | | | 0.0389 | | | | | | | -0.3420 | | | 0.7324 | | | | | | -0.0895 | | | | | 0.0629 |
| **Change in weight from baseline (proportion of males)** | | | | | | | | | | | | | | | | | | | | | | | | | | | | | | | | | | |
| Mixed-Effects Model (k = 46; tau^2 estimator: REML) | | | | | | | | | | | | | | | | | | | | | | | | | | | | | | | | | | |
| logLik | | | | | | | | | | deviance | | | | | | | AIC | | | | | | | | | BIC | | | | | | AICc | | |
| -124.5870 | | | | | | | | | | 249.1741 | | | | | | | 255.1741 | | | | | | | | | 260.5267 | | | | | | 255.7741 | | |
| tau^2 (estimated amount of residual heterogeneity): 15.8638 (SE = 3.5954); tau (square root of estimated tau^2 value): 3.9829  I^2 (residual heterogeneity / unaccounted variability): 94.07%; H^2 (unaccounted variability / sampling variability): 16.86; R^2 (amount of heterogeneity accounted for): 0.00% | | | | | | | | | | | | | | | | | | | | | | | | | | | | | | | | | | |
| Test for Residual Heterogeneity: QE(df = 44) = 742.0080, p-val < .0001 | | | | | | | | | | | | | | | | | | | | | | | | | | | | | | | | | | |
| Test of Moderators (coefficient 2): QM(df = 1) = 0.1497, p-val = 0.6988 | | | | | | | | | | | | | | | | | | | | | | | | | | | | | | | | | | |
| Model Results: | | | | | | | | | | | | | | | | | | | | | | | | | | | | | | | | | | |
|  | | | estimate | | | | | | | | | | se | | | | | | | zval | | | pval | | | | | | ci.lb | | | | | ci.ub |
| intrcpt | | | 1.0221 | | | | | | | | | | 2.2566 | | | | | | | 0.4529 | | | 0.6506 | | | | | | -3.4008 | | | | | 5.4450 |
| proportion of males | | | -1.9853 | | | | | | | | | | 5.1313 | | | | | | | -0.3869 | | | 0.6988 | | | | | | -12.0424 | | | | | 8.0718 |
| **Change in weight from baseline ((trial’s follow-up))** | | | | | | | | | | | | | | | | | | | | | | | | | | | | | | | | | | |
| Mixed-Effects Model (k = 48; tau^2 estimator: REML) | | | | | | | | | | | | | | | | | | | | | | | | | | | | | | | | | | |
| logLik | | | | | | | | | | deviance | | | | | | | AIC | | | | | | | | | BIC | | | | | | AICc | | |
| -126.5743 | | | | | | | | | | 253.1485 | | | | | | | 259.1485 | | | | | | | | | 264.6344 | | | | | | 259.7199 | | |
| tau^2 (estimated amount of residual heterogeneity): 13.3730 (SE = 2.9970); tau (square root of estimated tau^2 value): 3.6569  I^2 (residual heterogeneity / unaccounted variability): 93.04%; H^2 (unaccounted variability / sampling variability): 14.37; R^2 (amount of heterogeneity accounted for): 12.48% | | | | | | | | | | | | | | | | | | | | | | | | | | | | | | | | | | |
| Test for Residual Heterogeneity: QE(df = 46) = 661.1580, p-val < .0001 | | | | | | | | | | | | | | | | | | | | | | | | | | | | | | | | | | |
| Test of Moderators (coefficient 2): QM(df = 1) = 7.2361, p-val = 0.0071 | | | | | | | | | | | | | | | | | | | | | | | | | | | | | | | | | | |
| Model Results: | | | | | | | | | | | | | | | | | | | | | | | | | | | | | | | | | | |
|  | | | estimate | | | | | | | | | | se | | | | | | | zval | | | pval | | | | | | ci.lb | | | | | ci.ub |
| intrcpt | | | -2.3577 | | | | | | | | | | 1.0481 | | | | | | | -2.2494 | | | 0.0245 | | | | | | -4.4121 | | | | | -0.3034 |
| trial’s follow-up | | | 0.3456 | | | | | | | | | | 0.1285 | | | | | | | 2.6900 | | | 0.0071 | | | | | | 0.0938 | | | | | 0.5975 |
| **Percentage of participants achieving BMI reduction of at least 5% (mean age at baseline)** | | | | | | | | | | | | | | | | | | | | | | | | | | | | | | | | | | |
| Mixed-Effects Model (k = 12; tau^2 estimator: REML) | | | | | | | | | | | | | | | | | | | | | | | | | | | | | | | | | | |
| logLik | | | | | | | | | | deviance | | | | | | | AIC | | | | | | | | | BIC | | | | | | AICc | | |
| -11.5932 | | | | | | | | | | 23.1864 | | | | | | | 29.1864 | | | | | | | | | 30.0942 | | | | | | 33.1864 | | |
| tau^2 (estimated amount of residual heterogeneity): 0.5626 (SE = 0.2643); tau (square root of estimated tau^2 value): 0.7501  I^2 (residual heterogeneity / unaccounted variability): 97.50%; H^2 (unaccounted variability / sampling variability): 39.92; R^2 (amount of heterogeneity accounted for): 1.99% | | | | | | | | | | | | | | | | | | | | | | | | | | | | | | | | | | |
| Test for Residual Heterogeneity: QE(df = 10) = 221.0569, p-val < .0001 | | | | | | | | | | | | | | | | | | | | | | | | | | | | | | | | | | |
| Test of Moderators (coefficient 2): QM(df = 1) = 1.1438, p-val = 0.2849 | | | | | | | | | | | | | | | | | | | | | | | | | | | | | | | | | | |
| Model Results: | | | | | | | | | | | | | | | | | | | | | | | | | | | | | | | | | | |
|  | | | estimate | | | | | | | | | | se | | | | | | | zval | | | pval | | | | | | ci.lb | | | | | ci.ub |
| intrcpt | | | -2.2114 | | | | | | | | | | 2.5886 | | | | | | | -0.8543 | | | 0.3929 | | | | | | -7.2849 | | | | | 2.8621 |
| mean age at baseline | | | 0.1959 | | | | | | | | | | 0.1831 | | | | | | | 1.0695 | | | 0.2849 | | | | | | -0.1631 | | | | | 0.5548 |
| **Percentage of participants achieving BMI reduction of at least 5% (mean BMI at baseline)** | | | | | | | | | | | | | | | | | | | | | | | | | | | | | | | | | | |
| Mixed-Effects Model (k = 12; tau^2 estimator: REML) | | | | | | | | | | | | | | | | | | | | | | | | | | | | | | | | | | |
| logLik | | | | | | | | | | deviance | | | | | | | AIC | | | | | | | | | BIC | | | | | | AICc | | |
| -12.1239 | | | | | | | | | | 24.2479 | | | | | | | 30.2479 | | | | | | | | | 31.1556 | | | | | | 34.2479 | | |
| tau^2 (estimated amount of residual heterogeneity): 0.6316 (SE = 0.2947); tau (square root of estimated tau^2 value): 0.7947;  I^2 (residual heterogeneity / unaccounted variability): 97.73%; H^2 (unaccounted variability / sampling variability): 43.99; R^2 (amount of heterogeneity accounted for): 0.00% | | | | | | | | | | | | | | | | | | | | | | | | | | | | | | | | | | |
| Test for Residual Heterogeneity: QE(df = 10) = 242.5093, p-val < .0001 | | | | | | | | | | | | | | | | | | | | | | | | | | | | | | | | | | |
| Test of Moderators (coefficient 2): QM(df = 1) = 0.0002, p-val = 0.9878 | | | | | | | | | | | | | | | | | | | | | | | | | | | | | | | | | | |
| Model Results: | | | | | | | | | | | | | | | | | | | | | | | | | | | | | | | | | | |
|  | | | estimate | | | | | | | | | | se | | | | | | | zval | | | pval | | | | | | ci.lb | | | | | ci.ub |
| intrcpt | | | 0.4701 | | | | | | | | | | 4.9880 | | | | | | | 0.0943 | | | 0.9249 | | | | | | -9.3062 | | | | | 10.2464 |
| mean BMI at baseline | | | 0.0021 | | | | | | | | | | 0.1351 | | | | | | | 0.0153 | | | 0.9878 | | | | | | -0.2628 | | | | | 0.2669 |
| **Percentage of participants achieving BMI reduction of at least 5% (mean weight at baseline)** | | | | | | | | | | | | | | | | | | | | | | | | | | | | | | | | | | |
| Mixed-Effects Model (k = 10; tau^2 estimator: REML) | | | | | | | | | | | | | | | | | | | | | | | | | | | | | | | | | | |
| logLik | | | | | | | | | | deviance | | | | | | | AIC | | | | | | | | | BIC | | | | | | AICc | | |
| -10.4285 | | | | | | | | | | 20.8570 | | | | | | | 26.8570 | | | | | | | | | 27.0953 | | | | | | 32.8570 | | |
| tau^2 (estimated amount of residual heterogeneity): 0.7623 (SE = 0.3935); tau (square root of estimated tau^2 value): 0.8731  I^2 (residual heterogeneity / unaccounted variability): 98.28%; H^2 (unaccounted variability / sampling variability): 57.98; R^2 (amount of heterogeneity accounted for): 0.00% | | | | | | | | | | | | | | | | | | | | | | | | | | | | | | | | | | |
| Test for Residual Heterogeneity: QE(df = 8) = 223.7114, p-val < .0001 | | | | | | | | | | | | | | | | | | | | | | | | | | | | | | | | | | |
| Test of Moderators (coefficient 2): QM(df = 1) = 0.0877, p-val = 0.7671 | | | | | | | | | | | | | | | | | | | | | | | | | | | | | | | | | | |
| Model Results: | | | | | | | | | | | | | | | | | | | | | | | | | | | | | | | | | | |
|  | | | estimate | | | | | | | | | | se | | | | | | | zval | | | pval | | | | | | ci.lb | | | | | ci.ub |
| intrcpt | | | -0.8510 | | | | | | | | | | 4.8944 | | | | | | | -0.1739 | | | 0.8620 | | | | | | -10.4438 | | | | | 8.7419 |
| mean weight at baseline | | | 0.0138 | | | | | | | | | | 0.0465 | | | | | | | 0.2962 | | | 0.7671 | | | | | | -0.0774 | | | | | 0.1050 |
| **Percentage of participants achieving BMI reduction of at least 5% (proportion of males)** | | | | | | | | | | | | | | | | | | | | | | | | | | | | | | | | | | |
| Mixed-Effects Model (k = 12; tau^2 estimator: REML) | | | | | | | | | | | | | | | | | | | | | | | | | | | | | | | | | | |
| logLik | | | | | | | | | | deviance | | | | | | | AIC | | | | | | | | | BIC | | | | | | AICc | | |
| -12.0948 | | | | | | | | | | 24.1896 | | | | | | | 30.1896 | | | | | | | | | 31.0973 | | | | | | 34.1896 | | |
| tau^2 (estimated amount of residual heterogeneity): 0.6282 (SE = 0.2944); tau (square root of estimated tau^2 value): 0.7926  I^2 (residual heterogeneity / unaccounted variability): 97.53%; H^2 (unaccounted variability / sampling variability): 40.42; R^2 (amount of heterogeneity accounted for): 0.00% | | | | | | | | | | | | | | | | | | | | | | | | | | | | | | | | | | |
| Test for Residual Heterogeneity: QE(df = 10) = 249.7422, p-val < .0001 | | | | | | | | | | | | | | | | | | | | | | | | | | | | | | | | | | |
| Test of Moderators (coefficient 2): QM(df = 1) = 0.1009, p-val = 0.7508 | | | | | | | | | | | | | | | | | | | | | | | | | | | | | | | | | | |
| Model Results: | | | | | | | | | | | | | | | | | | | | | | | | | | | | | | | | | | |
|  | | | | estimate | | | | | | | | | se | | | | | | | zval | | | pval | | | | | | ci.lb | | | | | ci.ub |
| intrcpt | | | | 1.1005 | | | | | | | | | 1.7598 | | | | | | | 0.6254 | | | 0.5317 | | | | | | -2.3486 | | | | | 4.5496 |
| proportion of males | | | | -1.3698 | | | | | | | | | 4.3128 | | | | | | | -0.3176 | | | 0.7508 | | | | | | -9.8228 | | | | | 7.0832 |
| **Percentage of participants achieving BMI reduction of at least 5% (trial’s follow-up)** | | | | | | | | | | | | | | | | | | | | | | | | | | | | | | | | | | |
| Mixed-Effects Model (k = 12; tau^2 estimator: REML) | | | | | | | | | | | | | | | | | | | | | | | | | | | | | | | | | | |
| logLik | | | | | | | | | | deviance | | | | | | | AIC | | | | | | | | | BIC | | | | | | AICc | | |
| -11.2321 | | | | | | | | | | 22.4641 | | | | | | | 28.4641 | | | | | | | | | 29.3719 | | | | | | 32.4641 | | |
| tau^2 (estimated amount of residual heterogeneity): 0.5240 (SE = 0.2463); tau (square root of estimated tau^2 value): 0.7239  I^2 (residual heterogeneity / unaccounted variability): 97.40%; H^2 (unaccounted variability / sampling variability): 38.39; R^2 (amount of heterogeneity accounted for): 8.71% | | | | | | | | | | | | | | | | | | | | | | | | | | | | | | | | | | |
| Test for Residual Heterogeneity: QE(df = 10) = 211.7179, p-val < .0001 | | | | | | | | | | | | | | | | | | | | | | | | | | | | | | | | | | |
| Test of Moderators (coefficient 2): QM(df = 1) = 1.9431, p-val = 0.1633 | | | | | | | | | | | | | | | | | | | | | | | | | | | | | | | | | | |
| Model Results: | | | | | | | | | | | | | | | | | | | | | | | | | | | | | | | | | | |
|  | | | | estimate | | | | | | | | | se | | | | | | | zval | | | pval | | | | | | ci.lb | | | | | ci.ub |
| intrcpt | | | | -0.6511 | | | | | | | | | 0.8859 | | | | | | | -0.7349 | | | 0.4624 | | | | | | -2.3875 | | | | | 1.0853 |
| trial’s follow-up | | | | 0.0925 | | | | | | | | | 0.0664 | | | | | | | 1.3940 | | | 0.1633 | | | | | | -0.0376 | | | | | 0.2225 |
| **Percentage of participants achieving BMI reduction of at least 10% (mean age at baseline)** | | | | | | | | | | | | | | | | | | | | | | | | | | | | | | | | | | |
| Mixed-Effects Model (k = 12; tau^2 estimator: REML) | | | | | | | | | | | | | | | | | | | | | | | | | | | | | | | | | | |
| logLik | | | | | | | | | | deviance | | | | | | | AIC | | | | | | | | | BIC | | | | | | AICc | | |
| -6.3398 | | | | | | | | | | 12.6797 | | | | | | | 18.6797 | | | | | | | | | 19.5874 | | | | | | 22.6797 | | |
| tau^2 (estimated amount of residual heterogeneity): 0.1848 (SE = 0.0927); tau (square root of estimated tau^2 value): 0.4298  I^2 (residual heterogeneity / unaccounted variability): 93.68%; H^2 (unaccounted variability / sampling variability): 15.82; R^2 (amount of heterogeneity accounted for): 2.61% | | | | | | | | | | | | | | | | | | | | | | | | | | | | | | | | | | |
| Test for Residual Heterogeneity: QE(df = 10) = 110.6663, p-val < .0001 | | | | | | | | | | | | | | | | | | | | | | | | | | | | | | | | | | |
| Test of Moderators (coefficient 2): QM(df = 1) = 1.0684, p-val = 0.3013 | | | | | | | | | | | | | | | | | | | | | | | | | | | | | | | | | | |
| Model Results: | | | | | | | | | | | | | | | | | | | | | | | | | | | | | | | | | | |
|  | | | | estimate | | | | | | | | | se | | | | | | | zval | | | pval | | | | | | ci.lb | | | | | ci.ub |
| intrcpt | | | | -1.3310 | | | | | | | | | 1.5722 | | | | | | | -0.8466 | | | 0.3972 | | | | | | -4.4124 | | | | | 1.7504 |
| mean age at baseline | | | | 0.1150 | | | | | | | | | 0.1112 | | | | | | | 1.0336 | | | 0.3013 | | | | | | -0.1030 | | | | | 0.3330 |
| **Percentage of participants achieving BMI reduction of at least 10% (mean BMI at baseline)** | | | | | | | | | | | | | | | | | | | | | | | | | | | | | | | | | | |
| Mixed-Effects Model (k = 12; tau^2 estimator: REML) | | | | | | | | | | | | | | | | | | | | | | | | | | | | | | | | | | |
| logLik | | | | | | | | | | deviance | | | | | | | AIC | | | | | | | | | BIC | | | | | | AICc | | |
| -6.8176 | | | | | | | | | | 13.6353 | | | | | | | 19.6353 | | | | | | | | | 20.5430 | | | | | | 23.6353 | | |
| tau^2 (estimated amount of residual heterogeneity): 0.2077 (SE = 0.1028); tau (square root of estimated tau^2 value): 0.4558  I^2 (residual heterogeneity / unaccounted variability): 94.28%; H^2 (unaccounted variability / sampling variability): 17.49; R^2 (amount of heterogeneity accounted for): 0.00% | | | | | | | | | | | | | | | | | | | | | | | | | | | | | | | | | | |
| Test for Residual Heterogeneity: QE(df = 10) = 118.8396, p-val < .0001 | | | | | | | | | | | | | | | | | | | | | | | | | | | | | | | | | | |
| Test of Moderators (coefficient 2): QM(df = 1) = 0.0092, p-val = 0.9235 | | | | | | | | | | | | | | | | | | | | | | | | | | | | | | | | | | |
| Model Results: | | | | | | | | | | | | | | | | | | | | | | | | | | | | | | | | | | |
|  | | | | estimate | | | | | | | | | se | | | | | | | zval | | | pval | | | | | | ci.lb | | | | | ci.ub |
| intrcpt | | | | -0.0041 | | | | | | | | | 3.0430 | | | | | | | -0.0013 | | | 0.9989 | | | | | | -5.9683 | | | | | 5.9601 |
| mean BMI at baseline | | | | 0.0079 | | | | | | | | | 0.0826 | | | | | | | 0.0960 | | | 0.9235 | | | | | | -0.1539 | | | | | 0.1698 |
| **Percentage of participants achieving BMI reduction of at least 10% (mean weight at baseline)** | | | | | | | | | | | | | | | | | | | | | | | | | | | | | | | | | | |
| Mixed-Effects Model (k = 10; tau^2 estimator: REML) | | | | | | | | | | | | | | | | | | | | | | | | | | | | | | | | | | |
| logLik | | | | | | | | | | deviance | | | | | | | AIC | | | | | | | | | BIC | | | | | | AICc | | |
| -6.1346 | | | | | | | | | | 12.2691 | | | | | | | 18.2691 | | | | | | | | | 18.5075 | | | | | | 24.2691 | | |
| tau^2 (estimated amount of residual heterogeneity): 0.2484 (SE = 0.1341); tau (square root of estimated tau^2 value): 0.4984  I^2 (residual heterogeneity / unaccounted variability): 95.57%; H^2 (unaccounted variability / sampling variability): 22.58; R^2 (amount of heterogeneity accounted for): 0.00% | | | | | | | | | | | | | | | | | | | | | | | | | | | | | | | | | | |
| Test for Residual Heterogeneity: QE(df = 8) = 106.2560, p-val < .0001 | | | | | | | | | | | | | | | | | | | | | | | | | | | | | | | | | | |
| Test of Moderators (coefficient 2): QM(df = 1) = 0.1203, p-val = 0.7287 | | | | | | | | | | | | | | | | | | | | | | | | | | | | | | | | | | |
| Model Results: | | | | | | | | | | | | | | | | | | | | | | | | | | | | | | | | | | |
|  | | | | estimate | | | | | | | | | se | | | | | | | zval | | | pval | | | | | | ci.lb | | | | | ci.ub |
| intrcpt | | | | -0.6870 | | | | | | | | | 2.9004 | | | | | | | -0.2369 | | | 0.8128 | | | | | | -6.3718 | | | | | 4.9977 |
| mean weight at baseline | | | | 0.0096 | | | | | | | | | 0.0277 | | | | | | | 0.3468 | | | 0.7287 | | | | | | -0.0446 | | | | | 0.0638 |
| **Percentage of participants achieving BMI reduction of at least 10% (proportion of males)** | | | | | | | | | | | | | | | | | | | | | | | | | | | | | | | | | | |
| Mixed-Effects Model (k = 12; tau^2 estimator: REML) | | | | | | | | | | | | | | | | | | | | | | | | | | | | | | | | | | |
| logLik | | | | | | | | | | deviance | | | | | | | AIC | | | | | | | | | BIC | | | | | | AICc | | |
| -6.8663 | | | | | | | | | | 13.7327 | | | | | | | 19.7327 | | | | | | | | | 20.6404 | | | | | | 23.7327 | | |
| tau^2 (estimated amount of residual heterogeneity): 0.2103 (SE = 0.1049); tau (square root of estimated tau^2 value): 0.4586  I^2 (residual heterogeneity / unaccounted variability): 93.90%; H^2 (unaccounted variability / sampling variability): 16.39; R^2 (amount of heterogeneity accounted for): 0.00% | | | | | | | | | | | | | | | | | | | | | | | | | | | | | | | | | | |
| Test for Residual Heterogeneity: QE(df = 10) = 124.4733, p-val < .0001 | | | | | | | | | | | | | | | | | | | | | | | | | | | | | | | | | | |
| Test of Moderators (coefficient 2): QM(df = 1) = 0.0090, p-val = 0.9242 | | | | | | | | | | | | | | | | | | | | | | | | | | | | | | | | | | |
| Model Results: | | | | | | | | | | | | | | | | | | | | | | | | | | | | | | | | | | |
|  | | | | estimate | | | | | | | | | se | | | | | | | zval | | | pval | | | | | | ci.lb | | | | | ci.ub |
| intrcpt | | | | 0.3868 | | | | | | | | | 1.0526 | | | | | | | 0.3675 | | | 0.7132 | | | | | | -1.6762 | | | | | 2.4498 |
| proportion of males | | | | -0.2458 | | | | | | | | | 2.5837 | | | | | | | -0.0951 | | | 0.9242 | | | | | | -5.3098 | | | | | 4.8182 |
| **Percentage of participants achieving BMI reduction of at least 10% (trial’s follow-up)** | | | | | | | | | | | | | | | | | | | | | | | | | | | | | | | | | | |
| Mixed-Effects Model (k = 12; tau^2 estimator: REML) | | | | | | | | | | | | | | | | | | | | | | | | | | | | | | | | | | |
| logLik | | | | | | | | | | deviance | | | | | | | AIC | | | | | | | | | BIC | | | | | | AICc | | |
| -5.8219 | | | | | | | | | | 11.6438 | | | | | | | 17.6438 | | | | | | | | | 18.5515 | | | | | | 21.6438 | | |
| tau^2 (estimated amount of residual heterogeneity): 0.1670 (SE = 0.0842); tau (square root of estimated tau^2 value): 0.4086  I^2 (residual heterogeneity / unaccounted variability): 93.25%; H^2 (unaccounted variability / sampling variability): 14.81; R^2 (amount of heterogeneity accounted for): 11.99% | | | | | | | | | | | | | | | | | | | | | | | | | | | | | | | | | | |
| Test for Residual Heterogeneity: QE(df = 10) = 103.8869, p-val < .0001 | | | | | | | | | | | | | | | | | | | | | | | | | | | | | | | | | | |
| Test of Moderators (coefficient 2): QM(df = 1) = 2.1955, p-val = 0.1384 | | | | | | | | | | | | | | | | | | | | | | | | | | | | | | | | | | |
| Model Results: | | | | | | | | | | | | | | | | | | | | | | | | | | | | | | | | | | |
|  | | | | estimate | | | | | | | | | se | | | | | | | zval | | | pval | | | | | | ci.lb | | | | | ci.ub |
| intrcpt | | | | -0.5008 | | | | | | | | | 0.5477 | | | | | | | -0.9144 | | | 0.3605 | | | | | | -1.5743 | | | | | 0.5727 |
| trial’s follow-up | | | | 0.0602 | | | | | | | | | 0.0406 | | | | | | | 1.4817 | | | 0.1384 | | | | | | -0.0194 | | | | | 0.1399 |
| **Change in BMI z-score from baseline (mean age at baseline)** | | | | | | | | | | | | | | | | | | | | | | | | | | | | | | | | | | |
| Mixed-Effects Model (k = 8; tau^2 estimator: REML) | | | | | | | | | | | | | | | | | | | | | | | | | | | | | | | | | | |
| logLik | | | | | | | | | | deviance | | | | | | | AIC | | | | | | | | | BIC | | | | | | AICc | | |
| -5.5837 | | | | | | | | | | 11.1673 | | | | | | | 17.1673 | | | | | | | | | 16.5426 | | | | | | 29.1673 | | |
| tau^2 (estimated amount of residual heterogeneity): 0 (SE = 0.5774); tau (square root of estimated tau^2 value): 0  I^2 (residual heterogeneity / unaccounted variability): 0.00%; H^2 (unaccounted variability / sampling variability): 1.00; R^2 (amount of heterogeneity accounted for): 0.00% | | | | | | | | | | | | | | | | | | | | | | | | | | | | | | | | | | |
| Test for Residual Heterogeneity: QE(df = 6) = 0.1401, p-val = 0.9999 | | | | | | | | | | | | | | | | | | | | | | | | | | | | | | | | | | |
| Test of Moderators (coefficient 2): QM(df = 1) = 0.0003, p-val = 0.9861 | | | | | | | | | | | | | | | | | | | | | | | | | | | | | | | | | | |
| Model Results: | | | | | | | | | | | | | | | | | | | | | | | | | | | | | | | | | | |
|  | | | | estimate | | | | | | | | | se | | | | | | | zval | | | pval | | | | | | ci.lb | | | | | ci.ub |
| intrcpt | | | | -0.1190 | | | | | | | | | 3.1809 | | | | | | | -0.0374 | | | 0.9702 | | | | | | -6.3535 | | | | | 6.1155 |
| mean age at baseline | | | | 0.0040 | | | | | | | | | 0.2309 | | | | | | | 0.0175 | | | 0.9861 | | | | | | -0.4485 | | | | | 0.4566 |
| **Change in BMI z-score from baseline (mean BMI at baseline)** | | | | | | | | | | | | | | | | | | | | | | | | | | | | | | | | | | |
| Mixed-Effects Model (k = 10; tau^2 estimator: REML) | | | | | | | | | | | | | | | | | | | | | | | | | | | | | | | | | | |
| logLik | | | | | | | | | | deviance | | | | | | | AIC | | | | | | | | | BIC | | | | | | AICc | | |
| -7.4513 | | | | | | | | | | 14.9026 | | | | | | | 20.9026 | | | | | | | | | 21.1409 | | | | | | 26.9026 | | |
| tau^2 (estimated amount of residual heterogeneity): 0 (SE = 0.5000); tau (square root of estimated tau^2 value): 0;  I^2 (residual heterogeneity / unaccounted variability): 0.00%; H^2 (unaccounted variability / sampling variability): 1.00; R^2 (amount of heterogeneity accounted for): 0.00% | | | | | | | | | | | | | | | | | | | | | | | | | | | | | | | | | | |
| Test for Residual Heterogeneity: QE(df = 8) = 0.1996, p-val = 1.0000 | | | | | | | | | | | | | | | | | | | | | | | | | | | | | | | | | | |
| Test of Moderators (coefficient 2): QM(df = 1) = 0.1485, p-val = 0.7000 | | | | | | | | | | | | | | | | | | | | | | | | | | | | | | | | | | |
| Model Results: | | | | | | | | | | | | | | | | | | | | | | | | | | | | | | | | | | |
|  | | | | estimate | | | | | | | | | se | | | | | | | zval | | | pval | | | | | | ci.lb | | | | | ci.ub |
| intrcpt | | | | -1.2714 | | | | | | | | | 2.9246 | | | | | | | -0.4347 | | | 0.6638 | | | | | | -7.0034 | | | | | 4.4607 |
| mean BMI at baseline | | | | 0.0325 | | | | | | | | | 0.0844 | | | | | | | 0.3853 | | | 0.7000 | | | | | | -0.1329 | | | | | 0.1979 |
| **Change in BMI z-score from baseline (mean weight at baseline)** | | | | | | | | | | | | | | | | | | | | | | | | | | | | | | | | | | |
| Mixed-Effects Model (k = 8; tau^2 estimator: REML) | | | | | | | | | | | | | | | | | | | | | | | | | | | | | | | | | | |
| logLik | | | | | | | | | | deviance | | | | | | | AIC | | | | | | | | | BIC | | | | | | AICc | | |
| -5.5822 | | | | | | | | | | 11.1645 | | | | | | | 17.1645 | | | | | | | | | 16.5397 | | | | | | 29.1645 | | |
| tau^2 (estimated amount of residual heterogeneity): 0 (SE = 0.5774); tau (square root of estimated tau^2 value): 0;  I^2 (residual heterogeneity / unaccounted variability): 0.00%; H^2 (unaccounted variability / sampling variability): 1.00; R^2 (amount of heterogeneity accounted for): 0.00% | | | | | | | | | | | | | | | | | | | | | | | | | | | | | | | | | | |
| Test for Residual Heterogeneity: QE(df = 6) = 0.1372, p-val = 0.9999 | | | | | | | | | | | | | | | | | | | | | | | | | | | | | | | | | | |
| Test of Moderators (coefficient 2): QM(df = 1) = 0.1918, p-val = 0.6614 | | | | | | | | | | | | | | | | | | | | | | | | | | | | | | | | | | |
| Model Results: | | | | | | | | | | | | | | | | | | | | | | | | | | | | | | | | | | |
|  | | | | estimate | | | | | | | | | se | | | | | | | zval | | | pval | | | | | | ci.lb | | | | | ci.ub |
| intrcpt | | | | -0.9970 | | | | | | | | | 1.9296 | | | | | | | -0.5167 | | | 0.6054 | | | | | | -4.7788 | | | | | 2.7849 |
| mean weight at baseline | | | | 0.0091 | | | | | | | | | 0.0207 | | | | | | | 0.4379 | | | 0.6614 | | | | | | -0.0315 | | | | | 0.0496 |
| **Change in BMI z-score from baseline (proportion of males)** | | | | | | | | | | | | | | | | | | | | | | | | | | | | | | | | | | |
| Mixed-Effects Model (k = 12; tau^2 estimator: REML) | | | | | | | | | | | | | | | | | | | | | | | | | | | | | | | | | | |
| logLik | | | | | | | | | | deviance | | | | | | | AIC | | | | | | | | | BIC | | | | | | AICc | | |
| -9.4307 | | | | | | | | | | 18.8614 | | | | | | | 24.8614 | | | | | | | | | 25.7692 | | | | | | 28.8614 | | |
| tau^2 (estimated amount of residual heterogeneity): 0 (SE = 0.4472); tau (square root of estimated tau^2 value): 0;  I^2 (residual heterogeneity / unaccounted variability): 0.00%; H^2 (unaccounted variability / sampling variability): 1.00; R^2 (amount of heterogeneity accounted for): 0.00% | | | | | | | | | | | | | | | | | | | | | | | | | | | | | | | | | | |
| Test for Residual Heterogeneity: QE(df = 10) = 0.4827, p-val = 1.0000 | | | | | | | | | | | | | | | | | | | | | | | | | | | | | | | | | | |
| Test of Moderators (coefficient 2): QM(df = 1) = 0.0042, p-val = 0.9481 | | | | | | | | | | | | | | | | | | | | | | | | | | | | | | | | | | |
| Model Results: | | | | | | | | | | | | | | | | | | | | | | | | | | | | | | | | | | |
|  | | | | estimate | | | | | | | | | se | | | | | | | zval | | | pval | | | | | | ci.lb | | | | | ci.ub |
| intrcpt | | | | -0.2166 | | | | | | | | | 1.2739 | | | | | | | -0.1700 | | | 0.8650 | | | | | | -2.7133 | | | | | 2.2801 |
| proportion of males | | | | 0.1532 | | | | | | | | | 2.3528 | | | | | | | 0.0651 | | | 0.9481 | | | | | | -4.4583 | | | | | 4.7646 |
| **Change in BMI z-score from baseline (trial’s follow-up)** | | | | | | | | | | | | | | | | | | | | | | | | | | | | | | | | | | |
| Mixed-Effects Model (k = 12; tau^2 estimator: REML) | | | | | | | | | | | | | | | | | | | | | | | | | | | | | | | | | | |
| logLik | | | | | | | | | | deviance | | | | | | | AIC | | | | | | | | | BIC | | | | | | AICc | | |
| -9.4245 | | | | | | | | | | 18.8490 | | | | | | | 24.8490 | | | | | | | | | 25.7567 | | | | | | 28.8490 | | |
| tau^2 (estimated amount of residual heterogeneity): 0 (SE = 0.4472); tau (square root of estimated tau^2 value): 0;  I^2 (residual heterogeneity / unaccounted variability): 0.00%; H^2 (unaccounted variability / sampling variability): 1.00; R^2 (amount of heterogeneity accounted for): 0.00% | | | | | | | | | | | | | | | | | | | | | | | | | | | | | | | | | | |
| Test for Residual Heterogeneity: QE(df = 10) = 0.4702, p-val = 1.0000 | | | | | | | | | | | | | | | | | | | | | | | | | | | | | | | | | | |
| Test of Moderators (coefficient 2): QM(df = 1) = 0.0167, p-val = 0.8973 | | | | | | | | | | | | | | | | | | | | | | | | | | | | | | | | | | |
| Model Results: | | | | | | | | | | | | | | | | | | | | | | | | | | | | | | | | | | |
|  | | | | estimate | | | | | | | | | se | | | | | | | zval | | | pval | | | | | | ci.lb | | | | | ci.ub |
| intrcpt | | | | -0.1793 | | | | | | | | | 0.4432 | | | | | | | -0.4044 | | | 0.6859 | | | | | | -1.0480 | | | | | 0.6894 |
| trial’s follow-up | | | | 0.0043 | | | | | | | | | 0.0329 | | | | | | | 0.1291 | | | 0.8973 | | | | | | -0.0603 | | | | | 0.0688 |
| **Change in BMI SDS from baseline (mean age at baseline)** | | | | | | | | | | | | | | | | | | | | | | | | | | | | | | | | | | |
| Mixed-Effects Model (k = 20; tau^2 estimator: REML) | | | | | | | | | | | | | | | | | | | | | | | | | | | | | | | | | | |
| logLik | | | | | | | | | | deviance | | | | | | | AIC | | | | | | | | | BIC | | | | | | AICc | | |
| -16.8290 | | | | | | | | | | 33.6580 | | | | | | | 39.6580 | | | | | | | | | 42.3291 | | | | | | 41.3723 | | |
| tau^2 (estimated amount of residual heterogeneity): 0 (SE = 0.3333); tau (square root of estimated tau^2 value): 0;  I^2 (residual heterogeneity / unaccounted variability): 0.00%; H^2 (unaccounted variability / sampling variability): 1.00; R^2 (amount of heterogeneity accounted for): 0.00% | | | | | | | | | | | | | | | | | | | | | | | | | | | | | | | | | | |
| Test for Residual Heterogeneity: QE(df = 18) = 0.5762, p-val = 1.0000 | | | | | | | | | | | | | | | | | | | | | | | | | | | | | | | | | | |
| Test of Moderators (coefficient 2): QM(df = 1) = 0.1201, p-val = 0.7290 | | | | | | | | | | | | | | | | | | | | | | | | | | | | | | | | | | |
| Model Results: | | | | | | | | | | | | | | | | | | | | | | | | | | | | | | | | | | |
|  | | | | estimate | | | | | | | | | se | | | | | | | zval | | | pval | | | | | | ci.lb | | | | | ci.ub |
| intrcpt | | | | -0.6806 | | | | | | | | | 1.5814 | | | | | | | -0.4304 | | | 0.6669 | | | | | | -3.7801 | | | | | 2.4190 |
| mean age at baseline | | | | 0.0422 | | | | | | | | | 0.1218 | | | | | | | 0.3465 | | | 0.7290 | | | | | | -0.1965 | | | | | 0.2809 |
| **Change in BMI SDS from baseline (mean BMI at baseline)** | | | | | | | | | | | | | | | | | | | | | | | | | | | | | | | | | | |
| Mixed-Effects Model (k = 16; tau^2 estimator: REML) | | | | | | | | | | | | | | | | | | | | | | | | | | | | | | | | | | |
| logLik | | | | | | | | | | deviance | | | | | | | AIC | | | | | | | | | BIC | | | | | | AICc | | |
| -13.0202 | | | | | | | | | | 26.0404 | | | | | | | 32.0404 | | | | | | | | | 33.9576 | | | | | | 34.4404 | | |
| tau^2 (estimated amount of residual heterogeneity): 0 (SE = 0.3780); tau (square root of estimated tau^2 value): 0;  I^2 (residual heterogeneity / unaccounted variability): 0.00%; H^2 (unaccounted variability / sampling variability): 1.00; R^2 (amount of heterogeneity accounted for): 0.00% | | | | | | | | | | | | | | | | | | | | | | | | | | | | | | | | | | |
| Test for Residual Heterogeneity: QE(df = 14) = 0.3101, p-val = 1.0000 | | | | | | | | | | | | | | | | | | | | | | | | | | | | | | | | | | |
| Test of Moderators (coefficient 2): QM(df = 1) = 0.0426, p-val = 0.8365 | | | | | | | | | | | | | | | | | | | | | | | | | | | | | | | | | | |
| Model Results: | | | | | | | | | | | | | | | | | | | | | | | | | | | | | | | | | | |
|  | | | | estimate | | | | | | | | | se | | | | | | | zval | | | pval | | | | | | ci.lb | | | | | ci.ub |
| intrcpt | | | | -0.7671 | | | | | | | | | 3.0247 | | | | | | | -0.2536 | | | 0.7998 | | | | | | -6.6953 | | | | | 5.1612 |
| mean BMI at baseline | | | | 0.0183 | | | | | | | | | 0.0887 | | | | | | | 0.2063 | | | 0.8365 | | | | | | -0.1556 | | | | | 0.1922 |
| **Change in BMI SDS from baseline (mean weight at baseline)** | | | | | | | | | | | | | | | | | | | | | | | | | | | | | | | | | | |
| Mixed-Effects Model (k = 12; tau^2 estimator: REML) | | | | | | | | | | | | | | | | | | | | | | | | | | | | | | | | | | |
| logLik | | | | | | | | | | deviance | | | | | | | AIC | | | | | | | | | BIC | | | | | | AICc | | |
| -9.2333 | | | | | | | | | | 18.4666 | | | | | | | 24.4666 | | | | | | | | | 25.3744 | | | | | | 28.4666 | | |
| tau^2 (estimated amount of residual heterogeneity): 0 (SE = 0.4472); tau (square root of estimated tau^2 value): 0;  I^2 (residual heterogeneity / unaccounted variability): 0.00%; H^2 (unaccounted variability / sampling variability): 1.00; R^2 (amount of heterogeneity accounted for): 0.00% | | | | | | | | | | | | | | | | | | | | | | | | | | | | | | | | | | |
| Test for Residual Heterogeneity: QE(df = 10) = 0.0878, p-val = 1.0000 | | | | | | | | | | | | | | | | | | | | | | | | | | | | | | | | | | |
| Test of Moderators (coefficient 2): QM(df = 1) = 0.0518, p-val = 0.8199 | | | | | | | | | | | | | | | | | | | | | | | | | | | | | | | | | | |
| Model Results: | | | | | | | | | | | | | | | | | | | | | | | | | | | | | | | | | | |
|  | | | | estimate | | | | | | | | | se | | | | | | | zval | | | pval | | | | | | ci.lb | | | | | ci.ub |
| intrcpt | | | | -0.5217 | | | | | | | | | 1.7983 | | | | | | | -0.2901 | | | 0.7717 | | | | | | -4.0463 | | | | | 3.0029 |
| mean weight at baseline | | | | 0.0046 | | | | | | | | | 0.0201 | | | | | | | 0.2276 | | | 0.8199 | | | | | | -0.0348 | | | | | 0.0439 |
| **Change in BMI SDS from baseline (****proportion of males)** | | | | | | | | | | | | | | | | | | | | | | | | | | | | | | | | | | |
| Mixed-Effects Model (k = 20; tau^2 estimator: REML) | | | | | | | | | | | | | | | | | | | | | | | | | | | | | | | | | | |
| logLik | | | | | | | | | | deviance | | | | | | | AIC | | | | | | | | | BIC | | | | | | AICc | | |
| -16.8035 | | | | | | | | | | 33.6070 | | | | | | | 39.6070 | | | | | | | | | 42.2781 | | | | | | 41.3213 | | |
| tau^2 (estimated amount of residual heterogeneity): 0 (SE = 0.3333); tau (square root of estimated tau^2 value): 0;  I^2 (residual heterogeneity / unaccounted variability): 0.00%; H^2 (unaccounted variability / sampling variability): 1.00; R^2 (amount of heterogeneity accounted for): 0.00% | | | | | | | | | | | | | | | | | | | | | | | | | | | | | | | | | | |
| Test for Residual Heterogeneity: QE(df = 18) = 0.5252, p-val = 1.0000 | | | | | | | | | | | | | | | | | | | | | | | | | | | | | | | | | | |
| Test of Moderators (coefficient 2): QM(df = 1) = 0.1710, p-val = 0.6792 | | | | | | | | | | | | | | | | | | | | | | | | | | | | | | | | | | |
| Model Results: | | | | | | | | | | | | | | | | | | | | | | | | | | | | | | | | | | |
|  | | | | | estimate | | | | | | | | se | | | | | | | zval | | | pval | | | | | | ci.lb | | | | | ci.ub |
| intrcpt | | | | | 0.2639 | | | | | | | | 0.9973 | | | | | | | 0.2646 | | | 0.7913 | | | | | | -1.6908 | | | | | 2.2186 |
| proportion of males | | | | | -0.9357 | | | | | | | | 2.2624 | | | | | | | -0.4136 | | | 0.6792 | | | | | | -5.3699 | | | | | 3.4986 |
| **Change in BMI SDS from baseline (trial’s follow-up)** | | | | | | | | | | | | | | | | | | | | | | | | | | | | | | | | | | |
| Mixed-Effects Model (k = 20; tau^2 estimator: REML) | | | | | | | | | | | | | | | | | | | | | | | | | | | | | | | | | | |
| logLik | | | | | | | | | | deviance | | | | | | | AIC | | | | | | | | | BIC | | | | | | AICc | | |
| -16.8327 | | | | | | | | | | 33.6654 | | | | | | | 39.6654 | | | | | | | | | 42.3365 | | | | | | 41.3797 | | |
| tau^2 (estimated amount of residual heterogeneity): 0 (SE = 0.3333); tau (square root of estimated tau^2 value): 0;  I^2 (residual heterogeneity / unaccounted variability): 0.00%; H^2 (unaccounted variability / sampling variability): 1.00; R^2 (amount of heterogeneity accounted for): 0.00% | | | | | | | | | | | | | | | | | | | | | | | | | | | | | | | | | | |
| Test for Residual Heterogeneity: QE(df = 18) = 0.5836, p-val = 1.0000 | | | | | | | | | | | | | | | | | | | | | | | | | | | | | | | | | | |
| Test of Moderators (coefficient 2): QM(df = 1) = 0.1126, p-val = 0.7372 | | | | | | | | | | | | | | | | | | | | | | | | | | | | | | | | | | |
| Model Results: | | | | | | | | | | | | | | | | | | | | | | | | | | | | | | | | | | |
|  | | | | | estimate | | | | | | | | se | | | | | | | zval | | | pval | | | | | | ci.lb | | | | | ci.ub |
| intrcpt | | | | | 0.0013 | | | | | | | | 0.4718 | | | | | | | 0.0028 | | | 0.9978 | | | | | | -0.9234 | | | | | 0.9261 |
| trial’s follow-up | | | | | -0.0126 | | | | | | | | 0.0374 | | | | | | | -0.3356 | | | 0.7372 | | | | | | -0.0859 | | | | | 0.0608 |
| **Total gastrointestinal adverse events (mean age at baseline)** | | | | | | | | | | | | | | | | | | | | | | | | | | | | | | | | | | |
| Mixed-Effects Model (k = 30; tau^2 estimator: REML) | | | | | | | | | | | | | | | | | | | | | | | | | | | | | | | | | | |
| logLik | | | | | | | | | | deviance | | | | | | | AIC | | | | | | | | | BIC | | | | | | AICc | | |
| -45.3521 | | | | | | | | | | 90.7042 | | | | | | | 96.7042 | | | | | | | | | 100.7008 | | | | | | 97.7042 | | |
| tau^2 (estimated amount of residual heterogeneity): 1.1644 (SE = 0.3331); tau (square root of estimated tau^2 value): 1.0791;  I^2 (residual heterogeneity / unaccounted variability): 96.98%; H^2 (unaccounted variability / sampling variability): 33.16; R^2 (amount of heterogeneity accounted for): 0.00% | | | | | | | | | | | | | | | | | | | | | | | | | | | | | | | | | | |
| Test for Residual Heterogeneity: QE(df = 28) = 343.6861, p-val < .0001 | | | | | | | | | | | | | | | | | | | | | | | | | | | | | | | | | | |
| Test of Moderators (coefficient 2): QM(df = 1) = 0.2327, p-val = 0.6295 | | | | | | | | | | | | | | | | | | | | | | | | | | | | | | | | | | |
| Model Results: | | | | | | | | | | | | | | | | | | | | | | | | | | | | | | | | | | |
|  | | | | | estimate | | | | | | | | se | | | | | | | zval | | | pval | | | | | | ci.lb | | | | | ci.ub |
| intrcpt | | | | | 0.0328 | | | | | | | | 1.5858 | | | | | | | 0.0207 | | | 0.9835 | | | | | | -3.0753 | | | | | 3.1408 |
| mean age at baseline | | | | | 0.0555 | | | | | | | | 0.1151 | | | | | | | 0.4824 | | | 0.6295 | | | | | | -0.1701 | | | | | 0.2812 |
| **Total gastrointestinal adverse events (mean BMI at baseline)** | | | | | | | | | | | | | | | | | | | | | | | | | | | | | | | | | | |
| Mixed-Effects Model (k = 30; tau^2 estimator: REML) | | | | | | | | | | | | | | | | | | | | | | | | | | | | | | | | | | |
| logLik | | | | | | | | | | deviance | | | | | | | AIC | | | | | | | | | BIC | | | | | | AICc | | |
| -44.5372 | | | | | | | | | | 89.0745 | | | | | | | 95.0745 | | | | | | | | | 99.0711 | | | | | | 96.0745 | | |
| tau^2 (estimated amount of residual heterogeneity): 1.0757 (SE = 0.3065); tau (square root of estimated tau^2 value): 1.0371;  I^2 (residual heterogeneity / unaccounted variability): 97.21%; H^2 (unaccounted variability / sampling variability): 35.83; R^2 (amount of heterogeneity accounted for): 4.45% | | | | | | | | | | | | | | | | | | | | | | | | | | | | | | | | | | |
| Test for Residual Heterogeneity: QE(df = 28) = 348.4988, p-val < .0001 | | | | | | | | | | | | | | | | | | | | | | | | | | | | | | | | | | |
| Test of Moderators (coefficient 2): QM(df = 1) = 2.3562, p-val = 0.1248 | | | | | | | | | | | | | | | | | | | | | | | | | | | | | | | | | | |
| Model Results: | | | | | | | | | | | | | | | | | | | | | | | | | | | | | | | | | | |
|  | | | | | estimate | | | | | | | | se | | | | | | | zval | | | pval | | | | | | ci.lb | | | | | ci.ub |
| intrcpt | | | | | -2.2369 | | | | | | | | 1.9153 | | | | | | | -1.1679 | | | 0.2428 | | | | | | -5.9909 | | | | | 1.5170 |
| mean BMI at baseline | | | | | 0.0864 | | | | | | | | 0.0563 | | | | | | | 1.5350 | | | 0.1248 | | | | | | -0.0239 | | | | | 0.1966 |
| **Total gastrointestinal adverse events (mean weight at baseline)** | | | | | | | | | | | | | | | | | | | | | | | | | | | | | | | | | | |
| Mixed-Effects Model (k = 26; tau^2 estimator: REML) | | | | | | | | | | | | | | | | | | | | | | | | | | | | | | | | | | |
| logLik | | | | | | | | | | deviance | | | | | | | AIC | | | | | | | | | BIC | | | | | | AICc | | |
| -39.1178 | | | | | | | | | | 78.2355 | | | | | | | 84.2355 | | | | | | | | | 87.7697 | | | | | | 85.4355 | | |
| tau^2 (estimated amount of residual heterogeneity): 1.1982 (SE = 0.3670); tau (square root of estimated tau^2 value): 1.0946;  I^2 (residual heterogeneity / unaccounted variability): 97.61%; H^2 (unaccounted variability / sampling variability): 41.79; R^2 (amount of heterogeneity accounted for): 12.88% | | | | | | | | | | | | | | | | | | | | | | | | | | | | | | | | | | |
| Test for Residual Heterogeneity: QE(df = 24) = 320.0103, p-val < .0001 | | | | | | | | | | | | | | | | | | | | | | | | | | | | | | | | | | |
| Test of Moderators (coefficient 2): QM(df = 1) = 4.3824, p-val = 0.0363 | | | | | | | | | | | | | | | | | | | | | | | | | | | | | | | | | | |
| Model Results: | | | | | | | | | | | | | | | | | | | | | | | | | | | | | | | | | | |
|  | | | | | estimate | | | | | | | | se | | | | | | | zval | | | pval | | | | | | ci.lb | | | | | ci.ub |
| intrcpt | | | | | -1.8422 | | | | | | | | 1.2295 | | | | | | | -1.4983 | | | 0.1340 | | | | | | -4.2520 | | | | | 0.5676 |
| mean weight at baseline | | | | | 0.0285 | | | | | | | | 0.0136 | | | | | | | 2.0934 | | | 0.0363 | | | | | | 0.0018 | | | | | 0.0552 |
| **Total gastrointestinal adverse events (proportion of males)** | | | | | | | | | | | | | | | | | | | | | | | | | | | | | | | | | | |
| Mixed-Effects Model (k = 32; tau^2 estimator: REML) | | | | | | | | | | | | | | | | | | | | | | | | | | | | | | | | | | |
| logLik | | | | | | | | | | deviance | | | | | | | AIC | | | | | | | | | BIC | | | | | | AICc | | |
| -47.6100 | | | | | | | | | | 95.2200 | | | | | | | 101.2200 | | | | | | | | | 105.4236 | | | | | | 102.1430 | | |
| tau^2 (estimated amount of residual heterogeneity): 1.0741 (SE = 0.2957); tau (square root of estimated tau^2 value): 1.0364;  I^2 (residual heterogeneity / unaccounted variability): 97.19%; H^2 (unaccounted variability / sampling variability): 35.58; R^2 (amount of heterogeneity accounted for): 0.00% | | | | | | | | | | | | | | | | | | | | | | | | | | | | | | | | | | |
| Test for Residual Heterogeneity: QE(df = 30) = 360.1592, p-val < .0001 | | | | | | | | | | | | | | | | | | | | | | | | | | | | | | | | | | |
| Test of Moderators (coefficient 2): QM(df = 1) = 0.7112, p-val = 0.3991 | | | | | | | | | | | | | | | | | | | | | | | | | | | | | | | | | | |
| Model Results: | | | | | | | | | | | | | | | | | | | | | | | | | | | | | | | | | | |
|  | | | | | | | estimate | | | | | | se | | | | | | | zval | | | pval | | | | | | ci.lb | | | | | ci.ub |
| intrcpt | | | | | | | 1.3659 | | | | | | 0.7929 | | | | | | | 1.7227 | | | 0.0849 | | | | | | -0.1881 | | | | | 2.9200 |
| proportion of males | | | | | | | -1.4415 | | | | | | 1.7094 | | | | | | | -0.8433 | | | 0.3991 | | | | | | -4.7918 | | | | | 1.9088 |
| **Total gastrointestinal adverse events (trial’s follow-up)** | | | | | | | | | | | | | | | | | | | | | | | | | | | | | | | | | | |
| Mixed-Effects Model (k = 36; tau^2 estimator: REML) | | | | | | | | | | | | | | | | | | | | | | | | | | | | | | | | | | |
| logLik | | | | | | | | | | deviance | | | | | | | AIC | | | | | | | | | BIC | | | | | | AICc | | |
| -51.8458 | | | | | | | | | | 103.6915 | | | | | | | 109.6915 | | | | | | | | | 114.2706 | | | | | | 110.4915 | | |
| tau^2 (estimated amount of residual heterogeneity): 0.8833 (SE = 0.2316); tau (square root of estimated tau^2 value): 0.9398;  I^2 (residual heterogeneity / unaccounted variability): 96.34%; H^2 (unaccounted variability / sampling variability): 27.35; R^2 (amount of heterogeneity accounted for): 2.50% | | | | | | | | | | | | | | | | | | | | | | | | | | | | | | | | | | |
| Test for Residual Heterogeneity: QE(df = 34) = 325.3949, p-val < .0001 | | | | | | | | | | | | | | | | | | | | | | | | | | | | | | | | | | |
| Test of Moderators (coefficient 2): QM(df = 1) = 1.0779, p-val = 0.2992 | | | | | | | | | | | | | | | | | | | | | | | | | | | | | | | | | | |
| Model Results: | | | | | | | | | | | | | | | | | | | | | | | | | | | | | | | | | | |
|  | | | | | | | estimate | | | | | | se | | | | | | | zval | | | pval | | | | | | ci.lb | | | | | ci.ub |
| intrcpt | | | | | | | 0.4596 | | | | | | 0.2516 | | | | | | | 1.8271 | | | 0.0677 | | | | | | -0.0334 | | | | | 0.9527 |
| trial’s follow-up | | | | | | | 0.0244 | | | | | | 0.0235 | | | | | | | 1.0382 | | | 0.2992 | | | | | | -0.0216 | | | | | 0.0703 |
| **Discontinuation due to adverse events (mean age at baseline)** | | | | | | | | | | | | | | | | | | | | | | | | | | | | | | | | | | |
| Mixed-Effects Model (k = 34; tau^2 estimator: REML) | | | | | | | | | | | | | | | | | | | | | | | | | | | | | | | | | | |
| logLik | | | | | | | | | | deviance | | | | | | | AIC | | | | | | | | | BIC | | | | | | AICc | | |
| 25.3614 | | | | | | | | | | -50.7228 | | | | | | | -44.7228 | | | | | | | | | -40.3256 | | | | | | -43.8657 | | |
| tau^2 (estimated amount of residual heterogeneity): 0 (SE = 0.0029); tau (square root of estimated tau^2 value): 0;  I^2 (residual heterogeneity / unaccounted variability): 0.00%; H^2 (unaccounted variability / sampling variability): 1.00; R^2 (amount of heterogeneity accounted for): 0.00% | | | | | | | | | | | | | | | | | | | | | | | | | | | | | | | | | | |
| Test for Residual Heterogeneity: QE(df = 32) = 8.7730, p-val = 1.0000 | | | | | | | | | | | | | | | | | | | | | | | | | | | | | | | | | | |
| Test of Moderators (coefficient 2): QM(df = 1) = 0.1124, p-val = 0.7375 | | | | | | | | | | | | | | | | | | | | | | | | | | | | | | | | | | |
| Model Results: | | | | | | | | | | | | | | | | | | | | | | | | | | | | | | | | | | |
|  | | | | | | | estimate | | | | | | se | | | | | | | zval | | | pval | | | | | | ci.lb | | | | | ci.ub |
| intrcpt | | | | | | | -0.0501 | | | | | | 0.2679 | | | | | | | -0.1870 | | | 0.8517 | | | | | | -0.5751 | | | | | 0.4750 |
| mean age at baseline | | | | | | | 0.0064 | | | | | | 0.0191 | | | | | | | 0.3352 | | | 0.7375 | | | | | | -0.0310 | | | | | 0.0438 |
| **Discontinuation due to adverse events (mean BMI at baseline)** | | | | | | | | | | | | | | | | | | | | | | | | | | | | | | | | | | |
| Mixed-Effects Model (k = 34; tau^2 estimator: REML) | | | | | | | | | | | | | | | | | | | | | | | | | | | | | | | | | | |
| logLik | | | | | | | | | | deviance | | | | | | | AIC | | | | | | | | | BIC | | | | | | AICc | | |
| 25.4088 | | | | | | | | | | -50.8175 | | | | | | | -44.8175 | | | | | | | | | -40.4203 | | | | | | -43.9604 | | |
| tau^2 (estimated amount of residual heterogeneity): 0 (SE = 0.0029); tau (square root of estimated tau^2 value): 0;  I^2 (residual heterogeneity / unaccounted variability): 0.00%; H^2 (unaccounted variability / sampling variability): 1.00; R^2 (amount of heterogeneity accounted for): 0.00% | | | | | | | | | | | | | | | | | | | | | | | | | | | | | | | | | | |
| Test for Residual Heterogeneity: QE(df = 32) = 8.7007, p-val = 1.0000 | | | | | | | | | | | | | | | | | | | | | | | | | | | | | | | | | | |
| Test of Moderators (coefficient 2): QM(df = 1) = 0.1847, p-val = 0.6674 | | | | | | | | | | | | | | | | | | | | | | | | | | | | | | | | | | |
| Model Results: | | | | | | | | | | | | | | | | | | | | | | | | | | | | | | | | | | |
|  | | | | | | | estimate | | | | | | se | | | | | | | zval | | | pval | | | | | | ci.lb | | | | | ci.ub |
| intrcpt | | | | | | | 0.2259 | | | | | | 0.4344 | | | | | | | 0.5199 | | | 0.6031 | | | | | | -0.6256 | | | | | 1.0774 |
| mean BMI at baseline | | | | | | | -0.0053 | | | | | | 0.0123 | | | | | | | -0.4297 | | | 0.6674 | | | | | | -0.0294 | | | | | 0.0188 |
| **Discontinuation due to adverse events (mean weight at baseline)** | | | | | | | | | | | | | | | | | | | | | | | | | | | | | | | | | | |
| Mixed-Effects Model (k = 30; tau^2 estimator: REML) | | | | | | | | | | | | | | | | | | | | | | | | | | | | | | | | | | |
| logLik | | | | | | | | | | deviance | | | | | | | AIC | | | | | | | | | BIC | | | | | | AICc | | |
| 25.0608 | | | | | | | | | | -50.1217 | | | | | | | -44.1217 | | | | | | | | | -40.1250 | | | | | | -43.1217 | | |
| tau^2 (estimated amount of residual heterogeneity): 0 (SE = 0.0029); tau (square root of estimated tau^2 value): 0;  I^2 (residual heterogeneity / unaccounted variability): 0.00%; H^2 (unaccounted variability / sampling variability): 1.00; R^2 (amount of heterogeneity accounted for): 0.00% | | | | | | | | | | | | | | | | | | | | | | | | | | | | | | | | | | |
| Test for Residual Heterogeneity: QE(df = 28) = 4.8687, p-val = 1.0000 | | | | | | | | | | | | | | | | | | | | | | | | | | | | | | | | | | |
| Test of Moderators (coefficient 2): QM(df = 1) = 0.0140, p-val = 0.9059 | | | | | | | | | | | | | | | | | | | | | | | | | | | | | | | | | | |
| Model Results: | | | | | | | | | | | | | | | | | | | | | | | | | | | | | | | | | | |
|  | | | | | | | estimate | | | | | | se | | | | | | | zval | | | pval | | | | | | ci.lb | | | | | ci.ub |
| intrcpt | | | | | | | 0.0657 | | | | | | 0.2534 | | | | | | | 0.2592 | | | 0.7955 | | | | | | -0.4310 | | | | | 0.5624 |
| mean weight at baseline | | | | | | | -0.0003 | | | | | | 0.0026 | | | | | | | -0.1182 | | | 0.9059 | | | | | | -0.0054 | | | | | 0.0048 |
| **Discontinuation due to adverse events (proportion of males)** | | | | | | | | | | | | | | | | | | | | | | | | | | | | | | | | | | |
| Mixed-Effects Model (k = 32; tau^2 estimator: REML) | | | | | | | | | | | | | | | | | | | | | | | | | | | | | | | | | | |
| logLik | | | | | | | | | | deviance | | | | | | | AIC | | | | | | | | | BIC | | | | | | AICc | | |
| 27.0112 | | | | | | | | | | -54.0223 | | | | | | | -48.0223 | | | | | | | | | -43.8188 | | | | | | -47.0993 | | |
| tau^2 (estimated amount of residual heterogeneity): 0 (SE = 0.0030); tau (square root of estimated tau^2 value): 0;  I^2 (residual heterogeneity / unaccounted variability): 0.00%; H^2 (unaccounted variability / sampling variability): 1.00; R^2 (amount of heterogeneity accounted for): 0.00% | | | | | | | | | | | | | | | | | | | | | | | | | | | | | | | | | | |
| Test for Residual Heterogeneity: QE(df = 30) = 5.0527, p-val = 1.0000 | | | | | | | | | | | | | | | | | | | | | | | | | | | | | | | | | | |
| Test of Moderators (coefficient 2): QM(df = 1) = 0.0005, p-val = 0.9827 | | | | | | | | | | | | | | | | | | | | | | | | | | | | | | | | | | |
| Model Results: | | | | | | | | | | | | | | | | | | | | | | | | | | | | | | | | | | |
|  | | | | estimate | | | | | | | | | se | | | | | | | zval | | | pval | | | | | | ci.lb | | | | | ci.ub |
| intrcpt | | | | 0.0347 | | | | | | | | | 0.1037 | | | | | | | 0.3343 | | | 0.7382 | | | | | | -0.1687 | | | | | 0.2380 |
| proportion of males | | | | 0.0056 | | | | | | | | | 0.2598 | | | | | | | 0.0217 | | | 0.9827 | | | | | | -0.5036 | | | | | 0.5148 |
| **Discontinuation due to adverse events (trial’s follow-up)** | | | | | | | | | | | | | | | | | | | | | | | | | | | | | | | | | | |
| Mixed-Effects Model (k = 36; tau^2 estimator: REML) | | | | | | | | | | | | | | | | | | | | | | | | | | | | | | | | | | |
| logLik | | | | | | | | | | deviance | | | | | | | AIC | | | | | | | | | BIC | | | | | | AICc | | |
| 26.3145 | | | | | | | | | | -52.6291 | | | | | | | -46.6291 | | | | | | | | | -42.0500 | | | | | | -45.8291 | | |
| tau^2 (estimated amount of residual heterogeneity): 0 (SE = 0.0029); tau (square root of estimated tau^2 value): 0;  I^2 (residual heterogeneity / unaccounted variability): 0.00%; H^2 (unaccounted variability / sampling variability): 1.00; R^2 (amount of heterogeneity accounted for): 0.00% | | | | | | | | | | | | | | | | | | | | | | | | | | | | | | | | | | |
| Test for Residual Heterogeneity: QE(df = 34) = 8.9349, p-val = 1.0000 | | | | | | | | | | | | | | | | | | | | | | | | | | | | | | | | | | |
| Test of Moderators (coefficient 2): QM(df = 1) = 0.0087, p-val = 0.9255 | | | | | | | | | | | | | | | | | | | | | | | | | | | | | | | | | | |
| Model Results: | | | | | | | | | | | | | | | | | | | | | | | | | | | | | | | | | | |
|  | | | | estimate | | | | | | | | | se | | | | | | | zval | | | pval | | | | | | ci.lb | | | | | ci.ub |
| intrcpt | | | | 0.0446 | | | | | | | | | 0.0574 | | | | | | | 0.7767 | | | 0.4373 | | | | | | -0.0679 | | | | | 0.1571 |
| trial’s follow-up | | | | -0.0005 | | | | | | | | | 0.0049 | | | | | | | -0.0935 | | | 0.9255 | | | | | | -0.0101 | | | | | 0.0092 |
| **Serious adverse events (mean age at baseline)** | | | | | | | | | | | | | | | | | | | | | | | | | | | | | | | | | | |
| Mixed-Effects Model (k = 24; tau^2 estimator: REML) | | | | | | | | | | | | | | | | | | | | | | | | | | | | | | | | | | |
| logLik | | | | | | | | | | deviance | | | | | | | AIC | | | | | | | | | BIC | | | | | | AICc | | |
| 23.3316 | | | | | | | | | | -46.6631 | | | | | | | -40.6631 | | | | | | | | | -37.3900 | | | | | | -39.3298 | | |
| tau^2 (estimated amount of residual heterogeneity): 0 (SE = 0.0030); tau (square root of estimated tau^2 value): 0;  I^2 (residual heterogeneity / unaccounted variability): 0.00%; H^2 (unaccounted variability / sampling variability): 1.00; R^2 (amount of heterogeneity accounted for): 0.00% | | | | | | | | | | | | | | | | | | | | | | | | | | | | | | | | | | |
| Test for Residual Heterogeneity: QE(df = 22) = 3.1976, p-val = 1.0000 | | | | | | | | | | | | | | | | | | | | | | | | | | | | | | | | | | |
| Test of Moderators (coefficient 2): QM(df = 1) = 0.0188, p-val = 0.8911 | | | | | | | | | | | | | | | | | | | | | | | | | | | | | | | | | | |
| Model Results: | | | | | | | | | | | | | | | | | | | | | | | | | | | | | | | | | | |
|  | | | | estimate | | | | | | | | | se | | | | | | | zval | | | pval | | | | | | ci.lb | | | | | ci.ub |
| intrcpt | | | | -0.0168 | | | | | | | | | 0.4225 | | | | | | | -0.0397 | | | 0.9683 | | | | | | -0.8448 | | | | | 0.8113 |
| mean age at baseline | | | | 0.0040 | | | | | | | | | 0.0296 | | | | | | | 0.1369 | | | 0.8911 | | | | | | -0.0539 | | | | | 0.0620 |
| **Serious adverse events (mean BMI at baseline)** | | | | | | | | | | | | | | | | | | | | | | | | | | | | | | | | | | |
| Mixed-Effects Model (k = 22; tau^2 estimator: REML) | | | | | | | | | | | | | | | | | | | | | | | | | | | | | | | | | | |
| logLik | | | | | | | | | | deviance | | | | | | | AIC | | | | | | | | | BIC | | | | | | AICc | | |
| 20.3690 | | | | | | | | | | -40.7380 | | | | | | | -34.7380 | | | | | | | | | -31.7508 | | | | | | -33.2380 | | |
| tau^2 (estimated amount of residual heterogeneity): 0 (SE = 0.0031); tau (square root of estimated tau^2 value): 0;  I^2 (residual heterogeneity / unaccounted variability): 0.00%; H^2 (unaccounted variability / sampling variability): 1.00; R^2 (amount of heterogeneity accounted for): 0.00% | | | | | | | | | | | | | | | | | | | | | | | | | | | | | | | | | | |
| Test for Residual Heterogeneity: QE(df = 20) = 2.9753, p-val = 1.0000 | | | | | | | | | | | | | | | | | | | | | | | | | | | | | | | | | | |
| Test of Moderators (coefficient 2): QM(df = 1) = 0.0108, p-val = 0.9174 | | | | | | | | | | | | | | | | | | | | | | | | | | | | | | | | | | |
| Model Results: | | | | | | | | | | | | | | | | | | | | | | | | | | | | | | | | | | |
|  | | | | estimate | | | | | | | | | se | | | | | | | zval | | | pval | | | | | | ci.lb | | | | | ci.ub |
| intrcpt | | | | 0.1001 | | | | | | | | | 0.5374 | | | | | | | 0.1862 | | | 0.8523 | | | | | | -0.9533 | | | | | 1.1534 |
| mean BMI at baseline | | | | -0.0016 | | | | | | | | | 0.0151 | | | | | | | -0.1038 | | | 0.9174 | | | | | | -0.0312 | | | | | 0.0281 |
| **Serious adverse events (mean weight at baseline)** | | | | | | | | | | | | | | | | | | | | | | | | | | | | | | | | | | |
| Mixed-Effects Model (k = 22; tau^2 estimator: REML) | | | | | | | | | | | | | | | | | | | | | | | | | | | | | | | | | | |
| logLik | | | | | | | | | | deviance | | | | | | | AIC | | | | | | | | | BIC | | | | | | AICc | | |
| 20.5178 | | | | | | | | | | -41.0357 | | | | | | | -35.0357 | | | | | | | | | -32.0485 | | | | | | -33.5357 | | |
| tau^2 (estimated amount of residual heterogeneity): 0 (SE = 0.0031); tau (square root of estimated tau^2 value): 0;  I^2 (residual heterogeneity / unaccounted variability): 0.00%; H^2 (unaccounted variability / sampling variability): 1.00; R^2 (amount of heterogeneity accounted for): 0.00% | | | | | | | | | | | | | | | | | | | | | | | | | | | | | | | | | | |
| Test for Residual Heterogeneity: QE(df = 20) = 2.4671, p-val = 1.0000 | | | | | | | | | | | | | | | | | | | | | | | | | | | | | | | | | | |
| Test of Moderators (coefficient 2): QM(df = 1) = 0.0077, p-val = 0.9301 | | | | | | | | | | | | | | | | | | | | | | | | | | | | | | | | | | |
| Model Results: | | | | | | | | | | | | | | | | | | | | | | | | | | | | | | | | | | |
|  | | | | estimate | | | | | | | | | se | | | | | | | zval | | | pval | | | | | | ci.lb | | | | | ci.ub |
| intrcpt | | | | 0.0116 | | | | | | | | | 0.3390 | | | | | | | 0.0342 | | | 0.9727 | | | | | | -0.6529 | | | | | 0.6761 |
| mean weight at baseline | | | | 0.0003 | | | | | | | | | 0.0034 | | | | | | | 0.0877 | | | 0.9301 | | | | | | -0.0065 | | | | | 0.0071 |
| **Serious adverse events (proportion of males)** | | | | | | | | | | | | | | | | | | | | | | | | | | | | | | | | | | |
| Mixed-Effects Model (k = 24; tau^2 estimator: REML) | | | | | | | | | | | | | | | | | | | | | | | | | | | | | | | | | | |
| logLik | | | | | | | | | | deviance | | | | | | | AIC | | | | | | | | | BIC | | | | | | AICc | | |
| 23.2533 | | | | | | | | | | -46.5065 | | | | | | | -40.5065 | | | | | | | | | -37.2334 | | | | | | -39.1732 | | |
| tau^2 (estimated amount of residual heterogeneity): 0 (SE = 0.0030); tau (square root of estimated tau^2 value): 0;  I^2 (residual heterogeneity / unaccounted variability): 0.00%; H^2 (unaccounted variability / sampling variability): 1.00; R^2 (amount of heterogeneity accounted for): 0.00% | | | | | | | | | | | | | | | | | | | | | | | | | | | | | | | | | | |
| Test for Residual Heterogeneity: QE(df = 22) = 3.2055, p-val = 1.0000 | | | | | | | | | | | | | | | | | | | | | | | | | | | | | | | | | | |
| Test of Moderators (coefficient 2): QM(df = 1) = 0.0109, p-val = 0.9170 | | | | | | | | | | | | | | | | | | | | | | | | | | | | | | | | | | |
| Model Results: | | | | | | | | | | | | | | | | | | | | | | | | | | | | | | | | | | |
|  | | | | | estimate | | | | | | | | se | | | | | | | zval | | | pval | | | | | | ci.lb | | | | | ci.ub |
| intrcpt | | | | | 0.0524 | | | | | | | | 0.1112 | | | | | | | 0.4709 | | | 0.6377 | | | | | | -0.1656 | | | | | 0.2703 |
| proportion of males | | | | | -0.0293 | | | | | | | | 0.2815 | | | | | | | -0.1042 | | | 0.9170 | | | | | | -0.5810 | | | | | 0.5224 |
| **Serious adverse events (trial’s follow-up)** | | | | | | | | | | | | | | | | | | | | | | | | | | | | | | | | | | |
| Mixed-Effects Model (k = 26; tau^2 estimator: REML) | | | | | | | | | | | | | | | | | | | | | | | | | | | | | | | | | | |
| logLik | | | | | | | | | | deviance | | | | | | | AIC | | | | | | | | | BIC | | | | | | AICc | | |
| 24.3653 | | | | | | | | | | -48.7305 | | | | | | | -42.7305 | | | | | | | | | -39.1964 | | | | | | -41.5305 | | |
| tau^2 (estimated amount of residual heterogeneity): 0 (SE = 0.0029); tau (square root of estimated tau^2 value): 0;  I^2 (residual heterogeneity / unaccounted variability): 0.00%; H^2 (unaccounted variability / sampling variability): 1.00; R^2 (amount of heterogeneity accounted for): 0.00% | | | | | | | | | | | | | | | | | | | | | | | | | | | | | | | | | | |
| Test for Residual Heterogeneity: QE(df = 24) = 3.1444, p-val = 1.0000 | | | | | | | | | | | | | | | | | | | | | | | | | | | | | | | | | | |
| Test of Moderators (coefficient 2): QM(df = 1) = 0.0957, p-val = 0.7570 | | | | | | | | | | | | | | | | | | | | | | | | | | | | | | | | | | |
| Model Results: | | | | | | | | | | | | | | | | | | | | | | | | | | | | | | | | | | |
|  | | | | | estimate | | | | | | | | se | | | | | | | zval | | | pval | | | | | | ci.lb | | | | | ci.ub |
| intrcpt | | | | | 0.0231 | | | | | | | | 0.0609 | | | | | | | 0.3797 | | | 0.7042 | | | | | | -0.0962 | | | | | 0.1424 |
| trial’s follow-up | | | | | 0.0016 | | | | | | | | 0.0052 | | | | | | | 0.3094 | | | 0.7570 | | | | | | -0.0086 | | | | | 0.0118 |
| **Nausea events (mean age at baseline)** | | | | | | | | | | | | | | | | | | | | | | | | | | | | | | | | | | |
| Mixed-Effects Model (k = 32; tau^2 estimator: REML) | | | | | | | | | | | | | | | | | | | | | | | | | | | | | | | | | | |
| logLik | | | | | | | | | | deviance | | | | | | | AIC | | | | | | | | | BIC | | | | | | AICc | | |
| -17.7527 | | | | | | | | | | 35.5055 | | | | | | | 41.5055 | | | | | | | | | 45.7091 | | | | | | 42.4286 | | |
| tau^2 (estimated amount of residual heterogeneity): 0.0694 (SE = 0.0257); tau (square root of estimated tau^2 value): 0.2634;  I^2 (residual heterogeneity / unaccounted variability): 79.16%; H^2 (unaccounted variability / sampling variability): 4.80; R^2 (amount of heterogeneity accounted for): 0.00% | | | | | | | | | | | | | | | | | | | | | | | | | | | | | | | | | | |
| Test for Residual Heterogeneity: QE(df = 30) = 110.5285, p-val < .0001 | | | | | | | | | | | | | | | | | | | | | | | | | | | | | | | | | | |
| Test of Moderators (coefficient 2): QM(df = 1) = 0.0313, p-val = 0.8595 | | | | | | | | | | | | | | | | | | | | | | | | | | | | | | | | | | |
| Model Results: | | | | | | | | | | | | | | | | | | | | | | | | | | | | | | | | | | |
|  | | | | | estimate | | | | | | | | se | | | | | | | zval | | | pval | | | | | | ci.lb | | | | | ci.ub |
| intrcpt | | | | | 0.1383 | | | | | | | | 0.7236 | | | | | | | 0.1912 | | | 0.8484 | | | | | | -1.2798 | | | | | 1.5565 |
| mean age at baseline | | | | | 0.0088 | | | | | | | | 0.0500 | | | | | | | 0.1770 | | | 0.8595 | | | | | | -0.0891 | | | | | 0.1067 |
| **Nausea events (mean BMI at baseline)** | | | | | | | | | | | | | | | | | | | | | | | | | | | | | | | | | | |
| Mixed-Effects Model (k = 28; tau^2 estimator: REML) | | | | | | | | | | | | | | | | | | | | | | | | | | | | | | | | | | |
| logLik | | | | | | | | | | deviance | | | | | | | AIC | | | | | | | | | BIC | | | | | | AICc | | |
| -17.1734 | | | | | | | | | | 34.3467 | | | | | | | 40.3467 | | | | | | | | | 44.1210 | | | | | | 41.4376 | | |
| tau^2 (estimated amount of residual heterogeneity): 0.1022 (SE = 0.0377); tau (square root of estimated tau^2 value): 0.3196;  I^2 (residual heterogeneity / unaccounted variability): 84.94%; H^2 (unaccounted variability / sampling variability): 6.64; R^2 (amount of heterogeneity accounted for): 0.00% | | | | | | | | | | | | | | | | | | | | | | | | | | | | | | | | | | |
| Test for Residual Heterogeneity: QE(df = 26) = 113.2405, p-val < .0001 | | | | | | | | | | | | | | | | | | | | | | | | | | | | | | | | | | |
| Test of Moderators (coefficient 2): QM(df = 1) = 1.0506, p-val = 0.3054 | | | | | | | | | | | | | | | | | | | | | | | | | | | | | | | | | | |
| Model Results: | | | | | | | | | | | | | | | | | | | | | | | | | | | | | | | | | | |
|  | | | | | estimate | | | | | | | | se | | | | | | | zval | | | pval | | | | | | ci.lb | | | | | ci.ub |
| intrcpt | | | | | 1.4130 | | | | | | | | 1.0899 | | | | | | | 1.2964 | | | 0.1948 | | | | | | -0.7232 | | | | | 3.5492 |
| mean BMI at baseline | | | | | -0.0312 | | | | | | | | 0.0304 | | | | | | | -1.0250 | | | 0.3054 | | | | | | -0.0908 | | | | | 0.0284 |
| **Nausea events (mean weightat baseline)** | | | | | | | | | | | | | | | | | | | | | | | | | | | | | | | | | | |
| Mixed-Effects Model (k = 28; tau^2 estimator: REML) | | | | | | | | | | | | | | | | | | | | | | | | | | | | | | | | | | |
| logLik | | | | | | | | | | deviance | | | | | | | AIC | | | | | | | | | BIC | | | | | | AICc | | |
| -17.8639 | | | | | | | | | | 35.7278 | | | | | | | 41.7278 | | | | | | | | | 45.5021 | | | | | | 42.8187 | | |
| tau^2 (estimated amount of residual heterogeneity): 0.1001 (SE = 0.0375); tau (square root of estimated tau^2 value): 0.3164;  I^2 (residual heterogeneity / unaccounted variability): 84.49%; H^2 (unaccounted variability / sampling variability): 6.45; R^2 (amount of heterogeneity accounted for): 0.00% | | | | | | | | | | | | | | | | | | | | | | | | | | | | | | | | | | |
| Test for Residual Heterogeneity: QE(df = 26) = 109.5512, p-val < .0001 | | | | | | | | | | | | | | | | | | | | | | | | | | | | | | | | | | |
| Test of Moderators (coefficient 2): QM(df = 1) = 0.0467, p-val = 0.8290 | | | | | | | | | | | | | | | | | | | | | | | | | | | | | | | | | | |
| Model Results: | | | | | | | | | | | | | | | | | | | | | | | | | | | | | | | | | | |
|  | | | | | estimate | | | | | | | | se | | | | | | | zval | | | pval | | | | | | ci.lb | | | | | ci.ub |
| intrcpt | | | | | 0.1557 | | | | | | | | 0.6798 | | | | | | | 0.2290 | | | 0.8189 | | | | | | -1.1767 | | | | | 1.4881 |
| mean weight at baseline | | | | | 0.0015 | | | | | | | | 0.0070 | | | | | | | 0.2160 | | | 0.8290 | | | | | | -0.0121 | | | | | 0.0151 |
| **Nausea events (proportion of males)** | | | | | | | | | | | | | | | | | | | | | | | | | | | | | | | | | | |
| Mixed-Effects Model (k = 32; tau^2 estimator: REML) | | | | | | | | | | | | | | | | | | | | | | | | | | | | | | | | | | |
| logLik | | | | | | | | | | deviance | | | | | | | AIC | | | | | | | | | BIC | | | | | | AICc | | |
| -17.8247 | | | | | | | | | | 35.6494 | | | | | | | 41.6494 | | | | | | | | | 45.8530 | | | | | | 42.5725 | | |
| tau^2 (estimated amount of residual heterogeneity): 0.0731 (SE = 0.0269); tau (square root of estimated tau^2 value): 0.2703;  I^2 (residual heterogeneity / unaccounted variability): 80.04%; H^2 (unaccounted variability / sampling variability): 5.01; R^2 (amount of heterogeneity accounted for): 0.00% | | | | | | | | | | | | | | | | | | | | | | | | | | | | | | | | | | |
| Test for Residual Heterogeneity: QE(df = 30) = 115.5973, p-val < .0001 | | | | | | | | | | | | | | | | | | | | | | | | | | | | | | | | | | |
| Test of Moderators (coefficient 2): QM(df = 1) = 0.1364, p-val = 0.7119 | | | | | | | | | | | | | | | | | | | | | | | | | | | | | | | | | | |
| Model Results: | | | | | | | | | | | | | | | | | | | | | | | | | | | | | | | | | | |
|  | | | | | estimate | | | | | | | | se | | | | | | | zval | | | pval | | | | | | ci.lb | | | | | ci.ub |
| intrcpt | | | | | 0.3523 | | | | | | | | 0.2375 | | | | | | | 1.4834 | | | 0.1380 | | | | | | -0.1132 | | | | | 0.8178 |
| proportion of males | | | | | -0.2106 | | | | | | | | 0.5702 | | | | | | | -0.3693 | | | 0.7119 | | | | | | -1.3282 | | | | | 0.9071 |
| **Nausea events (trial’s follow-up)** | | | | | | | | | | | | | | | | | | | | | | | | | | | | | | | | | | |
| Mixed-Effects Model (k = 32; tau^2 estimator: REML) | | | | | | | | | | | | | | | | | | | | | | | | | | | | | | | | | | |
| logLik | | | | | | | | | | deviance | | | | | | | AIC | | | | | | | | | BIC | | | | | | AICc | | |
| -14.8177 | | | | | | | | | | 29.6354 | | | | | | | 35.6354 | | | | | | | | | 39.8390 | | | | | | 36.5585 | | |
| tau^2 (estimated amount of residual heterogeneity): 0.0506 (SE = 0.0204); tau (square root of estimated tau^2 value): 0.2250;  I^2 (residual heterogeneity / unaccounted variability): 73.38%; H^2 (unaccounted variability / sampling variability): 3.76; R^2 (amount of heterogeneity accounted for): 23.02% | | | | | | | | | | | | | | | | | | | | | | | | | | | | | | | | | | |
| Test for Residual Heterogeneity: QE(df = 30) = 102.8681, p-val < .0001 | | | | | | | | | | | | | | | | | | | | | | | | | | | | | | | | | | |
| Test of Moderators (coefficient 2): QM(df = 1) = 6.5092, p-val = 0.0107 | | | | | | | | | | | | | | | | | | | | | | | | | | | | | | | | | | |
| Model Results: | | | | | | | | | | | | | | | | | | | | | | | | | | | | | | | | | | |
|  | | | | | estimate | | | | | | | | se | | | | | | | zval | | | pval | | | | | | ci.lb | | | | | ci.ub |
| intrcpt | | | | | 0.0152 | | | | | | | | 0.1082 | | | | | | | 0.1407 | | | 0.8881 | | | | | | -0.1968 | | | | | 0.2273 |
| trial’s follow-up | | | | | 0.0259 | | | | | | | | 0.0101 | | | | | | | 2.5513 | | | 0.0107 | | | | | | 0.0060 | | | | | 0.0458 |
| **Vomiting events (mean age at baseline)** | | | | | | | | | | | | | | | | | | | | | | | | | | | | | | | | | | |
| Mixed-Effects Model (k = 24; tau^2 estimator: REML) | | | | | | | | | | | | | | | | | | | | | | | | | | | | | | | | | | |
| logLik | | | | | | | | | | deviance | | | | | | | AIC | | | | | | | | | BIC | | | | | | AICc | | |
| 4.8725 | | | | | | | | | | -9.7450 | | | | | | | -3.7450 | | | | | | | | | -0.4719 | | | | | | -2.4117 | | |
| tau^2 (estimated amount of residual heterogeneity): 0.0154 (SE = 0.0110); tau (square root of estimated tau^2 value): 0.1243;  I^2 (residual heterogeneity / unaccounted variability): 43.51%; H^2 (unaccounted variability / sampling variability): 1.77; R^2 (amount of heterogeneity accounted for): 2.28% | | | | | | | | | | | | | | | | | | | | | | | | | | | | | | | | | | |
| Test for Residual Heterogeneity: QE(df = 22) = 35.5722, p-val = 0.0337 | | | | | | | | | | | | | | | | | | | | | | | | | | | | | | | | | | |
| Test of Moderators (coefficient 2): QM(df = 1) = 0.4854, p-val = 0.4860 | | | | | | | | | | | | | | | | | | | | | | | | | | | | | | | | | | |
| Model Results: | | | | | | | | | | | | | | | | | | | | | | | | | | | | | | | | | | |
|  | | | | | estimate | | | | | | | | se | | | | | | | zval | | | pval | | | | | | ci.lb | | | | | ci.ub |
| intrcpt | | | | | -0.1858 | | | | | | | | 0.5044 | | | | | | | -0.3684 | | | 0.7126 | | | | | | -1.1745 | | | | | 0.8028 |
| mean age at baseline | | | | | 0.0243 | | | | | | | | 0.0348 | | | | | | | 0.6967 | | | 0.4860 | | | | | | -0.0440 | | | | | 0.0925 |
| **Vomiting events (mean BMI at baseline)** | | | | | | | | | | | | | | | | | | | | | | | | | | | | | | | | | | |
| Mixed-Effects Model (k = 20; tau^2 estimator: REML) | | | | | | | | | | | | | | | | | | | | | | | | | | | | | | | | | | |
| logLik | | | | | | | | | | deviance | | | | | | | AIC | | | | | | | | | BIC | | | | | | AICc | | |
| 4.1247 | | | | | | | | | | -8.2494 | | | | | | | -2.2494 | | | | | | | | | 0.4218 | | | | | | -0.5351 | | |
| tau^2 (estimated amount of residual heterogeneity): 0.0145 (SE = 0.0121); tau (square root of estimated tau^2 value): 0.1203;  I^2 (residual heterogeneity / unaccounted variability): 40.47%; H^2 (unaccounted variability / sampling variability): 1.68; R^2 (amount of heterogeneity accounted for): 24.21% | | | | | | | | | | | | | | | | | | | | | | | | | | | | | | | | | | |
| Test for Residual Heterogeneity: QE(df = 18) = 27.4390, p-val = 0.0711 | | | | | | | | | | | | | | | | | | | | | | | | | | | | | | | | | | |
| Test of Moderators (coefficient 2): QM(df = 1) = 2.9958, p-val = 0.0835 | | | | | | | | | | | | | | | | | | | | | | | | | | | | | | | | | | |
| Model Results: | | | | | | | | | | | | | | | | | | | | | | | | | | | | | | | | | | |
|  | | | | | estimate | | | | | | | | se | | | | | | | zval | | | pval | | | | | | ci.lb | | | | | ci.ub |
| intrcpt | | | | | -1.5298 | | | | | | | | 0.9872 | | | | | | | -1.5497 | | | 0.1212 | | | | | | -3.4646 | | | | | 0.4050 |
| mean BMI at baseline | | | | | 0.0489 | | | | | | | | 0.0282 | | | | | | | 1.7308 | | | 0.0835 | | | | | | -0.0065 | | | | | 0.1042 |
| **Vomiting events (mean weight at baseline)** | | | | | | | | | | | | | | | | | | | | | | | | | | | | | | | | | | |
| Mixed-Effects Model (k = 20; tau^2 estimator: REML) | | | | | | | | | | | | | | | | | | | | | | | | | | | | | | | | | | |
| logLik | | | | | | | | | | deviance | | | | | | | AIC | | | | | | | | | BIC | | | | | | AICc | | |
| 3.3319 | | | | | | | | | | -6.6638 | | | | | | | -0.6638 | | | | | | | | | 2.0073 | | | | | | 1.0505 | | |
| tau^2 (estimated amount of residual heterogeneity): 0.0159 (SE = 0.0129); tau (square root of estimated tau^2 value): 0.1261;  I^2 (residual heterogeneity / unaccounted variability): 42.25%; H^2 (unaccounted variability / sampling variability): 1.73; R^2 (amount of heterogeneity accounted for): 20.60% | | | | | | | | | | | | | | | | | | | | | | | | | | | | | | | | | | |
| Test for Residual Heterogeneity: QE(df = 18) = 28.5288, p-val = 0.0545 | | | | | | | | | | | | | | | | | | | | | | | | | | | | | | | | | | |
| Test of Moderators (coefficient 2): QM(df = 1) = 2.1279, p-val = 0.1446 | | | | | | | | | | | | | | | | | | | | | | | | | | | | | | | | | | |
| Model Results: | | | | | | | | | | | | | | | | | | | | | | | | | | | | | | | | | | |
|  | | | | | estimate | | | | | | | | se | | | | | | | zval | | | pval | | | | | | ci.lb | | | | | ci.ub |
| intrcpt | | | | | -0.5156 | | | | | | | | 0.4817 | | | | | | | -1.0703 | | | 0.2845 | | | | | | -1.4598 | | | | | 0.4286 |
| mean weight at baseline | | | | | 0.0073 | | | | | | | | 0.0050 | | | | | | | 1.4587 | | | 0.1446 | | | | | | -0.0025 | | | | | 0.0171 |
| **Vomiting events (proportion of males)** | | | | | | | | | | | | | | | | | | | | | | | | | | | | | | | | | | |
| Mixed-Effects Model (k = 24; tau^2 estimator: REML) | | | | | | | | | | | | | | | | | | | | | | | | | | | | | | | | | | |
| logLik | | | | | | | | | | deviance | | | | | | | AIC | | | | | | | | | BIC | | | | | | AICc | | |
| 4.4514 | | | | | | | | | | -8.9028 | | | | | | | -2.9028 | | | | | | | | | 0.3704 | | | | | | -1.5694 | | |
| tau^2 (estimated amount of residual heterogeneity): 0.0169 (SE = 0.0116); tau (square root of estimated tau^2 value): 0.1299;  I^2 (residual heterogeneity / unaccounted variability): 45.40%; H^2 (unaccounted variability / sampling variability): 1.83; R^2 (amount of heterogeneity accounted for): 0.00% | | | | | | | | | | | | | | | | | | | | | | | | | | | | | | | | | | |
| Test for Residual Heterogeneity: QE(df = 22) = 37.2003, p-val = 0.0225 | | | | | | | | | | | | | | | | | | | | | | | | | | | | | | | | | | |
| Test of Moderators (coefficient 2): QM(df = 1) = 0.0129, p-val = 0.9097 | | | | | | | | | | | | | | | | | | | | | | | | | | | | | | | | | | |
| Model Results: | | | | | | | | | | | | | | | | | | | | | | | | | | | | | | | | | | |
|  | | | | | estimate | | | | | | | | se | | | | | | | zval | | | pval | | | | | | ci.lb | | | | | ci.ub |
| intrcpt | | | | | 0.1834 | | | | | | | | 0.1727 | | | | | | | 1.0619 | | | 0.2883 | | | | | | -0.1551 | | | | | 0.5218 |
| proportion of males | | | | | -0.0460 | | | | | | | | 0.4055 | | | | | | | -0.1134 | | | 0.9097 | | | | | | -0.8407 | | | | | 0.7487 |
| **Vomiting events (trial’s follow-up)** | | | | | | | | | | | | | | | | | | | | | | | | | | | | | | | | | | |
| Mixed-Effects Model (k = 24; tau^2 estimator: REML) | | | | | | | | | | | | | | | | | | | | | | | | | | | | | | | | | | |
| logLik | | | | | | | | | | deviance | | | | | | | AIC | | | | | | | | | BIC | | | | | | AICc | | |
| 5.8239 | | | | | | | | | | -11.6478 | | | | | | | -5.6478 | | | | | | | | | -2.3747 | | | | | | -4.3145 | | |
| tau^2 (estimated amount of residual heterogeneity): 0.0110 (SE = 0.0096); tau (square root of estimated tau^2 value): 0.1048;  I^2 (residual heterogeneity / unaccounted variability): 34.51%; H^2 (unaccounted variability / sampling variability): 1.53; R^2 (amount of heterogeneity accounted for): 30.42% | | | | | | | | | | | | | | | | | | | | | | | | | | | | | | | | | | |
| Test for Residual Heterogeneity: QE(df = 22) = 29.7004, p-val = 0.1259 | | | | | | | | | | | | | | | | | | | | | | | | | | | | | | | | | | |
| Test of Moderators (coefficient 2): QM(df = 1) = 3.3499, p-val = 0.0672 | | | | | | | | | | | | | | | | | | | | | | | | | | | | | | | | | | |
| Model Results: | | | | | | | | | | | | | | | | | | | | | | | | | | | | | | | | | | |
|  | | | | | estimate | | | | | | | | se | | | | | | | zval | | | pval | | | | | | ci.lb | | | | | ci.ub |
| intrcpt | | | | | 0.0298 | | | | | | | | 0.0829 | | | | | | | 0.3599 | | | 0.7189 | | | | | | -0.1326 | | | | | 0.1923 |
| trial’s follow-up | | | | | 0.0147 | | | | | | | | 0.0081 | | | | | | | 1.8303 | | | 0.0672 | | | | | | -0.0010 | | | | | 0.0305 |
| **Diarrhea events (mean age at baseline)** | | | | | | | | | | | | | | | | | | | | | | | | | | | | | | | | | | |
| Mixed-Effects Model (k = 26; tau^2 estimator: REML) | | | | | | | | | | | | | | | | | | | | | | | | | | | | | | | | | | |
| logLik | | | | | | | | | | deviance | | | | | | | AIC | | | | | | | | | BIC | | | | | | AICc | | |
| -1.9584 | | | | | | | | | | 3.9169 | | | | | | | 9.9169 | | | | | | | | | 13.4510 | | | | | | 11.1169 | | |
| tau^2 (estimated amount of residual heterogeneity): 0.0029 (SE = 0.0061); tau (square root of estimated tau^2 value): 0.0541;  I^2 (residual heterogeneity / unaccounted variability): 12.20%; H^2 (unaccounted variability / sampling variability): 1.14; R^2 (amount of heterogeneity accounted for): 44.18% | | | | | | | | | | | | | | | | | | | | | | | | | | | | | | | | | | |
| Test for Residual Heterogeneity: QE(df = 24) = 44.4603, p-val = 0.0067 | | | | | | | | | | | | | | | | | | | | | | | | | | | | | | | | | | |
| Test of Moderators (coefficient 2): QM(df = 1) = 0.7791, p-val = 0.3774 | | | | | | | | | | | | | | | | | | | | | | | | | | | | | | | | | | |
| Model Results: | | | | | | | | | | | | | | | | | | | | | | | | | | | | | | | | | | |
|  | | | | | estimate | | | | | | | | se | | | | | | | zval | | | pval | | | | | | ci.lb | | | | | ci.ub |
| intrcpt | | | | | -0.1094 | | | | | | | | 0.3526 | | | | | | | -0.3103 | | | 0.7563 | | | | | | -0.8005 | | | | | 0.5817 |
| mean age at baseline | | | | | 0.0218 | | | | | | | | 0.0247 | | | | | | | 0.8827 | | | 0.3774 | | | | | | -0.0267 | | | | | 0.0703 |
| **Diarrhea events (mean BMI at baseline)** | | | | | | | | | | | | | | | | | | | | | | | | | | | | | | | | | | |
| Mixed-Effects Model (k = 24; tau^2 estimator: REML) | | | | | | | | | | | | | | | | | | | | | | | | | | | | | | | | | | |
| logLik | | | | | | | | | | deviance | | | | | | | AIC | | | | | | | | | BIC | | | | | | AICc | | |
| -0.6835 | | | | | | | | | | 1.3671 | | | | | | | 7.3671 | | | | | | | | | 10.6402 | | | | | | 8.7004 | | |
| tau^2 (estimated amount of residual heterogeneity): 0.0000 (SE = 0.0052); tau (square root of estimated tau^2 value): 0.0021;  I^2 (residual heterogeneity / unaccounted variability): 0.02%; H^2 (unaccounted variability / sampling variability): 1.00; R^2 (amount of heterogeneity accounted for): 99.92% | | | | | | | | | | | | | | | | | | | | | | | | | | | | | | | | | | |
| Test for Residual Heterogeneity: QE(df = 22) = 39.6704, p-val = 0.0118 | | | | | | | | | | | | | | | | | | | | | | | | | | | | | | | | | | |
| Test of Moderators (coefficient 2): QM(df = 1) = 4.9156, p-val = 0.0266 | | | | | | | | | | | | | | | | | | | | | | | | | | | | | | | | | | |
| Model Results: | | | | | | | | | | | | | | | | | | | | | | | | | | | | | | | | | | |
|  | | | | | estimate | | | | | | | | se | | | | | | | zval | | | pval | | | | | | ci.lb | | | | | ci.ub |
| intrcpt | | | | | -0.5071 | | | | | | | | 0.3172 | | | | | | | -1.5984 | | | 0.1100 | | | | | | -1.1289 | | | | | 0.1147 |
| mean BMI at baseline | | | | | 0.0208 | | | | | | | | 0.0094 | | | | | | | 2.2171 | | | 0.0266 | | | | | | 0.0024 | | | | | 0.0392 |
| **Diarrhea events (mean weight at baseline)** | | | | | | | | | | | | | | | | | | | | | | | | | | | | | | | | | | |
| Mixed-Effects Model (k = 22; tau^2 estimator: REML) | | | | | | | | | | | | | | | | | | | | | | | | | | | | | | | | | | |
| logLik | | | | | | | | | | deviance | | | | | | | AIC | | | | | | | | | BIC | | | | | | AICc | | |
| 2.6952 | | | | | | | | | | -5.3903 | | | | | | | 0.6097 | | | | | | | | | 3.5969 | | | | | | 2.1097 | | |
| tau^2 (estimated amount of residual heterogeneity): 0.0000 (SE = 0.0052); tau (square root of estimated tau^2 value): 0.0011;  I^2 (residual heterogeneity / unaccounted variability): 0.01%; H^2 (unaccounted variability / sampling variability): 1.00; R^2 (amount of heterogeneity accounted for): 99.90% | | | | | | | | | | | | | | | | | | | | | | | | | | | | | | | | | | |
| Test for Residual Heterogeneity: QE(df = 20) = 29.5434, p-val = 0.0776 | | | | | | | | | | | | | | | | | | | | | | | | | | | | | | | | | | |
| Test of Moderators (coefficient 2): QM(df = 1) = 6.5018, p-val = 0.0108 | | | | | | | | | | | | | | | | | | | | | | | | | | | | | | | | | | |
| Model Results: | | | | | | | | | | | | | | | | | | | | | | | | | | | | | | | | | | |
|  | | | | | estimate | | | | | | | | se | | | | | | | zval | | | pval | | | | | | ci.lb | | | | | ci.ub |
| intrcpt | | | | | -0.2870 | | | | | | | | 0.1864 | | | | | | | -1.5398 | | | 0.1236 | | | | | | -0.6524 | | | | | 0.0783 |
| mean weight at baseline | | | | | 0.0053 | | | | | | | | 0.0021 | | | | | | | 2.5499 | | | 0.0108 | | | | | | 0.0012 | | | | | 0.0093 |
| **Diarrhea events (proportion of males)** | | | | | | | | | | | | | | | | | | | | | | | | | | | | | | | | | | |
| Mixed-Effects Model (k = 28; tau^2 estimator: REML) | | | | | | | | | | | | | | | | | | | | | | | | | | | | | | | | | | |
| logLik | | | | | | | | | | deviance | | | | | | | AIC | | | | | | | | | BIC | | | | | | AICc | | |
| -0.1078 | | | | | | | | | | 0.2156 | | | | | | | 6.2156 | | | | | | | | | 9.9899 | | | | | | 7.3065 | | |
| tau^2 (estimated amount of residual heterogeneity): 0.0020 (SE = 0.0054); tau (square root of estimated tau^2 value): 0.0442;  I^2 (residual heterogeneity / unaccounted variability): 8.76%; H^2 (unaccounted variability / sampling variability): 1.10; R^2 (amount of heterogeneity accounted for): 17.96% | | | | | | | | | | | | | | | | | | | | | | | | | | | | | | | | | | |
| Test for Residual Heterogeneity: QE(df = 26) = 45.3695, p-val = 0.0107 | | | | | | | | | | | | | | | | | | | | | | | | | | | | | | | | | | |
| Test of Moderators (coefficient 2): QM(df = 1) = 1.0166, p-val = 0.3133 | | | | | | | | | | | | | | | | | | | | | | | | | | | | | | | | | | |
| Model Results: | | | | | | | | | | | | | | | | | | | | | | | | | | | | | | | | | | |
|  | | estimate | | | | | | | | | | | se | | | | | | | zval | | | pval | | | | | | ci.lb | | | | | ci.ub |
| intrcpt | | 0.3474 | | | | | | | | | | | 0.1578 | | | | | | | 2.2020 | | | 0.0277 | | | | | | 0.0382 | | | | | 0.6566 |
| proportion of males | | -0.3860 | | | | | | | | | | | 0.3828 | | | | | | | -1.0083 | | | 0.3133 | | | | | | -1.1362 | | | | | 0.3643 |
| **Diarrhea events (trial’s follow-up)** | | | | | | | | | | | | | | | | | | | | | | | | | | | | | | | | | | |
| Mixed-Effects Model (k = 28; tau^2 estimator: REML) | | | | | | | | | | | | | | | | | | | | | | | | | | | | | | | | | | |
| logLik | | | | | | | | | | deviance | | | | | | | AIC | | | | | | | | | BIC | | | | | | AICc | | |
| 5.5606 | | | | | | | | | | -11.1212 | | | | | | | -5.1212 | | | | | | | | | -1.3469 | | | | | | -4.0303 | | |
| tau^2 (estimated amount of residual heterogeneity): 0.0000 (SE = 0.0047); tau (square root of estimated tau^2 value): 0.0003;  I^2 (residual heterogeneity / unaccounted variability): 0.00%; H^2 (unaccounted variability / sampling variability): 1.00; R^2 (amount of heterogeneity accounted for): 100.00% | | | | | | | | | | | | | | | | | | | | | | | | | | | | | | | | | | |
| Test for Residual Heterogeneity: QE(df = 26) = 33.7011, p-val = 0.1427 | | | | | | | | | | | | | | | | | | | | | | | | | | | | | | | | | | |
| Test of Moderators (coefficient 2): QM(df = 1) = 12.8436, p-val = 0.0003 | | | | | | | | | | | | | | | | | | | | | | | | | | | | | | | | | | |
| Model Results: | | | | | | | | | | | | | | | | | | | | | | | | | | | | | | | | | | |
|  | | estimate | | | | | | | | | | | se | | | | | | | zval | | | pval | | | | | | ci.lb | | | | | ci.ub |
| intrcpt | | 0.0060 | | | | | | | | | | | 0.0574 | | | | | | | 0.1047 | | | 0.9166 | | | | | | -0.1064 | | | | | 0.1185 |
| trial’s follow-up | | 0.0208 | | | | | | | | | | | 0.0058 | | | | | | | 3.5838 | | | 0.0003 | | | | | | 0.0094 | | | | | 0.0322 |

### Appendix 10: Results of sensitivity analyses

**Note:** LMA, Lifestyle modification alone; GLP-1, glucagon-like peptide-1; Metformin_Fluoxetine, Metformin combined with Fluoxetine.

**Exclusion 1:** exclusion of studies with fewer than 40 participants

**Exclusion 2:** exclusion of studies without a placebo-control

**Exclusion 3:** exclusion of studies that did not report BMI change from baseline

**Exclusion 4:** exclusion of studies with treatment duration < 3 months

**Exclusion 5:** exclusion of the high risk of bias studies

| **Comparisons** | **Main estimates** | **Bayesian estimates** | **Exclusion 1 (P)** | **Exclusion 2 (I/C)** | **Exclusion 3 (O)** | **Exclusion 4 (T)** | **Exclusion 5 (R)** |
| --- | --- | --- | --- | --- | --- | --- | --- |
| **Outcome: change in BMI from baseline** | | | | | | | |
| Fluoxetine vs GLP-1 receptor agonists | 0.11 (-2.57, 2.79) | 0.04 (-2.05, 2.19) | 0.65 (-2.34, 3.65) | 0.15 (-1.84, 2.13) | - | 0.45 (-2.39, 3.30) | 0.11 (-2.56, 2.77) |
| Fluoxetine vs LMA | -1.70 (-4.00, 0.60) | -1.67 (-3.49, 0.11) | -1.65 (-3.97, 0.67) | -1.57 (-3.24, 0.10) | - | -1.70 (-4.04, 0.64) | -1.70 (-3.98, 0.59) |
| Fluoxetine vs Metformin | -0.60 (-2.90, 1.70) | -0.62 (-2.42, 1.19) | -0.65 (-2.97, 1.68) | -0.73 (-2.40, 0.94) | - | -0.60 (-2.94, 1.74) | -0.60 (-2.89, 1.68) |
| Fluoxetine vs Metformin_Fluoxetine | 0.30 (-2.32, 2.92) | 0.3 (-1.76, 2.35) | 0.30 (-2.34, 2.94) | 0.30 (-1.60, 2.20) | - | 0.30 (-2.37, 2.97) | 0.30 (-2.31, 2.91) |
| Fluoxetine vs Orlistat | -0.03 (-2.82, 2.75) | -0.13 (-2.3, 2.12) | 0.01 (-2.79, 2.82) | -0.66 (-3.22, 1.89) | - | 0.52 (-2.55, 3.58) | -0.92 (-3.83, 2.00) |
| Fluoxetine vs Phentermine-Topiramate | 3.13 (-0.36, 6.63) | 3.16 (0.4, 5.88) | 3.18 (-0.34, 6.70) | 3.26 ( 0.72, 5.80) | - | 3.13 (-0.42, 6.69) | 3.13 (-0.34, 6.60) |
| Fluoxetine vs Sitagliptin | -2.40 (-5.94, 1.14) | -2.38 (-5.18, 0.4) | -2.35 (-5.92, 1.21) | -2.27 (-4.87, 0.33) | - | -2.40 (-6.00, 1.20) | -2.40 (-5.92, 1.12) |
| Fluoxetine vs Topiramate | -0.84 (-4.57, 2.89) | -0.82 (-3.85, 2.2) | - | -0.71 (-3.57, 2.15) | - | -0.84 (-4.63, 2.95) | -0.84 (-4.55, 2.87) |
| GLP-1 receptor agonists vs LMA | -1.81 (-3.19, -0.43) | -1.72 (-2.88, -0.62) | -2.31 (-4.20, -0.42) | -1.72 (-2.78, -0.65) | - | -2.15 (-3.77, -0.54) | -1.81 (-3.18, -0.43) |
| GLP-1 receptor agonists vs Metformin | -0.71 (-2.25, 0.83) | -0.67 (-1.94, 0.58) | -1.30 (-3.36, 0.76) | -0.88 (-2.10, 0.34) | - | -1.05 (-2.82, 0.71) | -0.71 (-2.25, 0.83) |
| GLP-1 receptor agonists vs Metformin_Fluoxetine | 0.19 (-2.49, 2.87) | 0.25 (-1.89, 2.36) | -0.35 (-3.35, 2.64) | 0.15 (-1.83, 2.14) | - | -0.15 (-3.00, 2.69) | 0.19 (-2.47, 2.86) |
| GLP-1 receptor agonists vs Orlistat | -0.14 (-2.23, 1.95) | -0.17 (-1.86, 1.54) | -0.64 (-3.10, 1.82) | -0.81 (-3.01, 1.39) | - | 0.06 (-2.50, 2.62) | -1.02 (-3.30, 1.25) |
| GLP-1 receptor agonists vs Phentermine-Topiramate | 3.02 ( 0.05, 5.99) | 3.11 (0.72, 5.43) | 2.52 (-0.73, 5.77) | 3.11 ( 0.93, 5.30) | - | 2.68 (-0.45, 5.80) | 3.02 ( 0.07, 5.98) |
| GLP-1 receptor agonists vs Sitagliptin | -2.51 (-5.53, 0.51) | -2.42 (-4.88, -0.04) | -3.01 (-6.31, 0.29) | -2.42 (-4.67, -0.16) | - | -2.85 (-6.03, 0.32) | -2.51 (-5.51, 0.50) |
| GLP-1 receptor agonists vs Topiramate | -0.95 (-4.20, 2.30) | -0.86 (-3.57, 1.8) | - | -0.86 (-3.41, 1.69) | - | -1.29 (-4.68, 2.10) | -0.95 (-4.18, 2.29) |
| Metformin vs LMA | -1.10 (-1.79, -0.41) | -1.06 (-1.64, -0.51) | -1.01 (-1.82, -0.19) | -0.84 (-1.43, -0.24) | - | -1.10 (-1.80, -0.40) | -1.10 (-1.79, -0.41) |
| Metformin vs Metformin_Fluoxetine | 0.90 (-1.40, 3.20) | 0.92 (-0.89, 2.72) | 0.95 (-1.37, 3.27) | 1.03 (-0.64, 2.70) | - | 0.90 (-1.44, 3.24) | 0.90 (-1.38, 3.19) |
| Metformin vs Orlistat | 0.57 (-1.15, 2.28) | 0.49 (-0.87, 1.91) | 0.66 (-1.12, 2.43) | 0.07 (-1.95, 2.08) | - | 1.12 (-0.99, 3.22) | -0.32 (-2.25, 1.62) |
| Metformin vs Phentermine-Topiramate | 3.73 ( 1.01, 6.45) | 3.78 (1.62, 5.9) | 3.82 ( 1.06, 6.59) | 3.99 ( 1.99, 5.99) | - | 3.73 ( 0.96, 6.50) | 3.73 ( 1.03, 6.44) |
| Metformin vs Sitagliptin | -1.80 (-4.57, 0.98) | -1.76 (-3.98, 0.43) | -1.71 (-4.53, 1.12) | -1.54 (-3.61, 0.54) | - | -1.80 (-4.62, 1.02) | -1.80 (-4.56, 0.96) |
| Metformin vs Topiramate | -0.24 (-3.26, 2.78) | -0.19 (-2.71, 2.29) | - | 0.02 (-2.37, 2.41) | - | -0.24 (-3.30, 2.82) | -0.24 (-3.24, 2.77) |
| Metformin_Fluoxetine vs LMA | -2.00 (-4.30, 0.30) | -1.98 (-3.8, -0.19) | -1.95 (-4.27, 0.37) | -1.87 (-3.54, -0.20) | - | -2.00 (-4.34, 0.34) | -2.00 (-4.28, 0.29) |
| Metformin_Fluoxetine vs Orlistat | -0.33 (-3.12, 2.45) | -0.43 (-2.59, 1.81) | -0.29 (-3.09, 2.52) | -0.96 (-3.52, 1.59) | - | 0.22 (-2.85, 3.28) | -1.22 (-4.13, 1.70) |
| Metformin_Fluoxetine vs Phentermine-Topiramate | 2.83 (-0.66, 6.33) | 2.86 (0.1, 5.58) | 2.88 (-0.64, 6.40) | 2.96 ( 0.42, 5.50) | - | 2.83 (-0.72, 6.38) | 2.83 (-0.64, 6.30) |
| Metformin_Fluoxetine vs Sitagliptin | -2.70 (-6.24, 0.84) | -2.68 (-5.48, 0.1) | -2.65 (-6.22, 0.91) | -2.57 (-5.17, 0.03) | - | -2.70 (-6.30, 0.90) | -2.70 (-6.22, 0.82) |
| Metformin_Fluoxetine vs Topiramate | -1.14 (-4.87, 2.59) | -1.12 (-4.16, 1.9) | - | -1.01 (-3.87, 1.85) | - | -1.14 (-4.93, 2.65) | -1.14 (-4.85, 2.57) |
| Orlistat vs LMA | -1.66 (-3.23, -0.09) | -1.55 (-2.87, -0.32) | -1.67 (-3.24, -0.09) | -0.91 (-2.83, 1.02) | - | -2.22 (-4.20, -0.23) | -0.78 (-2.60, 1.03) |
| Orlistat vs Phentermine-Topiramate | 3.17 ( 0.10, 6.23) | 3.28 (0.81, 5.66) | 3.16 ( 0.08, 6.24) | 3.92 ( 1.21, 6.64) | - | 2.61 (-0.72, 5.94) | 4.05 ( 0.87, 7.23) |
| Orlistat vs Sitagliptin | -2.36 (-5.48, 0.75) | -2.25 (-4.79, 0.19) | -2.37 (-5.50, 0.76) | -1.61 (-4.38, 1.16) | - | -2.92 (-6.29, 0.46) | -1.48 (-4.71, 1.75) |
| Orlistat vs Topiramate | -0.80 (-4.14, 2.53) | -0.69 (-3.48, 2.02) | - | -0.05 (-3.06, 2.97) | - | -1.36 (-4.94, 2.22) | 0.08 (-3.36, 3.52) |
| Phentermine-Topiramate vs LMA | -4.83 (-7.46, -2.20) | -4.83 (-6.89, -2.77) | -4.83 (-7.48, -2.18) | -4.83 (-6.74, -2.92) | - | -4.83 (-7.51, -2.15) | -4.83 (-7.44, -2.22) |
| Phentermine-Topiramate vs Sitagliptin | -5.53 (-9.29, -1.77) | -5.53 (-8.51, -2.57) | -5.53 (-9.31, -1.75) | -5.53 (-8.29, -2.77) | - | -5.53 (-9.36, -1.70) | -5.53 (-9.27, -1.79) |
| Phentermine-Topiramate vs Topiramate | -3.97 (-7.91, -0.03) | -3.97 (-7.16, -0.79) | - | -3.97 (-6.97, -0.97) | - | -3.97 (-7.98, 0.04) | -3.97 (-7.89, -0.05) |
| Sitagliptin vs LMA | 0.70 (-1.99, 3.39) | 0.7 (-1.43, 2.84) | 0.70 (-2.00, 3.40) | 0.70 (-1.29, 2.69) | - | 0.70 (-2.03, 3.43) | 0.70 (-1.97, 3.37) |
| Sitagliptin vs Topiramate | 1.56 (-2.42, 5.54) | 1.56 (-1.67, 4.8) | - | 1.56 (-1.49, 4.61) | - | 1.56 (-2.48, 5.60) | 1.56 (-2.40, 5.52) |
| Topiramate vs LMA | -0.86 (-3.80, 2.08) | -0.86 (-3.29, 1.57) | - | -0.86 (-3.18, 1.46) | - | -0.86 (-3.84, 2.12) | -0.86 (-3.79, 2.07) |
| **Outcome: change in weight from baseline** | | | | | | | |
| GLP-1 receptor agonists vs LMA | -3.82 ( -6.47, -1.16) | -3.83 (-6.87, -0.83) | -4.49 ( -8.28, -0.70) | -3.82 ( -6.49, -1.15) | -3.82 ( -6.57, -1.08) | -4.35 ( -7.63, -1.07) | -3.80 ( -6.30, -1.30) |
| GLP-1 receptor agonists vs Metformin | -1.25 ( -4.38, 1.89) | -1.24 (-4.82, 2.35) | -1.44 ( -5.82, 2.94) | -1.30 ( -4.50, 1.91) | -1.28 ( -4.58, 2.02) | -1.79 ( -5.59, 2.02) | -1.25 ( -4.19, 1.68) |
| GLP-1 receptor agonists vs Orlistat | 0.46 ( -3.42, 4.35) | 0.48 (-3.9, 5.02) | -0.14 ( -5.02, 4.75) | -1.06 ( -6.18, 4.07) | 0.49 ( -3.54, 4.52) | 1.11 ( -3.92, 6.15) | -1.33 ( -5.22, 2.55) |
| GLP-1 receptor agonists vs Phentermine-Topiramate | 10.77 (5.30, 16.24) | 10.76 (4.24, 17.27) | 10.10 (3.59, 16.62) | 10.77 ( 5.26, 16.29) | 10.77 ( 5.05, 16.49) | 10.24 ( 3.94, 16.54) | 10.79 ( 5.77, 15.81) |
| GLP-1 receptor agonists vs Sitagliptin | -5.62 (-11.20, -0.03) | -5.63 (-12.26, 0.98) | -6.29 (-12.90, 0.32) | -5.62 (-11.24, 0.01) | -5.62 (-11.45, 0.20) | -6.15 (-12.54, 0.25) | -5.60 (-10.74, -0.46) |
| GLP-1 receptor agonists vs Topiramate | -1.39 ( -8.04, 5.26) | -1.39 (-8.92, 6.12) | - | -1.39 ( -8.07, 5.30) | -1.39 ( -8.25, 5.46) | -1.92 ( -9.27, 5.43) | -1.37 ( -7.66, 4.91) |
| Metformin vs LMA | -2.57 ( -4.23, -0.91) | -2.58 (-4.54, -0.7) | -3.05 ( -5.25, -0.85) | -2.52 ( -4.30, -0.74) | -2.54 ( -4.37, -0.72) | -2.56 ( -4.49, -0.63) | -2.55 ( -4.09, -1.00) |
| Metformin vs Orlistat | 1.71 (-1.58, 5.00) | 1.72 (-2.02, 5.6) | 1.30 ( -2.48, 5.09) | 0.24 ( -4.48, 4.96) | 1.77 ( -1.70, 5.24) | 2.90 ( -1.38, 7.18) | -0.08 ( -3.44, 3.28) |
| Metformin vs Phentermine-Topiramate | 12.02 (6.96, 17.08) | 12.01 (5.87, 18.05) | 11.54 (5.80, 17.28) | 12.07 ( 6.93, 17.21) | 12.05 ( 6.71, 17.39) | 12.03 ( 6.32, 17.74) | 12.04 ( 7.42, 16.66) |
| Metformin vs Sitagliptin | -4.37 ( -9.55, 0.81) | -4.39 (-10.61, 1.77) | -4.85 (-10.70, 0.99) | -4.32 ( -9.58, 0.94) | -4.34 ( -9.80, 1.11) | -4.36 (-10.18, 1.46) | -4.35 ( -9.10, 0.40) |
| Metformin vs Topiramate | -0.14 ( -6.46, 6.18) | -0.14 (-7.33, 6.97) | - | -0.09 ( -6.47, 6.29) | -0.11 ( -6.66, 6.43) | -0.13 ( -6.98, 6.72) | -0.12 ( -6.09, 5.85) |
| Orlistat vs LMA | -4.28 ( -7.12, -1.44) | -4.3 (-7.7, -1.11) | -4.35 ( -7.43, -1.27) | -2.76 ( -7.13, 1.61) | -4.32 ( -7.26, -1.37) | -5.46 ( -9.28, -1.64) | -2.47 ( -5.45, 0.51) |
| Orlistat vs Phentermine-Topiramate | 10.31 (4.75, 15.87) | 10.29 (3.52, 16.84) | 10.24 (4.11, 16.37) | 11.83 ( 5.32, 18.34) | 10.27 ( 4.45, 16.10) | 9.13 ( 2.53, 15.72) | 12.12 ( 6.85, 17.40) |
| Orlistat vs Sitagliptin | -6.08 (-11.75, -0.41) | -6.11 (-12.96, 0.55) | -6.15 (-12.38, 0.08) | -4.56 (-11.16, 2.05) | -6.12 (-12.04, -0.19) | -7.26 (-13.95, -0.57) | -4.27 ( -9.66, 1.12) |
| Orlistat vs Topiramate | -1.85 ( -8.57, 4.88) | -1.87 (-9.6, 5.67) | - | -0.33 ( -7.86, 7.20) | -1.89 ( -8.83, 5.06) | -3.03 (-10.63, 4.57) | -0.04 ( -6.53, 6.45) |
| Phentermine-Topiramate vs LMA | -14.59 (-19.37, -9.81) | -14.59 (-20.38, -8.81) | -14.59 (-19.89, -9.29) | -14.59 (-19.41, -9.77) | -14.59 (-19.61, -9.57) | -14.59 (-19.97, -9.21) | -14.59(-18.94, -10.24) |
| Phentermine-Topiramate vs Sitagliptin | -16.39 (-23.24, -9.54) | -16.39 (-24.64, -8.12) | -16.39 (-23.97, -8.81) | -16.39 (-23.30, -9.48) | -16.39 (-23.57, -9.21) | -16.39 (-24.07, -8.71) | -16.39(-22.65, -10.13) |
| Phentermine-Topiramate vs Topiramate | -12.16 (-19.91, -4.41) | -12.15 (-21.12, -3.18) | - | -12.16 (-19.96, -4.36) | -12.16 (-20.20, -4.12) | -12.16 (-20.65, -3.67) | -12.16 (-19.38, -4.94) |
| Sitagliptin vs LMA | 1.80 (-3.11, 6.71) | 1.8 (-4.08, 7.69) | 1.80 ( -3.62, 7.22) | 1.80 ( -3.15, 6.75) | 1.80 ( -3.34, 6.94) | 1.80 ( -3.69, 7.29) | 1.80 (-2.69, 6.29) |
| Sitagliptin vs Topiramate | 4.23 (-3.60, 12.06) | 4.25 (-4.81, 13.27) | - | 4.23 ( -3.65, 12.11) | 4.23 ( -3.89, 12.35) | 4.23 ( -4.33, 12.79) | 4.23 (-3.08, 11.54) |
| Topiramate vs LMA | -2.43 (-8.53, 3.67) | -2.44 (-9.32, 4.46) | - | -2.43 ( -8.56, 3.70) | -2.43 ( -8.71, 3.85) | -2.43 ( -9.00, 4.14) | -2.43 (-8.20, 3.34) |
| **Outcome: percentage of participants achieving BMI reduction of at least 5%** | | | | | | | |
| GLP-1 receptor agonists vs LMA | 4.86 (1.93, 12.24) | 5.1 (0.92, 24.53) | 4.86 (1.93, 12.24) | 4.86 (1.93, 12.24) | 3.65 (2.01, 6.63) | 4.86 (1.93, 12.24) | 4.86 (1.93, 12.24) |
| GLP-1 receptor agonists vs Orlistat | 2.58 (0.48, 13.90) | 2.71 (0.1, 60.5) | 2.58 (0.48, 13.90) | 2.58 (0.48, 13.90) | - | 2.58 (0.48, 13.90) | 2.58 (0.48, 13.90) |
| GLP-1 receptor agonists vs Phentermine-Topiramate | 0.35 (0.05, 2.60) | 0.31 (0.01, 8.22) | 0.35 (0.05,2.60) | 0.35 (0.05, 2.60) | 0.26 (0.07, 0.99) | 0.35 (0.05, 2.60) | 0.35 (0.05, 2.60) |
| GLP-1 receptor agonists vs Topiramate | 2.43 (0.20, 28.96) | 2.23 (0.05, 79.64) | - | 2.43 (0.20, 28.96) | 1.83 (0.25, 13.08) | 2.43 (0.20, 28.96) | 2.43 (0.20, 28.96) |
| Orlistat vs LMA | 1.88 (0.46, 7.68) | 1.89 (0.12, 29.16) | 1.88 (0.46,7.68) | 1.88 (0.46, 7.68) | - | 1.88 (0.46, 7.68) | 1.88 (0.46, 7.68) |
| Orlistat vs Phentermine-Topiramate | 0.13 (0.01, 1.31) | 0.116 (0.002, 6.304) | 0.13 (0.01,1.31) | 0.13 (0.01, 1.31) | - | 0.13 (0.01, 1.31) | 0.13 (0.01, 1.31) |
| Orlistat vs Topiramate | 0.94 (0.06, 13.94) | 0.83 (0.01, 57.3) | - | 0.94 (0.06, 13.94) | - | 0.94 (0.06, 13.94) | 0.94 (0.06, 13.94) |
| Phentermine-Topiramate vs LMA | 14.06 (2.34, 84.35) | 16.12 (0.89, 317.28) | 14.06 (2.34, 84.35) | 14.06 (2.34, 84.35) | 14.06 (4.22, 46.79) | 14.06 (2.34, 84.35) | 14.06 (2.34, 84.35) |
| Phentermine-Topiramate vs Topiramate | 7.03 (0.38, 129.65) | 7.18 (0.08, 575.44) | - | 7.03 (0.38, 129.65) | 7.03 (0.76, 65.29) | 7.03 (0.38, 129.65) | 7.03 (0.38, 129.65) |
| Topiramate vs LMA | 2.00 (0.20, 19.93) | 2.26 (0.09, 67.38) | - | 2.00 (0.20, 19.93) | 2.00 (0.31, 13.06) | 2.00 (0.20, 19.93) | 2.00 (0.20, 19.93) |
| **Outcome: percentage of participants achieving BMI reduction of at least 10%** | | | | | | | |
| GLP-1 receptor agonists vs LMA | 5.1996(1.4400, 18.7747) | 5.8014 (0.3395, 72.1668) | 5.1996 (1.4400, 18.7747) | 5.1996 (1.4400, 18.7747) | 4.6630 (2.1247, 10.2337) | 5.1996(1.4400, 18.7747) | 5.1996 (1.4400, 18.7747) |
| GLP-1 receptor agonists vs Orlistat | 1.5995(0.1418, 18.0367) | 1.7152 (0.0074, 277.6411) | 1.5995 (0.1418, 18.0367) | 1.5995 (0.1418, 18.0367) | - | 1.5995(0.1418, 18.0367) | 1.5995 (0.1418, 18.0367) |
| GLP-1 receptor agonists vs Phentermine-Topiramate | 0.0720(0.0019, 2.6937) | 0 (0, 0.1162) | 0.0720 (0.0019,2.6937) | 0.0720 (0.0019, 2.6937) | 0.0646 (0.0035, 1.1848) | 0.0720(0.0019, 2.6937) | 0.0720 (0.0019, 2.6937) |
| GLP-1 receptor agonists vs Topiramate | 5.9995(0.1521, 236.6757) | 6.6696 (0.0112, 3024.7293) | - | 5.9995 (0.1521, 236.6757) | 5.3804 (0.2746, 105.4310) | 5.9995(0.1521, 236.6757) | 5.9995 (0.1521, 236.6757) |
| Orlistat vs LMA | 3.2508(0.4166, 25.3670) | 3.4118 (0.0365, 323.0711) | 3.2508 (0.4166, 25.3670) | 3.2508 (0.4166, 25.3670) | - | 3.2508(0.4166, 25.3670) | 3.2508 (0.4166, 25.3670) |
| Orlistat vs Phentermine-Topiramate | 0.0450(0.0009, 2.3644) | 0 (0, 0.1007) | 0.0450 (0.0009,2.3644) | 0.0450 (0.0009, 2.3644) | - | 0.0450(0.0009, 2.3644) | 0.0450 (0.0009, 2.3644) |
| Orlistat vs Topiramate | 3.7509(0.0680, 206.8080) | 4.0658 (0.0029, 5219.9707) | - | 3.7509 (0.0680, 206.8080) | - | 3.7509(0.0680, 206.8080) | 3.7509 (0.0680, 206.8080) |
| Phentermine-Topiramate vs LMA | 72.2098(2.4421, 2135.1130) | 227912516528.795 (59.1108, 1.32902251362776e+40) | 72.2098 (2.4421, 2135.1130) | 72.2098 (2.4421, 2135.1130) | 72.2098 (4.3856, 1188.9561) | 72.2098(2.4421, 2135.1130) | 72.2098 (2.4421, 2135.1130) |
| Phentermine-Topiramate vs Topiramate | 83.3189(0.6656, 10430.3372) | 288981746135.12 (21.5022, 2.10413682194129e+40) | - | 83.3189 (0.6656, 10430.3372) | 83.3189 (1.5106, 4595.6076) | 83.3189(0.6656, 10430.3372) | 83.3189 (0.6656, 10430.3372) |
| Topiramate vs LMA | 0.8667(0.0277, 27.1216) | 0.8444 (0.003, 252.9043) | - | 0.8667 (0.0277, 27.1216) | 0.8667 (0.0492, 15.2791) | 0.8667(0.0277, 27.1216) | 0.8667 (0.0277, 27.1216) |
| **Outcome: change in BMI z-score from baseline** | | | | | | | |
| Metformin vs LMA | -0.16 (-0.21, -0.10) | -0.15 (-0.35, 0.05) | -0.15 (-0.24, -0.06) | -0.15 (-0.24, -0.06) | -0.15 (-0.22, -0.08) | -0.16 (-0.21, -0.10) | -0.16 (-0.21, -0.10) |
| Metformin vs Orlistat | -0.11 (-0.17, -0.05) | -0.10 (-0.5, 0.29) | -0.10 (-0.23, 0.03) | - | -0.10 (-0.20, 0.00) | - | -0.11 (-0.17, -0.05) |
| Metforminvs Topiramate | -0.13 (-0.22, -0.04) | -0.12 (-0.52, 0.28) | - | -0.12 (-0.26, 0.02) | -0.12 (-0.24, 0.00) | -0.13 (-0.22, -0.04) | -0.13 (-0.22, -0.04) |
| Orlistat vs LMA | -0.05 (-0.08, -0.02) | -0.05 (-0.39, 0.29) | -0.05 (-0.14, 0.04) | - | -0.05 (-0.12, 0.02) | - | -0.05 (-0.08, -0.02) |
| Orlistat vs Topiramate | -0.02 (-0.10, 0.06) | -0.02 (-0.51, 0.47) | - | - | -0.02 (-0.14, 0.10) | - | -0.02 (-0.10, 0.06) |
| Topiramate vs LMA | -0.03 (-0.10, 0.04) | -0.03 (-0.38, 0.32) | - | -0.03 (-0.14, 0.08) | -0.03 (-0.13, 0.07) | -0.03 (-0.10, 0.04) | -0.03 (-0.10, 0.04) |
| **Outcome: change in BMI SDS from baseline** | | | | | | | |
| GLP-1 receptor agonists vs LMA | -0.15 (-0.25, -0.04) | -0.15 (-0.31, -0.01) | -0.14 (-0.22, -0.06) | -0.15 (-0.25, -0.04) | -0.14 (-0.23, -0.06) | -0.15 (-0.25, -0.04) | -0.15 (-0.25, -0.04) |
| GLP-1 receptor agonists vs Metformin | -0.06 (-0.19, 0.06) | -0.06 (-0.23, 0.14) | -0.07 (-0.16, 0.02) | -0.06 (-0.19, 0.06) | -0.07 (-0.17, 0.02) | -0.06 (-0.19, 0.06) | -0.06 (-0.19, 0.06) |
| GLP-1 receptor agonists vs Orlistat | -0.09 (-0.25, 0.07) | -0.09 (-0.36, 0.16) | -0.08 (-0.19, 0.03) | -0.09 (-0.25, 0.07) | -0.08 (-0.20, 0.04) | -0.09 (-0.25, 0.07) | -0.09 (-0.25, 0.07) |
| Metformin vs LMA | -0.09 (-0.15, -0.02) | -0.08 (-0.22, -0.01) | -0.07 (-0.11, -0.03) | -0.09 (-0.15, -0.02) | -0.07 (-0.11, -0.02) | -0.09 (-0.15, -0.02) | -0.09 (-0.15, -0.02) |
| Metformin vs Orlistat | -0.03 (-0.16, 0.11) | -0.02 (-0.3, 0.18) | -0.01 (-0.09, 0.08) | -0.03 (-0.16, 0.11) | -0.01 (-0.10, 0.08) | -0.03 (-0.16, 0.11) | -0.03 (-0.16, 0.11) |
| Orlistat vs LMA | -0.06 (-0.18, 0.06) | -0.06 (-0.28, 0.16) | -0.06 (-0.13, 0.01) | -0.06 (-0.18, 0.06) | -0.06 (-0.14, 0.02) | -0.06 (-0.18, 0.06) | -0.06 (-0.18, 0.06) |
| **Outcome: total gastrointestinal adverse events** | | | | | | | |
| GLP-1 receptor agonists vs LMA | 2.97 (1.93, 4.55) | 3.12 (1.91, 5.79) | 2.83 (1.80, 4.46) | 2.91 (2.06, 4.11) | 3.31 (1.60, 6.84) | 2.82 ( 1.97, 4.03) | 2.91 (2.06, 4.11) |
| GLP-1 receptor agonists vs Metformin | 2.11 (1.10, 4.04) | 2.08 (0.96, 4.65) | 2.22 (1.12, 4.41) | 2.06 (1.18, 3.61) | 2.63 (1.02, 6.76) | 2.00 ( 1.14, 3.52) | 2.06 (1.18, 3.61) |
| GLP-1 receptor agonists vs Orlistat | 0.07 (0.01, 0.74) | 0 (0, 2e-06) | 0.06 (0.01, 0.71) | - | 0.07 (0.01, 0.90) | 0.00153 (0.00003 0.08) | 0.51 (0.03, 9.88) |
| GLP-1 receptor agonists vs Phentermine-Topiramate | 3.18 (1.12, 9.05) | 2.7 (0.68, 8.72) | 3.03 (1.05, 8.75) | 3.23 (1.29, 8.10) | 4.30 (1.12, 16.46) | 3.66 ( 1.41, 9.54) | 3.23 (1.29, 8.10) |
| Metformin vs LMA | 1.41 (0.86, 2.30) | 1.5 (0.87, 2.77) | 1.27 (0.76, 2.13) | 1.41 (0.91, 2.19) | 1.26 (0.69, 2.30) | 1.41 ( 0.91, 2.19) | 1.41 (0.91, 2.19) |
| Metformin vs Orlistat | 0.031(0.003, 0.357) | 0 (0, 1e-06) | 0.028 (0.002, 0.325) | - | 0.03 (0.002, 0.33) | 0.00076 (0.00001, 0.04) | 0.25 (0.01, 4.85) |
| Metformin vs Phentermine-Topiramate | 1.51 (0.52, 4.41) | 1.29 (0.32, 4.18) | 1.37 (0.46, 4.04) | 1.57 (0.60, 4.08) | 1.63 (0.45, 5.88) | 1.83 ( 0.68, 4.93) | 1.57 (0.60, 4.08) |
| Orlistat vs LMA | 45.15 (4.14, 492.24) | 9.91372297712241e+32 (131496896.26, 1.38737701725904e+59) | 45.17 (4.14, 492.80) | - | 45.94 (4.09, 516.06) | 1845.00 (34.98, 97302.39) | 5.71 (0.30, 108.40) |
| Orlistat vs Phentermine-Topiramate | 48.39 (3.69, 633.79) | 8.44516260829596e+32 (111030697.82, 1.09158530574297e+59) | 48.36 (3.69, 634.51) | - | 59.64 (4.13, 861.14) | 2395.26 (41.17, 139355.29) | 6.34 (0.30, 135.78) |
| Phentermine-Topiramate vs LMA | 0.93 (0.36, 2.42) | 1.16 (0.41, 4.36) | 0.93 (0.36, 2.43) | 0.90 (0.38, 2.11) | 0.77 (0.25, 2.39) | 0.77 ( 0.32, 1.87) | 0.90 (0.38, 2.11) |
| **Outcome: discontinuation due to adverse events** | | | | | | | |
| GLP-1 receptor agonists vs LMA | 2.03 (0.76, 5.42) | 6.79 (0.82, 108.44) | 2.06 (0.76, 5.58) | 1.96 (0.77, 5.02) | 11.41 (1.35, 96.73) | 1.96 (0.77, 4.98) | 1.96 (0.77, 5.02) |
| GLP-1 receptor agonists vs Metformin | 1.06 (0.25, 4.44) | 1.97 (0.08, 59.38) | 1.35 (0.25, 7.34) | 1.03 (0.26, 4.11) | 8.31 (0.75, 92.72) | 1.03 (0.26, 4.07) | 1.03 (0.26, 4.11) |
| GLP-1 receptor agonists vs Orlistat | 0.60 (0.13, 2.79) | 0.366 (0.004, 18.888) | 0.60 (0.12, 2.87) | 0.82 (0.18, 3.74) | 1.05 (0.05, 21.53) | 0.61 (0.14, 2.57) | 0.82 (0.18, 3.74) |
| GLP-1 receptor agonists vs Phentermine-Topiramate | 4.23 (0.47, 37.79) | 6.38 (0.05, 702.86) | 4.26 (0.46, 39.02) | 4.15 (0.49, 34.93) | 70.16 (2.78, 1771.36) | 12.04 (0.90, 161.11) | 4.15 (0.49, 34.93) |
| GLP-1 receptor agonists vs Sitagliptin | 0.66 (0.05, 8.54) | 1.62 (0.01, 591.58) | 0.67 (0.05, 8.95) | 0.64 (0.05, 7.60) | 3.72 (0.16, 84.83) | 0.64 (0.05, 7.52) | 0.64 (0.05, 7.60) |
| Metformin vs LMA | 1.92 (0.68, 5.43) | 3.45 (0.43, 43.38) | 1.53 (0.39, 6.02) | 1.90 (0.69, 5.25) | 1.37 (0.45, 4.19) | 1.90 (0.69, 5.23) | 1.90 (0.69, 5.25) |
| Metformin vs Orlistat | 0.57 (0.12, 2.74) | 0.1829 (0.004, 8.201) | 0.44 (0.07, 2.77) | 0.79 (0.17, 3.80) | 0.13 (0.01, 1.40) | 0.59 (0.13, 2.63) | 0.79 (0.17, 3.80) |
| Metformin vs Phentermine-Topiramate | 3.99 (0.43, 36.66) | 3.22 (0.02, 303.52) | 3.16 (0.29, 35.08) | 4.02 (0.46, 35.03) | 8.44 (0.59, 121.30) | 11.70 (0.85, 161.17) | 4.02 (0.46, 35.03) |
| Metformin vs Sitagliptin | 0.62 (0.05, 8.25) | 0.81 (0.003, 261.57) | 0.50 (0.03, 7.84) | 0.62 (0.05, 7.59) | 0.45 (0.04, 5.68) | 0.62 (0.05, 7.53) | 0.62 (0.05, 7.59) |
| Orlistat vs LMA | 3.39 (1.04, 11.09) | 18.21 (0.97, 1349.54) | 3.45 (1.02, 11.66) | 2.40 (0.73, 7.90) | 10.85 (1.29, 91.55) | 3.21 (1.07, 9.62) | 2.40 (0.73, 7.90) |
| Orlistat vs Phentermine-Topiramate | 7.06 (0.72, 69.59) | 17.31 (0.09, 5075.36) | 7.15 (0.70, 72.89) | 5.06 (0.53, 48.21) | 66.71 (2.65, 1679.09) | 19.75 (1.39, 281.48) | 5.06 (0.53, 48.21) |
| Orlistat vs Sitagliptin | 1.11 (0.08, 15.51) | 4.43 (0.01, 3935.15) | 1.13 (0.08, 16.47) | 0.78 (0.06, 10.33) | 3.54 (0.16, 80.40) | 1.05 (0.08, 13.17) | 0.78 (0.06, 10.33) |
| Phentermine-Topiramate vs LMA | 0.48 (0.07, 3.40) | 1.07 (0.02, 98.89) | 0.48 (0.07, 3.49) | 0.47 (0.07, 3.21) | 0.16 (0.01, 1.83) | 0.16 (0.01, 1.83) | 0.47 (0.07, 3.21) |
| Phentermine-Topiramate vs Sitagliptin | 0.16 (0.01, 3.36) | 0.25 (4e-04, 239.69) | 0.16 (0.01, 3.51) | 0.15 (0.01, 3.05) | 0.05 (0.002, 1.48) | 0.05 (0.002, 1.48) | 0.15 (0.01, 3.05) |
| Sitagliptin vs LMA | 3.07 (0.29, 32.45) | 4.21 (0.03, 988.95) | 3.07 (0.28, 33.46) | 3.07 (0.31, 30.22) | 3.07 (0.31, 30.01) | 3.07 (0.31, 30.01) | 3.07 (0.31, 30.22) |
| **Outcome: serious adverse events** | | | | | | | |
| GLP-1 receptor agonists vs LMA | 1.07 (0.60, 1.91) | 1.11 (0.45, 2.85) | 1.08 (0.60, 1.93) | 1.07 (0.60, 1.91) | 0.80 (0.23, 2.76) | 1.07 (0.58, 1.96) | 1.07 (0.60, 1.91) |
| GLP-1 receptor agonists vs Metformin | 0.36 (0.01, 9.53) | 0 (0, 2.12) | 0.36 (0.01, 9.58) | 0.36 (0.01, 9.53) | 0.27 (0.01, 8.52) | 0.36 (0.01, 9.70) | 0.36 (0.01, 9.53) |
| GLP-1 receptor agonists vs Orlistat | 1.15 (0.36, 3.67) | 1.54 (0.27, 15.14) | 1.15 (0.36, 3.71) | 1.15 (0.36, 3.67) | 4.10 (0.15, 110.02) | 1.16 (0.34, 3.91) | 1.15 (0.36, 3.67) |
| GLP-1 receptor agonists vs Phentermine-Topiramate | 0.65 (0.07, 6.51) | 0 (0, 1.07) | 0.65 (0.07, 6.55) | 0.65 (0.07, 6.51) | 0.47 (0.02, 12.66) | 0.63 (0.03, 14.29) | 0.65 (0.07, 6.51) |
| GLP-1 receptor agonists vs Sitagliptin | 0.53 (0.09, 3.23) | 0.49 (0.03, 6.3) | 0.53 (0.09, 3.25) | 0.53 (0.09, 3.23) | 0.39 (0.05, 3.27) | 0.52 (0.08, 3.34) | 0.53 (0.09, 3.23) |
| Metformin vs LMA | 3.00 (0.12, 75.96) | 64342.52 (0.56, 9246134027160556) | 3.00 (0.12, 75.96) | 3.00 (0.12, 75.96) | 3.00 (0.12, 75.96) | 3.00 (0.12, 77.28) | 3.00 (0.12, 75.96) |
| Metformin vs Orlistat | 3.21 (0.11,94.76) | 97215.23 (0.67, 14368643717900340) | 3.21 (0.11, 94.76) | 3.21 (0.11, 94.76) | 15.30 (0.18, 1301.33) | 3.25 (0.11, 98.99) | 3.21 (0.11, 94.76) |
| Metformin vs Phentermine-Topiramate | 1.82 (0.04, 92.30) | 5.39 (0, 3133832470610.71) | 1.82 (0.04, 92.30) | 1.82 (0.04, 92.30) | 1.76 (0.02, 149.68) | 1.76 (0.02, 153.45) | 1.82 (0.04, 92.30) |
| Metformin vs Sitagliptin | 1.47 (0.04, 57.14) | 28181.94 (0.16, 4739685003520172) | 1.47 (0.04, 57.14) | 1.47 (0.04, 57.14) | 1.47 (0.04, 57.14) | 1.47 (0.04, 58.89) | 1.47 (0.04, 57.14) |
| Orlistat vs LMA | 0.94 (0.34, 2.57) | 0.72 (0.09, 3.27) | 0.94 (0.34, 2.57) | 0.94 (0.34, 2.57) | 0.20 (0.01, 4.14) | 0.92 (0.32, 2.65) | 0.94 (0.34, 2.57) |
| Orlistat vs Phentermine-Topiramate | 0.57 (0.05, 6.57) | 0 (0, 0.75) | 0.57 (0.05, 6.57) | 0.57 (0.05, 6.57) | 0.11 (0.00, 8.58) | 0.54 (0.02, 13.88) | 0.57 (0.05, 6.57) |
| Orlistat vs Sitagliptin | 0.46 (0.06, 3.37) | 0.31 (0.01, 4.93) | 0.46 (0.06, 3.37) | 0.46 (0.06, 3.37) | 0.10 (0.003, 3.18) | 0.45 (0.06, 3.49) | 0.46 (0.06, 3.37) |
| Phentermine-Topiramate vs LMA | 1.65 (0.18, 15.30) | 9105.08 (1.13, 275979612148218) | 1.65 (0.18, 15.30) | 1.65 (0.18, 15.30) | 1.71 (0.08, 36.09) | 1.71 (0.08, 36.75) | 1.65 (0.18, 15.30) |
| Phentermine-Topiramate vs Sitagliptin | 0.81 (0.05, 13.47) | 4082.96 (0.31, 139249316945121) | 0.81 (0.05, 13.47) | 0.81 (0.05, 13.47) | 0.84 (0.03, 27.76) | 0.84 (0.02, 28.65) | 0.81 (0.05, 13.47) |
| Sitagliptin vs LMA | 2.04 (0.37, 11.44) | 2.27 (0.21, 31.17) | 2.04 (0.37, 11.44) | 2.04 (0.37, 11.44) | 2.04 (0.37, 11.44) | 2.04 (0.35, 11.81) | 2.04 (0.37, 11.44) |
| **Outcome: nausea events** | | | | | | | |
| GLP-1 receptor agonists vs LMA | 3.30 (2.31, 4.72) | 3.55 (2.26, 5.88) | 3.25 (2.25, 4.68) | 3.30 (2.31, 4.72) | 3.73 (2.21, 6.32) | 3.25 (2.25, 4.68) | 3.30 (2.31, 4.72) |
| GLP-1 receptor agonists vs Metformin | 0.89 (0.36, 2.24) | 0.78 (0.26, 2.2) | 1.01 (0.39, 2.60) | 0.89 (0.36, 2.24) | 1.16 (0.42, 3.22) | 1.01 (0.39, 2.60) | 0.89 (0.36, 2.24) |
| GLP-1 receptor agonists vs Orlistat | 2.81 (1.57, 5.04) | 3.02 (1.35, 7.14) | 2.76 (1.54, 4.98) | 2.81 (1.57, 5.04) | 3.32 (1.13, 9.82) | 2.76 (1.54, 4.98) | 2.81 (1.57, 5.04) |
| GLP-1 receptor agonists vs Phentermine-Topiramate | 2.80 (0.54, 14.43) | 2.64 (0.3, 15.31) | 2.75 (0.53, 14.21) | 2.80 (0.54, 14.43) | 3.16 (0.59, 17.06) | 2.75 (0.53, 14.21) | 2.80 (0.54, 14.43) |
| GLP-1 receptor agonists vs Sitagliptin | 0.63 (0.07, 5.68) | 0.52 (0.02, 4.59) | 0.62 (0.07, 5.59) | 0.63 (0.07, 5.68) | 0.72 (0.08, 6.65) | 0.62 (0.07, 5.59) | 0.63 (0.07, 5.68) |
| Metformin vs LMA | 3.69 (1.59, 8.59) | 4.58 (1.83, 12.67) | 3.23 (1.34, 7.78) | 3.69 (1.59, 8.59) | 3.23 (1.34, 7.78) | 3.23 (1.34, 7.78) | 3.69 (1.59, 8.59) |
| Metformin vs Orlistat | 3.14 (1.20, 8.23) | 3.89 (1.25, 13.34) | 2.75 (1.02, 7.42) | 3.14 (1.20, 8.23) | 2.87 (0.79, 10.46) | 2.75 (1.02, 7.42) | 3.14 (1.20, 8.23) |
| Metformin vs Phentermine-Topiramate | 3.13 (0.51, 19.11) | 3.42 (0.33, 23.98) | 2.73 (0.44, 16.98) | 3.13 (0.51, 19.11) | 2.73 (0.44, 16.98) | 2.73 (0.44, 16.98) | 3.13 (0.51, 19.11) |
| Metformin vs Sitagliptin | 0.71 (0.07, 7.23) | 0.66 (0.02, 6.97) | 0.62 (0.06, 6.40) | 0.71 (0.07, 7.23) | 0.62 (0.06, 6.40) | 0.62 (0.06, 6.40) | 0.71 (0.07, 7.23) |
| Orlistat vs LMA | 1.17 (0.74, 1.86) | 1.18 (0.59, 2.34) | 1.17 (0.74, 1.86) | 1.17 (0.74, 1.86) | 1.12 (0.44, 2.89) | 1.17 (0.74, 1.86) | 1.17 (0.74, 1.86) |
| Orlistat vs Phentermine-Topiramate | 0.99 (0.19, 5.26) | 0.87 (0.09, 5.36) | 0.99 (0.19, 5.26) | 0.99 (0.19, 5.26) | 0.95 (0.15, 6.11) | 0.99 (0.19, 5.26) | 0.99 (0.19, 5.26) |
| Orlistat vs Sitagliptin | 0.22 (0.02, 2.06) | 0.17 (0.01, 1.57) | 0.22 (0.02, 2.06) | 0.22 (0.02, 2.06) | 0.22 (0.02, 2.29) | 0.22 (0.02, 2.06) | 0.22 (0.02, 2.06) |
| Phentermine-Topiramate vs LMA | 1.18 (0.24, 5.86) | 1.34 (0.25, 11.52) | 1.18 (0.24, 5.86) | 1.18 (0.24, 5.86) | 1.18 (0.24, 5.86) | 1.18 (0.24, 5.86) | 1.18 (0.24, 5.86) |
| Phentermine-Topiramate vs Sitagliptin | 0.23 (0.02, 3.35) | 0.19 (0, 4.06) | 0.23 (0.02, 3.35) | 0.23 (0.02, 3.35) | 0.23 (0.02, 3.35) | 0.23 (0.02, 3.35) | 0.23 (0.02, 3.35) |
| Sitagliptin vs LMA | 5.22 (0.60, 45.57) | 6.91 (0.83, 194.95) | 5.22 (0.60, 45.57) | 5.22 (0.60, 45.57) | 5.22 (0.60, 45.57) | 5.22 (0.60, 45.57) | 5.22 (0.60, 45.57) |
| **Outcome: vomiting events** | | | | | | | |
| GLP-1 receptor agonists vs LMA | 5.62 (3.54, 8.93) | 7.13 (3.67, 18.7) | 5.75 (3.56, 9.28) | 5.62 (3.54, 8.93) | 8.71 (3.77, 20.13) | 5.75 (3.56, 9.28) | 5.62 (3.54, 8.93) |
| GLP-1 receptor agonists vs Metformin | 0.84 (0.09, 7.67) | 0.77 (0.02, 11.1) | 0.85 (0.09, 7.87) | 0.84 (0.09, 7.67) | 1.29 (0.12, 13.51) | 0.85 (0.09, 7.87) | 0.84 (0.09, 7.67) |
| GLP-1 receptor agonists vs Orlistat | 5.62 (1.72, 18.31) | 7.14 (1.14, 59.75) | 5.75 (1.75 , 18.84) | 5.62 (1.72 , 18.31) | 8.71 (2.13 , 35.57) | 5.75 (1.75 , 18.84) | 5.62 (1.72 , 18.31) |
| GLP-1 receptor agonists vs Sitagliptin | 2.75 (0.46, 16.35) | 3.26 (0.25, 38.1) | 2.81 (0.47, 16.79) | 2.75 (0.46, 16.35) | 4.26 (0.61, 29.65) | 2.81 (0.47, 16.79) | 2.75 (0.46, 16.35) |
| Metformin vs LMA | 6.73 (0.77, 58.82) | 9.39 (0.81, 349.99) | 6.73 (0.77, 58.82) | 6.73 (0.77, 58.82) | 6.73 (0.75, 60.16) | 6.73 (0.77, 58.82) | 6.73 (0.77, 58.82) |
| Metformin vs Orlistat | 6.73 (0.60, 76.06) | 9.69 (0.45, 532.98) | 6.73 (0.60 , 76.06) | 6.73 (0.60 , 76.06) | 6.73 (0.57 , 79.16) | 6.73 (0.60 , 76.06) | 6.73 (0.60 , 76.06) |
| Metformin vs Sitagliptin | 3.29 (0.21, 52.46) | 4.35 (0.12, 287.7) | 3.29 (0.21, 52.46) | 3.29 (0.21, 52.46) | 3.29 (0.20, 54.34) | 3.29 (0.21, 52.46) | 3.29 (0.21, 52.46) |
| Orlistat vs LMA | 1.00 (0.34, 2.96) | 1.00 (0.16, 6.08) | 1.00 (0.34 , 2.96) | 1.00 (0.34 , 2.96) | 1.00 (0.32 , 3.10) | 1.00 (0.34 , 2.96) | 1.00 (0.34 , 2.96) |
| Orlistat vs Sitagliptin | 0.49 (0.06, 3.75) | 0.45 (0.02, 7.82) | 0.49 (0.06 , 3.75) | 0.49 (0.06 , 3.75) | 0.49 (0.06 , 3.93) | 0.49 (0.06 , 3.75) | 0.49 (0.06 , 3.75) |
| Sitagliptin vs LMA | 2.04 (0.37, 11.44) | 2.22 (0.24, 28.76) | 2.04 (0.37, 11.44) | 2.04 (0.37, 11.44) | 2.04 (0.36, 11.76) | 2.04 (0.37, 11.44) | 2.04 (0.37, 11.44) |
| **Outcome: diarrhea events** | | | | | | | |
| GLP-1 receptor agonists vs LMA | 1.66 (1.17, 2.35) | 1.78 (1.16, 2.97) | 1.64 (1.15, 2.33) | 1.66 (1.17, 2.35) | 1.85 (1.05, 3.26) | 1.64 (1.15, 2.33) | 1.66 (1.17, 2.35) |
| GLP-1 receptor agonists vs Metformin | 1.08 (0.48, 2.41) | 1.09 (0.41, 2.86) | 1.07 (0.48, 2.39) | 1.08 (0.48, 2.41) | 1.20 (0.48, 3.02) | 1.07 (0.48, 2.39) | 1.08 (0.48, 2.41) |
| GLP-1 receptor agonists vs Orlistat | 0.54 (0.21, 1.38) | 0.57 (0.17, 1.94) | 0.54 (0.21 , 1.37) | 0.54 (0.21 , 1.38) | 0.60 (0.21 , 1.70) | 0.54 (0.21 , 1.37) | 0.54 (0.21 , 1.38) |
| GLP-1 receptor agonists vs Sitagliptin | 2.83 (0.63, 12.69) | 3.21 (0.6, 21.01) | 2.79 (0.62, 12.54) | 2.83 (0.63, 12.69) | 3.15 (0.66, 15.07) | 2.79 (0.62, 12.54) | 2.83 (0.63, 12.69) |
| Metformin vs LMA | 1.54 (0.74, 3.17) | 1.65 (0.72, 4.03) | 1.54 (0.74, 3.17) | 1.54 (0.74, 3.17) | 1.54 (0.74, 3.17) | 1.54 (0.74, 3.17) | 1.54 (0.74, 3.17) |
| Metformin vs Orlistat | 0.50 (0.16, 1.56) | 0.52 (0.13, 2.16) | 0.50 (0.16 , 1.56) | 0.50 (0.16 , 1.56) | 0.50 (0.16 , 1.56) | 0.50 (0.16 , 1.56) | 0.50 (0.16 , 1.56) |
| Metformin vs Sitagliptin | 2.62 (0.51, 13.35) | 2.98 (0.48, 22.03) | 2.62 (0.51, 13.35) | 2.62 (0.51, 13.35) | 2.62 (0.51, 13.35) | 2.62 (0.51, 13.35) | 2.62 (0.51, 13.35) |
| Orlistat vs LMA | 3.06 (1.28, 7.28) | 3.14 (1.06, 10.08) | 3.06 (1.28 , 7.28) | 3.06 (1.28 , 7.28) | 3.06 (1.28 , 7.28) | 3.06 (1.28 , 7.28) | 3.06 (1.28 , 7.28) |
| Orlistat vs Sitagliptin | 5.21 (0.95, 28.48) | 5.72 (0.8, 48.04) | 5.21 (0.95 , 28.48) | 5.21 (0.95 , 28.48) | 5.21 (0.95 , 28.48) | 5.21 (0.95 , 28.48) | 5.21 (0.95 , 28.48) |
| Sitagliptin vs LMA | 0.59 (0.14, 2.53) | 0.56 (0.09, 2.83) | 0.59 (0.14, 2.53) | 0.59 (0.14, 2.53) | 0.59 (0.14, 2.53) | 0.59 (0.14, 2.53) | 0.59 (0.14, 2.53) |

### Appendix 11: P-scores for each outcomes

| **Outcomes** | **P-score (common)** | **P-score (random)** |
| --- | --- | --- |
| **Change in BMI from baseline** | | |
| Phentermine-Topiramate | 1.0000 | 0.9787 |
| Metformin_Fluoxetine | 0.7111 | 0.6482 |
| GLP-1 receptor agonists | 0.848 | 0.6286 |
| Orlistat | 0.4547 | 0.5877 |
| Fluoxetine | 0.5335 | 0.5786 |
| Metformin | 0.0831 | 0.4287 |
| Topiramate | 0.5608 | 0.3956 |
| Lifestyle modification alone | 0.2615 | 0.1402 |
| Sitagliptin | 0.0473 | 0.1138 |
| **Change in weight from baseline** | | |
| Phentermine-Topiramate | 1.0000 | 0.9998 |
| Orlistat | 0.5688 | 0.6874 |
| GLP-1 receptor agonists | 0.773 | 0.6369 |
| Metformin | 0.4306 | 0.4731 |
| Topiramate | 0.5402 | 0.4597 |
| Lifestyle modification alone | 0.184 | 0.1644 |
| Sitagliptin | 0.0034 | 0.0787 |
| **Percentage of participants achieving BMI reduction of at least 5%** | | |
| Phentermine-Topiramate | 0.9725 | 0.9276 |
| GLP-1 receptor agonists | 0.7256 | 0.6939 |
| Topiramate | 0.3740 | 0.3940 |
| Orlistat | 0.3684 | 0.3673 |
| Lifestyle modification alone | 0.0596 | 0.1172 |
| **Percentage of participants achieving BMI reduction of at least 10%** | | |
| Phentermine-Topiramate | 0.9788 | 0.9543 |
| GLP-1 receptor agonists | 0.7214 | 0.6374 |
| Orlistat | 0.4754 | 0.5063 |
| Topiramate | 0.1890 | 0.2332 |
| Lifestyle modification alone | 0.1354 | 0.1688 |
| **Change in BMI z-score from baseline** | | |
| Metformin | 0.9996 | 0.9990 |
| Orlistat | 0.5718 | 0.5642 |
| Topiramate | 0.3652 | 0.3678 |
| Lifestyle modification alone | 0.0634 | 0.0689 |
| **Change in BMI SDS from baseline** | | |
| GLP-1 receptor agonists | 0.9712 | 0.9036 |
| Metformin | 0.5401 | 0.597 |
| Orlistat | 0.483 | 0.4424 |
| Lifestyle modification alone | 0.0057 | 0.057 |
| **Total gastrointestinal adverse events** | | |
| Lifestyle modification alone | 0.8355 | 0.8392 |
| Phentermine-Topiramate | 0.8521 | 0.8285 |
| Metformin | 0.5584 | 0.5742 |
| GLP-1 receptor agonists | 0.2497 | 0.2534 |
| Orlistat | 0.0043 | 0.0048 |
| **Discontinuation due to adverse events** | | |
| Phentermine-Topiramate | 0.8856 | 0.8788 |
| Lifestyle modification alone | 0.7698 | 0.7689 |
| Metformin | 0.4251 | 0.4308 |
| GLP-1 receptor agonists | 0.4095 | 0.4023 |
| Sitagliptin | 0.3013 | 0.3121 |
| Orlistat | 0.2087 | 0.2071 |
| **Serious adverse events** | | |
| Orlistat | 0.6691 | 0.6691 |
| Lifestyle modification alone | 0.6506 | 0.6506 |
| GLP-1 receptor agonists | 0.5886 | 0.5886 |
| Phentermine-Topiramate | 0.4381 | 0.4381 |
| Sitagliptin | 0.3389 | 0.3389 |
| Metformin | 0.3147 | 0.3147 |
| **Nausea events** | | |
| Lifestyle modification alone | 0.853 | 0.853 |
| Orlistat | 0.7293 | 0.7293 |
| Phentermine-Topiramate | 0.7117 | 0.7117 |
| GLP-1 receptor agonists | 0.2727 | 0.2727 |
| Metformin | 0.228 | 0.228 |
| Sitagliptin | 0.2054 | 0.2054 |
| **Vomiting events** | | |
| Lifestyle modification alone | 0.8124 | 0.8124 |
| Orlistat | 0.7976 | 0.7976 |
| Sitagliptin | 0.5302 | 0.5302 |
| Metformin | 0.1851 | 0.1851 |
| GLP-1 receptor agonists | 0.1746 | 0.1746 |
| **Diarrhea events** | | |
| Sitagliptin | 0.8807 | 0.8807 |
| Lifestyle modification alone | 0.7766 | 0.7766 |
| Metformin | 0.4263 | 0.4263 |
| GLP-1 receptor agonists | 0.3537 | 0.3537 |
| Orlistat | 0.0627 | 0.0627 |

### Appendix 12: Reference list for included studies

We screened 2072 records from the databases (PubMed, Embase, the Cochrane Library), ICTRP and ClinicalTrials.gov, 2030 studies were excluded according to the predefined criteria. A total of 42 studies were included in this network meta-analysis[10-51].

**Reference**

1.	Liu L, Bai H, Wang C, Seery S, Wang Z, Duan J, Li S, Xue P, Wang G, Sun Y *et al*: Efficacy and Safety of First-Line Immunotherapy Combinations for Advanced NSCLC: A Systematic Review and Network Meta-Analysis. (1556-1380 (Electronic)).

2.	Sterne JAC, Savović J, Page MJ, Elbers RG, Blencowe NS, Boutron I, Cates CJ, Cheng HY, Corbett MS, Eldridge SM *et al*: RoB 2: a revised tool for assessing risk of bias in randomised trials. *Bmj* 2019, 366:l4898.

3.	Roman YM, Burela PA, Pasupuleti V, Piscoya A, Vidal JE, Hernandez AV: Ivermectin for the Treatment of Coronavirus Disease 2019: A Systematic Review and Meta-analysis of Randomized Controlled Trials. *Clin Infect Dis* 2022, 74(6):1022-1029.

4.	Shi Q, Wang Y, Hao Q, Vandvik PO, Guyatt G, Li J, Chen Z, Xu S, Shen Y, Ge L *et al*: Pharmacotherapy for adults with overweight and obesity: a systematic review and network meta-analysis of randomised controlled trials. *Lancet* 2022, 399(10321):259-269.

5.	Doi S, Furuya-Kanamori L, Xu C, Lin L, Chivese T, Thalib L: Questionable utility of the relative risk in clinical research: A call for change to practice. *Journal of Clinical Epidemiology* 2020.

6.	Chen AK, Roberts CK, Barnard RJ: Effect of a short-term diet and exercise intervention on metabolic syndrome in overweight children. *Metabolism* 2006, 55(7):871-878.

7.	Food and Drug Administration. Guidance for Industry Developing Products for Weight Management [<https://www.fda.gov/regulatory-information/search-fda-guidance-documents/developing-products-weight-management-revision-1>]

8.	Shim S, Yoon BH, Shin IS, Bae JM: Network meta-analysis: application and practice using Stata. *Epidemiol Health* 2017, 39:e2017047.

9.	Cortese S, Del Giovane C, Chamberlain S, Philipsen A, Young S, Bilbow A, Cipriani A: Pharmacological and non-pharmacological interventions for adults with ADHD: protocol for a systematic review and network meta-analysis. *BMJ Open* 2022, 12(3):e058102.

10.	Maahs D, de Serna DG, Kolotkin RL, Ralston S, Sandate J, Qualls C, Schade DS: Randomized, double-blind, placebo-controlled trial of orlistat for weight loss in adolescents. *Endocr Pract* 2006, 12(1):18-28.

11.	Yu CC, Li AM, Chan KO, Chook P, Kam JT, Au CT, So RC, Sung RY, McManus AM: Orlistat improves endothelial function in obese adolescents: a randomised trial. *J Paediatr Child Health* 2013, 49(11):969-975.

12.	Chanoine JP, Hampl S, Jensen C, Boldrin M, Hauptman J: Effect of orlistat on weight and body composition in obese adolescents: a randomized controlled trial. *Jama* 2005, 293(23):2873-2883.

13.	Ozkan B, Bereket A, Turan S, Keskin S: Addition of orlistat to conventional treatment in adolescents with severe obesity. *Eur J Pediatr* 2004, 163(12):738-741.

14.	Li W, Li M, Kong D: Clinical efficacy of metformin combined with lifestyle intervention for treatment of childhood obesity with hyperinsulinemia. *International Journal of Clinical and Experimental Medicine* 2019, 12(6):7644-7650.

15.	Atabek ME, Pirgon O: Use of metformin in obese adolescents with hyperinsulinemia: a 6-month, randomized, double-blind, placebo-controlled clinical trial. *J Pediatr Endocrinol Metab* 2008, 21(4):339-348.

16.	Kelsey MM, Hilkin A, Pyle L, Severn C, Utzschneider K, Van Pelt RE, Zeitler PS, Nadeau KJ: Two-Year Treatment with Metformin during Puberty Does Not Preserve beta-Cell Function in Youth with Obesity. *Journal of Clinical Endocrinology and Metabolism* 2021, 106(7):E2622-E2632.

17.	Nadeau KJ, Ehlers LB, Zeitler PS, Love-Osborne K: Treatment of non-alcoholic fatty liver disease with metformin versus lifestyle intervention in insulin-resistant adolescents. *Pediatr Diabetes* 2009, 10(1):5-13.

18.	Rezvanian H, Hashemipour M, Kelishadi R, Tavakoli N, Poursafa P: A randomized, triple masked, placebo-controlled clinical trial for controlling childhood obesity. *World J Pediatr* 2010, 6(4):317-322.

19.	Vendrell J, Vilarrasa N, Bridger T, MacDonald S, Baltzer F, Rodd C: Randomized placebo-controlled trial of metformin for adolescents with polycystic ovary syndrome. *Sci Rep* 2006, 160(3):241-246.

20.	Mauras N, DelGiorno C, Hossain J, Bird K, Killen K, Merinbaum D, Weltman A, Damaso L, Balagopal P: Metformin use in children with obesity and normal glucose tolerance--effects on cardiovascular markers and intrahepatic fat. *J Pediatr Endocrinol Metab* 2012, 25(1-2):33-40.

21.	Warnakulasuriya LS, Fernando MMA, Adikaram AVN, Thawfeek ARM, Anurasiri WL, Silva RR, Sirasa MSF, Rytter E, Forslund AH, Samaranayake DL *et al*: Metformin in the Management of Childhood Obesity: A Randomized Control Trial. *Child Obes* 2018, 14(8):553-565.

22.	Kendall D, Vail A, Amin R, Barrett T, Dimitri P, Ivison F, Kibirige M, Mathew V, Matyka K, McGovern A *et al*: Metformin in obese children and adolescents: the MOCA trial. *J Clin Endocrinol Metab* 2013, 98(1):322-329.

23.	Clarson CL, Mahmud FH, Baker JE, Clark HE, McKay WM, Schauteet VD, Hill DJ: Metformin in combination with structured lifestyle intervention improved body mass index in obese adolescents, but did not improve insulin resistance. *Endocrine* 2009, 36(1):141-146.

24.	Pastor-Villaescusa B, Cañete MD, Caballero-Villarraso J, Hoyos R, Latorre M, Vázquez-Cobela R, Plaza-Díaz J, Maldonado J, Bueno G, Leis R *et al*: Metformin for Obesity in Prepubertal and Pubertal Children: A Randomized Controlled Trial. *Pediatrics* 2017, 140(1).

25.	Wilson DM, Abrams SH, Aye T, Lee PDK, Lenders C, Lustig RH, Osganian SV, Feldman HA, Fechner P, Robinson T *et al*: Metformin extended release treatment of adolescent obesity: A 48-week randomized, double-blind, placebo-controlled trial with 48-week follow-up. *Archives of Pediatrics and Adolescent Medicine* 2010, 164(2):116-123.

26.	Wiegand S, l'Allemand D, Hübel H, Krude H, Bürmann M, Martus P, Grüters A, Holl RW: Metformin and placebo therapy both improve weight management and fasting insulin in obese insulin-resistant adolescents: a prospective, placebo-controlled, randomized study. *Eur J Endocrinol* 2010, 163(4):585-592.

27.	van der Aa MP, Elst MAJ, van de Garde EMW, van Mil EGAH, Knibbe CAJ, van der Vorst MMJ: Long-term treatment with metformin in obese, insulin-resistant adolescents: results of a randomized double-blinded placebo-controlled trial. *Nutrition and Diabetes* 2016, 6(8):e228.

28.	Rynders C, Weltman A, Delgiorno C, Balagopal P, Damaso L, Killen K, Mauras N: Lifestyle intervention improves fitness independent of metformin in obese adolescents. *Med Sci Sports Exerc* 2012, 44(5):786-792.

29.	Evia-Viscarra ML, Rodea-Montero ER, Apolinar-Jiménez E, Muñoz-Noriega N, García-Morales LM, Leaños-Pérez C, Figueroa-Barrón M, Sánchez-Fierros D, Reyes-García JG: The effects of metformin on inflammatory mediators in obese adolescents with insulin resistance: controlled randomized clinical trial. *J Pediatr Endocrinol Metab* 2012, 25(1-2):41-49.

30.	Yanovski JA, Krakoff J, Salaita CG, McDuffie JR, Kozlosky M, Sebring NG, Reynolds JC, Brady SM, Calis KA: Effects of metformin on body weight and body composition in obese insulin-resistant children: a randomized clinical trial. *Diabetes* 2011, 60(2):477-485.

31.	Bassols J, Martínez-Calcerrada JM, Osiniri I, Díaz-Roldán F, Xargay-Torrent S, Mas-Parés B, Dorado-Ceballos E, Prats-Puig A, Carreras-Badosa G, de Zegher F *et al*: Effects of metformin administration on endocrine-metabolic parameters, visceral adiposity and cardiovascular risk factors in children with obesity and risk markers for metabolic syndrome: a pilot study. *PloS one* 2019, 14(12):e0226303.

32.	Garibay-Nieto N, Queipo-García G, Alvarez F, Bustos M, Villanueva E, Ramírez F, León M, Laresgoiti-Servitje E, Duggirala R, Macías T *et al*: Effects of Conjugated Linoleic Acid and Metformin on Insulin Sensitivity in Obese Children: randomized Clinical Trial. *Journal of clinical endocrinology and metabolism* 2017, 102(1):132‐140.

33.	Kay JP, Alemzadeh R, Langley G, D'Angelo L, Smith P, Holshouser S: Beneficial effects of metformin in normoglycemic morbidly obese adolescents. *Metabolism* 2001, 50(12):1457-1461.

34.	Burgert TS, Duran EJ, Goldberg-Gell R, Dziura J, Yeckel CW, Katz S, Tamborlane WV, Caprio S: Short-term metabolic and cardiovascular effects of metformin in markedly obese adolescents with normal glucose tolerance. *Pediatr Diabetes* 2008, 9(6):567-576.

35.	Shankar RR, Zeitler P, Deeb A, Jalaludin MY, Garcia R, Newfield RS, Samoilova Y, Rosario CA, Shehadeh N, Saha CK *et al*: A randomized clinical trial of the efficacy and safety of sitagliptin as initial oral therapy in youth with type 2 diabetes. *Pediatric Diabetes* 2022, 23(2):173-182.

36.	Fox CK, Clark JM, Rudser KD, Ryder JR, Gross AC, Nathan BM, Sunni M, Dengel DR, Billington CJ, Bensignor MO *et al*: Exenatide for weight-loss maintenance in adolescents with severe obesity: A randomized, placebo-controlled trial. *Obesity (Silver Spring, Md)* 2022, 30(5):1105‐1115.

37.	Kelly AS, Metzig AM, Rudser KD, Fitch AK, Fox CK, Nathan BM, Deering MM, Schwartz BL, Abuzzahab MJ, Gandrud LM *et al*: Exenatide as a weight-loss therapy in extreme pediatric obesity: a randomized, controlled pilot study. *Obesity (Silver Spring, Md)* 2012, 20(2):364‐370.

38.	Weghuber D, Forslund A, Ahlstrom H, Alderborn A, Bergstrom K, Brunner S, Cadamuro J, Ciba I, Dahlbom M, Heu V *et al*: A 6-month randomized, double-blind, placebo-controlled trial of weekly exenatide in adolescents with obesity. *Pediatric Obesity* 2020, 15(7):e12624.

39.	Arslanian SA, Hannon T, Zeitler P, Chao LC, Boucher-Berry C, Barrientos-Pérez M, Bismuth E, Dib S, Cho JI, Cox D: Once-Weekly Dulaglutide for the Treatment of Youths with Type 2 Diabetes. *N Engl J Med* 2022, 387(5):433-443.

40.	Kelly AS, Auerbach P, Barrientos-Perez M, Gies I, Hale PM, Marcus C, Mastrandrea LD, Prabhu N, Arslanian S: A Randomized, Controlled Trial of Liraglutide for Adolescents with Obesity. *N Engl J Med* 2020, 382(22):2117-2128.

41.	Tamborlane WV, Barrientos-Pérez M, Fainberg U, Frimer-Larsen H, Hafez M, Hale PM, Jalaludin MY, Kovarenko M, Libman I, Lynch JL *et al*: Liraglutide in Children and Adolescents with Type 2 Diabetes. *N Engl J Med* 2019, 381(7):637-646.

42.	Danne T, Biester T, Kapitzke K, Jacobsen SH, Jacobsen LV, Petri KCC, Hale PM, Kordonouri O: Liraglutide in an Adolescent Population with Obesity: A Randomized, Double-Blind, Placebo-Controlled 5-Week Trial to Assess Safety, Tolerability, and Pharmacokinetics of Liraglutide in Adolescents Aged 12-17 Years. *J Pediatr* 2017, 181:146-153.e143.

43.	Mastrandrea LD, Witten L, Carlsson Petri KC, Hale PM, Hedman HK, Riesenberg RA: Liraglutide effects in a paediatric (7-11 y) population with obesity: A randomized, double-blind, placebo-controlled, short-term trial to assess safety, tolerability, pharmacokinetics, and pharmacodynamics. *Pediatr Obes* 2019, 14(5):e12495.

44.	Fox CK, Kaizer AM, Rudser KD, Nathan BM, Gross AC, Sunni M, Jennifer Abuzzahab M, Schwartz BL, Kumar S, Petryk A *et al*: Meal replacements followed by topiramate for the treatment of adolescent severe obesity: A pilot randomized controlled trial. *Obesity (Silver Spring)* 2016, 24(12):2553-2561.

45.	Hsia DS, Gosselin NH, Williams J, Farhat N, Marier JF, Shih W, Peterson C, Siegel R: A randomized, double-blind, placebo-controlled, pharmacokinetic and pharmacodynamic study of a fixed-dose combination of phentermine/topiramate in adolescents with obesity. *Diabetes Obes Metab* 2020, 22(4):480-491.

46.	Barrientos-Pérez M, Hsia DS: A study on pharmacokinetics, pharmacodynamics and safety of lixisenatide in children and adolescents with type 2 diabetes. *Pediatr Obes* 2022, 23(6):641-648.

47.	Kelly AS, Bensignor MO, Hsia DS, Shoemaker AH, Shih W, Peterson C, Varghese ST: Phentermine/Topiramate for the Treatment of Adolescent Obesity. *NEJM Evid* 2022, 1(6).

48.	Weghuber D, Barrett T, Barrientos-Pérez M, Gies I, Hesse D, Jeppesen OK, Kelly AS, Mastrandrea LD, Sørrig R, Arslanian S: Once-Weekly Semaglutide in Adolescents with Obesity. *N Engl J Med* 2022, 387(24):2245-2257.

49.	Diene G, Angulo M, Hale PM, Jepsen CH, Hofman PL, Hokken-Koelega A, Ramesh C, Turan S, Tauber M: Liraglutide for Weight Management in Children and Adolescents With Prader-Willi Syndrome and Obesity. *J Clin Endocrinol Metab* 2022, 108(1):4-12.

50.	Safety and Efficacy of Xenical in Children and Adolescents With Obesity-Related Diseases [<https://clinicaltrials.gov/ct2/show/results/NCT00001723>]

51.	Impact of Metformin in Teens With Polycystic Ovary Syndrome (PCOS) on Oral Contraceptive Therapy [<https://clinicaltrials.gov/ct2/show/results/NCT00283816>]
